# Resurgence of SARS-CoV-2 in India: Potential role of the B.1.617.2 (Delta) variant and delayed interventions

**DOI:** 10.1101/2021.06.23.21259405

**Authors:** Maxwell Salvatore, Rupam Bhattacharyya, Soumik Purkayastha, Lauren Zimmermann, Debashree Ray, Aditi Hazra, Michael Kleinsasser, Thomas Mellan, Charlie Whittaker, Seth Flaxman, Samir Bhatt, Swapnil Mishra, Bhramar Mukherjee

## Abstract

India has seen a surge of SARS-CoV-2 infections and deaths in early part of 2021, despite having controlled the epidemic during 2020. Building on a two-strain, semi-mechanistic model that synthesizes mortality and genomic data, we find evidence that altered epidemiological properties of B.1.617.2 (Delta) variant play an important role in this resurgence in India. Under all scenarios of immune evasion, we find an increased transmissibility advantage for B.1617.2 against all previously circulating strains. Using an extended SIR model accounting for reinfections and wanning immunity, we produce evidence in support of how early public interventions in March 2021 would have helped to control transmission in the country. We argue that enhanced genomic surveillance along with constant assessment of risk associated with increased transmission is critical for pandemic responsiveness.

**One Sentence Summary:** Altered epidemiological characteristics of B.1.617.2 and delayed public health interventions contributed to the resurgence of SARS-CoV-2 in India from February to May 2021.

## INTRODUCTION

The first case of SARS-CoV-2 in India (where 18% of the world’s population lives (*1*)) was reported on January 30, 2020. India was proactive in implementing a suite of effective and timely public health interventions in the first wave of its epidemic. On March 3, 2020, with few positively confirmed COVID-19 cases and no reported deaths, India began border controls through travel bans and visa cancellations (*2*). Within two weeks of the World Health Organization (WHO) officiating COVID-19 as a pandemic (*3*), India made the historic and highly debated decision to implement a 21-day national lockdown starting March 25, 2020, with only 536 reported cases and 11 COVID-19 attributable deaths (*4*). The lockdown was later extended to four distinct phases that lasted until May 31, 2020 (*5–7*). During this lockdown, India scaled up testing (*8*) and treatment facilities (*9*) within the constraints of a low-resource setting. Public acceptance of masks, avoidance of social gatherings, and adoption of other non-pharmaceutical interventions (NPIs) was impressive during the initial phase (*10*). A gradual relaxation of nationwide restrictions started in monthly phases from June 1, 2020 (*11*). While countries such as Italy, China, and South Korea observed a rapid and sustained decline in case counts within a 3- to 4-week period of initiating lockdowns and/or other strict NPIs in 2020, India’s daily case counts continued to increase during and after lockdown, albeit with a substantially reduced doubling time (*12*). India’s daily new case counts peaked on September 16, 2020, with 97,860 cases and 1,281 daily new deaths reported. During this time, India also managed to reduce overall COVID-19 case fatality rates from what was predicted (*13*), particularly given its large vulnerable population size and fragile healthcare system (*4*).

Since the September peak, the incidence curve declined steadily to less than 10,000 daily new cases in February of 2021 (*14*). National serosurveys and epidemiological model estimates indicated a substantial infection under-ascertainment rate, suggesting only about 6% of infections in India were reported by the end of 2020 (*15–18*). There were discussions about urban metros in India approaching herd immunity thresholds with many of them reporting more than 50% seropositivity (*17, 19, 20*). The third national serosurvey in January 2021 indicated 21.5% of adults in India had evidence of a past COVID-19 infection (*21*). As the country further relaxed restrictions, COVID-appropriate behaviors diminished with time (*10*), crowded public transportation system restarted, large indoor and outdoor gatherings were taking place without meaningful adherence to proper face coverings. Social and religious events, weddings, political rallies, mass protests - all cultural facets that define the tapestry of life in India were in full force. The year 2021 started on an optimistic note with multiple vaccine trials going on in India and globally (*10*) with promising efficacy and safety results. India formalized operational guidelines for its national vaccine distribution with emergency-use approval for two vaccines (*22*), including prioritizing beneficiaries (*23*). Vaccination began on January 16 with an initial focus on healthcare workers. The vaccine roll-out in India has been sluggish, and initially this was in part due to an underappreciation of the threat that SARS-CoV-2 still posed. Only 8.7 million doses were administered in the first one month, and 0.65% of the population received at least one dose on February 15. By April 1, only 5.2% of the population had received at least one dose in India with less than 2% of the population fully vaccinated.

After a steady decline for about four months, an uptick in cases was noted in three Indian states in February 2021 (Maharashtra, Punjab and Chhattisgarh) with the national effective reproduction number crossing the threshold of one on February 14 (*24*). No strict control measures were reintroduced in the two months following the initial indications of a resurgence in transmission. Indeed, the first comprehensive lockdown only started on April 14 in Maharashtra (*25*), when India was already witnessing a staggering growth in infections. A massive humanitarian crisis unfolded that was termed a “national catastrophe”, and calls were made for an international alliance and collaboration (*26*). Healthcare infrastructure collapsed under surges in hospitalizations. The extent of this collapse was such that various parts of India suffered from acute shortages of oxygen, steroidal treatment medicine (*27*) and testing kits. Crematoriums and burial grounds were overflowing (*28, 29*). Multiple preprints and investigative reports suggest that the actual death toll far exceeds official numbers (*30, 31*). In addition to lack of timely and stringent preventive measures guided by public health, emerging variants became a large part of the conversation around India’s second wave (*32, 33*).

In many other parts of the world, winter of 2020 brought resurgent transmission, with new SARS-CoV-2 variants being identified in the UK (Alpha/B.1.1.7), Brazil (Gamma/P.1) and in South Africa (Beta/B.1.351) (*34*), concurrent with a globally changing pandemic trajectory (*35*). Many of these new variants were observed to have epidemiologically distinct changes in transmissibility as well as antigenic escape. In December 2020, the Ministry of Health and Family Welfare (MoHFW) in India launched a multi-laboratory genomic surveillance initiative formally referred to as the Indian SARS-CoV-2 Genome Sequencing Consortia (INSACOG) (*36*) to track the virus’s evolution and identify new Variants of Concern (VOC). The year 2020 was a period of comparative evolutionary stasis for SARS-CoV-2 with infections attributed to the previously circulated lineages (*37*).

The identification and global spread of other VOCs were mirrored by the identification of B.1.1.7, B.1.351, and P.1 in India (*36*). The B.1.617(.1/2/3) lineage was first detected in December 2020 in India and rapidly became a dominant lineage, particularly in Maharashtra (*38*). Between January and February 2021, the B.1.617 lineage, including Delta (B.1.617.2) and Kappa (B.1.617.1), was detected in about 60% of the 361 sequenced cases sampled in Maharashtra (*39*), and the B.1.617.2 sub-lineage was marked as a VOC in early-May by WHO (*40*). Considerable regional heterogeneity exists with respect to the dominant lineage in India. For instance, the B.1.36 lineage accounted for 43.7% of cases (November 22, 2020, to January 22, 2021) in Bengaluru, Karnataka (*41*). Over 80% of the 401 cases sampled in Punjab were attributed to the B.1.1.7 lineage (*42*). Another newly detected lineage, B.1.618, was reported to be increasingly circulating in the eastern state of West Bengal (*43*).

In this paper, we present an epidemiological analysis of the second wave of COVID-19 transmission in India. First, we compare the second wave to the first wave, nationally and across states and union territories, in terms of multiple public health metrics. Using a dynamic epidemiological model that integrates both genomic and COVID-19 mortality data, we then investigate the extent to which the emergence and altered epidemiological properties of the SARS-CoV-2 Delta variant (i.e., B.1.617.2 sub-lineage) might have driven the surge in the observed case and death counts in the second wave in India. Finally, we estimate the number of deaths that could have been averted through an early nationwide intervention (like a lockdown) at various time points in March and April 2021 during the onset of the second wave. We conclude with some recommendations moving forward.

### The second wave dwarfs the first wave in India

#### Comparison at a national level

We define the second wave as starting on February 14, 2021, the day the effective reproduction number, Rt, exceeded one (i.e., Rt > 1), indicating a growth in case counts after 4 months of steady decline. For comparative purposes, we consider June 13, 2020, as the starting point of the first wave (Wave 1), the first time the daily case counts were comparable to the daily case counts observed on February 14, 2021, the start of the second wave (Wave 2). Our analysis period ends on May 31, 2021, for the descriptive analysis. As shown in **Table 1**, Wave 2 is more severe than Wave 1 in nearly every metric of growth: higher maximum Rt, nearly 4 times higher maximum daily cases and deaths, and maximum daily test-positive rate (TPR) of 25% in Wave 2 versus 16% in Wave 1. There is growing consensus among epidemiologists that the second wave in India has had a greater severity than the first wave (*44, 45*). While there have been more cases and deaths in Wave 2, there is a lower reported case-fatality rate compared to Wave 1 (1.0% vs 1.4%). This could partly be explained by reports that younger age groups were infected more in Wave 2 where the clinical infection fatality rate is lower, but this assertion is yet to be verified (*46*). It is important to note that we do not have full follow-up data on Wave 2 and more deaths continue to be reported in June including some states like Bihar revising previously reported death numbers (*47*). There exists a systemic delay in updating death records and underreporting of case and death counts appear to be a challenge with this data (*31, 48*).

**Table 1.**
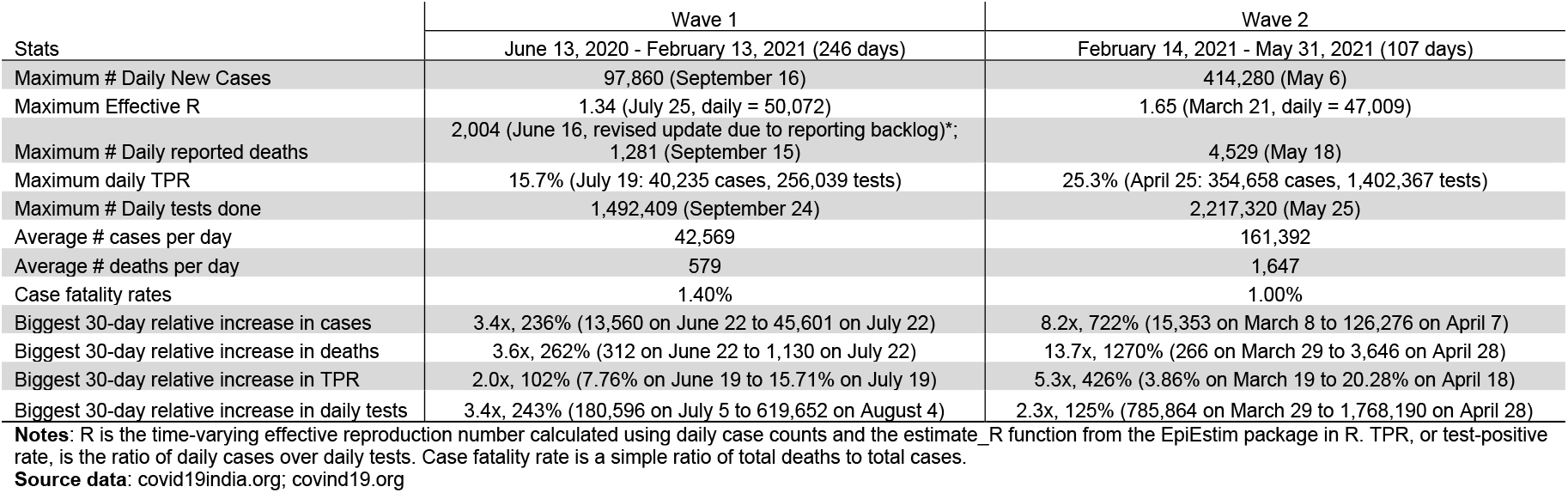
Describing COVID-19 Waves in India using selected metrics

|  | Wave 1 | Wave 2 |
| --- | --- | --- |
| Stats | June 13, 2020 - February 13, 2021 (246 days) | February 14, 2021 - May 31, 2021 (107 days) |
| Maximum # Daily New Cases | 97,860 (September 16) | 414,280 (May 6) |
| Maximum Effective R | 1.34 (July 25, daily = 50,072) | 1.65 (March 21, daily = 47,009) |
| Maximum # Daily reported deaths | 2,004 (June 16, revised update due to reporting backlog)*;<br>1,281 (September 15) | 4,529 (May 18) |
| Maximum daily TPR | 15.7% (July 19: 40,235 cases, 256,039 tests) | 25.3% (April 25: 354,658 cases, 1,402,367 tests) |
| Maximum # Daily tests done | 1,492,409 (September 24) | 2,217,320 (May 25) |
| Average # cases per day | 42,569 | 161,392 |
| Average # deaths per day | 579 | 1,647 |
| Case fatality rates | 1.40% | 1.00% |
| Biggest 30-day relative increase in cases | 3.4x, 236% (13,560 on June 22 to 45,601 on July 22) | 8.2x, 722% (15,353 on March 8 to 126,276 on April 7) |
| Biggest 30-day relative increase in deaths | 3.6x, 262% (312 on June 22 to 1,130 on July 22) | 13.7x, 1270% (266 on March 29 to 3,646 on April 28) |
| Biggest 30-day relative increase in TPR | 2.0x, 102% (7.76% on June 19 to 15.71% on July 19) | 5.3x, 426% (3.86% on March 19 to 20.28% on April 18) |
| Biggest 30-day relative increase in daily tests | 3.4x, 243% (180,596 on July 5 to 619,652 on August 4) | 2.3x, 125% (785,864 on March 29 to 1,768,190 on April 28) |

**Notes:** R is the time-varying effective reproduction number calculated using daily case counts and the estimate\_R function from the EpiEstim package in R. TPR, or test-positive rate, is the ratio of daily cases over daily tests. Case fatality rate is a simple ratio of total deaths to total cases.
**Source data:** covid19india.org; covind19.org

#### State-level comparisons

Multiple articles have now shown that there is substantial heterogeneity across Indian states and union territories, and the national data often masks this state-level variation (*12*). For this comparison, due to the differences in the lengths of our analysis periods for the waves (246 versus 107 days), we calculate standardized case and death counts by states across waves, as described in **Supplementary Material Section 1**. For example, in Wave 1, Delhi reported 599,972 cases in 246 days, yielding a standardized daily case rate of 2438.9, whereas in Wave 2, 789,444 cases were reported in a timespan of 107 days, yielding a daily case rate of 7377.9. Consequently, the ratio of daily case rates for Wave 2 versus Wave 1 in Delhi is 3.0. Only one union territory, namely the Andaman and Nicobar Islands, reports a less-than-unity ratio of 0.9, indicating a less severe Wave 2 when compared to Wave 1. All other states and union territories return ratios that are greater than unity, suggesting a more severe Wave 2. Across all states and union territories, the median ratio of daily case rate for Wave 2 versus Wave 1 is 3.8. The states with the four highest case intensity ratios are (in decreasing order): Uttarakhand (5.6), Himachal Pradesh (5.3), Punjab (5.2) and Gujarat (5.2). These findings are reflected in **Figure 1A**.

**Figure 1.**
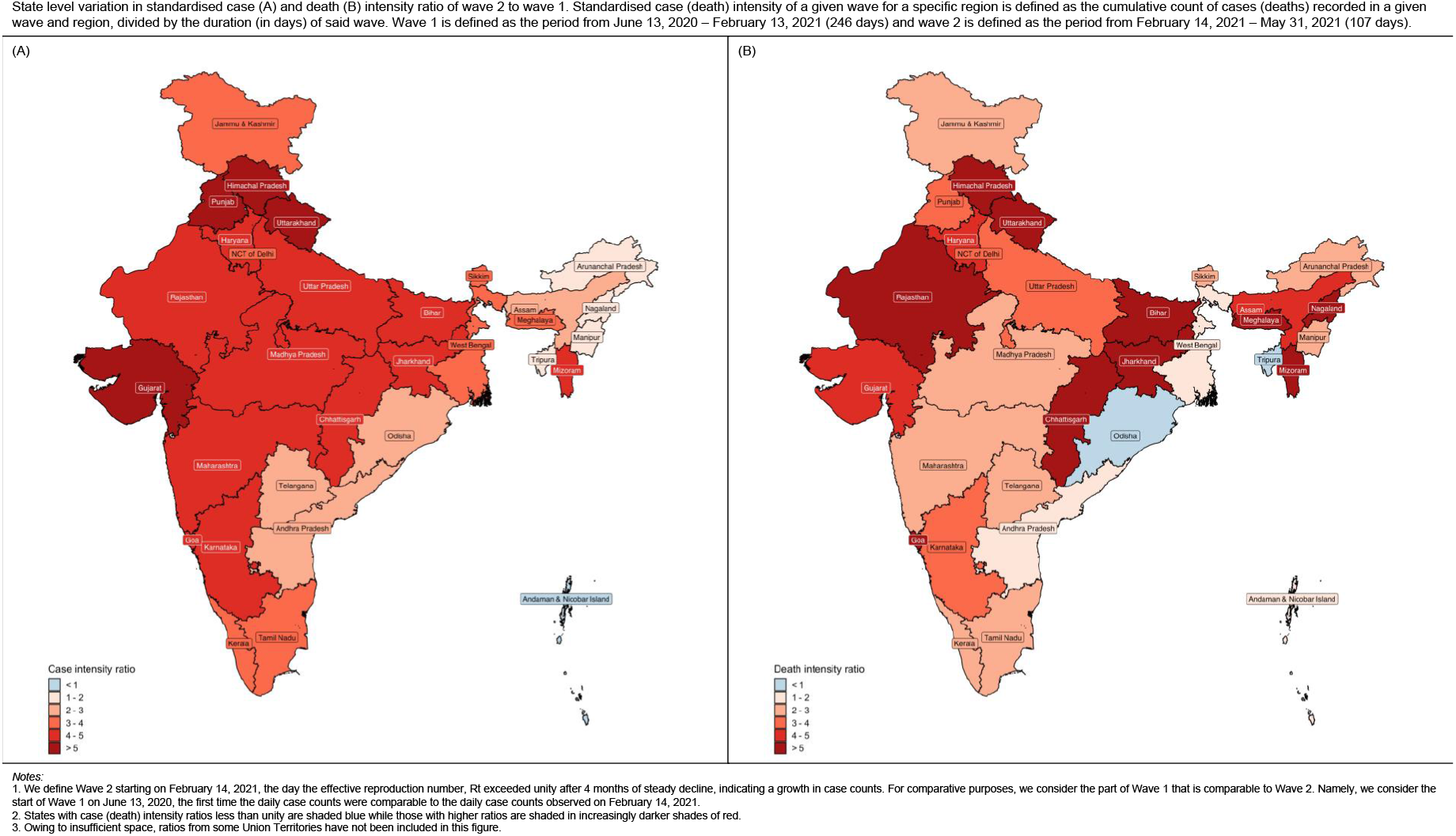
State level variation in standardized case (A) and death (B) rate ratio of Wave 2 to Wave 1. Standardized case (death) rate of a given wave for a specific region is defined as the cumulative count of cases (deaths) recorded in a given wave and region, divided by the duration (in days) of said wave. Wave 1 is defined as the period from June 13, 2020 – February 13, 2021 (246 days) and Wave 2 is defined as the period from February 14, 2021 – May 31, 2021 (107 days).

Following a similar approach for death counts, Delhi reported 9,675 deaths in Wave 1 over 246 days, yielding a standardized death intensity of 39.3. In Wave 2, there were 13,348 deaths reported over 107 days in Delhi, yielding a standardized death rate of 124.8. Consequently, the ratio of daily death rates for Wave 2 versus Wave 1 in Delhi is 3.2. Two states report ratios that are less than unity (Tripura 0.7 and Odisha 0.99), indicating a less severe Wave 2 when compared to Wave 1, while all other states and union territories return ratios that are greater than unity. Across all other states and union territories, the median death rate ratio for Wave 2 versus Wave 1 is 3.2. The states with the five highest death count intensity ratios are (in decreasing order): Jharkhand (8.4), Mizoram (7.9), Nagaland (7.2), Meghalaya (6.7) and Uttarakhand (6.6). These findings are reflected in **Figure 1B**. The exact numerical values are provided in **Table S1**.

#### State-level comparison of movement of the virus and peaks across Waves 1 and 2

The national peak occurred on September 16, 2020, with 97,860 daily cases during Wave 1 and on May 6, 2021, with 414,280 daily cases during Wave 2. However, we see significant variation in the timing of the peaks across states in both waves. For example, Wave 1 peaks in 2020 occurred as early as July 27 for Tamil Nadu and as late as November 29 for Himachal Pradesh. Similarly, Wave 2 peaks ranged from April 17, 2021 (Ladakh) through May 30, 2021 (Mizoram). Only 9 of the 36 states and union territories experienced a higher proportion of **total cases** in Wave 1 than in Wave 2 until May 31. Deaths are a lagged metric of infections, and we see the national peak of daily new deaths in Wave 2 was attained on May 18 with 4,529 new deaths reported, nearly 4 times the peak in Wave 1 (1,281 deaths). The peaks for daily number of deaths across states took place from the end of April to the end of May in 2021 for Wave 2 while they were dispersed from July to November of 2020 for Wave 1. Even with a limited follow-up time, twenty-eight states had already seen more deaths in Wave 2 than Wave 1. The timeline for the sequence of peaks across Indian states is presented in **Supplementary Table S1a** and **S1b**.

### New virus variants explain part of the Wave 2 surge

As shown in **Figure 1** and **Table S1** there is considerable spatial heterogeneity in infection burden and fatalities across states and union territories in India across the waves. The period just before and around the second wave has seen the introduction of multiple VOCs, and variants under investigation in India (*49, 50*). This period is marked by a rapid increase in the infection share of B.1.617.2 which has, as of late April 2021 (*51*), become the dominant strain (>99% of all sequenced genomes). A detailed literature review of the variant landscape in India is presented in **Table S2**.

We explore epidemiological factors underlying the resurgence of transmission observed across India, with a specific focus on the SARS-CoV-2 VOC B.1.617.2 (Delta variant under the WHO nomenclature (*34, 52*)) as a driver of the second wave. Recent *in vitro* work shows B.1.617.2 is associated with significantly reduced antibody neutralization (with reductions similar to those observed for the Beta VOC B.1.351) (*53*), whilst analysis of secondary attack rates in UK households have suggested a transmission advantage of B.1.617.2 over the Alpha VOC B.1.1.7 (*54*). We restrict our analyses to the state of Maharashtra which has had the largest COVID-19 burden to date in India and where detailed epidemiological and genomic data required to assess the contribution of B.1.617.2 to resurgence is available. **Table S3** and **Figure S1** provide a temporal distribution of this variant in Maharashtra, where the proportion of B.1.617.2 samples rose from 1.5% in February 2021 to 87% in May 2021.

We adapt a previous two-strain epidemiological model developed by Faria et al. (*55*) to explore the extent and nature of alterations to B.1.617.2’s characteristics (such as transmissibility, severity and ability to evade immunity elicited by prior natural infection) that are required to explain the extensive resurgence of transmission observed during the second wave. Within this framework (*55*), we explicitly include the possibility for other competing hypotheses to explain this resurgence, such as increased transmissibility or immune evasion capabilities for the new variant B.1.617.2, as well as waning of prior immunity from infections occurring during the first wave. The framework essentially fuses mortality data with genomic distribution data from GISAID. The mortality data is used in a renewal framework (*56, 57*) by linking it to the infections. The genomic data is used to estimate the fraction of infections for individual strains, i.e., B.1617.2 and non-B.1617.2.

In the absence of up-to-date and representative state-wide serological surveys enabling accurate ascertainment of the infection fatality ratio (IFR) across Maharashtra and the uncertainty surrounding the extent to which COVID-19 deaths might be missed due to testing constraints, we run a suite of different scenarios, exploring the extent and nature of the inferred changes to B.1.617.2’s epidemiological properties. We assume the introduction date for B.1617.2 variant as 1^st^ Dec 2020, as the first sequence for B.1.617.2 in GISAID for Maharashtra is dated 10^th^ Dec 2020 (*50*). The results presented in the main text are for an assumed IFR of 0.25% (*31*) and an under-reporting factor of 50% (suggesting 1 in 2 deaths are missed due to testing and other logistical constraints which is a very conservative estimate based on the literature (*31*)); results from a sensitivity analysis comprising eleven other scenarios are summarized in **Table S4**.

**Figure 2 (left column)** shows the fit for the model where under-reporting is 50% along with IFR 0.25% and protection immunity of 100%. The fraction of deaths attributed by the model to the B.1617.2 variant from April 1-May 15 is 55% [10%-90%] (95% Bayesian Credible Interval [CI]). However, this number varies from being 15% [5%-45%] on 1^st^ April to 83% [62%-96%] on 15^th^ May 2021. Furthermore, results shown in **Figure 2 (right column)** suggest the epidemiological characteristics of B.1.617.2 are different from those of previously circulating SARS-CoV-2 lineages (see **Table S2** for a list) - across the three plausible values of cross-infection (100% - no immunity escape; 75% - moderate possibility of reinfection; 50% - higher possibility of reinfection), our results consistently support the hypothesis of B.1.617.2 as being more transmissible than all previously circulating lineages. As seen in **Figure 2**, under scenarios of highest cross-immunity (100%, meaning previous infection with **non**-B.1.617.2 lineages provides robust protection to B.1.617.2 infection except in those individuals where immunity has waned), levels of population immunity to B.1.617.2 are higher and so a greater transmissibility increase, 83% [77%-90%], is required to generate the observed second wave. Conversely, when the degree of cross-immunity is lowest (50%), more individuals are available for infection with B.1.617.2 (including a fraction of those previously infected with non-B.1.617.2 lineages), and so a smaller increase in transmissibility, 49% [44%-55%] is required to explain the observed second wave. Nonetheless, higher transmissibility is indicated across all scenarios.

**Figure 2.**
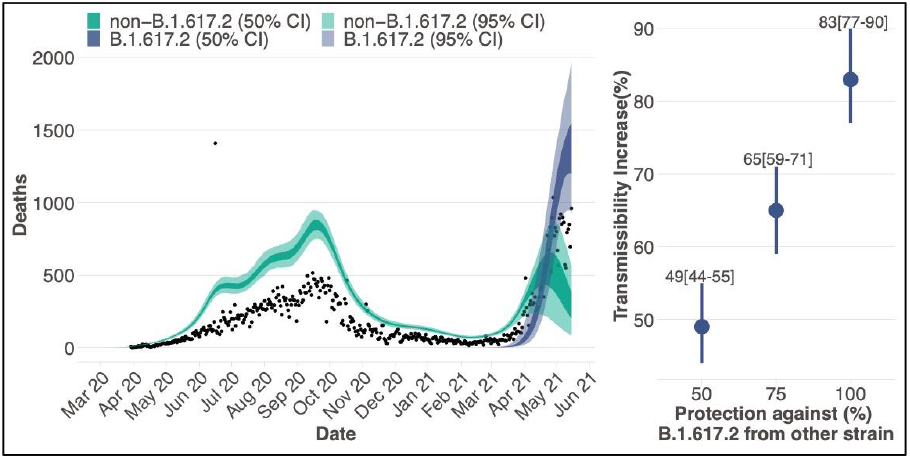
Estimates of the epidemiological characteristic of B.1.617.2 inferred from a multicategory Bayesian transmission model fitted to mortality and genomic data from Maharashtra, India. *Left column:* Daily incidence of COVID-19 mortality with black dots showing the observed data; the blue and green shaded region represent the 95% (50% with darker shades) Bayesian credible interval for estimated daily deaths with an assumed under-reporting factor of 50% disaggregated by non-B.1.617.2 and B.1.617.2 deaths estimated by a model assuming 100% protection. *Right column:* 95% Bayesian posterior credible intervals of the transmissibility increase against various fixed values of protection against infection from B.1.617.2 if earlier infected by another strain.

However, these results remain highly uncertain, particularly given the absence of reliable serological surveys to infer cumulative population infections to date and the IFR. Whilst we cannot quantify the exact extent and nature of the changes, our results highlight that immunity waning or relaxation of NPIs alone cannot explain the large second wave in Maharashtra. Instead, the results suggest that this was at least partially driven by the emergence of a variant with altered epidemiological properties. We would like to point out that our two-strain model doesn’t account for NPIs explicitly but observed changes to the reproduction number as a consequence of the NPIs are modelled as a 7-day random walk (*55*). We note that, our estimates of altered epidemiological properties for B.1.617.2 are qualitatively robust under various conservative scenarios of lower IFR and no under-reporting, though the numerical values change. For example, in a scenario with a low IFR of 0.15% (*58*), no under-reporting and 100% protection, the estimates of transmissibility increase are 82% [76-88%], which is aligned with the results in the corresponding result in the main text of 83% [77%-90%] transmissibility increase. For results under all twelve scenarios refer **Table S4**.

### The resurgent Wave 2 could have been prevented through early public health interventions

Could timely nationwide NPIs have mitigated the speed of Wave 2 in India? A summary of COVID-19-related public health interventions that were implemented in India from March 3, 2020, through May 31, 2021, is provided in **Table S5**. Using evidence from previously observed lockdown effects in India (nationwide, beginning March 25, 2020) and in Maharashtra (beginning April 14, 2021), we can assess what would have happened had an intervention with a similar effect taking place across India at various timepoints in March and April of 2021. Because of the observed strength of the 2020 national lockdown, we use it as a “strong lockdown effect” intervention schedule, while the 2021 Maharashtra lockdown represents a “moderate lockdown effect” intervention schedule. A detailed description of the predictive model used and how the modifying intervention effect schedules were elicited from real data based on rates of growth/decline in case counts over time are described in **Supplementary Material Section 3**.

#### Intervention effect on cases

As one would expect, implementing a lockdown during an optimal time window has short- and long-term benefits with respect to the degree of reduction in reported case counts. This result is consistent regardless of whether a strong lockdown effect (**Figure 3a**) or a moderate lockdown effect (**Figure 3b**) is used. Indeed, had a lockdown been instituted in mid-to late-March of 2021, the case counts would have almost immediately started to decline, with the daily case count peaking around 20 thousand and 49 thousand, respectively (instead of the observed 414,280). To put this in perspective, with a *moderate* lockdown starting on March 15, approximately 2.6 million cases (95% CI: [0.3, 2.9]) (**bolded** in **Table 2**) could have been avoided through April 15, representing a 91.6% (95% CI: [10.1%, 100.0%]) reduction in total case counts compared to observed numbers. We demonstrate that these reductions in the number of infected cases would continue to grow through May 15, resulting in the prevention of 12.9 million infections (95% CI: [9.5, 13.3]), a 97.0% reduction (95% CI: [71.7%, 100.0%]); (**bolded** in **Table 2**). Interestingly, a lockdown with *strong* effect beginning March 15 would also result in essentially the same number of infections being prevented (12.9 million with moderate effect vs 13.0 million with strong effect) by May 15 (98.2% reduction in cases, 95% CI: [73.7%, 100.0%]). This suggests that the timing of the intervention matters, and March 15-30 would have been the optimal time window for intervening.

**Figure 3.**
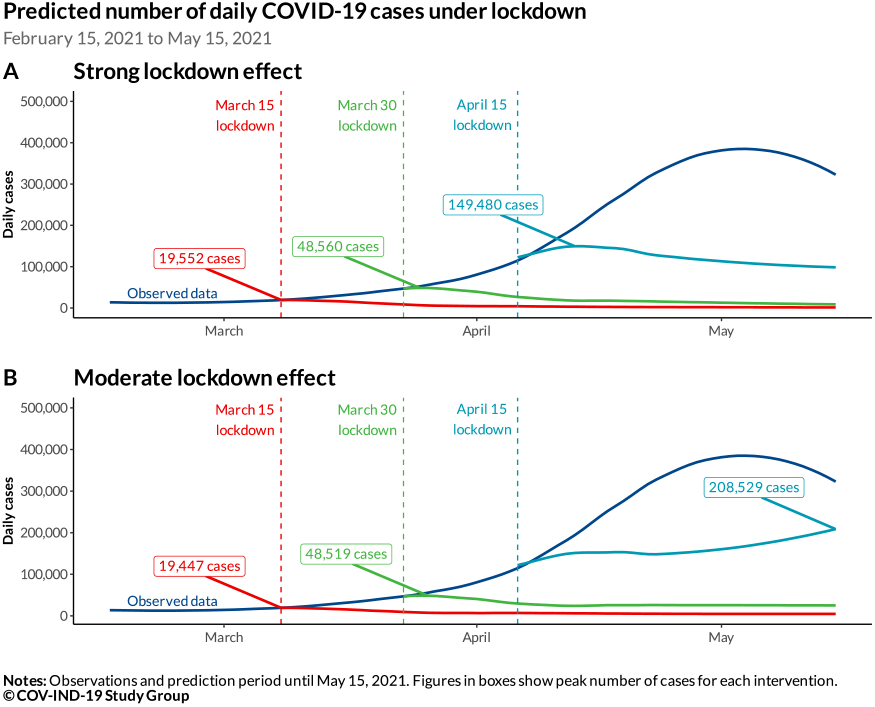
Observed and predicted daily cases under lockdown scenarios starting on different dates using the strong (**panel A**) and moderate (**panel B**) lockdown effect schedules from February 15 to May 15, 2021. Effect of lockdown drawn from relative reduction in time-varying effective reproduction number (R_t_) from India and Maharashtra after the implementation of their COVID-19 interventions in March 2020 and April 2021, respectively. The lockdown effect schedules are then smoothed using a LOESS smoother to account for day-to-day variations in R_t_. Three lockdown start dates are depicted: March 15 (red), March 30 (green), and April 15 (light blue). Peak daily case counts under lockdown scenarios are shown.

**Table 2.** Predicted case counts, cases averted and % reduction with corresponding 95% credible intervals under different lockdown interventions (in millions)

| Evaluation date | Metrics | Strong lockdown effect start date |  |  |  | Moderate lockdown effect start date |  |  |  |
| --- | --- | --- | --- | --- | --- | --- | --- | --- | --- |
|  |  | March 1 | March 15 | March 30 | April 15 | March 1 | March 15 | March 30 | April 15 |
| 3/15/21 | Observed | 0.3 | - | - | - | 0.3 | - | - | - |
|  | Predicted | 0.1 [0.0, 1.6] |  |  |  | 0.1 [0.0, 1.7] |  |  |  |
|  | Averted | 0.2 [-1.4, 0.3] |  |  |  | 0.2 [-1.4, 0.3] |  |  |  |
|  | % Reduction | 68.9% [-477.3%, 100.0%] |  |  |  | 65.9% [-480.7%, 100.0%] |  |  |  |
| 3/30/21 | Observed | 1.0 | 0.7 | - | - | 1.0 | 0.7 | - | - |
|  | Predicted | 0.1 [0.0, 2.3] | 0.1 [0.0, 1.7] |  |  | 0.2 [0.0, 2.4] | 0.1 [0.0, 1.8] |  |  |
|  | Averted | 0.9 [-1.3, 1.0] | 0.6 [-1.0, 0.7] |  |  | 0.9 [-1.4, 1.0] | 0.6 [-1.0, 0.7] |  |  |
|  | % Reduction | 86.4% [-128.6%, 100.0%] | 84.1% [-136.0%, 100.0%] |  |  | 83.0% [-133.4%, 100.0%] | 81.9% [-139.3%, 100.0%] |  |  |
| 4/15/21 | Observed | 3.2 | 2.9 | 2.1 | - | 3.2 | 2.9 | 2.1 | - |
|  | Predicted | 0.2 [0.0, 2.9] | 0.2 [0.0, 2.5] | 0.4 [0.0, 2.3] |  | 0.2 [0.0, 3.0] | 0.2 [0.0, 2.6] | 0.4 [0.0, 2.4] |  |
|  | Averted | 3.0 [0.3, 3.2] | 2.7 [0.4, 2.9] | 1.8 [-0.1, 2.1] |  | 2.9 [0.1, 3.2] | 2.6 [0.3, 2.9] | 1.7 [-0.2, 2.1] |  |
|  | % Reduction | 94.7% [8.2%, 100.0%] | 93.7% [12.9%, 100.0%] | 81.9% [-5.2%, 100.0%] |  | 92.3% [4.2%, 100.0%] | 91.6% [10.1%, 100.0%] | 79.1% [-11.2%, 100.0%] |  |
| 4/30/21 | Observed | 8.0 | 7.7 | 7.0 | 4.9 | 8.0 | 7.7 | 7.0 | 4.9 |
|  | Predicted | 0.2 [0.0, 3.3] | 0.2 [0.0, 3.0] | 0.6 [0.0, 3.3] | 1.9 [0.0, 4.6] | 0.3 [0.0, 3.5] | 0.3 [0.0, 3.2] | 0.8 [0.0, 3.7] | 2.3 [0.0, 5.2] |
|  | Averted | 7.9 [4.7, 8.0] | 7.5 [4.7, 7.7] | 6.4 [3.7, 7.0] | 2.9 [0.3, 4.9] | 7.7 [4.5, 8.0] | 7.4 [4.5, 7.7] | 6.2 [3.3, 7.0] | 2.6 [-0.3, 4.9] |
|  | % Reduction | 97.7% [58.3%, 100.0%] | 97.2% [60.7%, 100.0%] | 91.1% [53.3%, 100.0%] | 60.4% [6.2%, 100.0%] | 96.4% [56.1%, 100.0%] | 95.9% [58.5%, 100.0%] | 88.1% [47.0%, 100.0%] | 53.6% [-6.5%, 100.0%] |
| 5/15/21 | Observed | 13.6 | 13.3 | 12.5 | 10.4 | 13.6 | 13.3 | 12.5 | 10.4 |
|  | Predicted | 0.2 [0.0, 3.7] | 0.2 [0.0, 3.5] | 0.8 [0.0, 4.0] | 3.5 [0.0, 7.8] | 0.3 [0.0, 4.0] | 0.4 [0.0, 3.8] | 1.2 [0.0, 5.0] | 5.0 [0.5, 10.9] |
|  | Averted | 13.4 [9.8, 13.6] | 13.0 [9.8, 13.3] | 11.7 [8.5, 12.5] | 6.9 [2.6, 10.4] | 13.2 [9.6, 13.6] | 12.9 [9.5, 13.3] | 11.3 [7.5, 12.5] | 5.4 [-0.5, 9.9] |
|  | % Reduction | 98.6% [72.4%, 100.0%] | 98.2% [73.7%, 100.0%] | 93.7% [67.7%, 100.0%] | 66.2% [25.3%, 100.0%] | 97.6% [70.6%, 100.0%] | 97.0% [71.7%, 100.0%] | 90.3% [59.8%, 100.0%] | 51.9% [-5.0%, 95.3%] |

**Notes:** For lockdown schedules, each cell reports (1) the total number of observed cases since the start of lockdown in the first row, (2) total number of predicted cases since the start of lockdown in the second row (with 95% CI), (3) the number of cases averted (relative to observed) since the start of lockdown in the third row (with 95% CI), and (4) the relative reduction in cases (as a percent) under lockdown in the fourth row (with 95% CI). Cells that are highlighted in red represent a statistically significant reduction in the number of cases under lockdown at the 95% credible interval level. Cells that are bolded are emphasized in the main text. Numbers are reported in millions.

For lockdowns beginning in mid-April, the benefits are not as pronounced. Our findings indicate that a peak would have occurred shortly after a lockdown with a strong effect with around 150,000 cases at the peak. However, under a lockdown with moderate effect, a near term peak is not obvious and the peak is attained at around 209,000 daily new cases (**Figure 3A and 3B**); it would have induced a reduction (approximately 51.9% or 5.4 million cases) in total case counts from April 15 to May 15, 2021, but the 95% CI [-0.5, 9.9] includes the zero value so the decrease is not statistically significant. An end-of-April national lockdown would have been too late for any marked benefit because the second wave outbreak had already run its course (largely aided by state-level lockdowns in Maharashtra and Delhi (*59*)), as is evident by the slowing growth and eventual decline of observed daily cases after May 6 (*14*).

#### Intervention effect on deaths

To estimate the number of preventable deaths we multiply the predicted cases under each intervention effect with daily CFR estimates obtained under three different scenarios. Details are presented in **Supplementary Section 3**. Three daily CFR schedules, based on observed data from Kerala, India, and Maharashtra on exactly the same dates, are applied to daily predicted case counts. For simplicity, we refer to these as low, moderate, and high CFR schedules, respectively. A similar pattern as the case counts can be seen with respect to death counts. Clear benefits from lockdowns beginning in mid- and late-March are present under all CFR scenarios through mid-May (**Figure 4**). Under the *moderate* intervention effect, our estimates show roughly 97 to 109 thousand deaths could have been avoided by May 15, 2021 (**Table 3**), a reduction of 89.6-98.5% from the observed 112 thousand deaths observed from March 15-May 15, under all CFR schedules. Regardless of strictness and timing of the lockdown (**Figure 4C**), if there is sufficient healthcare capacity (i.e., low CFR scenario), we see that a lockdown would have remarkable benefits in terms of reducing death counts. For example, we see that a March 15 lockdown would have resulted in a 98.5% [95% CI: 85.2%, 100.0%] (109.7 thousand [95% CI: 94.9, 111.4] deaths averted) reduction by May 15 (**Table 3**). Even with a very late lockdown on April 15, in the low CFR situation 91.6% [95% CI: 81.6%, 99.4%] of the deaths could have been averted by May 15 while this proportion is 55.1%% [95% CI: 0.9%, 95.7%] for the moderate CFR and 37.9% [95% CI: -37.9%, 94.5%] for the high CFR scenario.

**Figure 4.**
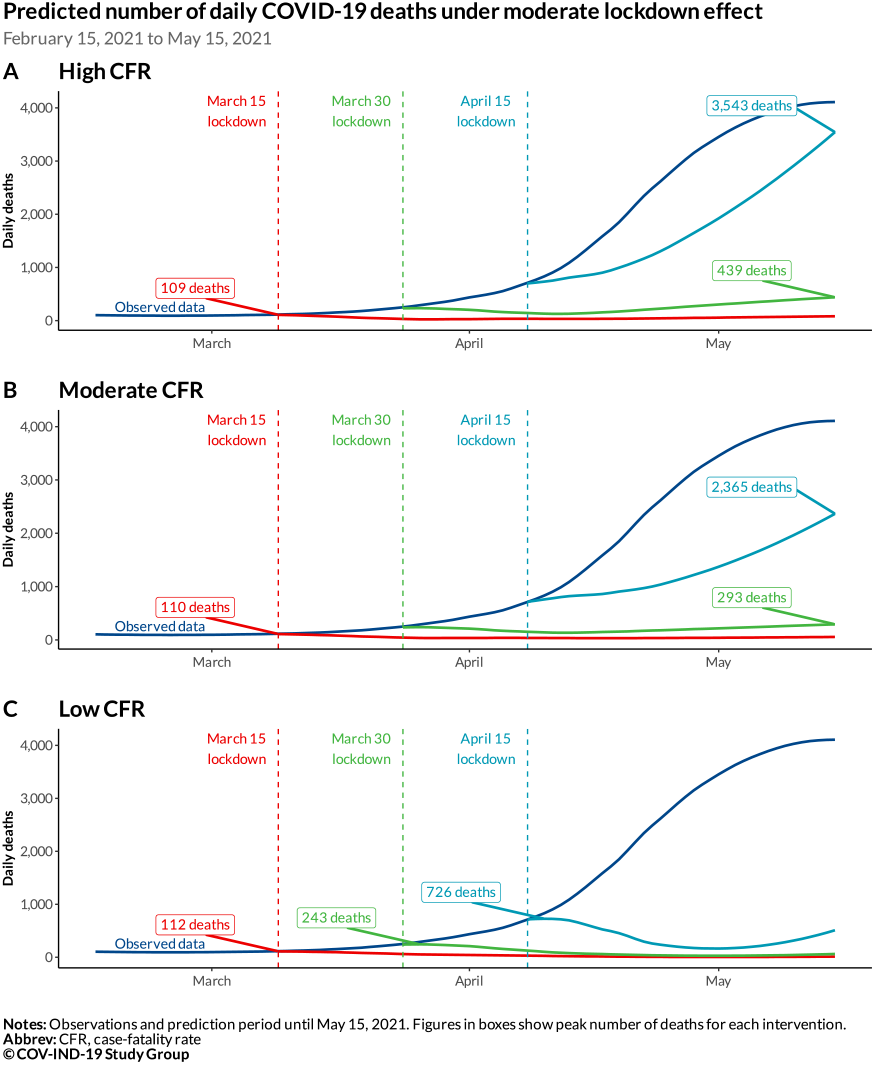
Observed and predicted daily deaths under lockdown with strong effect starting on different dates using high, moderate, and low CFR schedules from February 15 to May 15, 2021. Effect of lockdown drawn from relative reduction in time-varying effective reproduction number from Maharashtra after the implementation of interventions in April 2021. High, moderate, and low CFR schedules correspond to trailing 7-day daily CFR schedules from Maharashtra, India, and Kerala, respectively. The daily death estimates represent the estimated daily infection count multiplied by the respective CFR schedule. Three lockdown start dates are depicted: March 15 (red), March 30 (green), and April 15 (light blue). Peak daily death counts under lockdown scenarios are shown.

**Table 3.** Predicted total deaths counts, deaths averted and % reduction with corresponding 95% credible intervals under lockdown with moderate effect interventions (in thousands)

| Evaluation date | Metrics | Intervention Start Date (High CFR) |  |  |  | Intervention Start Date (Moderate CFR) |  |  |  | Intervention Start Date (Low CFR) |  |  |  |
| --- | --- | --- | --- | --- | --- | --- | --- | --- | --- | --- | --- | --- | --- |
|  |  | March 1 | March 15 | March 30 | April 15 | March 1 | March 15 | March 30 | April 15 | March 1 | March 15 | March 30 | April 15 |
| 3/15/21 | *Observed* | 1.6 | - | - | - | 1.6 | - | - | - | 1.6 | - | - | - |
|  | *Predicted* | 0.5 [0.0, 8.7] |  |  |  | 0.6 [0.0, 10.3] |  |  |  | 0.6 [0.0, 9.8] |  |  |  |
|  | *Averted* | 1.1 [-7.1, 1.6] |  |  |  | 1.0 [-8.7, 1.6] |  |  |  | 1.0 [-8.2, 1.6] |  |  |  |
|  | *% Reduction* | 68.7% [-439.8%, 100.0%] |  |  |  | 62.5% [-540.1%, 100.0%] |  |  |  | 64.5% [-513.0%, 100.0%] |  |  |  |
| 3/30/21 | *Observed* | 5.2 | 3.6 | - | - | 5.2 | 3.6 | - | - | 5.2 | 3.6 | - | - |
|  | *Predicted* | 0.8 [0.0, 10.6] | 0.5 [0.0, 6.2] |  |  | 1.0 [0.0, 13.5] | 0.7 [0.0, 8.9] |  |  | 1.1 [0.0, 15.7] | 1.0 [0.0, 12.7] |  |  |
|  | *Averted* | 4.4 [-5.4, 5.2] | 3.1 [-2.6, 3.6] |  |  | 4.2 [-8.2, 5.2] | 2.9 [-5.3, 3.6] |  |  | 4.1 [-10.5, 5.2] | 2.7 [-9.1, 3.6] |  |  |
|  | *% Reduction* | 85.3% [-103.6%, 100.0%] | 87.1% [-72.7%, 100.0%] |  |  | 81.2% [-157.9%, 100.0%] | 81.5% [-146.7%, 100.0%] |  |  | 78.5% [-200.8%, 100.0%] | 73.4% [-252.3%, 100.0%] |  |  |
| 4/15/21 | *Observed* | 17.1 | 15.4 | 11.8 | - | 17.1 | 15.4 | 11.8 | - | 17.1 | 15.4 | 11.8 | - |
|  | *Predicted* | 1.1 [0.0, 14.3] | 1.0 [0.0, 11.6] | 2.3 [0.0, 12.2] |  | 1.4 [0.0, 17.0] | 1.3 [0.0, 13.7] | 2.5 [0.0, 13.1] |  | 1.4 [0.0, 18.0] | 1.5 [0.0, 15.7] | 2.1 [0.0, 11.3] |  |
|  | *Averted* | 15.9 [2.7, 17.1] | 14.4 [3.9, 15.4] | 9.5 [-0.3, 11.8] |  | 15.7 [0.1, 17.1] | 14.2 [1.7, 15.4] | 9.4 [-1.3, 11.8] |  | 15.6 [-1.0, 17.1] | 14.0 [-0.2, 15.4] | 9.8 [0.5, 11.8] |  |
|  | *% Reduction* | 93.4% [16.0%, 100.0%] | 93.3% [25.2%, 100.0%] | 80.6% [-3.0%, 100.0%] |  | 92.0% [0.4%, 100.0%] | 91.7% [11.1%, 100.0%] | 79.1% [-10.9%, 100.0%] |  | 91.6% [-5.8%, 100.0%] | 90.5% [-1.4%, 100.0%] | 82.4% [4.2%, 100.0%] |  |
| 4/30/21 | *Observed* | 54.6 | 52.9 | 49.3 | 37.5 | 54.6 | 52.9 | 49.3 | 37.5 | 54.6 | 52.9 | 49.3 | 37.5 |
|  | *Predicted* | 1.5 [0.0, 20.9] | 1.7 [0.0, 19.6] | 5.6 [0.0, 25.7] | 19.0 [0.0, 44.1] | 1.7 [0.0, 21.0] | 1.8 [0.0, 18.8] | 5.1 [0.0, 23.0] | 15.4 [0.0, 35.4] | 1.5 [0.0, 18.4] | 1.6 [0.0, 16.1] | 2.7 [0.0, 12.8] | 3.5 [0.0, 8.3] |
|  | *Averted* | 53.1 [33.6, 54.6] | 51.3 [33.4, 52.9] | 43.8 [23.6, 49.3] | 18.5 [-6.6, 37.5] | 52.9 [33.5, 54.6] | 51.1 [34.1, 52.9] | 44.2 [26.4, 49.3] | 22.1 [2.1, 37.5] | 53.0 [36.2, 54.6] | 51.4 [36.8, 52.9] | 46.7 [36.5, 49.3] | 34.0 [29.2, 37.5] |
|  | *% Reduction* | 97.3% [61.6%, 100.0%] | 96.8% [63.0%, 100.0%] | 88.7% [47.8%, 100.0%] | 49.2% [-17.5%, 100.0%] | 96.9% [61.4%, 100.0%] | 96.6% [64.4%, 100.0%] | 89.6% [53.5%, 100.0%] | 58.9% [5.5%, 100.0%] | 97.2% [66.3%, 100.0%] | 97.0% [69.6%, 100.0%] | 94.6% [74.0%, 100.0%] | 90.6% [77.8%, 100.0%] |
| 5/15/21 | *Observed* | 113.0 | 111.4 | 107.8 | 96.0 | 113.0 | 111.4 | 107.8 | 96.0 | 113.0 | 111.4 | 107.8 | 96.0 |
|  | *Predicted* | 2.1 [0.0, 33.0] | 2.7 [0.0, 33.1] | 11.2 [0.0, 49.8] | 59.6 [5.3, 132.4] | 2.1 [0.0, 27.5] | 2.5 [0.0, 26.5] | 9.0 [0.0, 38.3] | 43.1 [4.1, 95.2] | 1.6 [0.0, 18.7] | 1.7 [0.0, 16.5] | 3.3 [0.0, 14.5] | 8.1 [0.6, 17.7] |
|  | *Averted* | 110.9 [80.0, 113.0] | 108.7 [78.3, 111.4] | 96.6 [58.1, 107.8] | 36.4 [-36.4, 90.7] | 111.0 [85.5, 113.0] | 108.9 [84.9, 111.4] | 98.9 [69.5, 107.8] | 52.9 [0.8, 91.8] | 111.5 [94.3, 113.0] | 109.7 [94.9, 111.4] | 104.5 [93.3, 107.8] | 87.9 [78.3, 95.4] |
|  | *% Reduction* | 98.2% [70.8%, 100.0%] | 97.5% [70.3%, 100.0%] | 89.6% [53.9%, 100.0%] | 37.9% [-37.9%, 94.5%] | 98.2% [75.6%, 100.0%] | 97.7% [76.2%, 100.0%] | 91.7% [64.4%, 100.0%] | 55.1% [0.9%, 95.7%] | 98.6% [83.4%, 100.0%] | 98.5% [85.2%, 100.0%] | 96.9% [86.6%, 100.0%] | 91.6% [81.6%, 99.4%] |

**Notes:** For lockdown schedules, each cell reports (1) the total number of observed deaths since the start of lockdown in the first row, (2) total number of predicted deaths since the start of lockdown in the second row (with 95% CI), (3) the number of deaths averted (relative to observed) since the start of lockdown in the third row (with 95% CI), and (4) the relative reduction in deaths (as a percent) under lockdown in the fourth row (with 95% CI). Cells that are highlighted in red represent a statistically significant reduction in the number of cases under lockdown at the 95% credible interval level. Cells that are bolded are emphasized in the main text. Numbers are reported in millions.

Projected deaths under the three CFR schedules under a lockdown with *strong* effect are presented in **Figure S3**. This shows slightly stronger effects suggesting a mid-late March lockdown could have prevented 94-99% of deaths by May 15. The analyses conducted using a model incorporating waning immunity are presented in **Supplementary Section S3.1**. Because the results are nearly indistinguishable from the results using a model without waning immunity (e.g., under strong intervention scenario, it projects 102-111 thousand deaths could be averted by a mid-to-late March lockdown regardless of intervention compared to 97-110 thousand deaths in the model without waning immunity), we present the results without waning immunity in the main text as it is a simpler model.

To summarize, had action taken place at any time in March, it is plausible that more than 90% of observed cases and deaths between March 1-May 15 could potentially be avoided under both strong and moderate intervention scenarios.

## DISCUSSION AND CONCLUSIONS

By virtually every metric, Wave 2 of the COVID-19 pandemic in India has been more acute and devastating than Wave 1. Owing to the development of COVID-19 vaccines, treatments and care, and the fact that variants of the virus have not yet demonstrated marked increases in lethality, the observed case fatality rate appears to be lower in Wave 2 than in Wave 1 (*31*). However, with daily infections growing exponentially, even with a small infection fatality rate, a large number of deaths are bound to occur, partly due to the natural infection related mortality but also due to the collapse of the healthcare system. We see a tragic example of this in India’s Wave 2. Despite the early and decisive actions taken in Wave 1, strong interventions were not enforced in Wave 2. Our analysis shows that a large fraction of cases and deaths could potentially be averted with early comprehensive nationwide interventions, with realistic data-driven effect sizes that are derived from lockdowns that took place in India.

### Study Limitations

We acknowledge there are certain overarching limitations in our work. First, under-reporting of cases and deaths attributed to COVID-19 is not aptly accounted for across these results. At the time of this report, officials have reported intermittent excess death calculations for selected cities (*60–62*) and for even fewer states in India, and hence, adjusting for under-reporting remains a challenge (*62*). All reports point to a large degree of underreporting, particularly in rural India (*48*). Similar considerations apply for infections. While we characterize effects of NPI on reported cases this only captures a small fraction of infections. The more recent serosurveys that are emerging indicate 55% seropositivity in age-groups 0-17 and 63% in adults in urban areas (*63*). Second, the lack of disaggregation of cases and deaths attributed to COVID-19 in the data available at the time of this study prohibits any investigation into differences within age-sex strata and identify vulnerable, underserved subgroups. This limits our ability to interpret some of the observations. For example, the apparent overall lower infection fatality rate in Wave 2 can be largely due to younger people getting infected. An age-specific fatality comparison is necessary but could not be performed. Third, our models do not incorporate vaccine roll-out. For further context, during the Wave 2 analysis period, about 3% of India was fully vaccinated and 10% received at least one dose (based on vaccine data available from covid19india.org (*14*) through May 15, 2021). Since age and occupation was at large the determining factor in vaccine eligibility, accounting for vaccine distribution during this period also requires age-stratified data which is not currently available in India. Fourth, our assessment period for the effect of intervention contains the period April 15-May 15 where many states in fact instituted partial lockdowns. Thus, the comparison with the observed data for this period is not with a no-intervention scenario but with the actual ground reality with a mix of mitigation strategies being compared to the idealized hypothetical lockdown effect. The comparison up to April 15 is clearer to interpret. Finally, we had to restrict our exploration of the epidemiologic properties of B.1.617.2 to the state of Maharashtra as the nationwide temporal distribution of this variant that is available in the GISAID database does not have adequate coverage of all Indian states. Amongst other states with genomic data, we chose Maharashtra not just because it contributes the largest share to India’s COVID case and death burden, but also because it instituted the earliest statewide lockdown (April 14, 2021). If we see a high transmissibility despite a statewide NPI, it provides stronger evidence in support of the hypothesis. However, we do acknowledge as shown in **Supplementary Figure S8 a**,**b**, growth of SARS-Cov-2 variants in India has very large spatiotemporal heterogeneity between March 1-May 15, 2021 (*64, 65*). A detailed future analysis is required to really capture this heterogeneity once reliable genomic, mortality and seroprevalence data is available for the second wave.

Relatedly, our inferences about the extent and nature of changes to B.1.617.2’s epidemiological properties remain highly uncertain. Though we mitigate these issues somewhat by running a suite of sensitivity analyses that consistently support altered epidemiological properties as being different to previously circulating lineages (even if uncertainty remains about the degree to which they have changed), our results underscore the urgent need for more detailed collection of genomic and epidemiological data (including mortality, estimates of underreporting and the carrying out of further serological surveys). An important assumption made in our analyses is that the genomic data (taken from GISAID) represent a random sample that accurately reflects the nature of transmission occurring in Maharashtra over the study period. Several potential sampling biases exist (including preferential sampling of symptomatic or severely ill individuals, preferential sequencing of samples based on PCR diagnostic results etc.), again highlighting the need for greater investment in routine genomic sequencing during viral epidemics. More broadly, our model makes the important simplifying assumption of homogeneous mixing between populations and various age groups. Hence, our model does not capture the more nuanced heterogeneities in spread likely to have occurred both across the state (e.g., between cities) and within-cities (such as between slum and non-slum dwelling populations). In spite of the limitations, such expanded sensitivity analyses can be helpful to understand the altered epidemiological characteristics of emerging sub-lineages like the “delta plus” (B.1.617.2.1) or PANGO lineage designation AY.1 variant with the K417N mutation. Finally, we recognize that retrospective analysis of the role of NPI is of limited use, many forecasting models alerted of this surge in February and March when such NPIs could actually save lives. However, we hope the present analysis helps to design preventions for future surges by quantifying the value of timely interventions.

### Conclusion

Rapid expansion of vaccination efforts as an effective, evidence-based intervention to reduce COVID-19 transmission and death is the way moving forward for India. Amidst emerging variants of concern, several small studies collectively suggest that antibodies are slightly less protective against B.1.617 relative to lineages that were present in the beginning of the pandemic. In Delhi, examination of serum from 30 re-infected healthcare workers, all fully vaccinated, revealed a resounding presence of B.1.617, thereby building evidence of this variant’s ability to evade antibodies (*66*). Likewise, German investigators found that the B.1.617 variant confers a reduction in antibody neutralization efficiency (50% among convalescent patients and 67% among those with both doses of the Pfizer vaccine) compared to common variants at the outset of the pandemic (*67*). While a comprehensive countrywide and region-specific investigation in India into the variant landscape, level of transmissibility, and immune evasion is needed, early evidence suggests attenuated yet effective levels of cross-immunity – vaccines work. The projected timeline to get the adult population of India vaccinated is by the end of this year (*68*). However, India is a very young population and roughly 40% of the population is in the age group of 0-18 years (*69*), for whom there are no vaccines available yet. Until there is a sufficient supply of vaccines to inoculate most of the Indian population, non-pharmaceutical public health interventions will be key for India as the country designs its current lockdown exit strategy. Such measures are crucial for curbing deaths, infections, and subsequent viral mutation; out analysis demonstrates that the earlier an intervention takes place, the better – timing matters.

Our findings echo the need for enhanced surveillance efforts, continued attention to the emergence of variants and examination of subsequent consequences for transmissibility, cross-immunity, and vaccine effectiveness. The scenarios we ran for the models with sequencing data show us that the resurgence of the second wave cannot be fully explained by the waning immunity or even the increased transmissibility. The extent of reinfections or immune evasion can only be substantiated after clinical studies of the variants, and we also need better availability of data so that we can move from modelling scenarios to more data-based inferences to provide better understanding of the epidemic landscape in India. We urge further ramping up of the sequencing effort in India to ensure sequencing-based epidemiological analyses are equipped with a sufficiently large, representative sample and to keep pace with, as well as predict, the various evolutionary paths of the virus. Strengthening the surveillance for COVID19 in every district is important for identifying clusters requiring further investigation. Among existent innovative strategies, wastewater analysis has also been shown to be a promising method for cross-validation of clinical data, for evaluating regional genomic sequencing efforts, and for in turn identifying novel SARS-CoV-2 strains otherwise undetected (*70*). Prioritizing sequencing of reinfections, as well as vaccine breakthrough infections, is necessary to understand vaccine effectiveness and development needs against emerging Variants of Concern. We hope that the lessons from Wave 2 lead to a bolstering of public health infrastructure, timely and comprehensive collection, and release of data as well as compel policymakers to act more proactively, thereby preparing India to respond to future waves and crises.

## Data Availability

We use data on reported infected cases and COVID-19-attributed deaths through May 31, 2021, for our descriptive analysis (Section 1) and May 15, 2021, for predictive modeling (Sections 2 and 3) from covid19india.org (14). Population data for India was obtained from Our World in Data (71). Genomic data was obtained from GISAID (49, 50). Analysis code are available at on the Center for Precision Health Data Science GitHub page: https://github.com/umich-cphds/covid_india_wave2.

https://www.covid19india.org

https://www.gisaid.org

## Acknowledgments

The authors thank Lili Wang for her advisement regarding the use of her eSIR R package. The authors also thank authors from the originating laboratories and submitting laboratories for their collection, generation, and proliferation of the GISAID data on which this study is based (full author list attached as supplementary material).

## Author Contributions

Conceptualization: SM, BM

Methodology: MS, MK, RB, SP, SM, TM, CW, SF, SB, BM

Investigation: MS, SP, LZ, SB, SM, BM

Visualization: MS, SP, SM

Funding acquisition: BM, SB, SF

Project administration: MS, SM, BM

Supervision: SM, BM

Writing – original draft: SM, RB, SP, LZ, DR, TM, CW, SF, SB, SM, BM

Writing – review & editing: MS, LZ, DR, SB, SM, BM

## Data and materials availability

We use data on reported infected cases and COVID-19-attributed deaths through May 31, 2021, for our descriptive analysis (Section 1) and May 15, 2021, for predictive modeling (Sections 2 and 3) from covid19india.org (*14*). Population data for India was obtained from Our World in Data (*71*). Genomic data was obtained from GISAID (*49, 50*). Analysis code are available at on the Center for Precision Health Data Science GitHub page: https://github.com/umich-cphds/covid_india_wave2.

## Supplementary materials

Materials and Methods Sections 1 – 3

Table S1 – S9

Fig S1 – S8

Supplementary references 1 – 27

## REFERENCES

1. M. A. Andrews, B. Areekal, K. R. Rajesh, J. Krishnan, R. Suryakala, B. Krishnan, C. P. Muraly, P. V. Santhosh, First confirmed case of COVID-19 infection in India: A case report. Indian J. Med. Res. 151, 490–492 (2020).

2. COVID-19 India Timeline. The Wire, (available at https://thewire.in/covid-19-india-timeline).

3. WHO Director-General’s opening remarks at the media briefing on COVID-19 - 11 March 2020, (available at https://www.who.int/director-general/speeches/detail/who-director-general-s-opening-remarks-at-the-media-briefing-on-covid-1911-march-2020).

4. D. Ray, M. Salvatore, R. Bhattacharyya, L. Wang, J. Du, S. Mohammed, S. Purkayastha, A. Halder, A. Rix, D. Barker, M. Kleinsasser, Y. Zhou, D. Bose, P. Song, M. Banerjee, V. Baladandayuthapani, P. Ghosh, B. Mukherjee, Predictions, Role of Interventions and Effects of a Historic National Lockdown in India’s Response to the the COVID-19 Pandemic: Data Science Call to Arms. Harv. Data Sci. Rev. (2020), doi:10.1162/99608f92.60e08ed5.

5. T. N. Service, Centre extends nationwide lockdown till May 31, new guidelines issued. Trib. News Serv., (available at https://www.tribuneindia.com/news/nation/centre-extends-nationwide-lockdown-till-may-31-new-guidelines-issued-86042).

6. J. Gettleman, K. Schultz, Modi Orders 3-Week Total Lockdown for All 1.3 Billion Indians. N. Y. Times (2020), (available at https://www.nytimes.com/2020/03/24/world/asia/india-coronavirus-lockdown.html).

7. U. Bhaskar, India to remain closed till 3 May, economy to open up gradually in lockdown 2.0. mint (2020), (available at https://www.livemint.com/news/india/pm-modi-announces-extension-of-lockdown-till-3-may-11586839412073.html).

8. ANI, India scales up testing capacity from 1 in January to over 7.7 crore in October: MoHFW. Econ. Times (2020), (available at https://economictimes.indiatimes.com/news/politics-and-nation/india-scales-up-testing-capacity-from-1-in-january-to-over-7-7-crore-in-october-mohfw/articleshow/78471855.cms?from=mdr).

9. Updates on COVID-19, (available at pib.gov.in/Pressreleaseshare.aspx?PRID=1615405).

10. G. R. Babu, D. Ray, R. Bhaduri, A. Halder, R. Kundu, G. I. Menon, B. Mukherjee, Glob. Health Sci. Pract., in press, doi:10.9745/GHSP-D-21-00233.

11. “Unlock1”: Malls, Restaurants, Places Of Worship To Reopen June 8. NDTV.com, (available at https://www.ndtv.com/india-news/lockdown-extended-till-june-30-malls-restaurants-can-reopen-from-june-8-except-in-containment-zones-2237910).

12. M. Salvatore, D. Basu, D. Ray, M. Kleinsasser, S. Purkayastha, R. Bhattacharyya, B. Mukherjee, Comprehensive public health evaluation of lockdown as a non-pharmaceutical intervention on COVID-19 spread in India: national trends masking state-level variations. BMJ Open. 10, e041778 (2020).

13. P. J. Datta, India can expect about 4,92,380 total deaths by December 1, says US study. @businessline, (available at https://www.thehindubusinessline.com/news/india-can-expect-about-492380-total-deaths-by-december-1-says-us-study/article32472763.ece).

14. Coronavirus in India: Latest Map and Case Count, (available at https://www.covid19india.org).

15. R. Bhattacharyya, R. Kundu, R. Bhaduri, D. Ray, L. J. Beesley, M. Salvatore, B. Mukherjee, Incorporating false negative tests in epidemiological models for SARS-CoV-2 transmission and reconciling with seroprevalence estimates. Sci. Rep. 11, 9748 (2021).

16. N. Sharma, P. Sharma, S. Basu, S. Saxena, R. Chawla, K. Dushyant, N. Mundeja, Z. S. Marak, S. Singh, G. K. Singh, R. Rustagi, medRxiv, in press, doi:10.1101/2020.12.13.20248123.

17. M. Mohanan, A. Malani, K. Krishnan, A. Acharya, Prevalence of SARS-CoV-2 in Karnataka, India. JAMA. 325, 1001 (2021).

18. S. Selvaraju, M. S. Kumar, J. W. V. Thangaraj, T. Bhatnagar, V. Saravanakumar, C. P. G. Kumar, K. Sekar, E. Ilayaperumal, R. Sabarinathan, M. Jagadeesan, M. S. Hemalatha, M. V. Murhekar, Population-Based Serosurvey for Severe Acute Respiratory Syndrome Coronavirus 2 Transmission, Chennai, India - Volume 27, Number 2—February 2021 - Emerging Infectious Diseases journal - CDC, doi:10.3201/eid2702.203938.

19. Delhi’s 5th sero survey: Over 56% people have antibodies against Covid-19. Hindustan Times (2021), (available at https://www.hindustantimes.com/cities/delhi-news/delhis-5th-sero-survey-over-56-people-have-antibodies-against-covid19-101612264534349.html).

20. A. Ghose, S. Bhattacharya, A. S. Karthikeyan, A. Kudale, J. M. Monteiro, A. Joshi, G. Medigeshi, G. Kang, V. Bal, S. Rath, L. S. Shashidhara, J. John, S. Chaudhuri, A. Nagarkar, “Community prevalence of antibodies to SARS-CoV-2 and correlates of protective immunity in an Indian metropolitan city” (preprint, Infectious Diseases (except HIV/AIDS), 2020),, doi:10.1101/2020.11.17.20228155.

21. ICMR sero survey: One in five Indians exposed to Covid-19. BBC News (2021), (available at https://www.bbc.com/news/world-asia-india-55945382).

22. E. Schmall, S. Yasir, India Approves Oxford-AstraZeneca Covid-19 Vaccine and 1 Other. N. Y. Times (2021), (available at https://www.nytimes.com/2021/01/03/world/asia/india-covid-19-vaccine.html).

23. Ministry of Health and Family Welfare, “COVID-19 Vaccines Operational Guidelines” (2020).

24. A New Variant May Be The Cause Of India’s COVID-19 Surge. NPR.org, (available at https://www.npr.org/2021/05/06/994376812/a-new-variant-may-be-the-cause-of-indias-covid-19-surge).

25. A. Deshpande, Maharashtra imposes ‘mini lockdown’ amid rising coronavirus cases. The Hindu (2021), (available at https://www.thehindu.com/news/national/coronavirus-curfew-in-maharashtra-for-15-days-from-april-14-says-uddhav-thackeray/article34313068.ece).

26. T. Lancet, India’s COVID-19 emergency. The Lancet. 397, 1683 (2021).

27. The RECOVERY Collaborative Group, Dexamethasone in Hospitalized Patients with Covid-19. N. Engl. J. Med. 384, 693–704 (2021).

28. India Covid: How bad is the second wave? BBC News (2021), (available at https://www.bbc.com/news/56987209).

29. Covid-19 in India: Why second coronavirus wave is devastating. BBC News (2021), (available at https://www.bbc.com/news/world-asia-india-56811315).

30. A. Gupta, M. Banaji, “The scale of Gujarat’s mortality crisis” (preprint, SocArXiv, 2021),, doi:10.31235/osf.io/2wae5.

31. S. Purkayastha, R. Kundu, R. Bhaduri, D. Barker, M. Kleinsasser, D. Ray, B. Mukherjee, Estimating the wave 1 and wave 2 infection fatality rates from SARS-CoV-2 in India. medRxiv (2021), doi:10.1101/2021.05.25.21257823.

32. M. S. Dhar, R. Marwal, V. Radhakrishnan, K. Ponnusamy, B. Jolly, R. C. Bhoyar, S. Fatihi, M. Datta, P. Singh, U. Sharma, R. Ujjainia, S. Naushin, N. Bhateja, M. K. Divakar, V. Sardana, M. K. Singh, M. Imran, V. Senthivel, R. Maurya, N. Jha, P. Mehta, M. Rophina, V. Arvinden, U. Chaudhary, L. Thukral, R. Pandey, D. Dash, M. Faruq, H. Lall, H. Gogia, P. Madan, S. Kulkarni, H. Chauhan, S. Sengupta, S. Kabra, The Indian SARS-CoV-2 Genomics Consortium (INSACOG), S. K. Singh, A. Agrawal, P. Rakshit, “Genomic characterization and Epidemiology of an emerging SARS-CoV-2 variant in Delhi, India” (preprint, Epidemiology, 2021),, doi:10.1101/2021.06.02.21258076.

33. R. Sarkar, R. Saha, P. Mallick, R. Sharma, A. Kaur, S. Dutta, M. Chawla-Sarkar, “Emergence of a new SARS-CoV-2 variant from GR clade with a novel S glycoprotein mutation V1230L in West Bengal, India” (preprint, Epidemiology, 2021),, doi:10.1101/2021.05.24.21257705.

34. Tracking SARS-CoV-2 variants, (available at https://www.who.int/activities/tracking-SARS-CoV-2-variants).

35. A. Chudik, M. H. Pesaran, A. Rebucci, COVID-19 time-varying reproduction numbers worldwide: an empirical analysis of mandatory and voluntary social distancing. NBER Work. Pap. Ser. (available at https://www.nber.org/system/files/working_papers/w28629/w28629.pdf).

36. Genome Sequencing by INSACOG shows variants of concern and a Novel variant in India, (available at pib.gov.in/Pressreleaseshare.aspx?PRID=1707177).

37. S. Alai, N. Gujar, M. Joshi, M. Gautam, S. Gairola, Pan-India novel coronavirus SARS-CoV-2 genomics and global diversity analysis in spike protein. Heliyon. 7 (2021), doi:10.1016/j.heliyon.2021.e06564.

38. S. Cherian, V. Potdar, S. Jadhav, P. Yadav, N. Gupta, M. Das, P. Rakshit, S. Singh, P. Abraham, S. Panda, NIC team, “Convergent evolution of SARS-CoV-2 spike mutations, L452R, E484Q and P681R, in the second wave of COVID-19 in Maharashtra, India” (preprint, Molecular Biology, 2021),, doi:10.1101/2021.04.22.440932.

39. Maharashtra: double mutant found in 61% samples tested. Indian Express (2021), (available at https://indianexpress.com/article/cities/mumbai/maharashtra-double-mutant-found-in-61-samples-tested-7272524/).

40. Weekly epidemiological update on COVID-19 - 11 May 2021, (available at https://www.who.int/publications/m/item/weekly-epidemiological-update-on-covid-19---11-may-2021).

41. C. Pattabiraman, P. Prasad, A. K. George, D. Sreenivas, R. Rasheed, N. V. K. Reddy, A. Desai, R. Vasanthapuram, Importation, circulation, and emergence of variants of SARS-CoV-2 in the South Indian state of Karnataka. Wellcome Open Res. 6, 110 (2021).

42. 81 per cent of Punjab COVID-19 samples sent for genome sequencing show new UK variant. New Indian Express, (available at https://www.newindianexpress.com/nation/2021/mar/23/81-per-cent-of-punjab-covid-19-samples-sent-for-genome-sequencing-show-new-uk-variant-2280394.html).

43. J. Koshy, Coronavirus | New virus lineage found in West Bengal. The Hindu (2021), (available at https://www.thehindu.com/sci-tech/health/new-coronavirus-variant-found-in-west-bengal/article34373083.ece).

44. Why India’s massive COVID-19 surge is breaking tragic records. Science (2021), (available at https://www.nationalgeographic.com/science/article/how-indias-second-wave-became-the-worst-covid-19-surge-in-the-world).

45. B. Parkin, J. Singh, S. Findlay, J. Burn-Murdoch, India’s devastating second wave: ‘It is much worse this time.’ Financ. Times (2021), (available at https://www.ft.com/content/683914a3-134f-40b6-989b-21e0ba1dc403).

46. R. Cai, P. Novosad, V. Tandel, S. Asher, A. Malani, Representative Estimates of COVID-19 Infection Fatality Rates from Three Locations in India. medRxiv (2021), doi:10.1101/2021.01.05.21249264.

47. Bihar revises Covid toll, deaths jump to over 9,000. Indian Express (2021), (available at https://indianexpress.com/article/india/bihar-covid-death-toll-revision-7352073/).

48. J. Gettleman, S. Yasir, H. Kumar, S. Raj, A. Loke, As Covid-19 Devastates India, Deaths Go Undercounted. N. Y. Times (2021), (available at https://www.nytimes.com/2021/04/24/world/asia/india-coronavirus-deaths.html).

49. S. Elbe, G. Buckland-Merrett, Data, disease and diplomacy: GISAID’s innovative contribution to global health: Data, Disease and Diplomacy. Glob. Chall. 1, 33–46 (2017).

50. Y. Shu, J. McCauley, GISAID: Global initiative on sharing all influenza data – from vision to reality. Eurosurveillance. 22 (2017), doi:10.2807/1560-7917.ES.2017.22.13.30494.

51. J. L. Mullen, G. Tsueng, A. Abdel Latif, M. Alkuzweny, M. Cano, E. Haag, J. Zhou, M. Zeller, N. Matteson, K. G. Andersen, C. Wu, A. I. Su, K. Gangavarapu, L. D. Hughes, The Center for Viral Systems Biology, outbreak.info. outbreak.info (2020), (available at https://outbreak.info/).

52. F. Konings, M. D. Perkins, J. H. Kuhn, M. J. Pallen, E. J. Alm, B. N. Archer, A. Barakat, T. Bedford, J. N. Bhiman, L. Caly, L. L. Carter, A. Cullinane, T. de Oliveira, J. Druce, I. El Masry, R. Evans, G. F. Gao, A. E. Gorbalenya, E. Hamblion, B. L. Herring, E. Hodcroft, E. C. Holmes, M. Kakkar, S. Khare, M. P. G. Koopmans, B. Korber, J. Leite, D. MacCannell, M. Marklewitz, S. Maurer-Stroh, J. A. M. Rico, V. J. Munster, R. Neher, B. O. Munnink, B. I. Pavlin, M. Peiris, L. Poon, O. Pybus, A. Rambaut, P. Resende, L. Subissi, V. Thiel, S. Tong, S. van der Werf, A. von Gottberg, J. Ziebuhr, M. D. Van Kerkhove, SARS-CoV-2 Variants of Interest and Concern naming scheme conducive for global discourse. Nat. Microbiol. (2021), doi:10.1038/s41564-021-00932-w.

53. E. C. Wall, M. Wu, R. Harvey, G. Kelly, S. Warchal, C. Sawyer, R. Daniels, P. Hobson, E. Hatipoglu, Y. Ngai, S. Hussain, J. Nicod, R. Goldstone, K. Ambrose, S. Hindmarsh, R. Beale, A. Riddell, S. Gamblin, M. Howell, G. Kassiotis, V. Libri, B. Williams, C. Swanton, S. Gandhi, D. L. Bauer, Neutralising antibody activity against SARS-CoV-2 VOCs B.1.617.2 and B.1.351 by BNT162b2 vaccination. The Lancet, S0140673621012903 (2021).

54. Public Health England, “3 June 2021 Risk assessment for SARS-CoV-2 variant: Delta (VOC-21 APR-02, B.1.617.2)” (2021), (available at https://assets.publishing.service.gov.uk/government/uploads/system/uploads/attachment_data/file/991135/3_June_2021_Risk_assessment_for_SARS-CoV-2_variant_DELTA.pdf).

55. N. R. Faria, T. A. Mellan, C. Whittaker, I. M. Claro, D. da S. Candido, S. Mishra, M. A. E. Crispim, F. C. S. Sales, I. Hawryluk, J. T. McCrone, R. J. G. Hulswit, L. A. M. Franco, M. S. Ramundo, J. G. de Jesus, P. S. Andrade, T. M. Coletti, G. M. Ferreira, C. A. M. Silva, E. R. Manuli, R. H. M. Pereira, P. S. Peixoto, M. U. G. Kraemer, N. Gaburo, C. da C. Camilo, H. Hoeltgebaum, W. M. Souza, E. C. Rocha, L. M. de Souza, M. C. de Pinho, L. J. T. Araujo, F. S. V. Malta, A. B. de Lima, J. do P. Silva, D. A. G. Zauli, A. C. de S. Ferreira, R. P. Schnekenberg, D. J. Laydon, P. G. T. Walker, H. M. Schlüter, A. L. P. dos Santos, M. S. Vidal, V. S. D. Caro, R. M. F. Filho, H. M. dos Santos, R. S. Aguiar, J.L. Proença-Modena, B. Nelson, J. A. Hay, M. Monod, X. Miscouridou, H. Coupland, R. Sonabend, M. Vollmer, A. Gandy, C. A. Prete, V. H. Nascimento, M. A. Suchard, T. A. Bowden, S. L. K. Pond, C.-H. Wu, O. Ratmann, N. M. Ferguson, C. Dye, N. J. Loman, P. Lemey, A. Rambaut, N. A. Fraiji, M. do P. S. S. Carvalho, O. G. Pybus, S. Flaxman, S. Bhatt, E. C. Sabino, Genomics and epidemiology of the P.1 SARS-CoV-2 lineage in Manaus, Brazil. Science. 372, 815– 821 (2021).

56. S. Flaxman, S. Mishra, A. Gandy, H. J. T. Unwin, T. A. Mellan, H. Coupland, C. Whittaker, H. Zhu, T. Berah, J. W. Eaton, M. Monod, Imperial College COVID-19 Response Team, P. N. Perez-Guzman, N. Schmit, L. Cilloni, K. E. C. Ainslie, M. Baguelin, A. Boonyasiri, O. Boyd, L. Cattarino, L. V. Cooper, Z. Cucunubá, G. Cuomo-Dannenburg, A. Dighe, B. Djaafara, I. Dorigatti, S. L. van Elsland, R. G. FitzJohn, K. A. M. Gaythorpe, L. Geidelberg, N. C. Grassly, W. D. Green, T. Hallett, A. Hamlet, W. Hinsley, B. Jeffrey, E. Knock, D. J. Laydon, G. Nedjati-Gilani, P. Nouvellet, K. V. Parag, I. Siveroni, H. A. Thompson, R. Verity, E. Volz, C. E. Walters, H. Wang, Y. Wang, O. J. Watson, P. Winskill, X. Xi, P. G. T. Walker, A. C. Ghani, C. A. Donnelly, S. Riley, M. A. C. Vollmer, N. M. Ferguson, L. C. Okell, S. Bhatt, Estimating the effects of non-pharmaceutical interventions on COVID-19 in Europe. Nature. 584, 257–261 (2020).

57. C. Fraser, Estimating Individual and Household Reproduction Numbers in an Emerging Epidemic. PLoS ONE. 2, e758 (2007).

58. M. Murhekar, T. Bhatnagar, S. Selvaraju, K. Rade, V. Saravanakumar, J. Vivian Thangaraj, M. Kumar, N. Shah, R. Sabarinathan, A. Turuk, P. Anand, S. Asthana, R. Balachandar, S. Bangar, A. Bansal, J. Bhat, D. Chakraborty, C. Rangaraju, V. Chopra, D. Das, A. Deb, K. Devi, G. Dwivedi, Sm. Salim Khan, I. Haq, Ms. Kumar, A. Laxmaiah, Madhuka A. Mahapatra, A. Mitra, A. Nirmala, A. Pagdhune, M. Qurieshi, T. Ramarao, S. Sahay, Y. Sharma, M. Shrinivasa, V. Shukla, P. Singh, A. Viramgami, V. Wilson, R. Yadav, C. Girish Kumar, H. Luke, U. Ranganathan, S. Babu, K. Sekar, P. Yadav, G. Sapkal, A. Das, P. Das, S. Dutta, R. Hemalatha, A. Kumar, K. Narain, S. Narasimhaiah, S. Panda, S. Pati, S. Patil, K. Sarkar, S. Singh, R. Kant, S. Tripathy, G. Toteja, G. Babu, S. Kant, J. Muliyil, R. Pandey, S. Sarkar, S. Singh, S. Zodpey, R. Gangakhedkar, D. S. Reddy, B. Bhargava, Prevalence of SARS-CoV-2 infection in India: Findings from the national serosurvey, May-June 2020. Indian J. Med. Res. 152, 48 (2020).

59. Covid-19 second wave: Here’s a list of states that have imposed full lockdown. Indian Express (2021), (available at https://indianexpress.com/article/india/covid-19-second-wave-heres-a-list-of-states-that-have-imposed-lockdowns-7306634/).

60. M. Banaji, “Estimating COVID-19 infection fatality rate in Mumbai during 2020” (preprint, Epidemiology, 2021),, doi:10.1101/2021.04.08.21255101.

61. M. Pons-Salort, J. John, O. J. Watson, N. F. Brazeau, R. Verity, G. Kang, N. C. Grassly, “Reconstructing the COVID-19 epidemic in Delhi, India: infection attack rate and reporting of deaths” (preprint, Infectious Diseases (except HIV/AIDS), 2021),, doi:10.1101/2021.03.23.21254092.

62. L. Zimmermann, M. Salvatore, G. Babu, B. Mukherjee, “Estimating COVID-19 Related Mortality in India: An Epidemiological Challenge with Insufficient Data” (preprint, LIFE SCIENCES, 2021),, doi:10.20944/preprints202105.0617.v1.

63. P. Misra, S. Kant, R. Guleria, S. K. Rai, WHO Unity Seroprevalence study team of AIIMS, “Serological prevalence of SARS-CoV-2 antibody among children and young age (between age 2-17 years) group in India: An interim result from a large multi-centric population-based seroepidemiological study” (preprint, Epidemiology, 2021),, doi:10.1101/2021.06.15.21258880.

64. Nextstrain / ncov / asia. Genomic Epidemiol. Nov. Coronavirus - Asia-Focus. Subsampling, (available at https://nextstrain.org/ncov/asia).

65. J. Hadfield, C. Megill, S. M. Bell, J. Huddleston, B. Potter, C. Callender, P. Sagulenko, T. Bedford, R. A. Neher, Nextstrain: real-time tracking of pathogen evolution. Bioinformatics. 34, 4121–4123 (2018).

66. Ferreira, R. Datir, S. Kemp, G. Papa, P. Rakshit, S. Singh, B. Meng, R. Pandey, K. Ponnusamy, V. S. Radhakrishnan, The Indian SARS-CoV-2 Genomics Consortium (INSACOG), The CITIID-NIHR BioResource COVID-19 Collaboration, K. Sato, L. James, A. Agrawal, R.K. Gupta, “SARS-CoV-2 B.1.617 emergence and sensitivity to vaccine-elicited antibodies” (preprint, Microbiology, 2021),, doi:10.1101/2021.05.08.443253.

67. M. Hoffmann, H. Hofmann-Winkler, N. Krüger, A. Kempf, I. Nehlmeier, L. Graichen, A. Sidarovich, A.-S. Moldenhauer, M. S. Winkler, S. Schulz, H.-M. Jäck, M. V. Stankov, G. M. N. Behrens, S. Pöhlmann, “SARS-CoV-2 variant B.1.617 is resistant to Bamlanivimab and evades antibodies induced by infection and vaccination” (preprint, Molecular Biology, 2021),, doi:10.1101/2021.05.04.442663.

68. A. S. N. DelhiMay 21, 2021UPDATED: May 21, 2021 20:37 Ist, By end of 2021, India will be in position to vaccinate all adults: Health Minister Harsh Vardhan. India Today, (available at https://www.indiatoday.in/coronavirus-outbreak/story/india-vaccinate-adults-covid-union-health-minister-1805469-2021-05-21).

69. Census of India Website : Office of the Registrar General & Census Commissioner, India, (available at https://censusindia.gov.in/2011census/C-series/C-13.html).

70. T. Dharmadhikari, V. Rajput, R. Yadav, R. Boaragaonkar, D. Panse, S. Kamble, S. Dastager, M. Dharne, “High throughput sequencing based detection of SARS-CoV-2 prevailing in wastewater of Pune, West India” (preprint, Epidemiology, 2021),, doi:10.1101/2021.06.08.21258563.

71. H. Ritchie, E. Ortiz-Ospina, D. Beltekian, E. Mathieu, J. Hasell, B. Macdonald, C. Giattino, C. Appel, L. Rodés-Guirao, M. Roser, Coronavirus Pandemic (COVID-19). Our World Data (2020) (available at https://ourworldindata.org/coronavirus/country/india).

