## Supplementary Material for "Resurgence of SARS-CoV-2 in India: Potential role of the B.1.617.2 (Delta) variant and delayed interventions"

**Section 1**. Methods for Comparison of First and Second Waves in India 1

**Section 2**. Methods for Characterizing the Epidemiologic Properties of B.1.617.2 4

**Section 3**. Methods for Characterizing the Potential Effect of Public Health Interventions 10

**Section 3.1**. **Forecast results with waning immunity** 23

**REFERENCES** 31

**Data Sources:** We use data on reported infected cases and COVID-19-attributed deaths through May 31, 2021, for our descriptive analysis (Section 1) and May 15, 2021, for predictive modeling (Sections 2 and 3) from covid19india.org (*1*). Population data for India was obtained from Our World in Data (*2*). Genomic data was obtained from GISAID (3, 4).

**Analysis Code are available at on the Center for Precision Health Data Science GitHub page:** https://github.com/umich-cphds/covid_india_wave2

#### **Section 1**. Methods for Comparison of First and Second Waves in India

We perform a comparative analysis of the first and second wave (Waves 1 and 2 respectively) of the pandemic in India by comparing cumulative case (death) counts associated with both the waves, standardized by the duration (in days) of each wave across the states and union territories.

We define the beginning of waves 1 and 2 of the pandemic as June 13, 2020, and February 14, 2021, respectively. This definition has been artificially created by other researchers (*5*) and is guided by the fact that the national effective reproduction number ($R_t$) crossed unity for the first time in 2021 on February 14, thereby marking the onset of the acceleration phase of the second wave. To ensure a fair comparison between the waves, we define the starting point of wave 1 as the first day of 2020 when the reported new infection count exceeds the number of new cases reported on February 14, 2021 (11,706 new cases were reported). Following these definitions, Wave 1 is the phase from June 14, 2020 - February 13, 2021 (lasting 246 days) and wave 2 is the phase from February 14, 2021 - May 31, 2021 (lasting 107 days). We define the case (death) rates of a specific wave as the cumulative case (death) counts reported during that wave, standardized by the duration of the corresponding wave. To compare waves 1 and 2 by means of this metric, we examine the ratio of wave 2 daily case (death) rates to wave 1 case (death) intensities for each state and plot them on a map of India (see ***Figure 1***). States with case (death) rate ratios less than unity are shaded blue while those with higher ratios are shaded in increasingly darker shades of red. The rate ratios above unity are grouped into five bins 1-2, 2-3, 3-4. 4-5 and 5+. Further, ratios from some small union territories (Ladakh, Dadra and Nagar Haveli, Lakshadweep) have not been included in this figure.

For comparing daily peaks in cases and deaths ***(Supplementary Table S1)*** we simply looked for the maximum reported case and death counts. The sequence of peaks show that India is a heterogeneous nation and while case counts came down in Delhi and Maharashtra with a strict lockdown, Southern and Eastern states reached their peak later. Similarly, for death counts the peaks were attained from late April to late May.

**Table S1a**. Comparison of peak timing of daily new cases across Indian states and union territories in Wave 1 vs Wave 2.

| | Wave 1 (June 13, 2020-February 13, 2021: 246 days) | | | | Wave 2 (February 14-May 15, 2021:107 days) | | | | Comparison Metrics | | |
|---|---|---|---|---|---|---|---|---|---|---|---|
| Place | Order | Peak Date | Peak Daily Cases | Total Cases | Order | Peak Date | Peak Daily Cases | Total Cases | % cases in Wave 2[a] | Case rate ratio[b] | Case tier[c] |
| Ladakh | 1 | 6/13/20* | 198 | 9,526 | 1 | 4/17/21 | 362 | 8,897 | 48.29% | 2.15 | 2-3 |
| Tamil Nadu | 2 | 7/27/20 | 6,993 | 803,952 | 32 | 5/21/21 | 36,184 | 1,251,866 | **60.89%** | 3.58 | 3-4 |
| Bihar | 3 | 8/11/20 | 4,071 | 255,594 | 9 | 4/30/21 | 15,853 | 445,071 | **63.52%** | 4.00 | 4-5 |
| Assam | 4 | 8/12/20 | 4,593 | 213,492 | 29 | 5/20/21 | 6,573 | 193,928 | 47.60% | 2.09 | 2-3 |
| Andaman and Nicobar Islands | 5 | 8/14/20 | 149 | 4,967 | 11 | 5/1/21 | 97 | 1,996 | 28.67% | 0.92 | < 1 |
| Sikkim | 6 | 8/14/20 | 149 | 6,067 | 35 | 5/28/21 | 420 | 9,201 | **60.26%** | 3.49 | 3-4 |
| Dadra and Nagar Haveli | 7 | 8/15/20 | 64 | 3,349 | 4 | 4/22/21 | 359 | 6,763 | **66.88%** | 4.73 | 4-5 |
| Andhra Pradesh | 8 | 8/26/20 | 10,830 | 883,178 | 27 | 5/16/21 | 24,171 | 804,271 | 47.66% | 2.09 | 2-3 |
| Telangana | 9 | 8/26/20 | 3,018 | 291,434 | 18 | 5/7/21 | 11,451 | 281,923 | 49.17% | 2.22 | 2-3 |
| Jharkhand | 10 | 8/31/20 | 3,221 | 117,602 | 8 | 4/28/21 | 8,075 | 218,490 | **65.01%** | 4.27 | 4-5 |
| Tripura | 11 | 9/5/20 | 691 | 32,387 | 28 | 5/19/21 | 879 | 17,780 | 35.44% | 1.26 | 1-2 |
| Mizoram | 12 | 9/10/20 | 141 | 4,288 | 37 | 5/30/21 | 329 | 7,695 | **64.22%** | 4.13 | 4-5 |
| Maharashtra | 13 | 9/11/20 | 24,886 | 1,959,045 | 2 | 4/18/21 | 68,631 | 3,686,706 | **65.30%** | 4.33 | 4-5 |
| Nagaland | 14 | 9/11/20 | 310 | 11,990 | 25 | 5/13/21 | 366 | 9,497 | 44.20% | 1.82 | 1-2 |
| Uttar Pradesh | 15 | 9/11/20 | 7,016 | 589,473 | 6 | 4/24/21 | 37,944 | 1,089,399 | **64.89%** | 4.25 | 4-5 |
| Goa | 16 | 9/12/20 | 740 | 53,730 | 16 | 5/7/21 | 4,195 | 101,473 | **65.38%** | 4.34 | 4-5 |
| Jammu and Kashmir | 17 | 9/12/20 | 1,698 | 120,538 | 17 | 5/7/21 | 5,443 | 165,197 | **57.81%** | 3.15 | 3-4 |
| Chandigarh | 18 | 9/13/20 | 449 | 20,875 | 22 | 5/9/21 | 895 | 38,826 | **65.03%** | 4.28 | 4-5 |
| **India** | **19** | **9/16/20** | **97,860** | **10,595,100** | **15** | **5/6/21** | **414,280** | **17,268,970** | **61.98%** | **3.75** | **3-4** |
| Punjab | 20 | 9/17/20 | 2,848 | 173,078 | 21 | 5/8/21 | 9,042 | 391,543 | **69.35%** | 5.20 | > 5 |
| Uttarakhand | 21 | 9/19/20 | 2,078 | 380,168 | 19 | 5/7/21 | 9,642 | 931,660 | **71.02%** | 5.63 | > 5 |
| Pondicherry | 22 | 9/24/20 | 668 | 39,279 | 23 | 5/11/21 | 2,049 | 65,013 | **62.34%** | 3.81 | 3-4 |
| Odisha | 23 | 9/26/20 | 4,356 | 332,603 | 33 | 5/23/21 | 12,852 | 428,896 | **56.32%** | 2.96 | 2-3 |
| Arunachal Pradesh | 24 | 9/28/20 | 328 | 16,749 | 34 | 5/28/21 | 497 | 10,441 | 38.40% | 1.43 | 1-2 |
| Meghalaya | 25 | 10/3/20 | 423 | 13,845 | 30 | 5/20/21 | 1,183 | 21,661 | **61.01%** | 3.58 | 3-4 |
| Karnataka | 26 | 10/7/20 | 10,947 | 938,340 | 14 | 5/5/21 | 50,112 | 1,659,575 | **63.88%** | 4.07 | 4-5 |
| Kerala | 27 | 10/10/20 | 11,755 | 997,201 | 24 | 5/12/21 | 43,529 | 1,527,056 | **60.50%** | 3.52 | 3-4 |
| Madhya Pradesh | 28 | 10/14/20 | 5,515 | 246,980 | 7 | 4/25/21 | 13,601 | 522,607 | **67.91%** | 4.86 | 4-5 |
| Manipur | 29 | 10/17/20 | 426 | 28,796 | 36 | 5/30/21 | 1,032 | 21,564 | 42.82% | 1.72 | 1-2 |
| West Bengal | 30 | 10/22/20 | 4,157 | 562,161 | 26 | 5/14/21 | 20,846 | 803,972 | **58.85%** | 3.29 | 3-4 |
| Delhi | 31 | 11/11/20 | 8,593 | 599,972 | 3 | 4/20/21 | 28,395 | 789,444 | **56.82%** | 3.03 | 3-4 |
| Haryana | 32 | 11/20/20 | 3,104 | 262,635 | 13 | 5/4/21 | 15,786 | 487,666 | **65.00%** | 4.27 | 4-5 |
| Rajasthan | 33 | 11/24/20 | 3,314 | 306,752 | 12 | 5/2/21 | 18,298 | 621,138 | **66.94%** | 4.66 | 4-5 |
| Gujarat | 34 | 11/27/20 | 1,607 | 242,434 | 10 | 4/30/21 | 14,605 | 544,172 | **69.18%** | 5.16 | > 5 |
| Himachal Pradesh | 35 | 11/29/20 | 1,026 | 57,715 | 20 | 5/8/21 | 5,424 | 132,129 | **69.60%** | 5.26 | > 5 |
| Chhattisgarh | 36 | 1/28/21 | 6,451 | 1,229,940 | 5 | 4/23/21 | 17,397 | 2,650,132 | **68.30%** | 4.95 | 4-5 |
| Lakshadweep | 37 | 2/13/21 | 36 | 224 | 31 | 5/21/21 | 345 | 7,853 | 97.23% | 80.60 | > 5 |

* Ladakh peak occurred prior to our defined Wave 1 (June 13, 2020 - February 14, 2021)

[a] Total cases in Wave 2/ Total cases reported on May 31 (Wave 1+Wave 2)

[b] Daily case rate in Wave 2/ Daily case rate in wave 2.

[c] Tier based on daily case rate ratio

**Table S1b**. Comparison of deaths peak timing across states in Wave 1 vs Wave 2

| | Wave 1 (June 13, 2020-February 13, 2021: 246 days) | | | | Wave 2 (February 14-May 15, 2021:107 days) | | | | Comparison Metrics | | |
|---|---|---|---|---|---|---|---|---|---|---|---|
| Place | Order | Peak Date | Peak Daily Deaths | Total Deaths | Order | Peak Date | Peak Daily Deaths | Total Deaths | % deaths in Wave 2[a] | Death ratio[b] | Death tier[c] |
| Dadra and Nagar Haveli | 1 | 7/7/20 | 1 | 2 | 1 | 4/15/21 | 2 | 2 | 50.00% | 2.30 | 2-3 |
| Gujarat | 2 | 7/21/20 | 34 | 2,955 | 4 | 4/29/21 | 180 | 5,433 | 64.77% | 4.19 | 4-5 |
| Tamil Nadu | 3 | 7/22/20 | 518 | 11,997 | 36 | 5/30/21 | 493 | 11,819 | 49.63% | 2.26 | 2-3 |
| Telangana | 4 | 7/31/20 | 14 | 1,436 | 10 | 5/7/21 | 87 | 1,667 | 53.72% | 2.66 | 2-3 |
| Andhra Pradesh | 5 | 8/8/20 | 97 | 7,080 | 26 | 5/22/21 | 118 | 3,768 | 34.73% | 1.22 | 1-2 |
| Jharkhand | 6 | 8/9/20 | 17 | 1,074 | 6 | 5/1/21 | 169 | 3,909 | 78.45% | 8.37 | > 5 |
| Andaman and Nicobar Islands | 7 | 8/16/20 | 4 | 62 | 33 | 5/28/21 | 4 | 53 | 46.09% | 1.97 | 1-2 |
| Bihar | 8 | 8/16/20 | 22 | 1,488 | 20 | 5/18/21 | 111 | 3,639 | 70.98% | 5.62 | > 5 |
| Punjab | 9 | 9/2/20 | 106 | 5,630 | 23 | 5/18/21 | 231 | 8,856 | 61.13% | 3.62 | 3-4 |
| Pondicherry | 10 | 9/4/20 | 20 | 653 | 29 | 5/23/21 | 34 | 880 | 57.40% | 3.10 | 3-4 |
| Chhattisgarh | 11 | 9/9/20 | 70 | 15,060 | 2 | 4/28/21 | 279 | 37,108 | 71.13% | 5.66 | > 5 |
| Sikkim | 12 | 9/11/20 | 4 | 135 | 14 | 5/13/21 | 9 | 118 | 46.64% | 2.01 | 2-3 |
| Assam | 13 | 9/12/20 | 23 | 1,085 | 16 | 5/17/21 | 92 | 2,278 | 67.74% | 4.85 | 4-5 |
| Tripura | 14 | 9/13/20 | 12 | 387 | 35 | 5/29/21 | 13 | 122 | 23.97% | 0.72 | < 1 |
| Goa | 15 | 9/14/20 | 14 | 778 | 12 | 5/11/21 | 75 | 1,871 | 70.63% | 5.53 | > 5 |
| **India** | **16** | **9/15/20** | **1,281** | **144,194** | **19** | **5/18/21** | **4,529** | **176,233** | **55.00%** | **2.76** | **2-3** |
| Maharashtra | 17 | 9/15/20 | 515 | 46,363 | 28 | 5/23/21 | 1,320 | 43,855 | 48.61% | 2.11 | 2-3 |
| Uttar Pradesh | 18 | 9/15/20 | 113 | 8,316 | 11 | 5/7/21 | 372 | 11,798 | 58.66% | 3.25 | 3-4 |
| Karnataka | 19 | 9/18/20 | 179 | 12,184 | 27 | 5/23/21 | 624 | 16,827 | 58.00% | 3.17 | 3-4 |
| Jammu and Kashmir | 20 | 9/21/20 | 23 | 1,895 | 17 | 5/17/21 | 73 | 1,958 | 50.82% | 2.37 | 2-3 |
| Chandigarh | 21 | 9/23/20 | 10 | 340 | 8 | 5/5/21 | 14 | 408 | 54.55% | 2.76 | 2-3 |
| Madhya Pradesh | 22 | 9/24/20 | 45 | 3,378 | 3 | 4/28/21 | 105 | 4,238 | 55.65% | 2.88 | 2-3 |
| Meghalaya | 23 | 10/5/20 | 5 | 147 | 30 | 5/24/21 | 26 | 430 | 74.52% | 6.73 | > 5 |
| Odisha | 24 | 10/7/20 | 18 | 1,950 | 32 | 5/26/21 | 35 | 844 | 30.21% | 1.00 | < 1 |
| Arunachal Pradesh | 25 | 10/13/20 | 4 | 56 | 15 | 5/14/21 | 5 | 59 | 51.30% | 2.42 | 2-3 |
| West Bengal | 26 | 10/14/20 | 64 | 9,769 | 25 | 5/20/21 | 162 | 5,311 | 35.22% | 1.25 | 1-2 |
| Nagaland | 27 | 10/17/20 | 4 | 82 | 31 | 5/25/21 | 18 | 275 | 77.03% | 7.18 | > 5 |
| Uttarakhand | 28 | 10/17/20 | 95 | 6,632 | 18 | 5/17/21 | 223 | 19,088 | 74.21% | 6.61 | > 5 |
| Mizoram | 29 | 10/28/20 | 1 | 9 | 37 | 5/31/21 | 4 | 31 | 77.50% | 7.92 | > 5 |
| Delhi | 30 | 11/18/20 | 131 | 9,238 | 7 | 5/3/21 | 448 | 13,348 | 59.10% | 3.17 | 3-4 |
| Himachal Pradesh | 31 | 11/23/20 | 22 | 972 | 21 | 5/18/21 | 78 | 2,148 | 68.85% | 5.08 | > 5 |
| Haryana | 32 | 11/25/20 | 42 | 2,951 | 9 | 5/5/21 | 181 | 5,264 | 64.08% | 4.08 | 4-5 |
| Ladakh | 33 | 11/26/20 | 5 | 129 | 22 | 5/18/21 | 5 | 59 | 31.38% | 1.05 | 1-2 |
| Manipur | 34 | 11/29/20 | 10 | 373 | 24 | 5/19/21 | 23 | 434 | 53.78% | 2.68 | 2-3 |
| Rajasthan | 35 | 11/30/20 | 20 | 2,502 | 13 | 5/11/21 | 169 | 5,604 | 69.13% | 5.14 | > 5 |
| Kerala | 36 | 12/9/20 | 35 | 3,951 | 34 | 5/29/21 | 198 | 4,845 | 55.08% | 2.82 | 2-3 |
| Lakshadweep* | - | - | - | - | - | 4/29/21 | 3 | 42 | 100.00% | - | - |

* No deaths were reported from data source (covid19india.org) for Lakshadweep in Wave 1. As such, it is omitted from peak ranking and the comparison metrics are undefined.

Note: In the event there were ties of peak daily deaths, the earliest date on which that daily death count was recorded was selected for "Peak Date."

a Total deaths in Wave 2/ (Total reported by May 31, Wave 1+ Wave 2)

b Daily death rate in Wave 2/ Daily case rate in wave 2.

c Tier based on daily death rate ratio

#### **Section 2**. Methods for Characterizing the Epidemiologic Properties of B.1.617.2

***Table S2.*** Summary of existing studies on the SARS-CoV-2 variant landscape in India, during 2020-21

| ***Main Lineage /Clade*** | ***Other*** | ***Location*** | ***Sample Size*** | ***Summary*** | ***Ref*** |
|---|---|---|---|---|---|
| Delta (B.1.617.2) sublineage | Alpha (B.1.1.7), Kappa (B.1.617.1), B.1.36, B.1 lineages | Delhi, India and states in North India | N=>9,000 sequences across regions, from Nov 2020 to May 2021 | Alpha increased to 40% of samples in Delhi during the period of January to March 2021. B.1.617 surpassed Alpha as predominant lineage comprising 60% of samples in Delhi, as of April 2021. Immune evasion of Delta appears to be slightly less than Beta and Gamma. Transmissibility is 50% higher for Delta compared to Alpha, but no increase in case fatality rate (CFR). | [1] |
| New lineage with proposed name: GRL or B.1.1/ S:V1230L | Alpha (B.1.1.7), Gamma (P.1), Epsilon (B.1.429), Beta (B.1.351), Eta (B.1.525), Delta (B.1.617.2), Kappa (B.1.617.1), B.1.618 | West Bengal, India | N=2,000 sequences, from Jan 2021 to Mar 2021 | 70 out of 412 genomes sequenced from West Bengal belonged to this new variant. | [2] |
| Alpha (B.1.1.7) lineage | Beta (B.1.351) lineage | Telangana, India | N=93 sequences, as of Feb 2021 | 12 out of the 93 COVID-19 genomes sequenced showed detection of Alpha and Beta was not detected in any of the samples. | [3] |
| Alpha (B.1.1.7) and B.1.36 lineages | B.1, B.1.1.74, and B.1.468 lineages | Karnataka, India | N=176 sequences, from Nov 2020 to Jan 2021 | 24 out of 73 (32.9%) genomes sequenced that were international travelers to the region showed presence of Alpha (B.1.1.7), i.e. the predominant imported lineage. 45 out of the 103 genomes sequenced from cases within Bengaluru showed presence of B.1.36, i.e. the main lineage circulating. | [4] |
| B.1 lineage A2a (i.e. G and GH clades) | B.1.113 lineage | Kerala, India | N=113 sequences, from Feb to Dec 2020 | 110 of the 113 showed presence of B.1, making this lineage dominant in the sample from Kerala. All 113 genomes sequenced clustered within the A2a clade. | [5] |
| B.1.36 lineage (in Ahmedabad) | B.1 and B.6 lineages (in Ahmedabad) | Gujarat, India | N=502 sequences, as of Aug 18, 2020 | Among the 63 deceased cases out of 361, the dominant mutations were G25563T and C28854T in Gujarat, as detected in 47.62% and 35.16% of deceased cases, respectively. | [6] |
| G clade | L, S, and V clades | India and select states | N=696 sequences, from Jan to Aug 2020 | Between March to May 2020, GH became predominant clade in southern states, whereas GR became prevalent in northern states. | [7] |
| B.6, B.1 lineages A2a clade | B.1.1 and B.1.36 lineages A3 clade | India and select states | N=611 sequences, as of Jun 6, 2020 | B.6, B.1 and B.1.36 the lineages and the A2a clade appear to be the most common in India. | [8] |
| B.6 lineage | A, B, B.1, B.1.80, B.1.1, B.4 lineages | Karnataka, India | N=91 sequences, as of May 21, 2020 | 47 of the 91 SARS-CoV-2 genomes sequenced showed B.6 lineage. | [9] |

*Note*: Entries are in descending order from most to least recent.


[1] M. S. Dhar *et al.*, "Genomic characterization and Epidemiology of an emerging SARS-CoV-2 variant in Delhi, India," *medRxiv*, p. 2021.06.02.21258076, Jun. 2021, doi: 10.1101/2021.06.02.21258076.

[2] R. Sarkar *et al.*, "Emergence of a new SARS-CoV-2 variant from GR clade with a novel S glycoprotein mutation V1230L in West Bengal, India," *medRxiv*, p. 2021.05.24.21257705, May 2021, doi: 10.1101/2021.05.24.21257705.

[3] S. M. Ahmed, S. R. Juvvadi, R. Kalapala, and J. B. Sreemanthula, "Detection of SARS-CoV-2 N501Y mutation by RT-PCR to identify the UK and the South African strains in the population of South Indian state of Telangana," *medRxiv*, p. 2021.03.27.21254107, Mar. 2021, doi: 10.1101/2021.03.27.21254107.

[4] C. Pattabiraman *et al.*, "Importation, circulation, and emergence of variants of SARS-CoV-2 in the South Indian State of Karnataka," *medRxiv*, p. 2021.03.17.21253810, Mar. 2021, doi: 10.1101/2021.03.17.21253810.

[5] C. Radhakrishnan *et al.*, "Initial Insights Into the Genetic Epidemiology of SARS-CoV-2 Isolates From Kerala Suggest Local Spread From Limited Introductions," *Front. Genet.*, vol. 12, p. 630542, 2021, doi: 10.3389/fgene.2021.630542.

[6] M. Joshi *et al.*, "Genomic Variations in SARS-CoV-2 Genomes From Gujarat: Underlying Role of Variants in Disease Epidemiology," *Front. Genet.*, vol. 12, Mar. 2021, doi: 10.3389/fgene.2021.586569.

[7] P. D. Yadav *et al.*, "An Epidemiological Analysis of SARS-CoV-2 Genomic Sequences from Different Regions of India," *Viruses*, vol. 13, no. 5, May 2021, doi: 10.3390/v13050925.

[8] R. Laskar and S. Ali, "Phylo-geo-network and haplogroup analysis of 611 novel coronavirus (SARS-CoV-2) genomes from India," *Life Sci. Alliance*, vol. 4, no. 5, May 2021, doi: 10.26508/lsa.202000925.

[9] C. Pattabiraman *et al.*, "Genomic epidemiology reveals multiple introductions and spread of SARS-CoV-2 in the Indian state of Karnataka," *PLOS ONE*, vol. 15, no. 12, p. e0243412, Dec. 2020, doi: 10.1371/journal.pone.0243412.

#### Two-strain Model

The model described here builds on a previously published model of SARS-CoV-2 transmission introduced in Flaxman et al, 2020 (*7*), subsequently extended into a two-category framework in Faria et al, 2020 (*6*).

The model describes two categories, denoted $s \in \{1,2\}$. The population-unadjusted reproduction number for the first category is defined as

$$R_{s=1,t} = \mu_0 2\sigma(X_t)$$

Where $\mu_0$ is a scale parameter (3.3), $\sigma$ is a logistic function, and $X_t$ is an second-order autoregressive process with weekly time innovations, as specified in earlier work (*8*). The population-unadjusted reproduction number of the second category is modelled as

$$R_{s=2,t} = \rho 1_{[t_2,\infty)} R_{1,t}$$

With

$$\rho \sim \text{Gamma}(5,5) \in [0, \infty)$$

Where $\rho$ is a parameter defining the relative transmissibility of category 2 compared to category 1 and $1_{[t_2,\infty)}$is an indicator function taking the value of 0 prior to $t_2$, and 1 thereafter, highlighting that category 2 does not contribute to the observed epidemic evolution before its emergence.

Infections arise for each category according to a discrete renewal process (*9*, *10*)

$$i_{s,t} = \left(1 - \frac{n_{s,t}}{N}\right) R_{s,t} \sum_{\tau<t} i_{s,\tau} g_{t-\tau}$$

Where $N$ is the total population size, $n_{s,t}$ is the total extent of population immunity to category $s$ present at time $t$, and $g$ is the generation interval distribution.

The susceptible depletion term for category $s$ is modelled as

$$n_{s,t} = \sum_{\tau<t} i_{s,t} W_{t-\tau} + \beta\left(1 - \alpha_{s,t}\right) \sum_{\tau<t} i_{s,\tau} W_{t-\tau}$$

under assumptions of symmetric fixed cross-immunity $\beta$. In models presented here we run scenarios with fixed cross immunity ($\beta$) of 1 (100%), 0.75 (75%), and 0.5 (50%). $W_{t-\tau}$ is the time-dependent waning of immunity elicited by previous infection (*11*, *12*), which is modelled as a Rayleigh survival-type function with Rayleigh parameter of sigma = 310, which produces 50% of individuals still immune after 1 year. The cross-immunity susceptible term $\alpha_{s,t}$ is modelled as

$$\alpha_{s,t} = \frac{(1-\beta) \sum_{\tau<t} i_{s,\tau} W_{t-\tau}}{N - \beta \sum_{\tau<t} i_{s,\tau} W_{t-\tau}}$$

Infections in Maharashtra are seeded for six days at the start of the epidemic as

$$i_{1,t_{1..6}} \sim \text{Exponential}\left(\frac{1}{\tau}\right)$$

With

$$\tau \sim \text{Exponential}(0.03)$$

and the second category for one day ($t_2$=01-12-2020) as

$$i_{2,t_2} \sim \text{Normal}(1, 20) \in [1, \infty)$$

Non-unit seeding of the B.1.617.2 variant and the diffuse prior represent our uncertainty in the precise date and magnitude of B.1.617.2's introduction/importation into Maharashtra.

The model generates deaths via the following mechanistic relationship:

$$d_t = \sum_s \text{ifr}_s \sum_{\tau<t} i_{s,\tau}\pi_{t-\tau}$$

The infection fatality ratios ($\text{ifr}_s$) of each of the categories are given moderately informative priors:

$$\text{ifr}_s \sim \text{Normal}(0.25, 0.02^2) \in [0, 100]$$

allowing some variation in the IFR while suggesting it is unlikely to be less than 0.15 or greater than 0.3.

The observation model uses two types of data.

In the first, the likelihood for the expected deaths $D_t$ with an under-reporting factor of $\nu$, is modelled as negative-binomially distributed,

$$D_t \sim \text{NegativeBinomial}(d_t * \nu, d_t * \nu + \frac{(d_t * \nu)^2}{\phi})$$

with mortality data $d_t$ and dispersion prior

$$\phi \sim \text{Normal}(0,5) \in [0, \infty)$$

In this work we run scenarios with underreporting factors $\nu$ as 0.5 (50%) and 1 (no under reporting).

The second likelihood is based on genomic data from individuals where infections were sequenced and where the sequence was uploaded to GISAID. Specifically, the proportion of sequenced genomes identified as B.1.617.2 at time $t$ are modelled with a binomial likelihood

$$G_t^+ \sim \text{Binomial}(G_t^+ + G_t^-, \theta_t)$$

with positive counts for B.1.617.2 denoted $G_t^+$ and counts for lineages not belonging to B.1.617.2 recorded as $G_t^-$. The success probability for B.1.617.2 positivity is modelled as the infection ratio

$$\theta_t = \frac{\tilde{\iota}_{2,t}}{\tilde{\iota}_{1,t} + \tilde{\iota}_{2,t}}$$

Where $\tilde{\iota}_{s,t}$ is given by

$$\tilde{\iota}_{s,t} = \sum_{\tau \leq t} \iota_{s,t} \kappa_{t-\tau}$$

to account for the time varying PCR positivity displayed over the natural course of a COVID-19 infection. The distribution $\kappa$ describes the probability of being PCR positive over time following infection, and is based on (*13*).

**Genomic Data Input:**

In a short span of two months, the variant of concern B.1.617.2, has become the dominant strain in the state of Maharashtra. The rise of variant in the state has coincided with the second wave (**Supplementary Figure S1**). Genomic data was obtained from GISAID (*3*, *4*).

**Figure S1.** Lineage prevalence over time in Maharashtra, India.

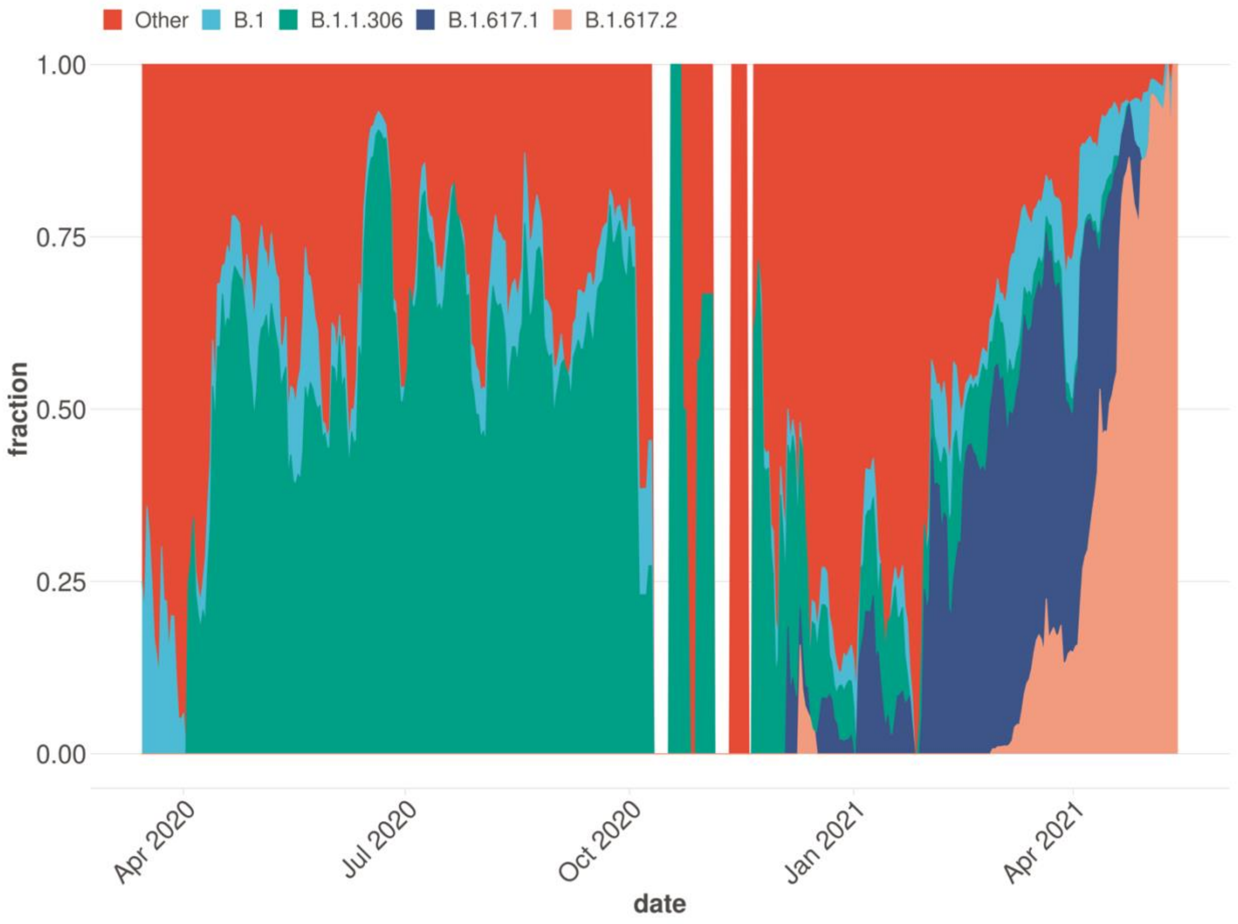

We consider two strains as: B.1.617.2 and non-B.1.617.2, where non-B.1617.2 is a mixture of previously circulating lineages other than B.1617.2.

All sequences are taken from GISAID (*3*, *4*) which have been uploaded for the state of Maharashtra. In total there are 4415 sequences in the dataset with 618 of them being labelled as B.1.617.2 for the period upto week starting 10$^{th}$ May 2021. In **Table S3** we summarize the distribution B.1.617.2 strain in genomes for the state of Maharashtra for each month from March 2020 to May 2021 (data for May includes data only till the week starting 10th May 2021). We can see from **Table S3** below**,** the astronomical rise of B.1.617.2 from mere 1.5% in Feb'21 to 87% in May'21.

**Table S3.** Distribution of B.1.617.2 strain genomic data from the State of Maharashtra (GISAID) for each month.

| Month | Total Samples | Total B.1.617.2 samples | Proportion of B.1.617.2 samples (%) |
|---|---|---|---|
| **March'20** | 53 | 0 | 0% |
| **April'20** | 130 | 0 | 0% |
| **May'20** | 255 | 0 | 0% |
| **June'20** | 297 | 0 | 0% |
| **July'20** | 255 | 0 | 0% |
| **August'20** | 201 | 0 | 0% |
| **September'20** | 205 | 0 | 0% |
| **October'20** | 18 | 0 | 0% |
| **November'20** | 81 | 0 | 0% |
| **December'20** | 141 | 1 | 0.71% |
| **January'21** | 112 | 0 | 0% |
| **February'21** | 769 | 12 | **1.5%** |
| **March'21** | 1251 | 190 | 15% |
| **April'21** | 562 | 341 | 61% |
| **May'21** | 85 | 74 | **87%** |

**Table S4**. Sensitivity analysis for B.1.617.2 strain under various scenarios of infection fatality rates (IFR), extent of underreporting of deaths and levels of protection from previous SARS-CoV-2 infections. The inferred changes in transmissibility increase of B.1.617.2 and estimated % deaths from B.1.617.2 between April 1-May 15, 2021, based on genomic data from the State of Maharashtra (GISAID) and daily death data from covidindia19.org are presented via the estimated posterior mean, with the Bayesian 95% credible interval (CI) in brackets. The results are derived from the stochastic renewal process model described in Faria et al. (*6*). The highlighted row corresponds to results presented in the main manuscript.

| Infection Fatality Rate | Under-Reporting of Deaths | Protection from previous SARS-CoV-2 Infections (%) | Transmissibility Increase from B.1.617.2* (95% CI) | Estimated % deaths from B 1.617.2 (April 1-May 15, 2021) |
|---|---|---|---|---|
| 0.25% | 0% | 100% | 1.78 (1.73-1.84) | 61 [15-91] |
| | 50% | 100% | 1.83 (1.77-1.90) | 55 [10-90] |
| | 0% | 75% | 1.69 (1.57-1.76) | 57 [11-90] |
| | 50% | 75% | 1.65 (1.59-1.71) | 61 [14-92] |
| | 0% | 50% | 1.63 (1.58-1.69) | 63 [16-92] |
| | 50% | 50% | 1.49 (1.44-1.45) | 64 [16-92] |
| 0.15% | 0% | 100% | 1.82 (1.76-1.88) | 59 [13-91] |
| | 50% | 100% | 1.87 (1.82-1.94) | 65 [19-90] |
| | 0% | 75% | 1.67 (1.61-1.73) | 60 [12-91] |
| | 50% | 75% | 1.46 (1.41-1.52) | 61 [14-90] |
| | 0% | 50% | 1.54 (1.50-1.60) | 63 [16-91] |
| | 50% | 50% | 1.20 (1.15-1.25) | 61 [12-91] |

*Inferred transmissibility advantage for B.1.617.2 is against the background of a mixture of all previous circulating variants in the state of Maharashtra. A number 1.5 means 50% increase in transmissibility.

#### **Section 3**. Methods for Characterizing the Potential Effect of Public Health Interventions

A comprehensive list of interventions introduced in India is provided in **Supplementary Table S4.** To estimate the impact of a potential public health intervention on the number of reported cases and deaths during the second wave of the COVID-19 pandemic in India, we implement a modified version of the traditional susceptible-infected-recovered (SIR) model, called an extended SIR (or eSIR), which allows for a time-varying effective reproduction number ($R_t$). This model was developed by Wang et al. (*14*) to study the COVID-19 outbreak in China and was made available in the eSIR R package (*15*).

Specifically, we use a modified version of the tvt.eSIR function from the eSIR R package with a modification that incorporates the possibility of re-infection and waning immunity over time. This re-infection could be from the previously circulated lineages/original wild strain or the newly emerging variants of interest/concern. The modified extended SIR model framework, depicted in **Figure S2**, is presented as follows:

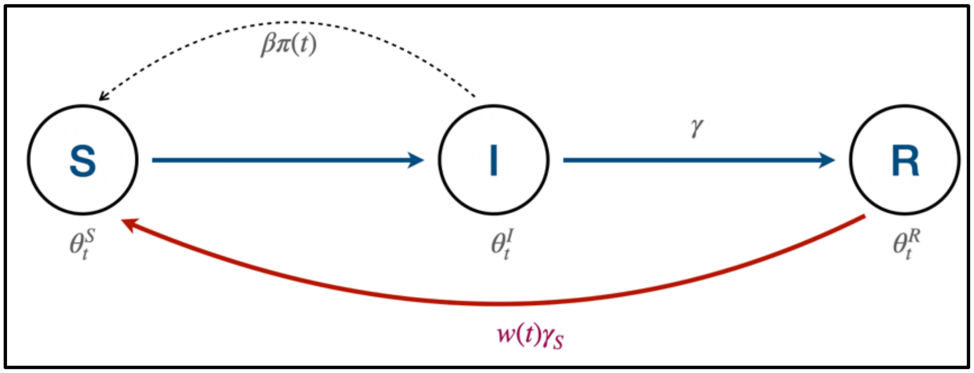


**Figure S2**. The compartments and transitions for the modified eSIR model with re-infection and waning immunity (red arc) incorporated.

The eSIR model works by assuming that the true underlying probabilities of the three compartments follow a latent Markov transition process, and that we only observe the daily proportions of infected cases and removed. First, let us set up some notations. Assume that the observed proportions of infected and removed cases on day $t$ are denoted by $Y_t^I$ and $Y_t^R$, respectively. Further, denote the true underlying probabilities of the S, I, and R compartments on day $t$ by $\theta_t^S$, $\theta_t^I$, and $\theta_t^R$, respectively, and assume that for any $t$, $\theta_t^S + \theta_t^I + \theta_t^R = 1$. Based on the modifications made with respect to the original eSIR model, this model can be described by the following set of differential equations.

$$f(\boldsymbol{\theta}_t, \beta, \gamma): \quad \frac{d\theta_t^S}{dt} = -\beta\pi(t)\theta_t^S\theta_t^I + \boldsymbol{w(t)\gamma_s\theta_t^R},$$

$$\frac{d\theta_t^I}{dt} = \beta\pi(t)\theta_t^S\theta_t^I - \gamma\theta_t^I,$$

$$\frac{d\theta_t^R}{dt} = \gamma\theta_t^I - \boldsymbol{w(t)\gamma_s\theta_t^R},$$

where $\boldsymbol{\theta}_t = (\theta_t^S, \theta_t^I, \theta_t^R)$. Here, $\beta > 0$ denotes the disease transmission rate, $\gamma > 0$ denotes the removal rate, $\gamma_s > 0$ denotes the re-infection rate. The basic reproduction number $R_0 := \frac{\beta}{\gamma}$ indicates the expected number of cases generated by one infected case in the absence of any intervention and assuming that the whole population is susceptible. At this stage, for the observed infected and removed proportions, we assume a Beta-Dirichlet state-space model, independent conditionally on the underlying process:

$$Y_t^I | \boldsymbol{\theta_t}, \boldsymbol{\tau} \sim Beta\left(\lambda^I \theta_t^I, \lambda^I (1 - \theta_t^I)\right),$$
$$Y_t^R | \boldsymbol{\theta_t}, \boldsymbol{\tau} \sim Beta\left(\lambda^R \theta_t^R, \lambda^R (1 - \theta_t^R)\right).$$

Further, the Markov process on the latent proportions is built as:

$$\boldsymbol{\theta_t} | \boldsymbol{\theta_{t-1}}, \boldsymbol{\tau} \sim Dirichlet\big(\kappa f(\boldsymbol{\theta_{t-1}}, \beta, \gamma)\big)$$

where $\boldsymbol{\theta_t}$ denotes the vector of the underlying population probabilities of the three compartments, whose mean is modeled as an unknown function of the probability vector from the previous time point, along with the transition parameters; $\boldsymbol{\tau} = (\beta, \gamma, \boldsymbol{\theta_0^T}, \lambda, \kappa)$ denotes the whole set of parameters where $\lambda^I, \lambda^R$ and $\kappa$ are parameters controlling variability of the observation and latent process, respectively. The function $f(.)$ is then solved as the mean transition probability determined by the SIR dynamical system, using a fourth order Runge-Kutta approximation.

We then follow the same MCMC scheme as in the original eSIR package, with added priors and hyperparameters for the additional parameter $\gamma_s$ (re-infection rate) and information to compute $w(t)$ (time-varying waning immunity rate). Specifically, our priors are as follows: $\boldsymbol{\theta}_0 \sim$ Dirichlet$(1 - Y_0^I - Y_0^R, Y_0^I, Y_0^R)$ ($Y_0^X$ denotes the observed proportions in compartment X on the first day when data is available), $R_0 \sim$ LogNormal$(0.582, 0.223)$ (so that the mean reproduction number $E(R_0) = 2$, and $SD(R_0) = 1$), $\gamma \sim$ LogNormal$(-2.955, 0.910)$ (so that the average infectious period is $\frac{1}{E(\gamma)} = \frac{1}{0.082} \approx 12$ days, and $SD(\gamma) = 0.1$) (*16*), $\beta = R_0\gamma$, $\gamma_s \sim$ LogNormal$(-8.660\,,\ 6.358)$ (so that average immunity period is $\frac{1}{E(\gamma_s)} \approx 8$ months, and $SD(\gamma_s) = 0.1$) (*11*). The prior mean of the basic reproduction number, 2.0, is approximately the average of the estimates from many other (wave 1) COVID-19 studies on the Indian population (*17–21*). The prior mean of the removal rate $\gamma$ indicates an average infectious period of 12 days, which is originally set using the estimation from SARS outbreak in Hong Kong (*22*) due to the similarity between the two viruses; and this value also aligns well with a few 2020 studies on COVID-19 in China (*23–25*). The prior mean for the average immunity period is based on a recent study by Hansen et al., 2021 (*11*).

For $w(t)$, our modified tvt.eSIR function allows the choices of $w(t) = 0\ \forall t$ (no waning immunity, reverts back to the original eSIR), a discrete time-varying vector of proportions (following the same structure as pi(t)), or a continuous function. For cases in this work where we want to incorporate waning immunity, we use a continuous function to better capture it over time. Specifically, following Faria et al., 2021, we model time-dependent waning of immunity as a Rayleigh distribution ($\sigma = 310$) to ensure 50% individuals are still immune after 1 year (*6*). The rest of the settings for the model and the MCMC scheme remain the same as the original eSIR package, and a detailed summary of these can be found in Ray et al., 2020 (*16*). Here, we briefly describe these for the sake of completeness.

For the variability parameters, the default choice is to set large variances in both observed and latent processes, which may be adjusted over the course of epidemic with more data becoming available.

$$\kappa,\ \lambda^I, \lambda^R \sim \ iid\ Gamma(2,\ 0.0001).$$

Denoting $t_0$ as the last date of data availability, and assuming that the forecast spans over the period $[t_0 + 1, T]$, our MCMC algorithm is as follows.

0. Take $M$ draws from the posterior $[\boldsymbol{\theta}_{1:t_0}, \boldsymbol{\tau} | \boldsymbol{Y}_{1:t_0}]$.
1. For each solution path $m \in \{1, \dots, M\}$, iterate between the following two steps via MCMC.
   i. Draw $\boldsymbol{\theta}_t^{(m)}$ from $\left[\boldsymbol{\theta}_t \middle| \boldsymbol{\theta}_{t-1}^{(m-1)}, \boldsymbol{\tau}^{(m)}\right], t \in \{t_0 + 1, \dots, T\}$.
   ii. Draw $\boldsymbol{Y}_t^{(m)}$ from $\left[\boldsymbol{Y}_t \middle| \boldsymbol{\theta}_t^{(m)}, \boldsymbol{\tau}^{(m)}\right], t \in \{t_0 + 1, \dots, T\}$.

We chose to present models where $w(t) = 0$ in the main text because the results are virtually identical and the model with this setting is simpler. Results with waning immunity are presented in **Section 3.1**. The modified version of the tvt.eSIR function used in our analyses can be accessed on the Center for Precision Health Data Science GitHub page here: https://github.com/umich-cphds/cov-ind-19/blob/master/other/tvt.eSIR.R.

We are able to simulate the effect of hypothetical nationwide lockdowns at various time points to assess the impact of a hypothetical intervention in this model by modifying $\beta$ by a time-varying modifier "pi schedule" (the $\pi(t)$ term in **Figure S2**). For a given intervention schedule we use the model to project the case counts over a future time window *[T+1, T+W]* using observed data up to time *T*. We incorporate the effect of a lockdown on these projections via incorporating a time-varying effect modifier "pi schedule". Namely, we impose a time-varying schedule $\pi(t)$= $R_t/R_0$, which represents the proportional change in $R_t$ over time under a given lockdown scenarios.

The primary challenge for obtaining a realistic lockdown intervention is the choice of an appropriate underlying intervention schedule or "pi schedule" in the eSIR modeling vocabulary. We make use of two actual intervention scenarios from India from which we derive a realistic pi schedule, namely: (1) the national lockdown beginning on March 25, 2020 in response to the first wave, a **strong lockdown effect**, and (2) the Maharashtra lockdown beginning on April 14 2021 in response to the second wave, **a moderate lockdown effect**. **Figure S3A** depicts the two pi schedules used to represent the impact of interventions. These adaptively estimated pi schedules capture the impact of a lockdown, representing the relative reduction of $R_t$ over time that was observed in India and in Maharashtra in 90 days following a lockdown. To compute the pi schedules, the effective time varying reproduction number $R_t$ is estimated using the estimate_R function from the EpiEstim R package ("parametric_si" method with mean_si = 7 and std_si = 4.5)
.

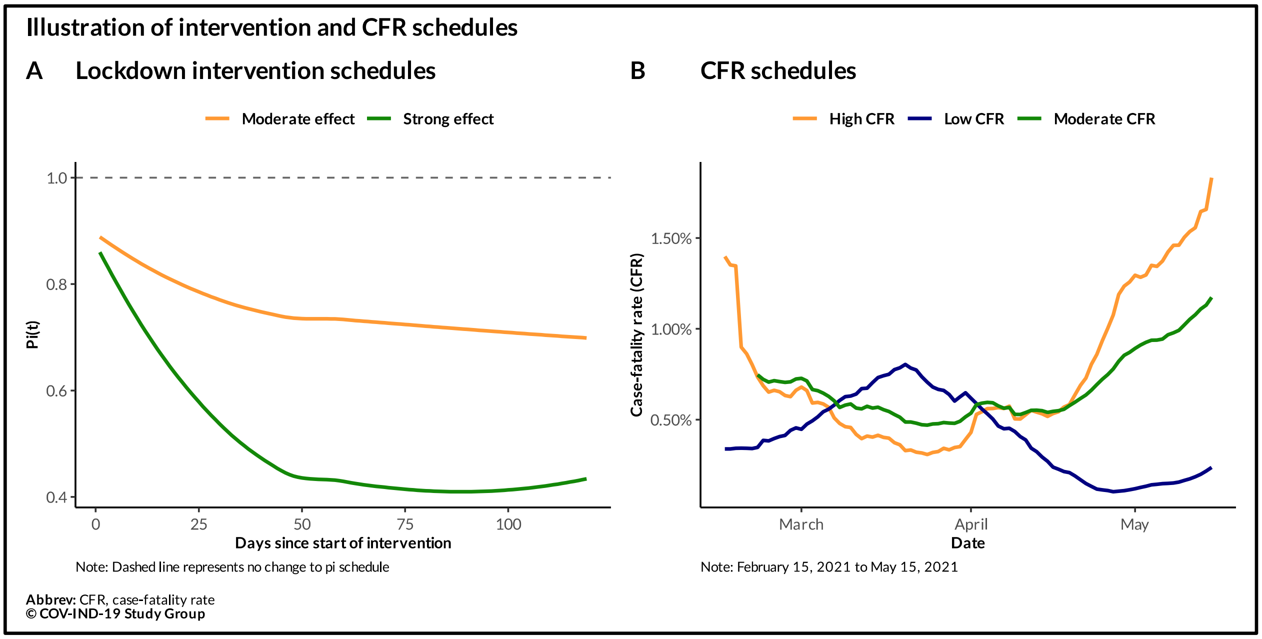


**Figure S3**. The lockdown intervention schedules $\pi(t)$for strong (green) and moderate (saffron) effects are presented in panel A. The strong effect represents a smoothed ratio of the estimate effective reproduction number, $R_t$, after the implementation of the nationwide lockdown in India, initiated on March 25, 2020. The moderate effect is derived from the Maharashtra interventions that began on April 14, 2021 (last observation carried forward to extend it to 120 days). The case-fatality rate (CFR) schedules for high (saffron), moderate (green), and low (navy) CFR using observed data from February 15, 2021, through May 15, 2021, are shown in panel B. These are trailing 7-day averages of the observed CFR in Maharashtra, India, and Kerala, respectively.

The India schedule is estimated from the nationwide lockdown which began during Wave 1, starting March 25, 2020. This lockdown was swift and had relatively strong immediate and long-term reductions in $R_t$ and represents a best-case or optimistic scenario (**Figure S3A**). The Maharashtra schedule is estimated from the statewide interventions which began during Wave 2 starting April 14, 2021. This lockdown had a more subdued impact on $R_t$ relative to the national lockdown in the previous year and represents what we consider to be a moderate intervention scenario. We construct the respective schedules as the ratio of the $R_t$ following the lockdown over the $R_t$ from the day the lockdown started, where estimates for each effective reproduction number were obtained from the previously detailed eSIR model. The schedule is smoothed using loess to account for day-to-day variations in $R_t$.

Because $R_t$ first crossed unity (i.e., rose above 1) in mid-February, an early warning sign of a forthcoming surge in cases, we assess lockdowns had they been implemented on March 1, March 15, March 30, April 15, and April 30, 2021. We project each scenario out to May 15 and compare the forecasted results to observed, reported counts. For example, for the March 15 start date of a lockdown, we apply the pi schedule from March 15 till May 15, a 60-day period.

We can also estimate the number of deaths under these forecasting scenarios under a hypothetical intervention. To do this, we apply a case-fatality rate (CFR) schedule, which was calculated as the observed, trailing 7-day average CFR (i.e., ratio of daily reported deaths to daily reported cases) for India (moderate CFR), for Maharashtra (high CFR), and for Kerala (low CFR) through May 15. We include the Kerala CFR schedule because of their low COVID-19 CFR relative to other large states in India and their robust healthcare system, which can be interpreted as a best-case scenario. The CFR schedules are based on observed data from February 15, 2021, through May 15, 2021, and are shown in **Figure S3B**. We then multiply the projected case counts by the CFR schedule to obtain an estimate of the daily number of COVID-19 deaths, under the presented lockdown scenarios. We then sum up the estimated daily deaths by summing up the daily incident cases.

**Figure S4** shows the estimated death projections for the three CFR schedules under a lockdown with moderate effect. This figure is analogous to Figure 4 in the main text (death projections under a lockdown with strong effect).

**Credible intervals:** One major advantage of the Bayesian implementation is that uncertainty associated with all parameters and functions of parameters can be calculated from exact posterior draws without relying on large-scale approximation or delta theorem. The credible intervals (CrI) for the prevalence and incidence are computed using the posterior distribution of proportions given the observed confirmed and removed prevalence, i.e. $Y^I_{(t_0+1):T} | Y^I_{1:t_0}, Y^R_{1:t_0}$ and $Y^R_{(t_0+1):T} | Y^I_{1:t_0}, Y^R_{1:t_0}$, where $t_0$ denotes the last observed date, and $T$ denotes the last forecast date. More specifically, suppose we want to compute the 95% posterior CrI for the observed proportion of confirmed cases on the first day of forecast, i.e., a CrI for the random variable $Y^I_{t_0+1}$. Then, from the $M$ solution paths of the posterior, we have the draws $\left\{Y^{I\,(m)}_{t_0+1}, 1 \le m \le M\right\}$. We construct a 95% posterior CrI for $Y^I_{t_0+1}$ by simply computing the 2.5th upper and lower quantiles from this set of M draws. The cumulative prevalence is sums of the draws from the I and R compartments at a given time, whereas the incidences are the sums of successive differences in these two compartments – thus the confidence interval for these can be calculated in a similar way. Case-counts can be obtained from prevalence by using population size. Similar techniques apply to $\theta^I_{t_0+j}$ (for quantifying uncertainties for the true counterparts to the observed prevalences and incidences) for any $1 \le j \le T - t_0$ and transmission parameters like $\beta$, $\gamma$ and $\gamma_S$. For instance, a 95% posterior CrI for $\beta$ can be constructed by calculating the 2.5th upper and lower quantiles of $\left\{\beta^{(m)}, 1 \le m \le M\right\}$. Therefore, we could simply define $R^{(m)} = \frac{\beta^{(m)}}{\gamma^{(m)}}\ \forall\ 1 \le m \le M$, and get the 95% posterior CrI for the effective reproduction number $R$ from $\left\{R^{(m)}, 1 \le m \le M\right\}$.

**Estimating the cases and deaths averted due to intervention:** We evaluate the intervention scenarios with different dates of introduction to illustrate the effect of timing of interventions. Let the start of the non-pharmaceutical public health intervention (NPI) be at day t (March 15, 30 and April 15 in our example) and the end of the follow-up period be denoted by t+T (May 15 in our case, where T is the number of days in the follow-up period where NPI takes place. We introduce the following notations.

$D_s$ = Observed total reported deaths on day $s$ , $C_s$ = Observed total reported cases on day $s$
$dcfr_s$ = observed CFR at time $s$, $\widehat{Y_s}$ = the predicted daily new case count on day *s* while $\widehat{P_s}$ is the predicted total case count on day *s*.

**For Cases we report:**

- Observed cases in period [t+1, t+T]: (1) $(C_{t+T} - C_t)$
- Predicted cases in period [t+1, t+T]: (2) $\sum_{s=t+1}^{T} \widehat{Y_s} = (\widehat{P_{t+T}} - \widehat{P_t})$
- Cases averted up to time t+T by introducing an intervention at time t:

  (3)= (1)-(2): $(C_{t+T} - C_t) - (\sum_{s=t+1}^{T} \widehat{Y_s})$
- Reduction in cases by introducing the intervention at time t:

  (Cases averted in [t+1, t+T]/Cases observed in [t+1, t+T])*100 or (4)= (3)/(1)*100

**For deaths we report:**

- Observed deaths in period [t+1, t+T]: (1) $(D_{t+T} - D_t)$
- Predicted deaths in period [t+1, t+T]: (2) $\sum_{s=t+1}^{T} (dcfr_s * \widehat{Y_s})$
- Deaths averted up to time *t+T* by introducing an intervention at time *t:*

  (3) = (1)-(2): $(D_{t+T} - D_t) - \sum_{s=t+1}^{T} (dcfr_s \ \widehat{Y_s})$
- Reduction in deaths by introducing the intervention at time t:

  (Deaths averted in [t+1, t+T]/Deaths observed in [t+1, t+T])*100 or (4)= (3)/(1)*100

We obtain the credible intervals from the MCMC draws of the daily number of cases predicted by the eSIR model and ignore the uncertainty in the observed CFR estimates used as weights to obtain the credible intervals for each of the above quantities which are linear combination of the posterior draws of predicted daily new cases.

**Table S5.** Timeline of COVID-19 interventions in India during the first and second waves.

| ***Wave 1: March 24, 2020 – February 14, 2021[a,b]*** | |
|---|---|
| ***Date*** | ***Intervention*** |
| Mar 3, 2020[1] | ◆ India issues travel ban on four countries - China, South Korea, Italy, and Iran |
| Mar 6, 2020 | ◆ Union Health Ministry issues advisory to avoid mass gatherings |
| Mar 7, 2020 | ◆ Mayor of Agra urges the Union government to close down historical monuments including Taj Mahal<br>◆ Kuwait suspends flights to India |
| Mar 9, 2020 | ◆ Qatar puts India on travel ban list |
| Mar 10, 2020 | ◆ Manipur closes its border with Myanmar |
| Mar 11, 2020 | ◆ India suspends all visas/e-visas granted to nationals of France, Germany, and Spain on or before today |
| Mar 12, 2020 | ◆ WHO declares the COVID-19 outbreak as 'pandemic'<br>◆ India suspends all visas excepting those for diplomatic, UN, or international bodies, official and employment purposes until April 15<br>◆ India reports 1st death |
| Mar 13, 2020 | ◆ India reports 2nd death<br>◆ Several academic institutions (e.g., JNU, IIT, IIM) cancel classes/convocations; some hostels close |
| Mar 16, 2020 | ◆ Central government proposes social distancing measures until March 31<br>◆ India bans passengers from EU countries, UK, and Turkey until March 31<br>◆ Central government recommends closure of educational institutions until March 31 |
| Mar 17, 2020 | ◆ Taj Mahal is shut until March 31; ASI closes 3,000 monuments and 200 museums<br>◆ Mandatory quarantine is imposed on passengers coming from UAE, Qatar, Oman, and Kuwait<br>◆ India is heading toward a countrywide lockdown mode |
| Mar 19, 2020 | ◆ India halts all incoming commercial international flights for one week<br>◆ Some state governments ban public transportation<br>◆ Prime Minister urges people of India to observe self-imposed curfew ('Janata Curfew') on March 22 |
| Mar 20, 2020 | ◆ Maharashtra announces lockdown in Mumbai, Nagpur, and Pune<br>◆ Jawaharlal Nehru University (JNU) in Delhi orders students to vacate hostels |
| Mar 21, 2020 | ◆ Private labs can conduct COVID-19 tests, says Maharashtra government<br>◆ Rajasthan government declares lockdown until March 31 |
| Mar 22, 2020 | ◆ 12 states, including Telangana and Delhi, announce lockdown until March 31<br>◆ International commercial passenger flights are disallowed to land in India for one week starting today<br>◆ Railways suspend all train services until March 31 |
| Mar 23, 2020 | ◆ Central government orders all states in India to impose lockdown<br>◆ Legal action is to be initiated against people violating lockdown measures |
| Mar 24, 2020 | ◆ Prime Minister of India announces lockdown for 21 days as country records 552 COVID-19 cases and 10 deaths |
| Mar 28, 2020 | ◆ Central government unveils stimulus package to help those hit by 21-day lockdown<br>◆ Priorities are to construct COVID-19 hospitals, sample testing, contact-tracing, and social distancing: Union Health ministry |
| Apr 2, 2020 | ◆ Common exit strategy necessary for 'staggered' relaxations after lockdown period ends, Prime Minister tells chief ministers |
| Apr 6, 2020 | ◆ Prime Minister instructs union ministers to prepare a graded plan to gradually open departments that are not COVID-19 hotspots |
| Apr 8, 2020 | ◆ Prime Minister and chief ministers decide on lockdown extension to April 11 |
| Apr 9, 2020 | ◆ Odisha extends lockdown until April 30 and becomes first Indian state to do so |
| Apr 11, 2020 | ◆ Prime Minister, after a meeting with chief ministers, extends lockdown by two weeks |
| Apr 14, 2020 | ◆ Prime Minister extends nationwide lockdown until May 3, and stated that states that could avoid being the hotspots may be allowed to resume important activities<br>◆ COVID-19 testing kits expected to last for six weeks, and additional RT-PCR kits have been ordered<br>◆ Railways to refund online customers for train cancellations through May 3, and those booked at the counters can collect refunds through July 31 |

| | |
|---|---|
| Apr 15, 2020 | Railways and air travel remain suspended through May 3<br>Central government issues new guidelines for second phase of countrywide lockdown<br>Select activities, e.g., agriculture and industries in rural areas, allowed after April 20 |
| Apr 16, 2020 | Railways mandate all officers above Deputy secretaries to be in-person at office from Monday<br>India supplies Hydroxychloroquine to fifty-five countries |
| Apr 17, 2020 | Rajasthan is first state to begin COVID-19 rapid testing<br>Andhra Pradesh secures 100,000 test kits from South Korea<br>Maharashtra prohibits house owners from collecting rent for three months |
| Apr 19, 2020 | Telangana extends lockdown until May 7 and Karnataka extends lockdown until April 21<br>Delhi has 79 containment zones with 3 newly added zones |
| Apr 22, 2020 | Karnataka relaxes rules of lockdown beginning April 23<br>Prepares rapid response teams for Bangladesh, Bhutan, Sri Lanka and Afghanistan |
| Apr 23, 2020 | Telangana extends home quarantine from 14 to 28 days<br>Railways suspend passenger trains until May 3<br>Agricultural activities resume in much of Arunachal Pradesh and Karnataka relaxes restrictions |
| Apr 24, 2020 | Police arrest 137 people for violating mask mandate in Guwahati, Assam<br>Odisha announces 60-hour shutdown in 3 districts<br>AIIMS announces plasma use from recovered patients has begun |
| Apr 28, 2020 | Prime Minister says India's pharma production will be available to aid in the worldwide COVID-19 effort<br>Andhra Pradesh initiates death audit committee<br>Delhi has a total of 100 containment zones |
| Apr 29, 2020 | Srinagar and Kerala mandate face masks after May 1 and from tomorrow, respectively<br>States require quarantine for travelers |
| Apr 30, 2020 | Bihar carries out door-to-door screening in hotspot districts<br>Gurugram extends restrictions on intra-district movement |
| May 5, 2020 | Countrywide lockdown has been extended until May 17 with Telangana extending until May 29 |
| May 7, 2020 | Assam mandates institutional quarantine for those returning from red zone |
| Jun 1, 2020[2] | Central government announces guidelines for first phase of easing countrywide lockdown, referred to as Unlock 1 |
| Jul 1, 2020 | Central government announces guidelines for second phase of lifting countrywide lockdown, referred to as Unlock 2.0 |
| Jul 15, 2020 | Covaxin, an Indian-based COVID-19 vaccine, enters phase I of clinical trials |
| Jul 25, 2020 | Central government issues guidelines for third phase of lifting countrywide lockdown restrictions, referred to as Unlock 3.0 |
| Aug 26, 2020 | Covishield, an Oxford-AstraZeneca vaccine, enters clinical trials |
| Aug 29, 2020 | Central government announces guidelines for fourth phase of lifting countrywide lockdown, referred to as Unlock 4.0 |
| Sep 30, 2020 | Central government announces guidelines for fifth phase of lifting countrywide lockdown, referred to as Unlock 5.0 |
| Oct 5, 2020 | Central government states that 200-250 million people will be vaccinated by July 25, 2021 |
| Oct 26, 2020 | Central government requests preparation of 3-tier approach to vaccine program |
| Nov 10, 2020 | Delhi experiences third wave of COVID-19 pandemic |
| Dec 21, 2020 | India imposes air travel ban on flights from the United Kingdom |
| Dec 29, 2020 | Six passengers from the United Kingdom tested positive for new B.1.1.7 lineage of SARS-CoV-2 |
| Dec 30, 2020 | Numerous cities issue a night curfew |
| Jan 16, 2021 | Nationwide vaccination program begins with initial prioritization of healthcare workers[3] |
| Jan 19, 2021 | Lakshadweep, an island territory off the coast of Kerala, is the last region in India to report first case[4] |
| ***Wave 2: February 15, 2021 – May 31, 2021[a]*** | |
| ***Date*** | ***Intervention*** |
| April 1, 2021 | Central government extends vaccine eligibility to adults aged ≥ 45, as announced on March 23[5] |
| April 5, 2021 | Maharashtra issues restrictions on malls, restaurants, religious buildings, and cinemas, as announced on April 4[6] |

| | |
|---|---|
| April 8, 2021 | ◆ Six states at least, including Andhra Pradesh, Chhattisgarh, Haryana, Maharashtra, Odisha and Telangana, report vaccine shortages[7]<br>◆ Madhya Pradesh imposes night curfew in all urban areas, as announced on April 7[8] |
| April 9, 2021 | ◆ Jammu and Kashmir imposes night curfew in urban regions across eight districts, as announced April 8[9]<br>◆ Maharashtra suspends vaccine administration in multiple districts[10]<br>◆ Chhattisgarh district of Raipur issues lockdown until April 19, as announced on May 7[11] |
| April 10, 2021 | ◆ Maharashtra enters statewide weekend lockdown, as announced on April 4[6], which was extended until June 15[12]<br>◆ Karnataka imposes night curfew until April 20, as announced on April 9[13] |
| April 16, 2021 | ◆ Delhi imposes a weekend curfew, as announced on April 16[14]<br>◆ Karnataka imposes restrictions on public gatherings and entertainment activities, as announced on April 16[13] |
| April 19, 2021 | ◆ Delhi imposes a week-long lockdown until April 26, as announced on April 19[14], which was extended until June 7[12] |
| April 20, 2021 | ◆ Telangana issues an immediate statewide night curfew, as announced on April 20[15] |
| April 22, 2021 | ◆ Jharkhand imposes lockdown-like restrictions until April 29, as announced on April 20[16] |
| April 23, 2021 | ◆ Puducherry imposes statewide lockdown, as announced on April 21[17], which was extended until June 7[12] |
| April 24, 2021 | ◆ Andhra Pradesh issues a statewide night curfew, as announced on April 23[18]<br>◆ Uttar Pradesh issues a statewide weekend lockdown, as announced on April 20[19], which was extended until June 1[12] |
| April 27, 2021 | ◆ Assam imposes statewide night curfew through May 7, as announced on April 27[20]<br>◆ Himachal Pradesh announces night curfew across 1/3 of all districts and issues weekend lockdown, as announced on April 25[21]<br>◆ Karnataka imposes a two-week statewide close down, as announced on April 26[22] |
| April 30, 2021 | ◆ Haryana issues a weekend curfew across nine districts, as announced on April 30[20]<br>◆ West Bengal issues restrictions including a ban on gatherings as well as shutdown of cinemas, gyms, malls and restaurants, as announced on April 30[20], which were extended until June 15[12]<br>◆ Nagaland enters statewide lockdown until May 14, as announced on April 27[23], which was extended until June 11[24] |
| May 1, 2021 | ◆ Central government extends vaccine eligibility to adults aged ≥ 18[25] |
| May 3, 2021 | ◆ Haryana imposes a statewide lockdown through May 10, as announced on May 3[20], which was extended until June 7[12]<br>◆ Select districts in Mizoram, including Aizawl, enter an eight-day lockdown, as announced on May 2[26], which was extended until June 6[27]<br>◆ Punjab issues a weekend lockdown and a night curfew through May 15, as announced on May 3[28], which was extended until June 10[12] |
| May 5, 2021 | ◆ Bihar issues statewide lockdown until May 15, as announced on May 4[20], which was extended until June 8[12]<br>◆ Odisha enters a 14-day statewide lockdown until May 19, as announced on May 2[29], which was extended until June 17[30]<br>◆ Andhra Pradesh enters a statewide 14-day partial curfew, as announced on May 3[31], which was extended until June 10[12] |
| May 7, 2021 | ◆ Gujarat issues night curfew on twenty-nine cities, and travel restrictions as well as social distancing measures through April 30, as announced May 7[32], which was extended until June 4[12] |
| May 8, 2021 | ◆ Kerala enters a statewide lockdown until May 16, as announced on May 6[20], which was extended until June 9[12] |
| May 9, 2021 | ◆ Goa imposes a statewide curfew until May 23, as announced on May 7[20] |
| May 10, 2021 | ◆ Karnataka imposes a statewide lockdown until May 24, as announced on May 7[20], which was extended until June 7[12]<br>◆ Rajasthan enters a statewide lockdown until May 24, as announced on May 6[20], which was extended to June 2[12]<br>◆ Tamil Nadu issues a statewide lockdown until May 24, as announced on May 8[20], which was extended until June 7[12] |
| May 12, 2021 | ◆ Telangana imposes statewide lockdown, as announced on May 11 [33], which was extended until June 9 [12] |

[a] Wave 1 is defined as starting from March 24, 2020, when the first nationwide lockdown was implemented in India. Wave 2 is defined as starting from February 15, 2021, when the national effective reproduction number for COVID-19 in India crossed unity.

[b] Entries for Wave 1 are updated from Table 1 in Ray et al., 2020 (Ray, D., Salvatore, M., Bhattacharyya, R., Wang, L., Du, J., Mohammed, S., … Mukherjee, B. (2020). Predictions, Role of Interventions and Effects of a Historic National Lockdown in India's Response to the COVID-19 Pandemic: Data Science Call to Arms. *Harvard Data Science Review*. doi.org:10.1162/99608f92.60e08ed5).

[1] Shalini Nair, "Covid-19 pandemic in India updates: Coronavirus status by city and state," *Pharmaceutical Technology*, Feb. 05, 2021. https://www.pharmaceutical-technology.com/news/india-covid-19-coronavirus-updates-status-by-state/ (accessed Jun. 07, 2021).

[2] Shuja Asrar, "Coronavirus calendar: How the Covid pandemic unfolded across India in 2020," *The Times of India*, Jan. 01, 2021. Accessed: Jun. 07, 2021. [Online]. Available: https://timesofindia.indiatimes.com/india/coronavirus-india-timeline/articleshow/80030867.cms

[3] "Coronavirus | World's largest vaccination programme begins in India on January 16," *The Hindu*, New Delhi, Jan. 15, 2021. Accessed: Jun. 07, 2021. [Online]. Available: https://www.thehindu.com/news/national/coronavirus-worlds-largest-vaccination-programme-begins-in-india-on-january-16/article33582069.ece

[4] N. C. Sharma, "Lakshadweep reports its first covid-19 case," *mint*, Jan. 19, 2021. Accessed: Jun. 07, 2021. [Online]. Available: https://www.livemint.com/news/india/lakshadweep-reports-its-first-covid-19-case-11611070751242.html

[5] "India expands vaccination drive, all above 45 years to be vaccinated from April 1," *Mumbai Mirror*, Mar. 23, 2021. Accessed: Jun. 07, 2021. [Online]. Available: https://mumbaimirror.indiatimes.com/coronavirus/news/covid-19-india-expands-vaccination-drive-all-above-45-years-to-be-vaccinated-from-april-1/articleshow/81649964.cms

[6] PTI, "COVID-19: Weekend lockdown in Maharashtra, stricter curbs from Monday," *The New Indian Express*, Apr. 04, 2021. Accessed: Jun. 07, 2021. [Online]. Available: https://www.newindianexpress.com/nation/2021/apr/04/covid-19-weekend-lockdown-in-maharashtra-stricter-curbs-from-monday-2285721.html

[7] "Odisha To Maharashtra: Six States Complain Of Vaccine Shortage, Centre Says 'No Scarcity Anywhere'," *Outlook*, Apr. 08, 2021. Accessed: Jun. 07, 2021. [Online]. Available: https://www.outlookindia.com/website/story/india-news-odisha-to-maharashtra-six-states-complain-of-vaccine-shortages-centre-says-no-scarcity-anywhere/379615

[8] "Madhya Pradesh govt announces night curfew amid Covid spike. Check details," *mint*, Apr. 07, 2021. Accessed: Jun. 07, 2021. [Online]. Available: https://www.livemint.com/news/india/madhya-pradesh-govt-announces-night-curfew-in-urban-areas-check-details-11617808196014.html

[9] "Night Curfew In Urban Areas Of 8 J&K Districts Amid Surge In Covid Cases," *NDTV.com*, Apr. 08, 2021. Accessed: Jun. 07, 2021. [Online]. Available: https://www.ndtv.com/india-news/night-curfew-in-urban-areas-of-8-districts-of-jammu-and-kashmir-amid-rising-covid-19-cases-2409420

[10] Julia Hollingsworth, Melissa Macaya, Melissa Mahtani, Veronica Rocha and Fernando Alfonso III, "The latest on the coronavirus pandemic and vaccines," *CNN*, Apr. 09, 2021. Accessed: Jun. 07, 2021. [Online]. Available: https://edition.cnn.com/world/live-news/coronavirus-pandemic-vaccine-updates-04-09-21/h_b617a849bbb8ba5584bde255f7833080

[11] "Coronavirus | Lockdown in Chhattisgarh's Raipur district from April 9 to 19," *The Hindu*, Raipur, Apr. 07, 2021. Accessed: Jun. 07, 2021. [Online]. Available: https://www.thehindu.com/news/national/other-states/coronavirus-lockdown-in-chhattisgarhs-raipur-district-from-april-9-to-19/article34265131.ece

[12] T. H. N. Desk, "Coronavirus second wave | List of States that have imposed restrictions, curfew and lockdowns," *The Hindu*, May 10, 2021. Accessed: Jun. 07, 2021. [Online]. Available: https://www.thehindu.com/news/national/coronavirus-second-wave-here-is-a-look-at-lockdowns-imposed-in-various-states/article34525655.ece

[13] Government of Karnataka, "Government Orders - COVID-19 INFORMATION PORTAL." https://covid19.karnataka.gov.in/new-page/Government%20Orders/en (accessed Jun. 07, 2021).

[14] "Lockdown in Delhi till April 26, health system may collapse with daily 25,000 Covid cases, says CM Kejriwal," *India Today*, Apr. 19, 2021. Accessed: Jun. 07, 2021. [Online]. Available: https://www.indiatoday.in/cities/delhi/story/lockdown-in-delhi-till-april-26-health-system-may-collapse-kejriwal-1792519-2021-04-19

[15] Rahul V Pisharody, "Telangana Covid-19 Night Curfew: Here's what is allowed and what isn't," *The Indian Express*, Apr. 20, 2021. Accessed: Jun. 07, 2021. [Online]. Available: https://indianexpress.com/article/india/telangana-covid-19-night-curfew-heres-what-is-allowed-and-what-isnt-7281397/

[16] "Jharkhand orders 7-day COVID-19 lockdown from 22 April; full list of what is allowed and what isn't," *Firstpost*, Apr. 20, 2021. Accessed: Jun. 07, 2021. [Online]. Available: https://www.firstpost.com/india/jharkhand-orders-7-day-covid-19-lockdown-from-22-april-full-list-of-what-is-allowed-and-what-is-not-9548451.html

[17] "Complete lockdown in Puducherry this weekend," *The Indian Express*, Apr. 21, 2021. Accessed: Jun. 07, 2021. [Online]. Available: https://indianexpress.com/article/india/complete-lockdown-in-puducherry-this-weekend-7282664/

[18] Victor Dasgupta, "Covid-19: Andhra Pradesh Govt To Impose Seven-Hour Night Curfew From April 24 | Details Here," *India.com*, Apr. 23, 2021. Accessed: Jun. 07, 2021. [Online]. Available: https://www.india.com/news/india/covid-19-andhra-pradesh-govt-to-impose-seven-hour-night-curfew-from-april-24-details-here-4609178/

[19] Shivendra Srivastava, "UP govt imposes weekend lockdown across state, night curfew in districts with over 500 active cases," *India Today*, Apr. 20, 2021. Accessed: Jun. 07, 2021. [Online]. Available: https://www.indiatoday.in/coronavirus-outbreak/story/up-govt-imposes-weekend-lockdown-across-state-night-curfew-in-districts-with-over-500-active-cases-1792981-2021-04-20

[20] "Covid-19 second wave: Here's a list of states that have imposed full lockdown," *The Indian Express*, May 09, 2021. Accessed: Jun. 07, 2021. [Online]. Available: https://indianexpress.com/article/india/covid-19-second-wave-heres-a-list-of-states-that-have-imposed-lockdowns-7306634/

[21] Anand Bodh, "Night curfew in Himachal Pradesh: Night curfew imposed in four districts of Himachal Pradesh," *The Times of India*, Apr. 25, 2021. Accessed: Jun. 07, 2021. [Online]. Available: https://timesofindia.indiatimes.com/city/shimla/covid-19-night-curfew-imposed-in-four-districts-of-himachal-pradesh/articleshow/82241827.cms

[22] "Karnataka 'close-down': What's allowed, what's not," *Deccan Herald*, Apr. 26, 2021. Accessed: Jun. 07, 2021. [Online]. Available: https://www.deccanherald.com/state/top-karnataka-stories/karnataka-close-down-whats-allowed-whats-not-979054.html

[23] "Nagaland to impose partial lockdown for a fortnight from April 30," *Outlook*, Apr. 27, 2021. Accessed: Jun. 07, 2021. [Online]. Available: https://www.outlookindia.com/newsscroll/nagaland-to-impose-partial-lockdown-for-a-fortnight-from-april-30/2072484

[24] "Nagaland Extends Total Lockdown Till June 11 as Covid-19 Cases Spike," *CNN-News 18*, May 28, 2021. Accessed: Jun. 07, 2021. [Online]. Available: https://www.news18.com/news/india/nagaland-extends-total-lockdown-till-june-11-as-covid-19-cases-spike-3787433.html

[25] "Everyone above the age of 18 to be eligible to get vaccine from May 1: Government," *The Economic Times*, Apr. 20, 2021. Accessed: Jun. 07, 2021. [Online]. Available: https://economictimes.indiatimes.com/news/india/everyone-above-the-age-of-18-to-be-eligible-to-get-vaccine-from-may-1-government/articleshow/82146974.cms?from=mdr

[26] "Mizoram: Eight-day lockdown imposed in Aizawl, other district headquarters," *mint*, May 02, 2021. Accessed: Jun. 07, 2021. [Online]. Available: https://www.livemint.com/news/india/mizoram-eight-day-lockdown-imposed-in-aizawl-other-district-headquarters-11619922863146.html

[27] "Mizoram extends lockdown in Aizawl till June 6," *The New Indian Express*, May 29, 2021. Accessed: Jun. 07, 2021. [Online]. Available: https://www.newindianexpress.com/nation/2021/may/29/mizoram-extends-lockdown-in-aizawl-till-june-6-2309255.html

[28] "Punjab imposes lockdown-like curbs till May 15; check out what's allowed, what's not," *Business Today*, May 03, 2021. Accessed: Jun. 07, 2021. [Online]. Available: https://www.businesstoday.in/coronavirus/punjab-imposes-lockdown-like-curbs-till-may-15-check-out-what-allowed-what-not/story/438098.html

[29] Debabrata Mohapatra, "Covid: Odisha announces 14-day lockdown from May 5," *The Times of India*, May 02, 2021. Accessed: Jun. 08, 2021. [Online]. Available: https://timesofindia.indiatimes.com/city/bhubaneswar/14-day-lockdown-in-odisha-from-may-5/articleshow/82352596.cms

[30] Debabrata Mohapatra, "Odisha lockdown news: Lockdown in Odisha extended till June 17," *The Times of India*, May 31, 2021. Accessed: Jun. 07, 2021. [Online]. Available: https://timesofindia.indiatimes.com/city/bhubaneswar/lockdown-extended-in-odisha-till-june-17/articleshow/83106749.cms

[31] "Andhra Pradesh Partial Curfew: Partial curfew in Andhra Pradesh for 14 days starting from May 5," *The Economic Times*, May 03, 2021. Accessed: Jun. 07, 2021. [Online]. Available: https://economictimes.indiatimes.com/news/india/partial-curfew-in-andhra-pradesh-for-14-days-starting-may-5/articleshow/82368569.cms

[32] "Night curfew in Gujarat till April 30 amid surge in COVID-19 cases," *Business Today*, Apr. 07, 2021. Accessed: Jun. 07, 2021. [Online]. Available: https://www.businesstoday.in/coronavirus/night-curfew-in-gujarat-till-april-30-amid-surge-in-covid-19-cases/story/435920.html

[33] "Telangana lockdown for 10 days starting tomorrow," *The Economic Times*, May 11, 2021. Accessed: Jun. 07, 2021. [Online]. Available: https://economictimes.indiatimes.com/news/india/covid-19-telangana-announces-10-day-lockdown-from-may-12/articleshow/82546448.cms?from=mdr

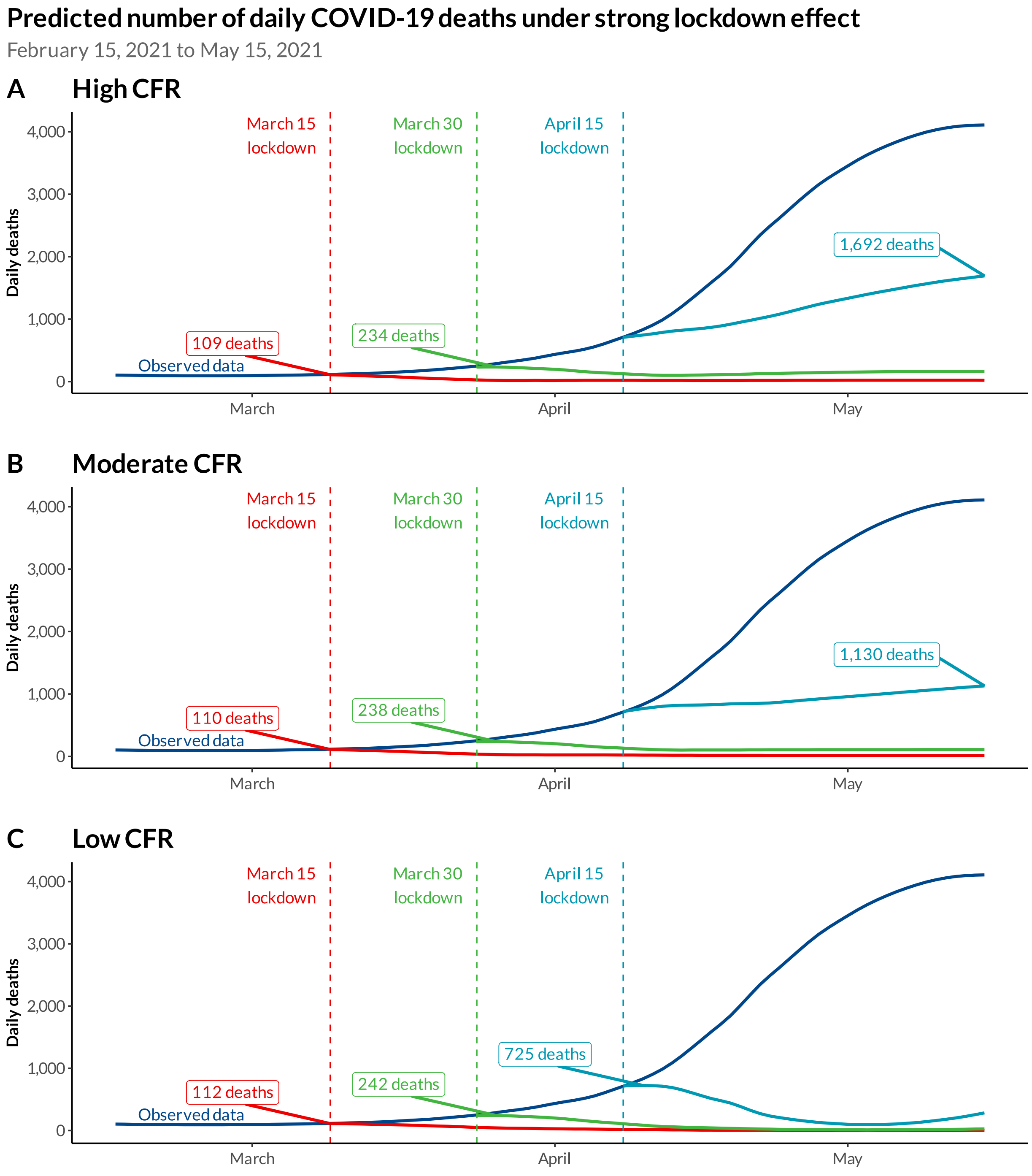


**Notes:** Observations and prediction period until May 15, 2021. Figures in boxes show peak number of deaths for each intervention.
**Abbrev:** CFR, case-fatality rate

**©COV-IND-19 Study Group**

**Figure S4**. Observed and predicted daily deaths under lockdown with moderate effect starting on different dates using high, moderate, and low CFR schedules from February 15 to May 15, 2021.

**Table S6. Predicted total deaths counts, deaths averted and % reduction with corresponding 95% credible intervals under lockdown with strong effect interventions (in thousands)**

| Evaluation date | Metrics | High CFR | | | | Moderate CFR | | | | Low CFR | | | |
|---|---|---|---|---|---|---|---|---|---|---|---|---|---|
| | | March 1 | March 15 | March 30 | April 15 | March 1 | March 15 | March 30 | `April 15 | March 1 | March 15 | March 30 | April 15 |
| 3/15/21 | *Observed* | 1.6 | - | - | - | 1.6 | - | - | - | 1.6 | - | - | - |
| | *Predicted* | 0.5 [0.0, 8.6] | | | | 0.6 [0.0, 10.2] | | | | 0.5 [0.0, 9.8] | | | |
| | *Averted* | 1.1 [-7.0, 1.6] | | | | 1.1 [-8.6, 1.6] | | | | 1.1 [-8.2, 1.6] | | | |
| | *% Reduction* | 71.0% [-437.3%, 100.0%] | | | | 65.6% [-536.6%, 100.0%] | | | | 68.0% [-509.1%, 100.0%] | | | |
| 3/30/21 | *Observed* | 5.2 | 3.6 | - | - | 5.2 | 3.6 | - | - | 5.2 | 3.6 | - | - |
| | *Predicted* | 0.6 [0.0, 10.4] | 0.4 [0.0, 6.1] | | | 0.8 [0.0, 13.2] | 0.6 [0.0, 8.8] | | | 0.9 [0.0, 15.3] | 0.9 [0.0, 12.6] | | |
| | *Averted* | 4.6 [-5.2, 5.2] | 3.2 [-2.5, 3.6] | | | 4.4 [-8.0, 5.2] | 3.0 [-5.2, 3.6] | | | 4.3 [-10.1, 5.2] | 2.8 [-8.9, 3.6] | | |
| | *% Reduction* | 87.7% [-100.1%, 100.0%] | 88.5% [-70.3%, 100.0%] | | | 84.6% [-153.4%, 100.0%] | 83.6% [-143.3%, 100.0%] | | | 83.2% [-193.8%, 100.0%] | 76.4% [-247.7%, 100.0%] | | |
| 4/15/21 | *Observed* | 17.1 | 15.4 | 11.8 | - | 17.1 | 15.4 | 11.8 | - | 17.1 | 15.4 | 11.8 | - |
| | *Predicted* | 0.8 [0.0, 13.8] | 0.8 [0.0, 11.1] | 2.0 [0.0, 11.6] | | 1.0 [0.0, 16.3] | 1.0 [0.0, 13.3] | 2.1 [0.0, 12.4] | | 1.0 [0.0, 17.4] | 1.2 [0.0, 15.2] | 1.8 [0.0, 10.8] | |
| | *Averted* | 16.3 [3.3, 17.1] | 14.7 [4.3, 15.4] | 9.9 [0.3, 11.8] | | 16.1 [0.7, 17.1] | 14.5 [2.1, 15.4] | 9.7 [-0.6, 11.8] | | 16.0 [-0.3, 17.1] | 14.3 [0.2, 15.4] | 10.0 [1.0, 11.8] | |
| | *% Reduction* | 95.4% [19.3%, 100.0%] | 95.1% [27.9%, 100.0%] | 83.3% [2.2%, 100.0%] | | 94.4% [4.2%, 100.0%] | 93.8% [13.9%, 100.0%] | 81.9% [-5.0%, 100.0%] | | 94.1% [-2.0%, 100.0%] | 92.5% [1.5%, 100.0%] | 84.4% [8.4%, 100.0%] | |
| 4/30/21 | *Observed* | 54.6 | 52.9 | 49.3 | 37.5 | 54.6 | 52.9 | 49.3 | 37.5 | 54.6 | 52.9 | 49.3 | 37.5 |
| | *Predicted* | 0.9 [0.0, 19.9] | 1.0 [0.0, 18.4] | 3.9 [0.0, 22.4] | 15.8 [0.0, 37.9] | 1.1 [0.0, 19.9] | 1.2 [0.0, 17.8] | 3.7 [0.0, 20.1] | 12.9 [0.0, 30.8] | 1.0 [0.0, 17.6] | 1.2 [0.0, 15.6] | 2.2 [0.0, 11.8] | 3.1 [0.0, 7.6] |
| | *Averted* | 53.6 [34.6, 54.6] | 51.9 [34.5, 52.9] | 45.4 [26.9, 49.3] | 21.7 [-0.4, 37.5] | 53.5 [34.7, 54.6] | 51.8 [35.2, 52.9] | 45.6 [29.2, 49.3] | 24.6 [6.7, 37.5] | 53.5 [36.9, 54.6] | 51.7 [37.4, 52.9] | 47.1 [37.5, 49.3] | 34.4 [29.9, 37.5] |
| | *% Reduction* | 98.3% [63.5%, 100.0%] | 98.0% [65.2%, 100.0%] | 92.1% [54.6%, 100.0%] | 57.9% [-1.1%, 100.0%] | 98.1% [63.5%, 100.0%] | 97.8% [66.5%, 100.0%] | 92.4% [59.3%, 100.0%] | 65.5% [17.9%, 100.0%] | 98.1% [67.7%, 100.0%] | 97.7% [70.6%, 100.0%] | 95.5% [76.0%, 100.0%] | 91.7% [79.9%, 100.0%] |
| 5/15/21 | *Observed* | 113.0 | 111.4 | 107.8 | 96.0 | 113.0 | 111.4 | 107.8 | 96.0 | 113.0 | 111.4 | 107.8 | 96.0 |
| | *Predicted* | 1.1 [0.0, 31.2] | 1.4 [0.0, 30.6] | 6.2 [0.0, 38.5] | 38.9 [0.0, 88.6] | 1.2 [0.0, 26.0] | 1.4 [0.0, 24.6] | 5.3 [0.0, 29.9] | 28.8 [0.0, 64.8] | 1.1 [0.0, 17.8] | 1.3 [0.0, 15.8] | 2.5 [0.0, 12.6] | 5.7 [0.0, 12.7] |
| | *Averted* | 111.9 [81.8, 113.0] | 110.0 [80.8, 111.4] | 101.6 [69.3, 107.8] | 57.1 [7.3, 96.0] | 111.8 [87.0, 113.0] | 110.0 [86.8, 111.4] | 102.5 [77.9, 107.8] | 67.2 [31.2, 96.0] | 112.0 [95.2, 113.0] | 110.2 [95.6, 111.4] | 105.3 [95.2, 107.8] | 90.3 [83.3, 96.0] |
| | *% Reduction* | 99.0% [72.4%, 100.0%] | 98.8% [72.5%, 100.0%] | 94.2% [64.3%, 100.0%] | 59.5% [7.7%, 100.0%] | 99.0% [77.0%, 100.0%] | 98.7% [77.9%, 100.0%] | 95.1% [72.3%, 100.0%] | 70.0% [32.5%, 100.0%] | 99.1% [84.2%, 100.0%] | 98.9% [85.8%, 100.0%] | 97.7% [88.3%, 100.0%] | 94.1% [86.8%, 100.0%] |

**Notes:** For lockdown schedules, each cell reports (1) the total number of observed deaths since the start of lockdown in the first row, (2) total number of predicted deaths since the start of lockdown in the second row (with 95% CI), (3) the number of deaths averted (relative to observed) since the start of lockdown in the third row (with 95% CI), and (4) the relative reduction in deaths (as a percent) under lockdown in the fourth row (with 95% CI). Cells that are highlighted in red represent a statistically significant reduction in the number of cases under lockdown at the 95% credible interval level. Cells that are bolded are emphasized in the main text. Numbers are reported in millions.

#### Section 3.1. Forecast results with waning immunity

This section shows the data for the projections that allow for waning immunity. The plots demonstrate virtually indistinguishable results from the projections without waning immunity.

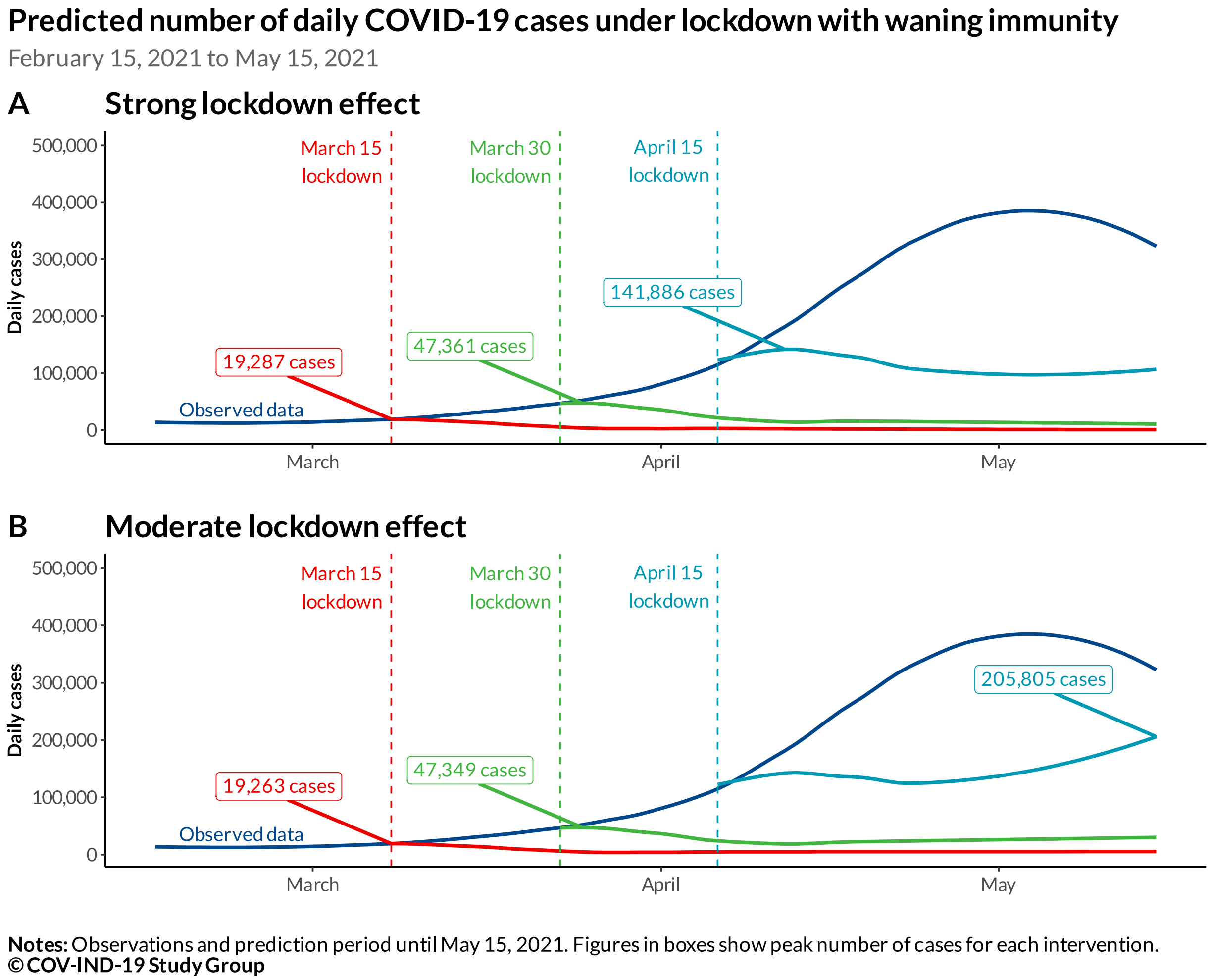


**Figure S5**. Observed and predicted daily cases under lockdown scenarios starting on different dates using the strong (**panel A**) and moderate (**panel B**) lockdown effect schedules with waning immunity from February 15 to May 15, 2021.

**Table S7.** Predicted total case counts and cases averted under different lockdown interventions under waning immunity (in millions)

| Evaluation date | Metrics | Strong lockdown effect | | | | Moderate lockdown effect | | | |
|---|---|---|---|---|---|---|---|---|---|
| | | March 1 | March 15 | March 30 | April 15 | March 1 | March 15 | March 30 | April 15 |
| 3/15/21 | *Observed* | 0.3 | - | - | - | 0.3 | - | - | - |
| | *Predicted* | 0.0 [0.0, 1.6] | | | | 0.0 [0.0, 1.6] | | | |
| | *Averted* | 0.2 [-1.3, 0.3] | | | | 0.2 [-1.3, 0.3] | | | |
| | *% Reduction* | 84.9% [-466.2%, 100.0%] | | | | 83.2% [-469.0%, 100.0%] | | | |
| 3/30/21 | *Observed* | 1.0 | 0.7 | - | - | 1.0 | 0.7 | - | - |
| | *Predicted* | 0.1 [0.0, 2.3] | 0.1 [0.0, 1.7] | | | 0.1 [0.0, 2.3] | 0.1 [0.0, 1.7] | | |
| | *Averted* | 1.0 [-1.3, 1.0] | 0.7 [-1.0, 0.7] | | | 0.9 [-1.3, 1.0] | 0.7 [-1.0, 0.7] | | |
| | *% Reduction* | 93.4% [-125.0%, 100.0%] | 90.9% [-132.0%, 100.0%] | | | 91.0% [-128.6%, 100.0%] | 89.4% [-132.8%, 100.0%] | | |
| 4/15/21 | *Observed* | 3.2 | 2.9 | 2.1 | - | 3.2 | 2.9 | 2.1 | - |
| | *Predicted* | 0.1 [0.0, 2.9] | 0.1 [0.0, 2.4] | 0.3 [0.0, 2.1] | | 0.1 [0.0, 3.0] | 0.2 [0.0, 2.5] | 0.3 [0.0, 2.2] | |
| | *Averted* | 3.1 [0.3, 3.2] | 2.8 [0.4, 2.9] | 1.8 [0.0, 2.1] | | 3.0 [0.2, 3.2] | 2.7 [0.4, 2.9] | 1.8 [-0.1, 2.1] | |
| | *% Reduction* | 97.5% [9.3%, 100.0%] | 96.0% [14.9%, 100.0%] | 86.0% [0.5%, 100.0%] | | 95.7% [5.7%, 100.0%] | 94.5% [12.8%, 100.0%] | 83.9% [-2.8%, 100.0%] | |
| 4/30/21 | *Observed* | 8.0 | 7.7 | 7.0 | 4.9 | 8.0 | 7.7 | 7.0 | 4.9 |
| | *Predicted* | 0.1 [0.0, 3.3] | 0.1 [0.0, 3.0] | 0.5 [0.0, 3.1] | 1.6 [0.0, 4.0] | 0.2 [0.0, 3.5] | 0.2 [0.0, 3.2] | 0.7 [0.0, 3.5] | 1.9 [0.0, 4.5] |
| | *Averted* | 8.0 [4.7, 8.0] | 7.6 [4.7, 7.7] | 6.5 [3.9, 7.0] | 3.3 [0.8, 4.9] | 7.9 [4.5, 8.0] | 7.5 [4.6, 7.7] | 6.3 [3.5, 7.0] | 3.0 [0.4, 4.9] |
| | *% Reduction* | 99.0% [58.7%, 100.0%] | 98.2% [61.0%, 100.0%] | 92.5% [55.3%, 100.0%] | 67.1% [17.0%, 100.0%] | 97.8% [56.4%, 100.0%] | 97.0% [59.3%, 100.0%] | 90.0% [50.6%, 100.0%] | 61.7% [7.4%, 100.0%] |
| 5/15/21 | *Observed* | 13.6 | 13.3 | 12.5 | 10.4 | 13.6 | 13.3 | 12.5 | 10.4 |
| | *Predicted* | 0.1 [0.0, 3.7] | 0.2 [0.0, 3.5] | 0.7 [0.0, 4.0] | 3.1 [0.0, 7.2] | 0.2 [0.0, 4.0] | 0.3 [0.0, 3.8] | 1.1 [0.0, 4.9] | 4.4 [0.2, 10.0] |
| | *Averted* | 13.5 [9.8, 13.6] | 13.1 [9.8, 13.3] | 11.8 [8.5, 12.5] | 7.3 [3.2, 10.4] | 13.3 [9.5, 13.6] | 13.0 [9.5, 13.3] | 11.4 [7.6, 12.5] | 6.0 [0.4, 10.2] |
| | *% Reduction* | 99.5% [72.6%, 100.0%] | 98.8% [73.8%, 100.0%] | 94.3% [68.2%, 100.0%] | 69.9% [30.4%, 100.0%] | 98.4% [70.4%, 100.0%] | 97.6% [71.7%, 100.0%] | 91.0% [61.0%, 100.0%] | 57.7% [4.0%, 97.9%] |

**Notes:** For lockdown schedules, each cell reports (1) the total number of observed cases since the start of lockdown in the first row, (2) total number of predicted cases since the start of lockdown in the second row (with 95% CI), (3) the number of cases averted (relative to observed) since the start of lockdown in the third row (with 95% CI), and (4) the relative reduction in cases (as a percent) under lockdown in the fourth row (with 95% CI). Cells that are highlighted in red represent a statistically significant reduction in the number of cases under lockdown at the 95% credible interval level. Numbers are reported in millions.

**Predicted number of daily COVID-19 deaths under strong lockdown effect with waning immunity**

February 15, 2021 to May 15, 2021

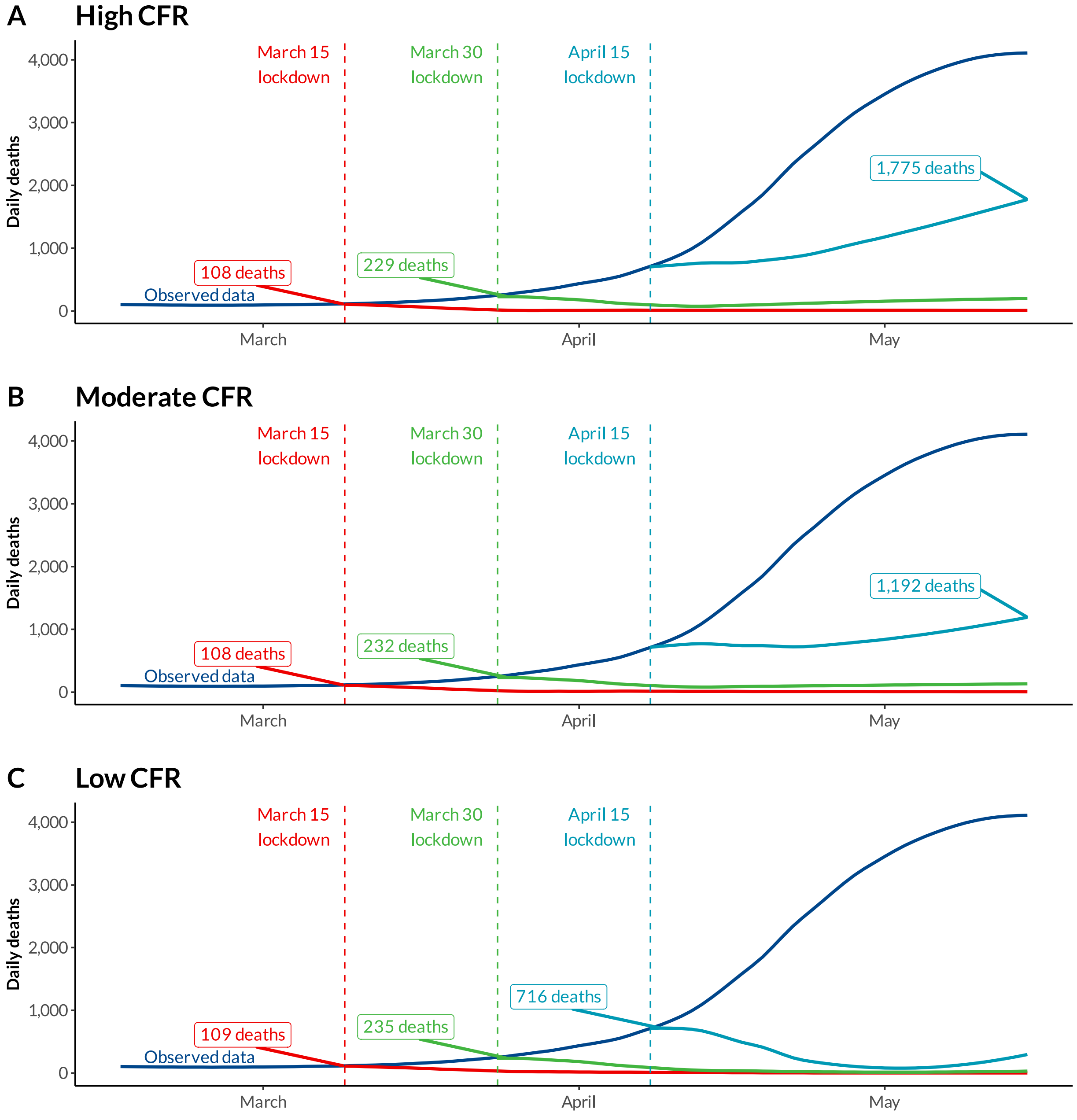


**Notes:** Observations and prediction period until May 15, 2021. Figures in boxes show peak number of deaths for each intervention.
**Abbrev:** CFR, case-fatality rate
**© COV-IND-19 Study Group**

**Figure S6**. Observed and predicted daily deaths under lockdown with strong effect and waning immunity starting on different dates using high, moderate, and low CFR schedules from February 15 to May 15, 2021.

**Table S8.** Predicted total deaths counts and deaths averted under lockdown with strong effect and waning immunity (in thousands)

| Evaluation date | Metrics | High CFR | | | | Moderate CFR | | | | Low CFR | | | |
|---|---|---|---|---|---|---|---|---|---|---|---|---|---|
| | | March 1 | March 15 | March 30 | April 15 | March 1 | March 15 | March 30 | April 15 | March 1 | March 15 | March 30 | April 15 |
| 3/15/21 | *Observed* | 1.6 | - | - | - | 1.6 | - | - | - | 1.6 | - | - | - |
| | *Predicted* | 0.2 [0.0, 8.5] | | | | 0.3 [0.0, 10.0] | | | | 0.3 [0.0, 9.6] | | | |
| | *Averted* | 1.4 [-6.9, 1.6] | | | | 1.3 [-8.4, 1.6] | | | | 1.4 [-8.0, 1.6] | | | |
| | *% Reduction* | 86.1% [-426.9%, 100.0%] | | | | 83.4% [-525.2%, 100.0%] | | | | 84.4% [-497.6%, 100.0%] | | | |
| 3/30/21 | *Observed* | 5.2 | 3.6 | - | - | 5.2 | 3.6 | - | - | 5.2 | 3.6 | - | - |
| | *Predicted* | 0.3 [0.0, 10.2] | 0.2 [0.0, 6.0] | | | 0.4 [0.0, 12.9] | 0.3 [0.0, 8.6] | | | 0.4 [0.0, 15.1] | 0.5 [0.0, 12.3] | | |
| | *Averted* | 4.9 [-5.0, 5.2] | 3.4 [-2.4, 3.6] | | | 4.8 [-7.7, 5.2] | 3.3 [-5.0, 3.6] | | | 4.8 [-9.9, 5.2] | 3.1 [-8.7, 3.6] | | |
| | *% Reduction* | 94.1% [-96.2%, 100.0%] | 93.5% [-67.4%, 100.0%] | | | 92.5% [-148.0%, 100.0%] | 90.7% [-139.2%, 100.0%] | | | 91.7% [-190.1%, 100.0%] | 86.6% [-241.6%, 100.0%] | | |
| 4/15/21 | *Observed* | 17.1 | 15.4 | 11.8 | - | 17.1 | 15.4 | 11.8 | - | 17.1 | 15.4 | 11.8 | - |
| | *Predicted* | 0.4 [0.0, 13.6] | 0.5 [0.0, 10.9] | 1.5 [0.0, 10.9] | | 0.4 [0.0, 16.1] | 0.6 [0.0, 13.0] | 1.7 [0.0, 11.7] | | 0.5 [0.0, 17.2] | 0.7 [0.0, 14.9] | 1.4 [0.0, 10.2] | |
| | *Averted* | 16.7 [3.5, 17.1] | 15.0 [4.5, 15.4] | 10.3 [0.9, 11.8] | | 16.6 [1.0, 17.1] | 14.8 [2.5, 15.4] | 10.2 [0.1, 11.8] | | 16.6 [-0.1, 17.1] | 14.7 [0.6, 15.4] | 10.4 [1.6, 11.8] | |
| | *% Reduction* | 97.9% [20.4%, 100.0%] | 96.9% [29.2%, 100.0%] | 87.1% [7.9%, 100.0%] | | 97.4% [5.7%, 100.0%] | 96.1% [15.9%, 100.0%] | 86.0% [0.8%, 100.0%] | | 97.2% [-0.7%, 100.0%] | 95.5% [3.8%, 100.0%] | 88.2% [13.6%, 100.0%] | |
| 4/30/21 | *Observed* | 54.6 | 52.9 | 49.3 | 37.5 | 54.6 | 52.9 | 49.3 | 37.5 | 54.6 | 52.9 | 49.3 | 37.5 |
| | *Predicted* | 0.4 [0.0, 19.8] | 0.7 [0.0, 18.5] | 3.4 [0.0, 21.8] | 13.2 [0.0, 33.9] | 0.4 [0.0, 19.8] | 0.8 [0.0, 17.7] | 3.2 [0.0, 19.3] | 10.8 [0.0, 27.3] | 0.5 [0.0, 17.4] | 0.7 [0.0, 15.2] | 1.8 [0.0, 11.2] | 2.6 [0.0, 6.6] |
| | *Averted* | 54.2 [34.7, 54.6] | 52.2 [34.4, 52.9] | 45.9 [27.5, 49.3] | 24.3 [3.6, 37.5] | 54.1 [34.8, 54.6] | 52.2 [35.2, 52.9] | 46.2 [30.0, 49.3] | 26.7 [10.2, 37.5] | 54.1 [37.2, 54.6] | 52.2 [37.7, 52.9] | 47.6 [38.1, 49.3] | 34.9 [30.9, 37.5] |
| | *% Reduction* | 99.4% [63.7%, 100.0%] | 98.7% [65.1%, 100.0%] | 93.1% [55.8%, 100.0%] | 64.7% [9.6%, 100.0%] | 99.2% [63.8%, 100.0%] | 98.5% [66.6%, 100.0%] | 93.5% [60.8%, 100.0%] | 71.2% [27.1%, 100.0%] | 99.1% [68.1%, 100.0%] | 98.6% [71.3%, 100.0%] | 96.4% [77.3%, 100.0%] | 93.2% [82.3%, 100.0%] |
| 5/15/21 | *Observed* | 113.0 | 111.4 | 107.8 | 96.0 | 113.0 | 111.4 | 107.8 | 96.0 | 113.0 | 111.4 | 107.8 | 96.0 |
| | *Predicted* | 0.3 [0.0, 31.2] | 0.9 [0.0, 30.8] | 6.1 [0.0, 39.0] | 35.6 [0.0, 84.7] | 0.4 [0.0, 25.8] | 0.9 [0.0, 24.7] | 5.0 [0.0, 30.0] | 26.1 [0.0, 61.4] | 0.5 [0.0, 17.6] | 0.8 [0.0, 15.5] | 2.1 [0.0, 12.1] | 5.0 [0.0, 11.8] |
| | *Averted* | 112.8 [81.9, 113.0] | 110.5 [80.6, 111.4] | 101.7 [68.8, 107.8] | 60.4 [11.3, 96.0] | 112.6 [87.2, 113.0] | 110.5 [86.8, 111.4] | 102.8 [77.8, 107.8] | 69.9 [34.6, 96.0] | 112.6 [95.4, 113.0] | 110.7 [95.9, 111.4] | 105.8 [95.7, 107.8] | 90.9 [84.2, 96.0] |
| | *% Reduction* | 99.8% [72.4%, 100.0%] | 99.2% [72.3%, 100.0%] | 94.4% [63.9%, 100.0%] | 62.9% [11.8%, 100.0%] | 99.7% [77.2%, 100.0%] | 99.2% [77.9%, 100.0%] | 95.3% [72.2%, 100.0%] | 72.8% [36.1%, 100.0%] | 99.6% [84.4%, 100.0%] | 99.3% [86.1%, 100.0%] | 98.1% [88.8%, 100.0%] | 94.8% [87.7%, 100.0%] |

**Notes:** For lockdown schedules, each cell reports (1) the total number of observed deaths since the start of lockdown in the first row, (2) total number of predicted deaths since the start of lockdown in the second row (with 95% CI), (3) the number of deaths averted (relative to observed) since the start of lockdown in the third row (with 95% CI), and (4) the relative reduction in deaths (as a percent) under lockdown in the fourth row (with 95% CI). Cells that are highlighted in red represent a statistically significant reduction in the number of cases under lockdown at the 95% credible interval level. Numbers are reported in millions.

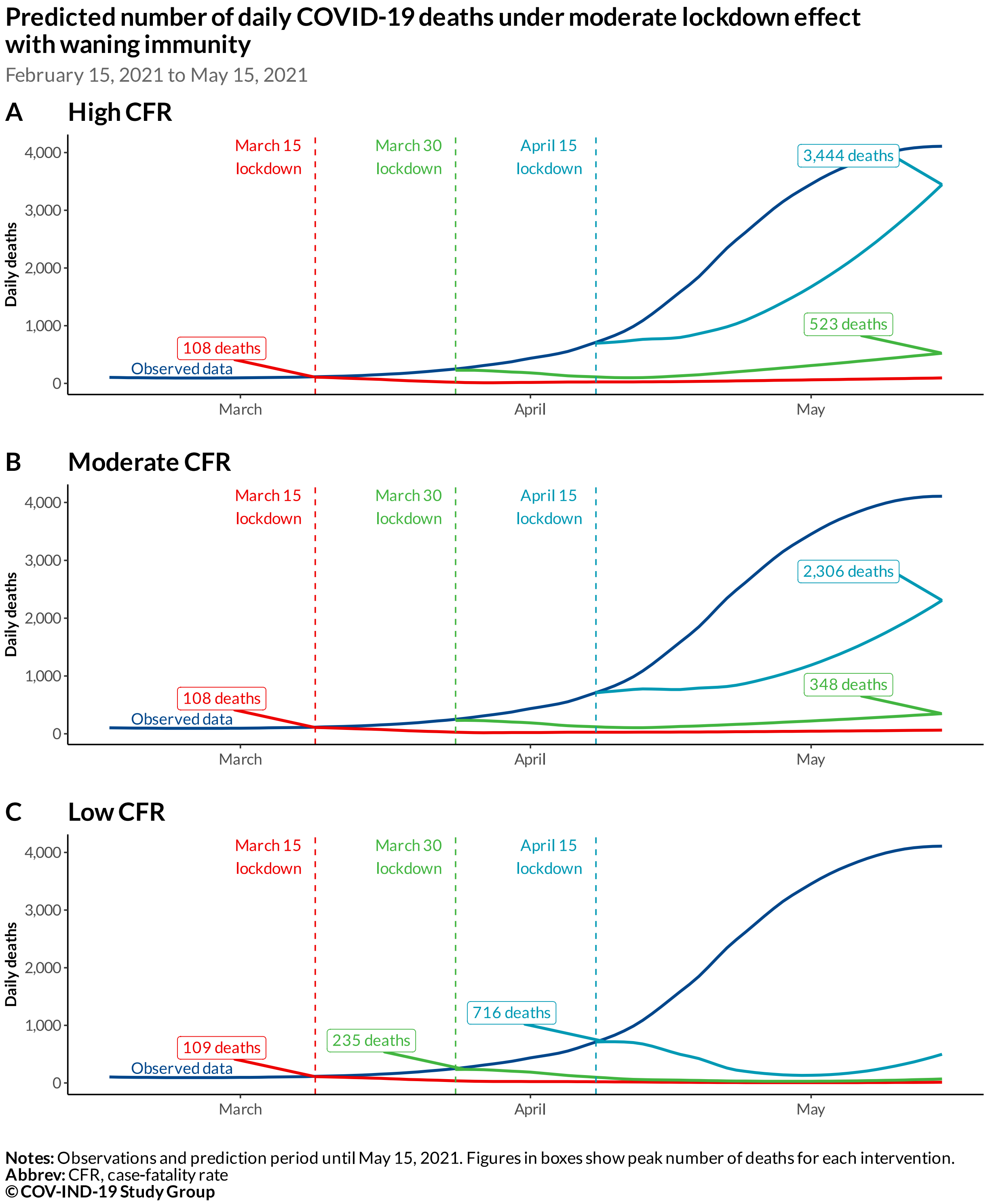


**Figure S7**. Observed and predicted daily deaths under lockdown with moderate effect and waning immunity starting on different dates using high, moderate, and low CFR schedules from February 15 to May 15, 2021.

**Table S9.** Predicted total death counts and deaths averted under lockdown with moderate effect and waning immunity (in thousands)

| Evaluation date | Metrics | High CFR | | | | Moderate CFR | | | | Low CFR | | | |
|---|---|---|---|---|---|---|---|---|---|---|---|---|---|
| | | March 1 | March 15 | March 30 | April 15 | March 1 | March 15 | March 30 | April 15 | March 1 | March 15 | March 30 | April 15 |
| 3/15/21 | *Observed* | 1.6 | - | - | - | 1.6 | - | - | - | 1.6 | - | - | - |
| | *Predicted* | 0.2 [0.0, 8.5] | | | | 0.3 [0.0, 10.1] | | | | 0.3 [0.0, 9.6] | | | |
| | *Averted* | 1.4 [-6.9, 1.6] | | | | 1.3 [-8.5, 1.6] | | | | 1.3 [-8.0, 1.6] | | | |
| | *% Reduction* | 84.7% [-430.1%, 100.0%] | | | | 81.6% [-527.9%, 100.0%] | | | | 82.3% [-499.0%, 100.0%] | | | |
| 3/30/21 | *Observed* | 5.2 | 3.6 | - | - | 5.2 | 3.6 | - | - | 5.2 | 3.6 | - | - |
| | *Predicted* | 0.4 [0.0, 10.4] | 0.3 [0.0, 6.1] | | | 0.5 [0.0, 13.1] | 0.4 [0.0, 8.7] | | | 0.6 [0.0, 15.3] | 0.6 [0.0, 12.4] | | |
| | *Averted* | 4.8 [-5.2, 5.2] | 3.3 [-2.5, 3.6] | | | 4.7 [-7.9, 5.2] | 3.2 [-5.1, 3.6] | | | 4.6 [-10.1, 5.2] | 3.0 [-8.8, 3.6] | | |
| | *% Reduction* | 92.4% [-99.0%, 100.0%] | 92.4% [-68.1%, 100.0%] | | | 90.2% [-152.1%, 100.0%] | 89.1% [-140.5%, 100.0%] | | | 88.6% [-194.2%, 100.0%] | 84.4% [-242.8%, 100.0%] | | |
| 4/15/21 | *Observed* | 17.1 | 15.4 | 11.8 | - | 17.1 | 15.4 | 11.8 | - | 17.1 | 15.4 | 11.8 | - |
| | *Predicted* | 0.6 [0.0, 14.1] | 0.7 [0.0, 11.2] | 1.8 [0.0, 11.3] | | 0.8 [0.0, 16.7] | 0.8 [0.0, 13.3] | 1.9 [0.0, 12.1] | | 0.8 [0.0, 17.7] | 0.9 [0.0, 15.2] | 1.6 [0.0, 10.5] | |
| | *Averted* | 16.4 [2.9, 17.1] | 14.8 [4.2, 15.4] | 10.1 [0.5, 11.8] | | 16.3 [0.3, 17.1] | 14.6 [2.1, 15.4] | 9.9 [-0.3, 11.8] | | 16.2 [-0.7, 17.1] | 14.5 [0.3, 15.4] | 10.3 [1.3, 11.8] | |
| | *% Reduction* | 96.3% [17.3%, 100.0%] | 95.5% [27.3%, 100.0%] | 85.0% [4.5%, 100.0%] | | 95.5% [2.0%, 100.0%] | 94.6% [13.6%, 100.0%] | 83.9% [-2.5%, 100.0%] | | 95.3% [-4.0%, 100.0%] | 94.0% [1.9%, 100.0%] | 86.6% [11.3%, 100.0%] | |
| 4/30/21 | *Observed* | 54.6 | 52.9 | 49.3 | 37.5 | 54.6 | 52.9 | 49.3 | 37.5 | 54.6 | 52.9 | 49.3 | 37.5 |
| | *Predicted* | 1.0 [0.0, 21.0] | 1.4 [0.0, 19.5] | 4.8 [0.0, 24.4] | 15.9 [0.0, 38.7] | 1.0 [0.0, 20.9] | 1.4 [0.0, 18.5] | 4.4 [0.0, 21.6] | 12.8 [0.0, 30.9] | 0.9 [0.0, 18.1] | 1.0 [0.0, 15.6] | 2.1 [0.0, 11.8] | 2.9 [0.0, 7.2] |
| | *Averted* | 53.6 [33.6, 54.6] | 51.6 [33.5, 52.9] | 44.5 [24.9, 49.3] | 21.6 [-1.2, 37.5] | 53.5 [33.7, 54.6] | 51.6 [34.4, 52.9] | 45.0 [27.8, 49.3] | 24.7 [6.6, 37.5] | 53.7 [36.4, 54.6] | 51.9 [37.4, 52.9] | 47.2 [37.5, 49.3] | 34.6 [30.3, 37.5] |
| | *% Reduction* | 98.2% [61.5%, 100.0%] | 97.4% [63.3%, 100.0%] | 90.2% [50.5%, 100.0%] | 57.6% [-3.1%, 100.0%] | 98.1% [61.7%, 100.0%] | 97.4% [65.0%, 100.0%] | 91.1% [56.3%, 100.0%] | 65.9% [17.6%, 100.0%] | 98.4% [66.8%, 100.0%] | 98.0% [70.5%, 100.0%] | 95.7% [76.1%, 100.0%] | 92.3% [80.8%, 100.0%] |
| 5/15/21 | *Observed* | 113.0 | 111.4 | 107.8 | 96.0 | 113.0 | 111.4 | 107.8 | 96.0 | 113.0 | 111.4 | 107.8 | 96.0 |
| | *Predicted* | 1.6 [0.0, 33.7] | 2.5 [0.0, 33.8] | 11.1 [0.0, 50.2] | 53.4 [2.4, 123.4] | 1.5 [0.0, 28.1] | 2.2 [0.0, 26.9] | 8.6 [0.0, 38.2] | 38.4 [2.0, 88.3] | 0.9 [0.0, 18.5] | 1.2 [0.0, 16.1] | 2.8 [0.0, 13.6] | 7.1 [0.2, 16.3] |
| | *Averted* | 111.5 [79.3, 113.0] | 108.9 [77.7, 111.4] | 96.7 [57.7, 107.8] | 42.6 [-27.4, 93.6] | 111.6 [85.0, 113.0] | 109.3 [84.6, 111.4] | 99.2 [69.7, 107.8] | 57.6 [7.7, 94.0] | 112.1 [94.5, 113.0] | 110.3 [95.3, 111.4] | 105.0 [94.2, 107.8] | 88.9 [79.7, 95.8] |
| | *% Reduction* | 98.6% [70.2%, 100.0%] | 97.7% [69.7%, 100.0%] | 89.7% [53.5%, 100.0%] | 44.4% [-28.6%, 97.5%] | 98.7% [75.2%, 100.0%] | 98.1% [75.9%, 100.0%] | 92.0% [64.6%, 100.0%] | 60.0% [8.0%, 98.0%] | 99.2% [83.6%, 100.0%] | 99.0% [85.5%, 100.0%] | 97.4% [87.4%, 100.0%] | 92.6% [83.1%, 99.8%] |

**Notes:** For lockdown schedules, each cell reports (1) the total number of observed deaths since the start of lockdown in the first row, (2) total number of predicted deaths since the start of lockdown in the second row (with 95% CI), (3) the number of deaths averted (relative to observed) since the start of lockdown in the third row (with 95% CI), and (4) the relative reduction in deaths (as a percent) under lockdown in the fourth row (with 95% CI). Cells that are highlighted in red represent a statistically significant reduction in the number of cases under lockdown at the 95% credible interval level. Numbers are reported in millions.

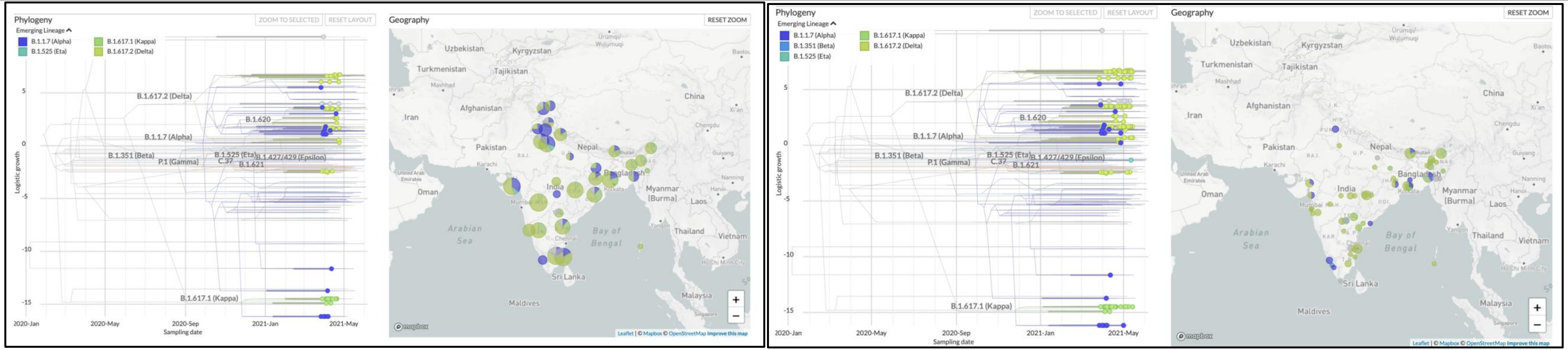


Figure S8a: A temporal depiction of the dynamic variant landscape in India. The left panel shows data from March 15-April 25, 2021 whereas the right panel covers sampling dates from March 1-May 15 of 2021. Maps extracted from https://nextstrain.org/ncov/asia.

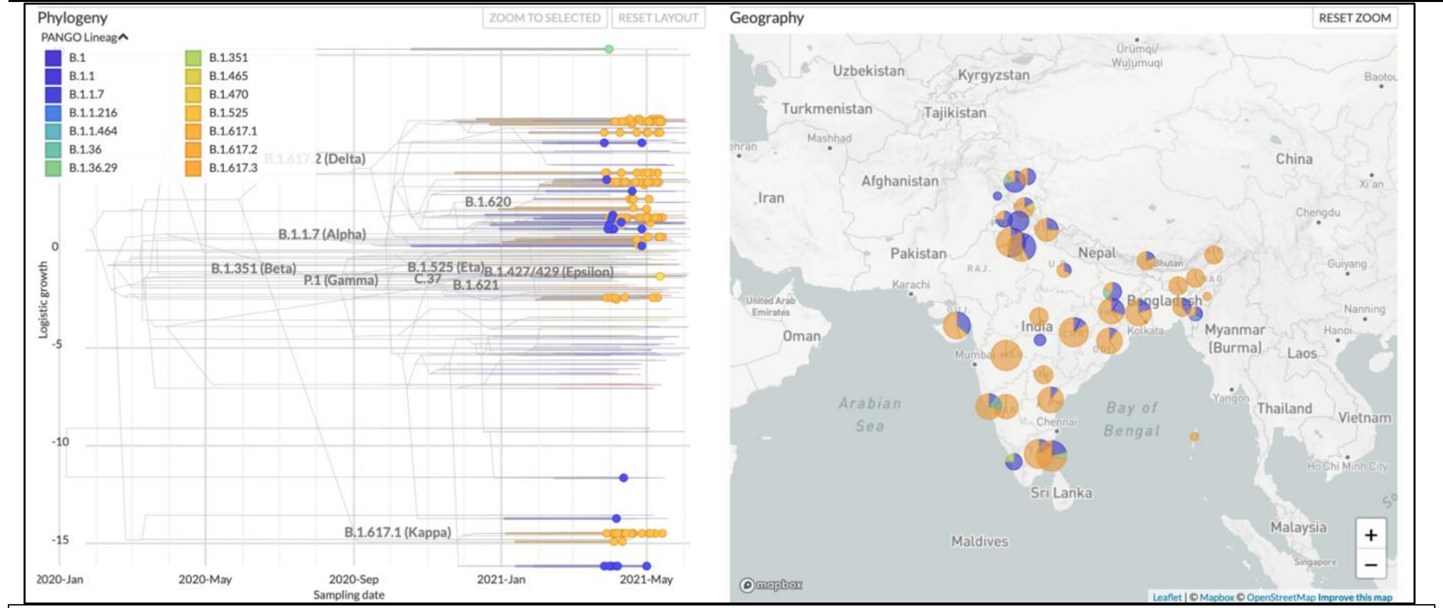


**Figure S8b:** Latest depiction of the variant landscape in India with a sampling period of March1-May 15 where variants are classified according to PANGO nomenclature (https://cov-lineages.org). Maps extracted from https://nextstrain.org/ncov/asia.

### REFERENCES

1. Coronavirus in India: Latest Map and Case Count, (available at https://www.covid19india.org).

2. H. Ritchie, E. Ortiz-Ospina, D. Beltekian, E. Mathieu, J. Hasell, B. Macdonald, C. Giattino, C. Appel, L. Rodés-Guirao, M. Roser, Coronavirus Pandemic (COVID-19). *Our World in Data* (2020) (available at https://ourworldindata.org/coronavirus/country/india).

3. S. Elbe, G. Buckland-Merrett, Data, disease and diplomacy: GISAID's innovative contribution to global health: Data, Disease and Diplomacy. *Global Challenges*. **1**, 33–46 (2017).

4. Y. Shu, J. McCauley, GISAID: Global initiative on sharing all influenza data – from vision to reality. *Eurosurveillance*. **22** (2017), doi:10.2807/1560-7917.ES.2017.22.13.30494.

5. S. Purkayastha, R. Kundu, R. Bhaduri, D. Barker, M. Kleinsasser, D. Ray, B. Mukherjee, Estimating the wave 1 and wave 2 infection fatality rates from SARS-CoV-2 in India. *medRxiv* (2021), doi:10.1101/2021.05.25.21257823.

6. N. R. Faria, T. A. Mellan, C. Whittaker, I. M. Claro, D. da S. Candido, S. Mishra, M. A. E. Crispim, F. C. S. Sales, I. Hawryluk, J. T. McCrone, R. J. G. Hulswit, L. A. M. Franco, M. S. Ramundo, J. G. de Jesus, P. S. Andrade, T. M. Coletti, G. M. Ferreira, C. A. M. Silva, E. R. Manuli, R. H. M. Pereira, P. S. Peixoto, M. U. G. Kraemer, N. Gaburo, C. da C. Camilo, H. Hoeltgebaum, W. M. Souza, E. C. Rocha, L. M. de Souza, M. C. de Pinho, L. J. T. Araujo, F. S. V. Malta, A. B. de Lima, J. do P. Silva, D. A. G. Zauli, A. C. de S. Ferreira, R. P. Schnekenberg, D. J. Laydon, P. G. T. Walker, H. M. Schlüter, A. L. P. dos Santos, M. S. Vidal, V. S. D. Caro, R. M. F. Filho, H. M. dos Santos, R. S. Aguiar, J. L. Proença-Modena, B. Nelson, J. A. Hay, M. Monod, X. Miscouridou, H. Coupland, R. Sonabend, M. Vollmer, A. Gandy, C. A. Prete, V. H. Nascimento, M. A. Suchard, T. A. Bowden, S. L. K. Pond, C.-H. Wu, O. Ratmann, N. M. Ferguson, C. Dye, N. J. Loman, P. Lemey, A. Rambaut, N. A. Fraiji, M. do P. S. S. Carvalho, O. G. Pybus, S. Flaxman, S. Bhatt, E. C. Sabino, Genomics and epidemiology of the P.1 SARS-CoV-2 lineage in Manaus, Brazil. *Science*. **372**, 815–821 (2021).

7. S. Flaxman, S. Mishra, A. Gandy, H. J. T. Unwin, T. A. Mellan, H. Coupland, C. Whittaker, H. Zhu, T. Berah, J. W. Eaton, M. Monod, Imperial College COVID-19 Response Team, P. N. Perez-Guzman, N. Schmit, L. Cilloni, K. E. C. Ainslie, M. Baguelin, A. Boonyasiri, O. Boyd, L. Cattarino, L. V. Cooper, Z. Cucunubá, G. Cuomo-Dannenburg, A. Dighe, B. Djaafara, I. Dorigatti, S. L. van Elsland, R. G. FitzJohn, K. A. M. Gaythorpe, L. Geidelberg, N. C. Grassly, W. D. Green, T. Hallett, A. Hamlet, W. Hinsley, B. Jeffrey, E. Knock, D. J. Laydon, G. Nedjati-Gilani, P. Nouvellet, K. V. Parag, I. Siveroni, H. A. Thompson, R. Verity, E. Volz, C. E. Walters, H. Wang, Y. Wang, O. J. Watson, P. Winskill, X. Xi, P. G. T. Walker, A. C. Ghani, C. A. Donnelly, S. Riley, M. A. C. Vollmer, N. M. Ferguson, L. C. Okell, S. Bhatt, Estimating the effects of non-pharmaceutical interventions on COVID-19 in Europe. *Nature*. **584**, 257–261 (2020).

8. H. J. T. Unwin, S. Mishra, V. C. Bradley, A. Gandy, T. A. Mellan, H. Coupland, J. Ish-Horowicz, M. A. C. Vollmer, C. Whittaker, S. L. Filippi, X. Xi, M. Monod, O. Ratmann, M. Hutchinson, F. Valka, H. Zhu, I. Hawryluk, P. Milton, K. E. C. Ainslie, M. Baguelin, A. Boonyasiri, N. F. Brazeau, L. Cattarino, Z. Cucunuba, G. Cuomo-Dannenburg, I. Dorigatti, O. D. Eales, J. W. Eaton, S. L. van Elsland, R. G. FitzJohn, K. A. M. Gaythorpe, W. Green, W. Hinsley, B. Jeffrey, E. Knock, D. J. Laydon, J. Lees, G. Nedjati-Gilani, P. Nouvellet, L. Okell, K. V. Parag, I. Siveroni, H. A. Thompson, P. Walker, C. E. Walters, O. J. Watson, L. K. Whittles, A. C. Ghani, N. M. Ferguson, S. Riley, C. A. Donnelly, S. Bhatt, S. Flaxman, State-level tracking of COVID-19 in the United States. *Nat Commun*. **11**, 6189 (2020).

9. W. Feller, On the Integral Equation of Renewal Theory. *Ann. Math. Statist.* **12**, 243–267 (1941).

10. R. Bellman, T. Harris, On Age-Dependent Binary Branching Processes. *The Annals of Mathematics*. **55**, 280 (1952).

11. C. H. Hansen, D. Michlmayr, S. M. Gubbels, K. Mølbak, S. Ethelberg, Assessment of protection against reinfection with SARS-CoV-2 among 4 million PCR-tested individuals in Denmark in 2020: a population-level observational study. *The Lancet*. **397**, 1204–1212 (2021).

12. V. Hall, S. Foulkes, A. Charlett, A. Atti, E. Monk, R. Simmons, E. Wellington, M. Cole, A. Saei, B. Oguti, K. Munro, S. Wallace, P. Kirwan, M. Shrotri, A. Vusirikala, S. Rokadiya, M. Kall, M. Zambon, M. Ramsay, T. Brooks, SIREN Study Group, C. Brown, M. Chand, S. Hopkins, "Do antibody positive healthcare workers have lower SARS-CoV-2 infection rates than antibody negative healthcare workers? Large multi-centre prospective cohort study (the SIREN study), England: June to November 2020" (preprint, Epidemiology, 2021), , doi:10.1101/2021.01.13.21249642.

13. The SAFER Investigators and Field Study Team, The Crick COVID-19 Consortium, CMMID COVID-19 working group, J. Hellewell, T. W. Russell, R. Beale, G. Kelly, C. Houlihan, E. Nastouli, A. J. Kucharski, Estimating the effectiveness of routine asymptomatic PCR testing at different frequencies for the detection of SARS-CoV-2 infections. *BMC Med*. **19**, 106 (2021).

14. L. Wang, Y. Zhou, J. He, B. Zhu, F. Wang, L. Tang, M. Kleinsasser, D. Barker, M. C. Eisenberg, P. X. K. Song, An epidemiological forecast model and software assessing interventions on the COVID-19 epidemic in China. *Journal of Data Science*. **18**, 409–432 (2021).

15. L. Wang, *lilywang1988/eSIR* (2021; https://github.com/lilywang1988/eSIR).

16. D. Ray, M. Salvatore, R. Bhattacharyya, L. Wang, J. Du, S. Mohammed, S. Purkayastha, A. Halder, A. Rix, D. Barker, M. Kleinsasser, Y. Zhou, D. Bose, P. Song, M. Banerjee, V. Baladandayuthapani, P. Ghosh, B. Mukherjee, Predictions, Role of Interventions and Effects of a Historic National Lockdown in India's Response to the the COVID-19 Pandemic: Data Science Call to Arms. *Harvard Data Science Review* (2020), doi:10.1162/99608f92.60e08ed5.

17. S. Das, Prediction of COVID-19 Disease Progression in India : Under the Effect of National Lockdown. *arXiv:2004.03147 [cs, q-bio]* (2020) (available at http://arxiv.org/abs/2004.03147).

18. S. Deb, M. Majumdar, A time series method to analyze incidence pattern and estimate reproduction number of COVID-19. *arXiv:2003.10655 [q-bio, stat]* (2020) (available at http://arxiv.org/abs/2003.10655).

19. R. Ranjan, "Predictions for COVID-19 outbreak in India using Epidemiological models" (preprint, Epidemiology, 2020), , doi:10.1101/2020.04.02.20051466.

20. T. Sardar, S. S. Nadim, S. Rana, J. Chattopadhyay, Assessment of lockdown effect in some states and overall India: A predictive mathematical study on COVID-19 outbreak. *Chaos, Solitons & Fractals*. **139**, 110078 (2020).

21. R. Singh, R. Adhikari, Age-structured impact of social distancing on the COVID-19 epidemic in India. *arXiv:2003.12055 [cond-mat, q-bio]* (2020) (available at http://arxiv.org/abs/2003.12055).

22. T. Mkhatshwa, A. Mummert, Modeling Super-spreading Events for Infectious Diseases: Case Study SARS. *arXiv:1007.0908 [q-bio]* (2010) (available at http://arxiv.org/abs/1007.0908).

23. N. Chen, M. Zhou, X. Dong, J. Qu, F. Gong, Y. Han, Y. Qiu, J. Wang, Y. Liu, Y. Wei, J. Xia, T. Yu, X. Zhang, L. Zhang, Epidemiological and clinical characteristics of 99 cases of 2019 novel coronavirus pneumonia in Wuhan, China: a descriptive study. *The Lancet*. **395**, 507–513 (2020).

24. Q. Li, X. Guan, P. Wu, X. Wang, L. Zhou, Y. Tong, R. Ren, K. S. M. Leung, E. H. Y. Lau, J. Y. Wong, X. Xing, N. Xiang, Y. Wu, C. Li, Q. Chen, D. Li, T. Liu, J. Zhao, M. Liu, W. Tu, C. Chen, L. Jin, R. Yang, Q. Wang, S. Zhou, R. Wang, H. Liu, Y. Luo, Y. Liu, G. Shao, H. Li, Z. Tao, Y. Yang, Z. Deng, B. Liu, Z. Ma, Y. Zhang, G. Shi, T. T. Y. Lam, J. T. Wu, G. F. Gao, B. J. Cowling, B. Yang, G. M. Leung, Z. Feng, Early Transmission Dynamics in Wuhan, China, of Novel Coronavirus–Infected Pneumonia. *N Engl J Med*. **382**, 1199–1207 (2020).

25. S. Ryu, B. C. Chun, An interim review of the epidemiological characteristics of 2019 novel coronavirus. *Epidemiol Health*. **42**, e2020006 (2020).

26. *mrc-ide/EpiEstim* (MRC Centre for Global Infectious Disease Analysis, 2021; https://github.com/mrc-ide/EpiEstim).

27. A. Cori, N. M. Ferguson, C. Fraser, S. Cauchemez, A New Framework and Software to Estimate Time-Varying Reproduction Numbers During Epidemics. *American Journal of Epidemiology*. **178**, 1505–1512 (2013).

We gratefully acknowledge the following Authors from the Originating laboratories responsible for obtaining the specimens, as well as the Submitting laboratories where the genome data were generated and shared via GISAID, on which this research is based.

All Submitters of data may be contacted directly via www.gisaid.org

Authors are sorted alphabetically.

| Accession ID | Originating Laboratory | Submitting Laboratory | Authors |
|---|---|---|---|
| EPI_ISL_1034156, EPI_ISL_1034168, EPI_ISL_1034169, EPI_ISL_1034170, EPI_ISL_1034175, EPI_ISL_1034176, EPI_ISL_1034179, EPI_ISL_1034183, EPI_ISL_1034184, EPI_ISL_1034185 | NIV Influenza | NIV Influenza | Potdar V |
| EPI_ISL_1039249, EPI_ISL_1055383 | Virus Research and Diagnostic Laboratory, Medical College Level, Kasturba Hospital for Infectious Diseases | Indian Council of Medical Research-National Institute of Virology, Microbial Containment Complex | Pragya D. Yadav, Jayanti Shastri |
| EPI_ISL_1055384, EPI_ISL_1055768, EPI_ISL_1055770, EPI_ISL_1055771, EPI_ISL_1055785, EPI_ISL_1055788, EPI_ISL_1055789, EPI_ISL_1063400, EPI_ISL_1063401, EPI_ISL_1063494, EPI_ISL_1063526, EPI_ISL_1063527, EPI_ISL_1063529, EPI_ISL_1063531, EPI_ISL_1081834, EPI_ISL_1081835, EPI_ISL_1129216, EPI_ISL_1129217, EPI_ISL_1129218, EPI_ISL_1129219, EPI_ISL_1129220, EPI_ISL_1129221, EPI_ISL_1129222, EPI_ISL_1129223, EPI_ISL_1129224, EPI_ISL_1129225, EPI_ISL_1129226, EPI_ISL_1129227 | | | |
| see above | Virus Research and Diagnostic Laboratory, Medical College Level, Kasturba Hospital for Infectious Diseases | National Institute of Virology-Microbial Containment Complex, Indian Council of Medical Research | Pragya D. Yadav, Jayanti Shastri |
| EPI_ISL_1129251, EPI_ISL_1129252, EPI_ISL_1129253, EPI_ISL_1129254, EPI_ISL_1129255, EPI_ISL_1129256, EPI_ISL_1129257, EPI_ISL_1129258, EPI_ISL_1129259, EPI_ISL_1129260, EPI_ISL_1129261, EPI_ISL_1129262, EPI_ISL_1129263, EPI_ISL_1129264, EPI_ISL_1129265, EPI_ISL_1129266, EPI_ISL_1129267, EPI_ISL_1129268, EPI_ISL_1129269, EPI_ISL_1129270, EPI_ISL_1129271, EPI_ISL_1129272, EPI_ISL_1129273, EPI_ISL_1129274 | | | |
| see above | VRDL, IGGMC, Nagpur | Indian Council of Medical Research-National Institute of Virology, Microbial Containment Complex | Pragya D. Yadav, Sharmila Raut |
| EPI_ISL_1129275, EPI_ISL_1129276, EPI_ISL_1129277, EPI_ISL_1129278 | Indian Council of Medical Research-National Institute of Virology, Pune | Indian Council of Medical Research-National Institute of Virology, Microbial Containment Complex | Pragya D. Yadav |
| EPI_ISL_1251268 | VRDL, IGGMC, Nagpur | Indian Council of Medical Research-National Institute of Virology, Microbial Containment Complex | Pragya D. Yadav, Sharmila Raut |
| EPI_ISL_1278285, EPI_ISL_1278286, EPI_ISL_1312365 | Indian Council of Medical Research-National Institute of Virology, Microbial Containment Complex | Indian Council of Medical Research-National Institute of Virology, Microbial Containment Complex | Pragya D. Yadav |
| EPI_ISL_1334317, EPI_ISL_1334318, EPI_ISL_1334319, EPI_ISL_1334320, EPI_ISL_1334321, EPI_ISL_1334322, EPI_ISL_1334323, EPI_ISL_1334324, EPI_ISL_1334325 | Clinical Specimen Feces | Enteric Viruses Group, ICMR-National Institute of Virology | Dr. Mallika Lavania, Dr. Varsha Potdar |
| EPI_ISL_1357695 | INSACOG-WB | National Institute of Biomedical Genomics | Arindam Maitra, Bhaswati Bandyopadhyay, Nidhan Kumar Biswas, Tamal Ghosh, Sreedhar Chinnaswamy, Ajay Chakraborti, Saumitra Das |
| EPI_ISL_1360295, EPI_ISL_1360296 | CSIR-National Environmental Engineering Research Institute | CSIR-Centre for Cellular and Molecular Biology - INSACOG | Onkar Kulkarni, Lamuk Zaveri, Ara Sreenivas, Sofia Banu, Shreekant Verma, Amareshwar Vodapalli , Viswagithe S L, B Himasri, Sharath Chandra Thota, Karthik Bharadwaj Tallapaka, Krishna Khairnar, Rakesh K Mishra,Divya Tej Sowpati |
| EPI_ISL_1360297 | CSIR-National Environmental Engineering Research Institute | CSIR-Centre for Cellular and Molecular Biology - INSACOG | Lamuk Zaveri, Ara Sreenivas, Sofia Banu, Onkar Kulkarni, Shreekant Verma, Amareshwar Vodapalli , Viswagithe S L, B Himasri, Sharath Chandra Thota, Karthik Bharadwaj Tallapaka, Krishna Khairnar, Rakesh K Mishra,Divya Tej Sowpati |
| EPI_ISL_1360298 | CSIR-National Environmental Engineering Research Institute | CSIR-Centre for Cellular and Molecular Biology - INSACOG | Lamuk Zaveri, Ara Sreenivas, Onkar Kulkarni, Sofia Banu, Shreekant Verma,Amareshwar Vodapalli, Viswagithe S L, B Himasri, Sharath Chandra Thota, Karthik Bharadwaj Tallapaka, Krishna Khairnar, Rakesh K Mishra,Divya Tej Sowpati |
| EPI_ISL_1360299, EPI_ISL_1360300 | CSIR-National Environmental Engineering Research Institute | CSIR-Centre for Cellular and Molecular Biology - INSACOG | Ara Sreenivas, Onkar Kulkarni, Lamuk Zaveri, Sofia Banu, Shreekant Verma, Amareshwar Vodapalli , Viswagithe S L, B Himasri, Sharath Chandra Thota, Karthik Bharadwaj Tallapaka, Krishna Khairnar, Rakesh K Mishra,Divya Tej Sowpati |
| EPI_ISL_1360301, EPI_ISL_1360302 | CSIR-National Environmental Engineering Research Institute | CSIR-Centre for Cellular and Molecular Biology - INSACOG | Sofia Banu, Lamuk Zaveri, Ara Sreenivas, Shreekant Verma, Onkar Kulkarni, Amareshwar Vodapalli ,Viswagithe S L, B Himasri, Sharath Chandra Thota, Karthik Bharadwaj Tallapaka, Krishna Khairnar, Rakesh K Mishra,Divya Tej Sowpati |
| EPI_ISL_1360303, EPI_ISL_1360304 | CSIR-National Environmental Engineering Research Institute | CSIR-Centre for Cellular and Molecular Biology - INSACOG | Onkar Kulkarni, Lamuk Zaveri, Ara Sreenivas, Sofia Banu, Shreekant Verma, Amareshwar Vodapalli , Viswagithe S L, B Himasri, Sharath Chandra Thota, Karthik Bharadwaj Tallapaka, Krishna Khairnar, Rakesh K Mishra,Divya Tej Sowpati |
| EPI_ISL_1360305 | CSIR-National Environmental Engineering Research Institute | CSIR-Centre for Cellular and Molecular Biology - INSACOG | Lamuk Zaveri, Ara Sreenivas, Sofia Banu, Onkar Kulkarni, Shreekant Verma, Amareshwar Vodapalli , Viswagithe S L, B Himasri, Sharath Chandra Thota, Karthik Bharadwaj Tallapaka, Krishna Khairnar, Rakesh K Mishra,Divya Tej Sowpati |
| EPI_ISL_1360306 | CSIR-National Environmental Engineering Research Institute | CSIR-Centre for Cellular and Molecular Biology - INSACOG | Lamuk Zaveri, Ara Sreenivas, Onkar Kulkarni, Sofia Banu, Shreekant Verma,Amareshwar Vodapalli, Viswagithe S L, B Himasri, Sharath Chandra Thota, Karthik Bharadwaj Tallapaka, Krishna Khairnar, Rakesh K Mishra,Divya Tej Sowpati |
| EPI_ISL_1360307, EPI_ISL_1360308 | CSIR-National Environmental Engineering Research Institute | CSIR-Centre for Cellular and Molecular Biology - INSACOG | Ara Sreenivas, Onkar Kulkarni, Lamuk Zaveri, Sofia Banu, Shreekant Verma, Amareshwar Vodapalli , Viswagithe S L, B Himasri, Sharath Chandra Thota, Karthik Bharadwaj Tallapaka, Krishna Khairnar, Rakesh K Mishra,Divya Tej Sowpati |
| EPI_ISL_1360309, EPI_ISL_1360310 | CSIR-National Environmental Engineering Research Institute | CSIR-Centre for Cellular and Molecular Biology - INSACOG | Sofia Banu, Lamuk Zaveri, Ara Sreenivas, Shreekant Verma, Onkar Kulkarni, Amareshwar Vodapalli ,Viswagithe S L, B Himasri, Sharath Chandra Thota, Karthik Bharadwaj Tallapaka, Krishna Khairnar, Rakesh K Mishra,Divya Tej Sowpati |
| EPI_ISL_1360311, EPI_ISL_1360312 | CSIR-National Environmental Engineering Research Institute | CSIR-Centre for Cellular and Molecular Biology - INSACOG | Onkar Kulkarni, Lamuk Zaveri, Ara Sreenivas, Sofia Banu, Shreekant Verma, Amareshwar Vodapalli , Viswagithe S L, B Himasri, Sharath Chandra Thota, Karthik Bharadwaj Tallapaka, Krishna Khairnar, Rakesh K Mishra,Divya Tej Sowpati |
| EPI_ISL_1360313 | CSIR-National Environmental Engineering Research Institute | CSIR-Centre for Cellular and Molecular Biology - INSACOG | Lamuk Zaveri, Ara Sreenivas, Sofia Banu, Onkar Kulkarni, Shreekant Verma, Amareshwar Vodapalli , Viswagithe S L, B Himasri, Sharath Chandra Thota, Karthik Bharadwaj Tallapaka, Krishna Khairnar, Rakesh K Mishra,Divya Tej Sowpati |
| EPI_ISL_1360314 | CSIR-National Environmental Engineering Research Institute | CSIR-Centre for Cellular and Molecular Biology - INSACOG | Lamuk Zaveri, Ara Sreenivas, Onkar Kulkarni, Sofia Banu, Shreekant Verma,Amareshwar Vodapalli, Viswagithe S L, B Himasri, Sharath Chandra Thota, Karthik Bharadwaj Tallapaka, Krishna Khairnar, Rakesh K Mishra,Divya Tej Sowpati |
| EPI_ISL_1360315, EPI_ISL_1360316 | CSIR-National Environmental Engineering Research Institute | CSIR-Centre for Cellular and Molecular Biology - INSACOG | Ara Sreenivas, Onkar Kulkarni, Lamuk Zaveri, Sofia Banu, Shreekant Verma, Amareshwar Vodapalli , Viswagithe S L, B Himasri, Sharath Chandra Thota, Karthik Bharadwaj Tallapaka, Krishna Khairnar, Rakesh K Mishra,Divya Tej Sowpati |
| EPI_ISL_1360317, EPI_ISL_1360318 | CSIR-National Environmental Engineering Research Institute | CSIR-Centre for Cellular and Molecular Biology - INSACOG | Sofia Banu, Lamuk Zaveri, Ara Sreenivas, Shreekant Verma, Onkar Kulkarni, Amareshwar Vodapalli ,Viswagithe S L, B Himasri, Sharath Chandra Thota, Karthik Bharadwaj Tallapaka, Krishna Khairnar, Rakesh K Mishra,Divya Tej Sowpati |
| EPI_ISL_1360319, EPI_ISL_1360320 | CSIR-National Environmental Engineering Research Institute | CSIR-Centre for Cellular and Molecular Biology - INSACOG | Onkar Kulkarni, Lamuk Zaveri, Ara Sreenivas, Sofia Banu, Shreekant Verma, Amareshwar Vodapalli , Viswagithe S L, B Himasri, Sharath Chandra Thota, Karthik Bharadwaj Tallapaka, Krishna Khairnar, Rakesh K Mishra,Divya Tej Sowpati |
| EPI_ISL_1360321 | CSIR-National Environmental Engineering Research Institute | CSIR-Centre for Cellular and Molecular Biology - INSACOG | Lamuk Zaveri, Ara Sreenivas, Sofia Banu, Onkar Kulkarni, Shreekant Verma, Amareshwar Vodapalli , Viswagithe S L, B Himasri, Sharath Chandra Thota, Karthik Bharadwaj Tallapaka, Krishna Khairnar, Rakesh K Mishra,Divya Tej Sowpati |

| | | | |
|---|---|---|---|
| EPI_ISL_1360322 | CSIR-National Environmental Engineering Research Institute | CSIR-Centre for Cellular and Molecular Biology - INSACOG | Lamuk Zaveri, Ara Sreenivas, Onkar Kulkarni, Sofia Banu, Shreekant Verma,Amareshwar Vodapalli, Viswagithe S L, B Himasri, Sharath Chandra Thota, Karthik Bharadwaj Tallapaka, Krishna Khairnar, Rakesh K Mishra,Divya Tej Sowpati |
| EPI_ISL_1360323, EPI_ISL_1360324 | CSIR-National Environmental Engineering Research Institute | CSIR-Centre for Cellular and Molecular Biology - INSACOG | Ara Sreenivas, Onkar Kulkarni, Lamuk Zaveri, Sofia Banu, Shreekant Verma, Amareshwar Vodapalli , Viswagithe S L, B Himasri, Sharath Chandra Thota, Karthik Bharadwaj Tallapaka, Krishna Khairnar, Rakesh K Mishra,Divya Tej Sowpati |
| EPI_ISL_1360325, EPI_ISL_1360326 | CSIR-National Environmental Engineering Research Institute | CSIR-Centre for Cellular and Molecular Biology - INSACOG | Sofia Banu, Lamuk Zaveri, Ara Sreenivas, Shreekant Verma, Onkar Kulkarni, Amareshwar Vodapalli ,Viswagithe S L, B Himasri, Sharath Chandra Thota, Karthik Bharadwaj Tallapaka, Krishna Khairnar, Rakesh K Mishra,Divya Tej Sowpati |
| EPI_ISL_1360327, EPI_ISL_1360328 | CSIR-National Environmental Engineering Research Institute | CSIR-Centre for Cellular and Molecular Biology - INSACOG | Onkar Kulkarni, Lamuk Zaveri, Ara Sreenivas, Sofia Banu, Shreekant Verma, Amareshwar Vodapalli , Viswagithe S L, B Himasri, Sharath Chandra Thota, Karthik Bharadwaj Tallapaka, Krishna Khairnar, Rakesh K Mishra,Divya Tej Sowpati |
| EPI_ISL_1360329 | CSIR-National Environmental Engineering Research Institute | CSIR-Centre for Cellular and Molecular Biology - INSACOG | Lamuk Zaveri, Ara Sreenivas, Sofia Banu, Onkar Kulkarni, Shreekant Verma, Amareshwar Vodapalli , Viswagithe S L, B Himasri, Sharath Chandra Thota, Karthik Bharadwaj Tallapaka, Krishna Khairnar, Rakesh K Mishra,Divya Tej Sowpati |
| EPI_ISL_1360330 | CSIR-National Environmental Engineering Research Institute | CSIR-Centre for Cellular and Molecular Biology - INSACOG | Lamuk Zaveri, Ara Sreenivas, Onkar Kulkarni, Sofia Banu, Shreekant Verma,Amareshwar Vodapalli, Viswagithe S L, B Himasri, Sharath Chandra Thota, Karthik Bharadwaj Tallapaka, Krishna Khairnar, Rakesh K Mishra,Divya Tej Sowpati |
| EPI_ISL_1360331, EPI_ISL_1360332 | CSIR-National Environmental Engineering Research Institute | CSIR-Centre for Cellular and Molecular Biology - INSACOG | Ara Sreenivas, Onkar Kulkarni, Lamuk Zaveri, Sofia Banu, Shreekant Verma, Amareshwar Vodapalli , Viswagithe S L, B Himasri, Sharath Chandra Thota, Karthik Bharadwaj Tallapaka, Krishna Khairnar, Rakesh K Mishra,Divya Tej Sowpati |
| EPI_ISL_1360333, EPI_ISL_1360334 | CSIR-National Environmental Engineering Research Institute | CSIR-Centre for Cellular and Molecular Biology - INSACOG | Sofia Banu, Lamuk Zaveri, Ara Sreenivas, Shreekant Verma, Onkar Kulkarni, Amareshwar Vodapalli ,Viswagithe S L, B Himasri, Sharath Chandra Thota, Karthik Bharadwaj Tallapaka, Krishna Khairnar, Rakesh K Mishra,Divya Tej Sowpati |
| EPI_ISL_1360335, EPI_ISL_1360336 | CSIR-National Environmental Engineering Research Institute | CSIR-Centre for Cellular and Molecular Biology - INSACOG | Onkar Kulkarni, Lamuk Zaveri, Ara Sreenivas, Sofia Banu, Shreekant Verma, Amareshwar Vodapalli , Viswagithe S L, B Himasri, Sharath Chandra Thota, Karthik Bharadwaj Tallapaka, Krishna Khairnar, Rakesh K Mishra,Divya Tej Sowpati |
| EPI_ISL_1360337 | CSIR-National Environmental Engineering Research Institute | CSIR-Centre for Cellular and Molecular Biology - INSACOG | Lamuk Zaveri, Ara Sreenivas, Sofia Banu, Onkar Kulkarni, Shreekant Verma, Amareshwar Vodapalli , Viswagithe S L, B Himasri, Sharath Chandra Thota, Karthik Bharadwaj Tallapaka, Krishna Khairnar, Rakesh K Mishra,Divya Tej Sowpati |
| EPI_ISL_1360338 | CSIR-National Environmental Engineering Research Institute | CSIR-Centre for Cellular and Molecular Biology - INSACOG | Lamuk Zaveri, Ara Sreenivas, Onkar Kulkarni, Sofia Banu, Shreekant Verma,Amareshwar Vodapalli, Viswagithe S L, B Himasri, Sharath Chandra Thota, Karthik Bharadwaj Tallapaka, Krishna Khairnar, Rakesh K Mishra,Divya Tej Sowpati |
| EPI_ISL_1360339, EPI_ISL_1360340 | CSIR-National Environmental Engineering Research Institute | CSIR-Centre for Cellular and Molecular Biology - INSACOG | Ara Sreenivas, Onkar Kulkarni, Lamuk Zaveri, Sofia Banu, Shreekant Verma, Amareshwar Vodapalli , Viswagithe S L, B Himasri, Sharath Chandra Thota, Karthik Bharadwaj Tallapaka, Krishna Khairnar, Rakesh K Mishra,Divya Tej Sowpati |
| EPI_ISL_1360341, EPI_ISL_1360342 | CSIR-National Environmental Engineering Research Institute | CSIR-Centre for Cellular and Molecular Biology - INSACOG | Sofia Banu, Lamuk Zaveri, Ara Sreenivas, Shreekant Verma, Onkar Kulkarni, Amareshwar Vodapalli ,Viswagithe S L, B Himasri, Sharath Chandra Thota, Karthik Bharadwaj Tallapaka, Krishna Khairnar, Rakesh K Mishra,Divya Tej Sowpati |
| EPI_ISL_1360343, EPI_ISL_1360344 | CSIR-National Environmental Engineering Research Institute | CSIR-Centre for Cellular and Molecular Biology - INSACOG | Onkar Kulkarni, Lamuk Zaveri, Ara Sreenivas, Sofia Banu, Shreekant Verma, Amareshwar Vodapalli , Viswagithe S L, B Himasri, Sharath Chandra Thota, Karthik Bharadwaj Tallapaka, Krishna Khairnar, Rakesh K Mishra,Divya Tej Sowpati |
| EPI_ISL_1360345 | CSIR-National Environmental Engineering Research Institute | CSIR-Centre for Cellular and Molecular Biology - INSACOG | Lamuk Zaveri, Ara Sreenivas, Sofia Banu, Onkar Kulkarni, Shreekant Verma, Amareshwar Vodapalli , Viswagithe S L, B Himasri, Sharath Chandra Thota, Karthik Bharadwaj Tallapaka, Krishna Khairnar, Rakesh K Mishra,Divya Tej Sowpati |
| EPI_ISL_1360346 | CSIR-National Environmental Engineering Research Institute | CSIR-Centre for Cellular and Molecular Biology - INSACOG | Lamuk Zaveri, Ara Sreenivas, Onkar Kulkarni, Sofia Banu, Shreekant Verma,Amareshwar Vodapalli, Viswagithe S L, B Himasri, Sharath Chandra Thota, Karthik Bharadwaj Tallapaka, Krishna Khairnar, Rakesh K Mishra,Divya Tej Sowpati |
| EPI_ISL_1360347, EPI_ISL_1360348 | CSIR-National Environmental Engineering Research Institute | CSIR-Centre for Cellular and Molecular Biology - INSACOG | Ara Sreenivas, Onkar Kulkarni, Lamuk Zaveri, Sofia Banu, Shreekant Verma, Amareshwar Vodapalli , Viswagithe S L, B Himasri, Sharath Chandra Thota, Karthik Bharadwaj Tallapaka, Krishna Khairnar, Rakesh K Mishra,Divya Tej Sowpati |
| EPI_ISL_1360349, EPI_ISL_1360350 | CSIR-National Environmental Engineering Research Institute | CSIR-Centre for Cellular and Molecular Biology - INSACOG | Sofia Banu, Lamuk Zaveri, Ara Sreenivas, Shreekant Verma, Onkar Kulkarni, Amareshwar Vodapalli ,Viswagithe S L, B Himasri, Sharath Chandra Thota, Karthik Bharadwaj Tallapaka, Krishna Khairnar, Rakesh K Mishra,Divya Tej Sowpati |
| EPI_ISL_1360351, EPI_ISL_1360352 | CSIR-National Environmental Engineering Research Institute | CSIR-Centre for Cellular and Molecular Biology - INSACOG | Onkar Kulkarni, Lamuk Zaveri, Ara Sreenivas, Sofia Banu, Shreekant Verma, Amareshwar Vodapalli , Viswagithe S L, B Himasri, Sharath Chandra Thota, Karthik Bharadwaj Tallapaka, Krishna Khairnar, Rakesh K Mishra,Divya Tej Sowpati |
| EPI_ISL_1360353 | CSIR-National Environmental Engineering Research Institute | CSIR-Centre for Cellular and Molecular Biology - INSACOG | Lamuk Zaveri, Ara Sreenivas, Sofia Banu, Onkar Kulkarni, Shreekant Verma, Amareshwar Vodapalli , Viswagithe S L, B Himasri, Sharath Chandra Thota, Karthik Bharadwaj Tallapaka, Krishna Khairnar, Rakesh K Mishra,Divya Tej Sowpati |
| EPI_ISL_1360354, EPI_ISL_1360355 | CSIR-National Environmental Engineering Research Institute | CSIR-Centre for Cellular and Molecular Biology - INSACOG | Ara Sreenivas, Onkar Kulkarni, Lamuk Zaveri, Sofia Banu, Shreekant Verma, Amareshwar Vodapalli , Viswagithe S L, B Himasri, Sharath Chandra Thota, Karthik Bharadwaj Tallapaka, Krishna Khairnar, Rakesh K Mishra,Divya Tej Sowpati |
| EPI_ISL_1360356, EPI_ISL_1360357 | CSIR-National Environmental Engineering Research Institute | CSIR-Centre for Cellular and Molecular Biology - INSACOG | Sofia Banu, Lamuk Zaveri, Ara Sreenivas, Shreekant Verma, Onkar Kulkarni, Amareshwar Vodapalli ,Viswagithe S L, B Himasri, Sharath Chandra Thota, Karthik Bharadwaj Tallapaka, Krishna Khairnar, Rakesh K Mishra,Divya Tej Sowpati |
| EPI_ISL_1360358 | CSIR-National Environmental Engineering Research Institute | CSIR-Centre for Cellular and Molecular Biology - INSACOG | Onkar Kulkarni, Lamuk Zaveri, Ara Sreenivas, Sofia Banu, Shreekant Verma, Amareshwar Vodapalli , Viswagithe S L, B Himasri, Sharath Chandra Thota, Karthik Bharadwaj Tallapaka, Krishna Khairnar, Rakesh K Mishra,Divya Tej Sowpati |
| EPI_ISL_1360359 | CSIR-National Environmental Engineering Research Institute | CSIR-Centre for Cellular and Molecular Biology - INSACOG | Lamuk Zaveri, Ara Sreenivas, Sofia Banu, Onkar Kulkarni, Shreekant Verma, Amareshwar Vodapalli , Viswagithe S L, B Himasri, Sharath Chandra Thota, Karthik Bharadwaj Tallapaka, Krishna Khairnar, Rakesh K Mishra,Divya Tej Sowpati |
| EPI_ISL_1360360 | CSIR-National Environmental Engineering Research Institute | CSIR-Centre for Cellular and Molecular Biology - INSACOG | Lamuk Zaveri, Ara Sreenivas, Onkar Kulkarni, Sofia Banu, Shreekant Verma,Amareshwar Vodapalli, Viswagithe S L, B Himasri, Sharath Chandra Thota, Karthik Bharadwaj Tallapaka, Krishna Khairnar, Rakesh K Mishra,Divya Tej Sowpati |
| EPI_ISL_1360361, EPI_ISL_1360362 | CSIR-National Environmental Engineering Research Institute | CSIR-Centre for Cellular and Molecular Biology - INSACOG | Ara Sreenivas, Onkar Kulkarni, Lamuk Zaveri, Sofia Banu, Shreekant Verma, Amareshwar Vodapalli , Viswagithe S L, B Himasri, Sharath Chandra Thota, Karthik Bharadwaj Tallapaka, Krishna Khairnar, Rakesh K Mishra,Divya Tej Sowpati |
| EPI_ISL_1360363, EPI_ISL_1360364 | CSIR-National Environmental Engineering Research Institute | CSIR-Centre for Cellular and Molecular Biology - INSACOG | Sofia Banu, Lamuk Zaveri, Ara Sreenivas, Shreekant Verma, Onkar Kulkarni, Amareshwar Vodapalli ,Viswagithe S L, B Himasri, Sharath Chandra Thota, Karthik Bharadwaj Tallapaka, Krishna Khairnar, Rakesh K Mishra,Divya Tej Sowpati |
| EPI_ISL_1360365, EPI_ISL_1360366 | CSIR-National Environmental Engineering Research Institute | CSIR-Centre for Cellular and Molecular Biology - INSACOG | Onkar Kulkarni, Lamuk Zaveri, Ara Sreenivas, Sofia Banu, Shreekant Verma, Amareshwar Vodapalli , Viswagithe S L, B Himasri, Sharath Chandra Thota, Karthik Bharadwaj Tallapaka, Krishna Khairnar, Rakesh K Mishra,Divya Tej Sowpati |
| EPI_ISL_1360367 | CSIR-National Environmental Engineering Research Institute | CSIR-Centre for Cellular and Molecular Biology - INSACOG | Lamuk Zaveri, Ara Sreenivas, Sofia Banu, Onkar Kulkarni, Shreekant Verma, Amareshwar Vodapalli , Viswagithe S L, B Himasri, Sharath Chandra Thota, Karthik Bharadwaj Tallapaka, Krishna Khairnar, Rakesh K Mishra,Divya Tej Sowpati |
| EPI_ISL_1360368 | CSIR-National Environmental Engineering Research Institute | CSIR-Centre for Cellular and Molecular Biology - INSACOG | Lamuk Zaveri, Ara Sreenivas, Onkar Kulkarni, Sofia Banu, Shreekant Verma,Amareshwar Vodapalli, Viswagithe S L, B Himasri, Sharath Chandra Thota, Karthik Bharadwaj Tallapaka, Krishna Khairnar, Rakesh K Mishra,Divya Tej Sowpati |
| EPI_ISL_1360369, EPI_ISL_1360370 | CSIR-National Environmental Engineering Research Institute | CSIR-Centre for Cellular and Molecular Biology - INSACOG | Ara Sreenivas, Onkar Kulkarni, Lamuk Zaveri, Sofia Banu, Shreekant Verma, Amareshwar Vodapalli , Viswagithe S L, B Himasri, Sharath Chandra Thota, Karthik Bharadwaj Tallapaka, Krishna Khairnar, Rakesh K Mishra,Divya Tej Sowpati |
| EPI_ISL_1360371, EPI_ISL_1360372 | CSIR-National Environmental Engineering Research Institute | CSIR-Centre for Cellular and Molecular Biology - INSACOG | Sofia Banu, Lamuk Zaveri, Ara Sreenivas, Shreekant Verma, Onkar Kulkarni, Amareshwar Vodapalli ,Viswagithe S L, B Himasri, Sharath Chandra Thota, Karthik Bharadwaj Tallapaka, Krishna Khairnar, Rakesh K Mishra,Divya Tej Sowpati |
| EPI_ISL_1360373, EPI_ISL_1360374 | CSIR-National Environmental Engineering Research Institute | CSIR-Centre for Cellular and Molecular Biology - INSACOG | Onkar Kulkarni, Lamuk Zaveri, Ara Sreenivas, Sofia Banu, Shreekant Verma, Amareshwar Vodapalli , Viswagithe S L, B Himasri, Sharath Chandra Thota, Karthik Bharadwaj Tallapaka, Krishna Khairnar, Rakesh K Mishra,Divya Tej Sowpati |
| EPI_ISL_1360375 | CSIR-National Environmental Engineering Research Institute | CSIR-Centre for Cellular and Molecular Biology - INSACOG | Lamuk Zaveri, Ara Sreenivas, Sofia Banu, Onkar Kulkarni, Shreekant Verma, Amareshwar Vodapalli , Viswagithe S L, B Himasri, Sharath Chandra Thota, Karthik Bharadwaj Tallapaka, Krishna Khairnar, Rakesh K Mishra,Divya Tej Sowpati |
| EPI_ISL_1360376 | CSIR-National Environmental Engineering Research Institute | CSIR-Centre for Cellular and Molecular Biology - INSACOG | Lamuk Zaveri, Ara Sreenivas, Onkar Kulkarni, Sofia Banu, Shreekant Verma,Amareshwar Vodapalli, Viswagithe S L, B Himasri, Sharath Chandra Thota, Karthik Bharadwaj Tallapaka, Krishna Khairnar, Rakesh K Mishra,Divya Tej Sowpati |

| | | | |
|---|---|---|---|
| EPI_ISL_1360377, EPI_ISL_1360378 | CSIR-National Environmental Engineering Research Institute | CSIR-Centre for Cellular and Molecular Biology - INSACOG | Ara Sreenivas, Onkar Kulkarni, Lamuk Zaveri, Sofia Banu, Shreekant Verma, Amareshwar Vodapalli , Viswagithe S L, B Himasri, Sharath Chandra Thota, Karthik Bharadwaj Tallapaka, Krishna Khairnar, Rakesh K Mishra,Divya Tej Sowpati |
| EPI_ISL_1360379, EPI_ISL_1360380 | CSIR-National Environmental Engineering Research Institute | CSIR-Centre for Cellular and Molecular Biology - INSACOG | Sofia Banu, Lamuk Zaveri, Ara Sreenivas, Shreekant Verma, Onkar Kulkarni, Amareshwar Vodapalli ,Viswagithe S L, B Himasri, Sharath Chandra Thota, Karthik Bharadwaj Tallapaka, Krishna Khairnar, Rakesh K Mishra,Divya Tej Sowpati |
| EPI_ISL_1360381, EPI_ISL_1360382 | CSIR-National Environmental Engineering Research Institute | CSIR-Centre for Cellular and Molecular Biology - INSACOG | Onkar Kulkarni, Lamuk Zaveri, Ara Sreenivas, Sofia Banu, Shreekant Verma, Amareshwar Vodapalli , Viswagithe S L, B Himasri, Sharath Chandra Thota, Karthik Bharadwaj Tallapaka, Krishna Khairnar, Rakesh K Mishra,Divya Tej Sowpati |
| EPI_ISL_1360383 | CSIR-National Environmental Engineering Research Institute | CSIR-Centre for Cellular and Molecular Biology - INSACOG | Lamuk Zaveri, Ara Sreenivas, Sofia Banu, Onkar Kulkarni, Shreekant Verma, Amareshwar Vodapalli , Viswagithe S L, B Himasri, Sharath Chandra Thota, Karthik Bharadwaj Tallapaka, Krishna Khairnar, Rakesh K Mishra,Divya Tej Sowpati |
| EPI_ISL_1360384 | CSIR-National Environmental Engineering Research Institute | CSIR-Centre for Cellular and Molecular Biology - INSACOG | Lamuk Zaveri, Ara Sreenivas, Onkar Kulkarni, Sofia Banu, Shreekant Verma,Amareshwar Vodapalli, Viswagithe S L, B Himasri, Sharath Chandra Thota, Karthik Bharadwaj Tallapaka, Krishna Khairnar, Rakesh K Mishra,Divya Tej Sowpati |
| EPI_ISL_1360385, EPI_ISL_1360386 | CSIR-National Environmental Engineering Research Institute | CSIR-Centre for Cellular and Molecular Biology - INSACOG | Ara Sreenivas, Onkar Kulkarni, Lamuk Zaveri, Sofia Banu, Shreekant Verma, Amareshwar Vodapalli , Viswagithe S L, B Himasri, Sharath Chandra Thota, Karthik Bharadwaj Tallapaka, Krishna Khairnar, Rakesh K Mishra,Divya Tej Sowpati |
| EPI_ISL_1360387, EPI_ISL_1360388 | CSIR-National Environmental Engineering Research Institute | CSIR-Centre for Cellular and Molecular Biology - INSACOG | Sofia Banu, Lamuk Zaveri, Ara Sreenivas, Shreekant Verma, Onkar Kulkarni, Amareshwar Vodapalli ,Viswagithe S L, B Himasri, Sharath Chandra Thota, Karthik Bharadwaj Tallapaka, Krishna Khairnar, Rakesh K Mishra,Divya Tej Sowpati |
| EPI_ISL_1372279 | CSIR-National Environmental Engineering Research Institute | CSIR-Centre for Cellular and Molecular Biology - INSACOG | Lamuk Zaveri, Ara Sreenivas, Onkar Kulkarni, Sofia Banu, Shreekant Verma,Amareshwar Vodapalli, Viswagithe S L, B Himasri, Sharath Chandra Thota, Karthik Bharadwaj Tallapaka, Krishna Khairnar, Rakesh K Mishra,Divya Tej Sowpati |
| EPI_ISL_1384813, EPI_ISL_1384814, EPI_ISL_1384815, EPI_ISL_1384816, EPI_ISL_1384817 | National Centre For Cell Science | National Centre For Cell Science - INSACOG | Dhiraj Paul, Mitali Inamdar, Sonal Manik Chavan, Mohak P Gujare, Shivang P. Bhanushali, Manoj Kumar Bhat, Ajay Pillai, INSACOG Consortium team, Yogesh Shouche. |
| EPI_ISL_1384818 | National Centre For Cell Science | National Centre For Cell Science | Dhiraj Paul, Sonal Manik Chavan, Mohak P Gujare, Shivang P. Bhanushali, Mitali Inamdar, Manoj Kumar Bhat, Ajay Pillai, Yogesh Shouche |
| EPI_ISL_1384819, EPI_ISL_1384820, EPI_ISL_1384821, EPI_ISL_1384822, EPI_ISL_1384823, EPI_ISL_1384824, EPI_ISL_1384825, EPI_ISL_1384826, EPI_ISL_1384827, EPI_ISL_1384828, EPI_ISL_1384829, EPI_ISL_1384830, EPI_ISL_1384831, EPI_ISL_1384832, EPI_ISL_1384833, EPI_ISL_1384834, EPI_ISL_1384835, EPI_ISL_1384836, EPI_ISL_1384837, EPI_ISL_1384838, EPI_ISL_1384839, EPI_ISL_1384840, EPI_ISL_1384841, EPI_ISL_1384842, EPI_ISL_1384843, EPI_ISL_1384844, EPI_ISL_1384845, EPI_ISL_1384846, EPI_ISL_1384847, EPI_ISL_1384848, EPI_ISL_1384849, EPI_ISL_1384850, EPI_ISL_1384851, EPI_ISL_1384852, EPI_ISL_1384854, EPI_ISL_1384855, EPI_ISL_1384862, EPI_ISL_1384864, EPI_ISL_1384865, EPI_ISL_1384866, EPI_ISL_1384867, EPI_ISL_1384868, EPI_ISL_1415087, EPI_ISL_1415088, EPI_ISL_1415089, EPI_ISL_1415090, EPI_ISL_1415091, EPI_ISL_1415092, EPI_ISL_1415093, EPI_ISL_1415094, EPI_ISL_1415095, EPI_ISL_1415096, EPI_ISL_1415098, EPI_ISL_1415099, EPI_ISL_1415100, EPI_ISL_1415101, EPI_ISL_1415102, EPI_ISL_1415103, EPI_ISL_1415104, EPI_ISL_1415105, EPI_ISL_1415106, EPI_ISL_1415107, EPI_ISL_1415108, EPI_ISL_1415109, EPI_ISL_1415110, EPI_ISL_1415111, EPI_ISL_1415112, EPI_ISL_1415114, EPI_ISL_1415115, EPI_ISL_1415118, EPI_ISL_1415120, EPI_ISL_1415121, EPI_ISL_1415122, EPI_ISL_1415125, EPI_ISL_1415126, EPI_ISL_1415127, EPI_ISL_1415128, EPI_ISL_1415132, EPI_ISL_1415133, EPI_ISL_1415137, EPI_ISL_1415138, EPI_ISL_1415139, EPI_ISL_1415140, EPI_ISL_1415142, EPI_ISL_1415143, EPI_ISL_1415144, EPI_ISL_1415145, EPI_ISL_1415146, EPI_ISL_1415147, EPI_ISL_1415148, EPI_ISL_1415149, EPI_ISL_1415150, EPI_ISL_1415151, EPI_ISL_1415152, EPI_ISL_1415153, EPI_ISL_1415154, EPI_ISL_1415155, EPI_ISL_1415156, EPI_ISL_1415157, EPI_ISL_1415158, EPI_ISL_1415159, EPI_ISL_1415160, EPI_ISL_1415162, EPI_ISL_1415164, EPI_ISL_1415165, EPI_ISL_1415166, EPI_ISL_1415168, EPI_ISL_1415169, EPI_ISL_1415171, EPI_ISL_1415172, EPI_ISL_1415173, EPI_ISL_1415174, EPI_ISL_1415175, EPI_ISL_1415176, EPI_ISL_1415177, EPI_ISL_1415178, EPI_ISL_1415179, EPI_ISL_1415180, EPI_ISL_1415181, EPI_ISL_1415182, EPI_ISL_1415185, EPI_ISL_1415186, EPI_ISL_1415187, EPI_ISL_1415188, EPI_ISL_1415189, EPI_ISL_1415190, EPI_ISL_1415192, EPI_ISL_1415193, EPI_ISL_1415194, EPI_ISL_1415196, EPI_ISL_1415197, EPI_ISL_1415199, EPI_ISL_1415200, EPI_ISL_1415201, EPI_ISL_1415203, EPI_ISL_1415206, EPI_ISL_1415208, EPI_ISL_1415210, EPI_ISL_1415212, EPI_ISL_1415213, EPI_ISL_1415214, EPI_ISL_1415215, EPI_ISL_1415216, EPI_ISL_1415218, EPI_ISL_1415219, EPI_ISL_1415220, EPI_ISL_1415225, EPI_ISL_1415226, EPI_ISL_1415227, EPI_ISL_1415228, EPI_ISL_1415230, EPI_ISL_1415232, EPI_ISL_1415233, EPI_ISL_1415234, EPI_ISL_1415235, EPI_ISL_1415236, EPI_ISL_1415237, EPI_ISL_1415238, EPI_ISL_1415242, EPI_ISL_1415243, EPI_ISL_1415244, EPI_ISL_1415246, EPI_ISL_1415248, EPI_ISL_1415250, EPI_ISL_1415252, EPI_ISL_1415253, EPI_ISL_1415254, EPI_ISL_1415255, EPI_ISL_1415256, EPI_ISL_1415257, EPI_ISL_1415258, EPI_ISL_1415261, EPI_ISL_1415262, EPI_ISL_1415263, EPI_ISL_1415264, EPI_ISL_1415265, EPI_ISL_1415266, EPI_ISL_1415267, EPI_ISL_1415268, EPI_ISL_1415269, EPI_ISL_1415270, EPI_ISL_1415271, EPI_ISL_1415272, EPI_ISL_1415273, EPI_ISL_1415274, EPI_ISL_1415275, EPI_ISL_1415276, EPI_ISL_1415277, EPI_ISL_1415278, EPI_ISL_1415280, EPI_ISL_1415281, EPI_ISL_1415282, EPI_ISL_1415283, EPI_ISL_1415284, EPI_ISL_1415285, EPI_ISL_1415286, EPI_ISL_1415287, EPI_ISL_1415288, EPI_ISL_1415289, EPI_ISL_1415290, EPI_ISL_1415291, EPI_ISL_1415292, EPI_ISL_1415294, EPI_ISL_1415295, EPI_ISL_1415296, EPI_ISL_1415297, EPI_ISL_1415298, EPI_ISL_1415299, EPI_ISL_1415300, EPI_ISL_1415301, EPI_ISL_1415302, EPI_ISL_1415303, EPI_ISL_1415304, EPI_ISL_1415305, EPI_ISL_1415306, EPI_ISL_1415307, EPI_ISL_1415308, EPI_ISL_1415309, EPI_ISL_1415310, EPI_ISL_1415311, EPI_ISL_1415312, EPI_ISL_1415313, EPI_ISL_1415314, EPI_ISL_1415315, EPI_ISL_1415316, EPI_ISL_1415317, EPI_ISL_1415318, EPI_ISL_1415319, EPI_ISL_1415320, EPI_ISL_1415321, EPI_ISL_1415322, EPI_ISL_1415323, EPI_ISL_1415324, EPI_ISL_1415325, EPI_ISL_1415327, EPI_ISL_1415328, EPI_ISL_1415330, EPI_ISL_1415332, EPI_ISL_1415333, EPI_ISL_1415335, EPI_ISL_1415337, EPI_ISL_1415338, EPI_ISL_1415340, EPI_ISL_1415341, EPI_ISL_1415342, EPI_ISL_1415347, EPI_ISL_1415348, EPI_ISL_1415349, EPI_ISL_1415350, EPI_ISL_1415352, EPI_ISL_1415353, EPI_ISL_1415355, EPI_ISL_1415356, EPI_ISL_1415357, EPI_ISL_1415358, EPI_ISL_1415359, EPI_ISL_1415361, EPI_ISL_1415362, EPI_ISL_1415364, EPI_ISL_1415366, EPI_ISL_1415370, EPI_ISL_1415371, EPI_ISL_1415372, EPI_ISL_1415373, EPI_ISL_1415375, EPI_ISL_1415377, EPI_ISL_1415378, EPI_ISL_1415380, EPI_ISL_1415381, EPI_ISL_1415382, EPI_ISL_1415383, EPI_ISL_1415384, EPI_ISL_1415386, EPI_ISL_1415387, EPI_ISL_1415390, EPI_ISL_1415391 | | | |
| see above | National Centre For Cell Science | National Centre For Cell Science - INSACOG | Dhiraj Paul, Mitali Inamdar, Sonal Manik Chavan, Mohak P Gujare, Shivang P. Bhanushali, Manoj Kumar Bhat, Ajay Pillai, INSACOG Consortium team, Yogesh Shouche. |
| EPI_ISL_1419052 | INSACOG-WB | National Institute of Biomedical Genomics - INSACOG | Arindam Maitra, Bhaswati Bandyopadhyay, Nidhan Kumar Biswas, Tamal Ghosh, Sreedhar Chinnaswamy, Ajay Chakraborti, Saumitra Das |
| EPI_ISL_1511201, EPI_ISL_1511202, EPI_ISL_1533729, EPI_ISL_1533732, EPI_ISL_1533734, EPI_ISL_1533735, EPI_ISL_1533739, EPI_ISL_1533741, EPI_ISL_1533771, EPI_ISL_1533772, EPI_ISL_1533773, EPI_ISL_1533774, EPI_ISL_1533775, EPI_ISL_1533776, EPI_ISL_1533777, EPI_ISL_1533778, EPI_ISL_1533779, EPI_ISL_1533780, EPI_ISL_1533781, EPI_ISL_1533782, EPI_ISL_1533783, EPI_ISL_1533784, EPI_ISL_1533785, EPI_ISL_1533786, EPI_ISL_1533787, EPI_ISL_1533788, EPI_ISL_1533789, EPI_ISL_1533790, EPI_ISL_1533791, EPI_ISL_1533792, EPI_ISL_1533793, EPI_ISL_1533794, EPI_ISL_1533795, EPI_ISL_1533796, EPI_ISL_1533797, EPI_ISL_1533798, EPI_ISL_1533799, EPI_ISL_1533800, EPI_ISL_1533801 | | | |
| see above | National Centre For Cell Science | National Centre For Cell Science - INSACOG | Dhiraj Paul, Sonal Manik Chavan, Mohak P Gujare, Shivang P. Bhanushali, Mitali Inamdar, Manoj Kumar Bhat, Ajay Pillai, INSACOG Consortium team, Yogesh Shouche |
| EPI_ISL_1540451, EPI_ISL_1540452, EPI_ISL_1540453, EPI_ISL_1540454, EPI_ISL_1540455, EPI_ISL_1540456, EPI_ISL_1540457 | National Centre For Cell Science | National Centre For Cell Science - INSACOG | Dhiraj Paul, Mohak P Gujare, Shivang P. Bhanushali, Mitali Inamdar, Sonal Manik Chavan, Manoj Kumar Bhat, Ajay Pillai, INSACOG Consortium team, Yogesh Shouche |
| EPI_ISL_1543988, EPI_ISL_1543989, EPI_ISL_1543990, EPI_ISL_1543991, EPI_ISL_1543992, EPI_ISL_1543993, EPI_ISL_1543994, EPI_ISL_1543995, EPI_ISL_1543996, EPI_ISL_1543997, EPI_ISL_1543998, EPI_ISL_1543999, EPI_ISL_1544000, EPI_ISL_1544001, EPI_ISL_1544002, EPI_ISL_1544003, EPI_ISL_1544004, EPI_ISL_1544005, EPI_ISL_1544006, EPI_ISL_1544007, EPI_ISL_1544008, EPI_ISL_1544009, EPI_ISL_1544010, EPI_ISL_1544011, EPI_ISL_1544012, EPI_ISL_1544013, EPI_ISL_1544014, EPI_ISL_1544015, EPI_ISL_1544016, EPI_ISL_1544017, EPI_ISL_1544018, EPI_ISL_1544019, EPI_ISL_1544020, EPI_ISL_1544021, EPI_ISL_1544022, EPI_ISL_1544023, EPI_ISL_1544024, EPI_ISL_1544025, EPI_ISL_1544026, EPI_ISL_1544027, EPI_ISL_1544028, EPI_ISL_1544029, EPI_ISL_1544030, EPI_ISL_1544031, EPI_ISL_1544032, EPI_ISL_1544033, EPI_ISL_1544034, EPI_ISL_1544035, EPI_ISL_1544036, EPI_ISL_1544037, EPI_ISL_1544038, EPI_ISL_1544039, EPI_ISL_1544040, EPI_ISL_1544041, EPI_ISL_1544042, EPI_ISL_1544043, EPI_ISL_1544044, EPI_ISL_1544045, EPI_ISL_1544046, EPI_ISL_1544047, EPI_ISL_1544048, EPI_ISL_1544049, EPI_ISL_1544050, EPI_ISL_1544051, EPI_ISL_1544052, EPI_ISL_1544053, EPI_ISL_1544054, EPI_ISL_1544055, EPI_ISL_1544056, EPI_ISL_1544057, EPI_ISL_1544058, EPI_ISL_1544059, EPI_ISL_1544060, EPI_ISL_1544061, EPI_ISL_1544062, EPI_ISL_1544063, EPI_ISL_1544064, EPI_ISL_1544065, EPI_ISL_1544066, EPI_ISL_1544067, EPI_ISL_1544068, EPI_ISL_1544069, EPI_ISL_1544070, EPI_ISL_1544071, EPI_ISL_1544072, EPI_ISL_1544073, EPI_ISL_1544074 | | | |
| see above | National Centre For Cell Science | National Centre For Cell Science - INSACOG | Dhiraj Paul, Mohak P Gujare, Shivang P. Bhanushali, Mitali Inamdar, Sonal Manik Chavan, Manoj Kumar Bhat, Ajay Pillai, INSACOG Consortium team, Yogesh Shouche |
| EPI_ISL_1547802, EPI_ISL_1547803, EPI_ISL_1547804, EPI_ISL_1547805 | CSIR-National Environmental Engineering Research Institute | CSIR-Centre for Cellular and Molecular Biology - INSACOG | Onkar Kulkarni, Lamuk Zaveri, Ara Sreenivas, Sofia Banu, Shreekant Verma, Amareshwar Vodapalli , Viswagithe S L, B Himasri, Sharath Chandra Thota, Karthik Bharadwaj Tallapaka, Krishna Khairnar, Rakesh K Mishra,Divya Tej Sowpati |
| EPI_ISL_1647423, EPI_ISL_1647424, EPI_ISL_1647425, EPI_ISL_1647426, EPI_ISL_1647427, EPI_ISL_1647428, EPI_ISL_1647429, EPI_ISL_1647430, EPI_ISL_1647431, EPI_ISL_1647432, EPI_ISL_1647433, EPI_ISL_1647434, EPI_ISL_1647435, EPI_ISL_1647436, EPI_ISL_1647437, EPI_ISL_1647438, EPI_ISL_1647439, EPI_ISL_1647440, EPI_ISL_1647441, EPI_ISL_1647442, EPI_ISL_1647443, EPI_ISL_1647444, EPI_ISL_1647445, EPI_ISL_1647446, EPI_ISL_1647447, EPI_ISL_1647448, EPI_ISL_1647449, EPI_ISL_1647450, EPI_ISL_1647451, EPI_ISL_1647452, EPI_ISL_1647453, EPI_ISL_1647454, EPI_ISL_1647455 | | | |
| see above | Indian Council of Medical Research-National Institute of Virology, Microbial Containment Complex | Indian Council of Medical Research-National Institute of Virology, Microbial Containment Complex | Pragya D Yadav |
| EPI_ISL_1663363, EPI_ISL_1663364, EPI_ISL_1663365, EPI_ISL_1663366, EPI_ISL_1663367, EPI_ISL_1663368, EPI_ISL_1663369, EPI_ISL_1663370, EPI_ISL_1663371, EPI_ISL_1663372, EPI_ISL_1663373, EPI_ISL_1663374, EPI_ISL_1663375, EPI_ISL_1663376, EPI_ISL_1663377, EPI_ISL_1663378, EPI_ISL_1663379, EPI_ISL_1663380, EPI_ISL_1663381, EPI_ISL_1663382, EPI_ISL_1663383, EPI_ISL_1663384, EPI_ISL_1663385, EPI_ISL_1663386, EPI_ISL_1663387, EPI_ISL_1663388, EPI_ISL_1663389, EPI_ISL_1663390, EPI_ISL_1663391, EPI_ISL_1663392, EPI_ISL_1663393, EPI_ISL_1663394, EPI_ISL_1663395, EPI_ISL_1663396, EPI_ISL_1663397, EPI_ISL_1663398, EPI_ISL_1663399, EPI_ISL_1663400, EPI_ISL_1663401, EPI_ISL_1663402, EPI_ISL_1663403, EPI_ISL_1663405, EPI_ISL_1663406, EPI_ISL_1663407, EPI_ISL_1663408, EPI_ISL_1663409, EPI_ISL_1663410, EPI_ISL_1663411, EPI_ISL_1663412, EPI_ISL_1663413, EPI_ISL_1663414, EPI_ISL_1663415, EPI_ISL_1663416, EPI_ISL_1663417, EPI_ISL_1663418, EPI_ISL_1663419, EPI_ISL_1663420, EPI_ISL_1663421, EPI_ISL_1663422, EPI_ISL_1663423, EPI_ISL_1663424, EPI_ISL_1663425, EPI_ISL_1663426, EPI_ISL_1663427, EPI_ISL_1663428, EPI_ISL_1663429, EPI_ISL_1663430, EPI_ISL_1663431, EPI_ISL_1663432, EPI_ISL_1663433, EPI_ISL_1663434, EPI_ISL_1663435, EPI_ISL_1663436, EPI_ISL_1663437, EPI_ISL_1663438, EPI_ISL_1663439, EPI_ISL_1663440, EPI_ISL_1663441, EPI_ISL_1663442, EPI_ISL_1663443, EPI_ISL_1663444, EPI_ISL_1663445, EPI_ISL_1663446, EPI_ISL_1663447, EPI_ISL_1663448, EPI_ISL_1663449, EPI_ISL_1663450, EPI_ISL_1663451, EPI_ISL_1663452, EPI_ISL_1663453, EPI_ISL_1663454, EPI_ISL_1663455, EPI_ISL_1663456, EPI_ISL_1663457, EPI_ISL_1663458, EPI_ISL_1663459, EPI_ISL_1663460, EPI_ISL_1663461, EPI_ISL_1663462, EPI_ISL_1663463, EPI_ISL_1663464, EPI_ISL_1663465, EPI_ISL_1663466, EPI_ISL_1663467, EPI_ISL_1663468, EPI_ISL_1663469, EPI_ISL_1663470, EPI_ISL_1663471, EPI_ISL_1663472, EPI_ISL_1663473, EPI_ISL_1663474, EPI_ISL_1663475, EPI_ISL_1663476, EPI_ISL_1663477, EPI_ISL_1663478, EPI_ISL_1663479, EPI_ISL_1663480, EPI_ISL_1663481, EPI_ISL_1663482, EPI_ISL_1663483, EPI_ISL_1663484, EPI_ISL_1663485, EPI_ISL_1663486, EPI_ISL_1663487 | | | |
| see above | NCCS, Pune | Institute of Life Sciences - INSACOG | Sunil K. Raghav, Safal Walia, Arup Ghosh, Atimukta Jha, Amol M. Kanampalliwar, Shifu Aggarwal, Rupesh Dash, Rajeeb Swain, Punit Prasad, INSACOG Consortium, Ajay Parida |
| EPI_ISL_1669766, EPI_ISL_1669767, EPI_ISL_1669768, EPI_ISL_1669769, EPI_ISL_1669770, EPI_ISL_1669771, EPI_ISL_1669772, EPI_ISL_1669773, EPI_ISL_1669774, EPI_ISL_1669775, EPI_ISL_1669776, EPI_ISL_1669777, EPI_ISL_1669778 | | | |
| see above | Indian Council of Medical Research-National Institute of | Indian Council of Medical Research-National Institute of | Pragya D Yadav |

| | | | |
|---|---|---|---|
| | Virology, Microbial Containment Complex | Virology, Microbial Containment Complex | |
| EPI_ISL_1703635, EPI_ISL_1703636, EPI_ISL_1703637, EPI_ISL_1703640, EPI_ISL_1703641, EPI_ISL_1703642, EPI_ISL_1703644, EPI_ISL_1703645, EPI_ISL_1703646, EPI_ISL_1703647, EPI_ISL_1703648, EPI_ISL_1703649, EPI_ISL_1703650, EPI_ISL_1703651, EPI_ISL_1703652, EPI_ISL_1703656, EPI_ISL_1703657, EPI_ISL_1703658, EPI_ISL_1703659, EPI_ISL_1703660, EPI_ISL_1703661, EPI_ISL_1703662, EPI_ISL_1703663, EPI_ISL_1703664, EPI_ISL_1703666, EPI_ISL_1703667, EPI_ISL_1703668, EPI_ISL_1703669, EPI_ISL_1703670, EPI_ISL_1703671, EPI_ISL_1703672, EPI_ISL_1703674, EPI_ISL_1703675, EPI_ISL_1703676, EPI_ISL_1703679, EPI_ISL_1703680, EPI_ISL_1703681, EPI_ISL_1703682, EPI_ISL_1703683, EPI_ISL_1703684, EPI_ISL_1703685, EPI_ISL_1703688, EPI_ISL_1703689, EPI_ISL_1703690, EPI_ISL_1703691, EPI_ISL_1703693, EPI_ISL_1703694, EPI_ISL_1703695, EPI_ISL_1703696, EPI_ISL_1703697, EPI_ISL_1703700, EPI_ISL_1703702, EPI_ISL_1703703, EPI_ISL_1703705, EPI_ISL_1703707, EPI_ISL_1703710, EPI_ISL_1703713, EPI_ISL_1703717, EPI_ISL_1703718, EPI_ISL_1703722, EPI_ISL_1703723, EPI_ISL_1703724, EPI_ISL_1703725, EPI_ISL_1703727, EPI_ISL_1703729, EPI_ISL_1703730, EPI_ISL_1703731, EPI_ISL_1703735, EPI_ISL_1703738, EPI_ISL_1703740, EPI_ISL_1703744, EPI_ISL_1703746, EPI_ISL_1703747, EPI_ISL_1703749, EPI_ISL_1703751, EPI_ISL_1703752, EPI_ISL_1703753, EPI_ISL_1703755, EPI_ISL_1703756, EPI_ISL_1703757, EPI_ISL_1703758, EPI_ISL_1703759, EPI_ISL_1703762, EPI_ISL_1703765, EPI_ISL_1703767, EPI_ISL_1703768, EPI_ISL_1703769, EPI_ISL_1703770, EPI_ISL_1703771, EPI_ISL_1703772, EPI_ISL_1703774, EPI_ISL_1703775, EPI_ISL_1703776, EPI_ISL_1703777, EPI_ISL_1703778, EPI_ISL_1703782, EPI_ISL_1703784, EPI_ISL_1703785, EPI_ISL_1703787, EPI_ISL_1703788, EPI_ISL_1703791, EPI_ISL_1703793, EPI_ISL_1703794, EPI_ISL_1703796, EPI_ISL_1703798, EPI_ISL_1703800, EPI_ISL_1703803, EPI_ISL_1703804, EPI_ISL_1703805, EPI_ISL_1703806, EPI_ISL_1703808, EPI_ISL_1703809, EPI_ISL_1703810, EPI_ISL_1703811, EPI_ISL_1703812, EPI_ISL_1703815, EPI_ISL_1703817, EPI_ISL_1703819, EPI_ISL_1703820, EPI_ISL_1703821, EPI_ISL_1703822, EPI_ISL_1703823, EPI_ISL_1703825, EPI_ISL_1703826, EPI_ISL_1703828, EPI_ISL_1703831, EPI_ISL_1703835, EPI_ISL_1703837, EPI_ISL_1703838, EPI_ISL_1703839, EPI_ISL_1703841, EPI_ISL_1703842, EPI_ISL_1703843, EPI_ISL_1703847, EPI_ISL_1703850, EPI_ISL_1703851, EPI_ISL_1703853, EPI_ISL_1703855, EPI_ISL_1703860, EPI_ISL_1703862, EPI_ISL_1703863, EPI_ISL_1703864, EPI_ISL_1703866, EPI_ISL_1703868, EPI_ISL_1703869, EPI_ISL_1703870, EPI_ISL_1703872, EPI_ISL_1703873, EPI_ISL_1703876, EPI_ISL_1703877, EPI_ISL_1703878, EPI_ISL_1703879, EPI_ISL_1703880, EPI_ISL_1703881, EPI_ISL_1703882, EPI_ISL_1703884, EPI_ISL_1703886, EPI_ISL_1703888, EPI_ISL_1703889, EPI_ISL_1703891, EPI_ISL_1703892, EPI_ISL_1703895, EPI_ISL_1703898, EPI_ISL_1703899, EPI_ISL_1703901, EPI_ISL_1703903, EPI_ISL_1703904, EPI_ISL_1703913, EPI_ISL_1703914, EPI_ISL_1703915, EPI_ISL_1703916, EPI_ISL_1703917, EPI_ISL_1703918, EPI_ISL_1703923, EPI_ISL_1703924, EPI_ISL_1703925, EPI_ISL_1703957, EPI_ISL_1703959, EPI_ISL_1703960, EPI_ISL_1703961, EPI_ISL_1703964, EPI_ISL_1703969, EPI_ISL_1703983, EPI_ISL_1703984, EPI_ISL_1703985, EPI_ISL_1703986, EPI_ISL_1704005, EPI_ISL_1704006, EPI_ISL_1704007, EPI_ISL_1704008, EPI_ISL_1704009, EPI_ISL_1704011, EPI_ISL_1704012, EPI_ISL_1704013, EPI_ISL_1704014, EPI_ISL_1704015, EPI_ISL_1704016, EPI_ISL_1704017, EPI_ISL_1704018, EPI_ISL_1704019, EPI_ISL_1704020, EPI_ISL_1704021, EPI_ISL_1704022, EPI_ISL_1704024, EPI_ISL_1704025, EPI_ISL_1704026, EPI_ISL_1704027, EPI_ISL_1704028, EPI_ISL_1704029, EPI_ISL_1704030, EPI_ISL_1704032, EPI_ISL_1704033, EPI_ISL_1704034, EPI_ISL_1704035, EPI_ISL_1704036, EPI_ISL_1704037, EPI_ISL_1704038, EPI_ISL_1704039, EPI_ISL_1704040, EPI_ISL_1704042, EPI_ISL_1704043, EPI_ISL_1704044, EPI_ISL_1704045, EPI_ISL_1704046, EPI_ISL_1704047, EPI_ISL_1704048, EPI_ISL_1704049, EPI_ISL_1704050, EPI_ISL_1704052, EPI_ISL_1704053, EPI_ISL_1704054, EPI_ISL_1704060, EPI_ISL_1704061, EPI_ISL_1704062, EPI_ISL_1704066, EPI_ISL_1704067, EPI_ISL_1704069, EPI_ISL_1704070, EPI_ISL_1704071, EPI_ISL_1704076, EPI_ISL_1704078, EPI_ISL_1704082, EPI_ISL_1704084, EPI_ISL_1704085, EPI_ISL_1704089, EPI_ISL_1704090, EPI_ISL_1704091, EPI_ISL_1704092, EPI_ISL_1704325, EPI_ISL_1704326, EPI_ISL_1704327, EPI_ISL_1704328, EPI_ISL_1704329, EPI_ISL_1704330, EPI_ISL_1704331, EPI_ISL_1704332, EPI_ISL_1704333, EPI_ISL_1704334, EPI_ISL_1704335, EPI_ISL_1704336, EPI_ISL_1704337, EPI_ISL_1704338, EPI_ISL_1704339, EPI_ISL_1704340, EPI_ISL_1704341, EPI_ISL_1704342, EPI_ISL_1704343, EPI_ISL_1704344, EPI_ISL_1704345, EPI_ISL_1704346, EPI_ISL_1704347, EPI_ISL_1704348, EPI_ISL_1704349, EPI_ISL_1704350, EPI_ISL_1704351, EPI_ISL_1704352, EPI_ISL_1704353, EPI_ISL_1704354, EPI_ISL_1704355, EPI_ISL_1704356, EPI_ISL_1704357, EPI_ISL_1704358, EPI_ISL_1704359, EPI_ISL_1704360, EPI_ISL_1704361, EPI_ISL_1704362, EPI_ISL_1704363, EPI_ISL_1704364, EPI_ISL_1704365, EPI_ISL_1704366, EPI_ISL_1704367, EPI_ISL_1704368, EPI_ISL_1704369, EPI_ISL_1704370, EPI_ISL_1704371, EPI_ISL_1704372, EPI_ISL_1704373, EPI_ISL_1704374, EPI_ISL_1704375, EPI_ISL_1704376, EPI_ISL_1704377, EPI_ISL_1704378, EPI_ISL_1704379, EPI_ISL_1704380, EPI_ISL_1704381, EPI_ISL_1704382, EPI_ISL_1704383, EPI_ISL_1704384, EPI_ISL_1704385, EPI_ISL_1704386, EPI_ISL_1704387, EPI_ISL_1704388, EPI_ISL_1704389, EPI_ISL_1704390, EPI_ISL_1704391, EPI_ISL_1704392, EPI_ISL_1704393, EPI_ISL_1704394, EPI_ISL_1704395, EPI_ISL_1704396, EPI_ISL_1704397, EPI_ISL_1704398, EPI_ISL_1704399, EPI_ISL_1704400, EPI_ISL_1704401, EPI_ISL_1704402, EPI_ISL_1704403, EPI_ISL_1704404, EPI_ISL_1704405, EPI_ISL_1704406, EPI_ISL_1704407, EPI_ISL_1704408, EPI_ISL_1704409, EPI_ISL_1704410, EPI_ISL_1704411, EPI_ISL_1704412, EPI_ISL_1704413, EPI_ISL_1704414, EPI_ISL_1704415, EPI_ISL_1704416, EPI_ISL_1704417, EPI_ISL_1704418, EPI_ISL_1704419, EPI_ISL_1704420, EPI_ISL_1704421, EPI_ISL_1704422, EPI_ISL_1704423, EPI_ISL_1704424, EPI_ISL_1704425, EPI_ISL_1704426, EPI_ISL_1704427, EPI_ISL_1704428, EPI_ISL_1704429, EPI_ISL_1704430, EPI_ISL_1704431, EPI_ISL_1704432, EPI_ISL_1704433, EPI_ISL_1704434, EPI_ISL_1704435, EPI_ISL_1704436, EPI_ISL_1704437, EPI_ISL_1704438, EPI_ISL_1704439, EPI_ISL_1704440, EPI_ISL_1704441, EPI_ISL_1704442, EPI_ISL_1704443, EPI_ISL_1704444, EPI_ISL_1704445, EPI_ISL_1704446, EPI_ISL_1704447, EPI_ISL_1704448, EPI_ISL_1704449, EPI_ISL_1704450, EPI_ISL_1704451, EPI_ISL_1704452, EPI_ISL_1704453, EPI_ISL_1704454, EPI_ISL_1704455, EPI_ISL_1704456, EPI_ISL_1704457, EPI_ISL_1704458, EPI_ISL_1704459, EPI_ISL_1704460, EPI_ISL_1704461, EPI_ISL_1704462, EPI_ISL_1704463, EPI_ISL_1704464, EPI_ISL_1704465, EPI_ISL_1704466, EPI_ISL_1704467, EPI_ISL_1704468, EPI_ISL_1704469, EPI_ISL_1704470, EPI_ISL_1704471, EPI_ISL_1704472, EPI_ISL_1704473, EPI_ISL_1704474, EPI_ISL_1704475, EPI_ISL_1704476, EPI_ISL_1704477, EPI_ISL_1704478, EPI_ISL_1704479, EPI_ISL_1704480, EPI_ISL_1704481, EPI_ISL_1704482, EPI_ISL_1704483, EPI_ISL_1704484, EPI_ISL_1704485, EPI_ISL_1704486, EPI_ISL_1704487, EPI_ISL_1704488, EPI_ISL_1704489, EPI_ISL_1704490, EPI_ISL_1704491, EPI_ISL_1704492, EPI_ISL_1704493, EPI_ISL_1704494, EPI_ISL_1704495, EPI_ISL_1704496, EPI_ISL_1704497, EPI_ISL_1704498, EPI_ISL_1704499, EPI_ISL_1704500, EPI_ISL_1704501, EPI_ISL_1704502, EPI_ISL_1704503, EPI_ISL_1704504, EPI_ISL_1704505, EPI_ISL_1704506, EPI_ISL_1704507, EPI_ISL_1704508, EPI_ISL_1704509, EPI_ISL_1704510, EPI_ISL_1704511, EPI_ISL_1704512, EPI_ISL_1704513, EPI_ISL_1704514, EPI_ISL_1704515, EPI_ISL_1704516, EPI_ISL_1704517, EPI_ISL_1704518, EPI_ISL_1704519, EPI_ISL_1704520, EPI_ISL_1704521, EPI_ISL_1704522, EPI_ISL_1704523, EPI_ISL_1704524, EPI_ISL_1704525, EPI_ISL_1704526, EPI_ISL_1704527, EPI_ISL_1704528, EPI_ISL_1704529, EPI_ISL_1704530, EPI_ISL_1704531, EPI_ISL_1704532, EPI_ISL_1704533, EPI_ISL_1704534, EPI_ISL_1704535, EPI_ISL_1704536, EPI_ISL_1704537, EPI_ISL_1704538, EPI_ISL_1704539, EPI_ISL_1704540, EPI_ISL_1704541, EPI_ISL_1704542, EPI_ISL_1704543, EPI_ISL_1704544, EPI_ISL_1704545, EPI_ISL_1704546, EPI_ISL_1704547, EPI_ISL_1704548, EPI_ISL_1704549, EPI_ISL_1704550, EPI_ISL_1704551, EPI_ISL_1704552, EPI_ISL_1704553, EPI_ISL_1704554, EPI_ISL_1704555, EPI_ISL_1704556, EPI_ISL_1704557, EPI_ISL_1704558, EPI_ISL_1704559, EPI_ISL_1704560, EPI_ISL_1704561, EPI_ISL_1704562, EPI_ISL_1704563, EPI_ISL_1704564, EPI_ISL_1704565, EPI_ISL_1704566, EPI_ISL_1704567, EPI_ISL_1704568, EPI_ISL_1704569, EPI_ISL_1704570, EPI_ISL_1704571, EPI_ISL_1704572, EPI_ISL_1704573, EPI_ISL_1704574, EPI_ISL_1704575, EPI_ISL_1704576, EPI_ISL_1704577, EPI_ISL_1704578, EPI_ISL_1704579, EPI_ISL_1704580, EPI_ISL_1704581, EPI_ISL_1704582, EPI_ISL_1704583, EPI_ISL_1704584, EPI_ISL_1704585, EPI_ISL_1704586, EPI_ISL_1704587, EPI_ISL_1704588, EPI_ISL_1704589, EPI_ISL_1704590, EPI_ISL_1704591, EPI_ISL_1704592, EPI_ISL_1704593, EPI_ISL_1704594, EPI_ISL_1704595, EPI_ISL_1704596, EPI_ISL_1704597, EPI_ISL_1704598, EPI_ISL_1704599, EPI_ISL_1704600, EPI_ISL_1704601, EPI_ISL_1704602, EPI_ISL_1704603, EPI_ISL_1704604, EPI_ISL_1704605, EPI_ISL_1704606, EPI_ISL_1704607, EPI_ISL_1704608, EPI_ISL_1704609, EPI_ISL_1704610, EPI_ISL_1704611, EPI_ISL_1704612, EPI_ISL_1704613, EPI_ISL_1704614, EPI_ISL_1704615, EPI_ISL_1704616, EPI_ISL_1704617, EPI_ISL_1704618, EPI_ISL_1704619, EPI_ISL_1704620, EPI_ISL_1704621, EPI_ISL_1704622, EPI_ISL_1704623, EPI_ISL_1704624, EPI_ISL_1704625, EPI_ISL_1704626, EPI_ISL_1704627, EPI_ISL_1704628, EPI_ISL_1704629, EPI_ISL_1704630, EPI_ISL_1704631, EPI_ISL_1704632, EPI_ISL_1704633, EPI_ISL_1704634, EPI_ISL_1704635, EPI_ISL_1704636, EPI_ISL_1704637, EPI_ISL_1704638, EPI_ISL_1704639, EPI_ISL_1704640, EPI_ISL_1704641, EPI_ISL_1704642, EPI_ISL_1704643, EPI_ISL_1704644, EPI_ISL_1704645, EPI_ISL_1704646, EPI_ISL_1704647, EPI_ISL_1704648, EPI_ISL_1704649, EPI_ISL_1704650, EPI_ISL_1704651, EPI_ISL_1704652, EPI_ISL_1704653, EPI_ISL_1704654, EPI_ISL_1704655, EPI_ISL_1704656, EPI_ISL_1704657 | | | |
| see above | ICMR-National Institute of Virology - INSACOG | NIV Influenza | Dr. Varsha Potdar |
| EPI_ISL_1710598 | Communicable Diseases, Interactive Research School for Health Affairs | Communicable Diseases, Interactive Research School for Health Affairs | Shubham Shrivastava, Meera Modak, Rashmi Virkar, Shamburaje S Pisal, Akhilesh Chandra Mishra, Vidya A Arankalle |
| EPI_ISL_1718261 | CSIR-National Environmental Engineering Research Institute | CSIR-Centre for Cellular and Molecular Biology - INSACOG | Onkar Kulkarni, Lamuk Zaveri, Ara Sreenivas, Sofia Banu, Shreekant Verma, Amareshwar Vodapalli , Viswagithe S L, B Himasri, Sharath Chandra Thota, Karthik Bharadwaj Tallapaka, Krishna Khairnar, Rakesh K Mishra,Divya Tej Sowpati |
| EPI_ISL_1761637, EPI_ISL_1761638 | Virus Research and Diagnostic Laboratory, Medical College Level, Kasturba Hospital for Infectious Diseases | National Institute of Virology-Microbial Containment Complex, Indian Council of Medical Research | Pragya D. Yadav, Jayanti Shastri |
| EPI_ISL_1837988, EPI_ISL_1837991, EPI_ISL_1837993, EPI_ISL_1837996, EPI_ISL_1837998, EPI_ISL_1838001, EPI_ISL_1838004, EPI_ISL_1838007, EPI_ISL_1838010, EPI_ISL_1838012, EPI_ISL_1838015, EPI_ISL_1838018, EPI_ISL_1838021, EPI_ISL_1838023, EPI_ISL_1838026, EPI_ISL_1838029, EPI_ISL_1838032, EPI_ISL_1838035, EPI_ISL_1838037, EPI_ISL_1838040, EPI_ISL_1838042, EPI_ISL_1838045, EPI_ISL_1838047, EPI_ISL_1838050, EPI_ISL_1838052, EPI_ISL_1838055, EPI_ISL_1838057, EPI_ISL_1838060, EPI_ISL_1838063, EPI_ISL_1838065, EPI_ISL_1838068, EPI_ISL_1838070, EPI_ISL_1838073, EPI_ISL_1838075, EPI_ISL_1838078, EPI_ISL_1838080, EPI_ISL_1838083, EPI_ISL_1838086, EPI_ISL_1838088, EPI_ISL_1838091, EPI_ISL_1838094, EPI_ISL_1838097, EPI_ISL_1838100, EPI_ISL_1838102, EPI_ISL_1838106, EPI_ISL_1838109, EPI_ISL_1838111, EPI_ISL_1838114, EPI_ISL_1838117, EPI_ISL_1838119, EPI_ISL_1838122, EPI_ISL_1838123, EPI_ISL_1838125, EPI_ISL_1838126, EPI_ISL_1838128, EPI_ISL_1838129, EPI_ISL_1838131, EPI_ISL_1838132, EPI_ISL_1838134, EPI_ISL_1838136, EPI_ISL_1838139, EPI_ISL_1838140, EPI_ISL_1838142, EPI_ISL_1838143, EPI_ISL_1838145, EPI_ISL_1838148, EPI_ISL_1838149, EPI_ISL_1838151, EPI_ISL_1838152, EPI_ISL_1838153, EPI_ISL_1838155, EPI_ISL_1838156, EPI_ISL_1838158, EPI_ISL_1838159, EPI_ISL_1838161, EPI_ISL_1838162, EPI_ISL_1838164, EPI_ISL_1838165, EPI_ISL_1838166, EPI_ISL_1838168, EPI_ISL_1838169, EPI_ISL_1838171, EPI_ISL_1838172, EPI_ISL_1838173, EPI_ISL_1838175, EPI_ISL_1838176, EPI_ISL_1838178, EPI_ISL_1838179, EPI_ISL_1838182, EPI_ISL_1838185, EPI_ISL_1838186, EPI_ISL_1838188, EPI_ISL_1838189, EPI_ISL_1838191, EPI_ISL_1838194, EPI_ISL_1838195, EPI_ISL_1838196, EPI_ISL_1838198, EPI_ISL_1838200, EPI_ISL_1838201, EPI_ISL_1838203, EPI_ISL_1838205, EPI_ISL_1838207, EPI_ISL_1838209, EPI_ISL_1838210, EPI_ISL_1838212, EPI_ISL_1838214, EPI_ISL_1838215, EPI_ISL_1838217, EPI_ISL_1838219, EPI_ISL_1838220, EPI_ISL_1838222, EPI_ISL_1838224, EPI_ISL_1838225, EPI_ISL_1838227, EPI_ISL_1838229, EPI_ISL_1838230, EPI_ISL_1838232, EPI_ISL_1838234, EPI_ISL_1838235, EPI_ISL_1838237, EPI_ISL_1838239, EPI_ISL_1838241, EPI_ISL_1838242, EPI_ISL_1838244, EPI_ISL_1838246, EPI_ISL_1838248, EPI_ISL_1838250, EPI_ISL_1838251, EPI_ISL_1838253, EPI_ISL_1838254, EPI_ISL_1838256, EPI_ISL_1838257, EPI_ISL_1838259, EPI_ISL_1838260, EPI_ISL_1838262, EPI_ISL_1838264, EPI_ISL_1838265, EPI_ISL_1838267, EPI_ISL_1838269, EPI_ISL_1838271, EPI_ISL_1838272, EPI_ISL_1838274, EPI_ISL_1838275, EPI_ISL_1838277, EPI_ISL_1838278, EPI_ISL_1838280, EPI_ISL_1838282, EPI_ISL_1838284, EPI_ISL_1838285, EPI_ISL_1838287, EPI_ISL_1838288, EPI_ISL_1838290, EPI_ISL_1838292, EPI_ISL_1838293, EPI_ISL_1838295, EPI_ISL_1838296, EPI_ISL_1838298, EPI_ISL_1838299, EPI_ISL_1838301, EPI_ISL_1838303, EPI_ISL_1838304, EPI_ISL_1838306, EPI_ISL_1838307, EPI_ISL_1838309, EPI_ISL_1838311, EPI_ISL_1838312, EPI_ISL_1838314, EPI_ISL_1838315, EPI_ISL_1838317, EPI_ISL_1838318, EPI_ISL_1838320, EPI_ISL_1838322, EPI_ISL_1838323, EPI_ISL_1838325, EPI_ISL_1838327, EPI_ISL_1838328, EPI_ISL_1838330, EPI_ISL_1838332, EPI_ISL_1838333, EPI_ISL_1838335, EPI_ISL_1838337, EPI_ISL_1838338, EPI_ISL_1838340, EPI_ISL_1838342, EPI_ISL_1838343, EPI_ISL_1838345, EPI_ISL_1838346 | | | |
| see above | The National Centre for Cell Science | CSIR-Centre for Cellular and Molecular Biology-INSACOG | Dhiraj Paul, Mohak P Gujare, Shivang P. Bhanushali, Mitali Inamdar, Sonal Manik Chavan, Manoj Kumar Bhat, Ajay Pillai, INSACOG Consortium team, Yogesh Shouche,Payel Mukherjee,Lamuk Zaveri,Tulasi Nagabandi, Ara Sreenivas,Valli Nagalakshmi Undamatla,Shreekant Verma, Amareshwar Vodapalli ,Blessy B John,Viswagithe S L,B Himasri,Onkar Kulkarni,Sofia Banu,Archana Bharadwaj Siva,Sharath Chandra Thota,Karthik Bharadwaj Tallapaka,Rakesh K Mishra,Divya Tej Sowpati |
| EPI_ISL_1841286, EPI_ISL_1841287, EPI_ISL_1841288, EPI_ISL_1841289, EPI_ISL_1841290, EPI_ISL_1841291, EPI_ISL_1841292, EPI_ISL_1841293, EPI_ISL_1841294, EPI_ISL_1841295, EPI_ISL_1841296, EPI_ISL_1841297, EPI_ISL_1841298, EPI_ISL_1841299, EPI_ISL_1841300, EPI_ISL_1841301, EPI_ISL_1841302, EPI_ISL_1841303, EPI_ISL_1841304, EPI_ISL_1841305, EPI_ISL_1841306, EPI_ISL_1841307, EPI_ISL_1841308, EPI_ISL_1841309, EPI_ISL_1841310, EPI_ISL_1841311, EPI_ISL_1841312, EPI_ISL_1841313, EPI_ISL_1841314, EPI_ISL_1841315, EPI_ISL_1841316, EPI_ISL_1841317, EPI_ISL_1841318, EPI_ISL_1841319, EPI_ISL_1841320, EPI_ISL_1841321, EPI_ISL_1841322, EPI_ISL_1841323, EPI_ISL_1841324, EPI_ISL_1841325, EPI_ISL_1841326, EPI_ISL_1841327, EPI_ISL_1841328, EPI_ISL_1841329, EPI_ISL_1841330, EPI_ISL_1841331, EPI_ISL_1841332, EPI_ISL_1841333, EPI_ISL_1841334, EPI_ISL_1841335, EPI_ISL_1841336, EPI_ISL_1841337, EPI_ISL_1841338, EPI_ISL_1841339, EPI_ISL_1841340, EPI_ISL_1841341, EPI_ISL_1841342, EPI_ISL_1841343, EPI_ISL_1841344, EPI_ISL_1841345, EPI_ISL_1841346, EPI_ISL_1841347, EPI_ISL_1841348, EPI_ISL_1841349, EPI_ISL_1841350, EPI_ISL_1841351, EPI_ISL_1841352, EPI_ISL_1841353, EPI_ISL_1841354, EPI_ISL_1841355, EPI_ISL_1841356, EPI_ISL_1841357, EPI_ISL_1841358, EPI_ISL_1841359, EPI_ISL_1841360, EPI_ISL_1841361, EPI_ISL_1841362, EPI_ISL_1841363, EPI_ISL_1841364, EPI_ISL_1841365, EPI_ISL_1841366, EPI_ISL_1841367, EPI_ISL_1841368, EPI_ISL_1841369, EPI_ISL_1841370, EPI_ISL_1841371, EPI_ISL_1841372, EPI_ISL_1841373, EPI_ISL_1841374, EPI_ISL_1841375, EPI_ISL_1841376, EPI_ISL_1841377, EPI_ISL_1841378, EPI_ISL_1841379, EPI_ISL_1841380, EPI_ISL_1841381, EPI_ISL_1841382, EPI_ISL_1841383 | | | |
| see above | ICMR-National Institute of Virology - INSACOG | NIV Influenza | Dr. Varsha Potdar |
| EPI_ISL_1916266, EPI_ISL_1916267, EPI_ISL_1916268, EPI_ISL_1916269, EPI_ISL_1916270, EPI_ISL_1916271, EPI_ISL_1916272, EPI_ISL_1916273, EPI_ISL_1916274, EPI_ISL_1916275, EPI_ISL_1916276, EPI_ISL_1916277, EPI_ISL_1916278, EPI_ISL_1916279, EPI_ISL_1916280, EPI_ISL_1916281, EPI_ISL_1916282, EPI_ISL_1916283, EPI_ISL_1916284, EPI_ISL_1916285, EPI_ISL_1916286, EPI_ISL_1916287, EPI_ISL_1916288, EPI_ISL_1916289, EPI_ISL_1916290, EPI_ISL_1916291, EPI_ISL_1916292, EPI_ISL_1916293, EPI_ISL_1916294, EPI_ISL_1916295, EPI_ISL_1916296, EPI_ISL_1916297, EPI_ISL_1916298, EPI_ISL_1916299, EPI_ISL_1916300, EPI_ISL_1916301, EPI_ISL_1916302, EPI_ISL_1916303, EPI_ISL_1916304, EPI_ISL_1916305, EPI_ISL_1916306, EPI_ISL_1916307, EPI_ISL_1916308, EPI_ISL_1916309, EPI_ISL_1916310 | | | |
| see above | CSIR-National Environmental Engineering Research Institute | CSIR-Centre for Cellular and Molecular Biology - INSACOG | Sofia Banu, Onkar Kulkarni, Lamuk Zaveri, Ara Sreenivas, Shreekant Verma, Amareshwar Vodapalli , Viswagithe S L, B Himasri, Sharath Chandra Thota, Karthik Bharadwaj Tallapaka, Krishna Khairnar, Rakesh K Mishra,Divya Tej Sowpati |
| EPI_ISL_1916311, EPI_ISL_1916312, EPI_ISL_1916313, EPI_ISL_1916361, EPI_ISL_1916362, EPI_ISL_1916363, EPI_ISL_1916364, EPI_ISL_1916365, EPI_ISL_1916366, EPI_ISL_1916367, EPI_ISL_1916368, EPI_ISL_1916369, EPI_ISL_1916370, EPI_ISL_1916371, EPI_ISL_1916372, EPI_ISL_1916373, EPI_ISL_1916374, EPI_ISL_1916375, EPI_ISL_1916376, EPI_ISL_1916377, EPI_ISL_1916378, EPI_ISL_1916379, EPI_ISL_1916380, EPI_ISL_1916381, EPI_ISL_1916382, EPI_ISL_1916383, EPI_ISL_1916384, EPI_ISL_1916385, EPI_ISL_1916386, EPI_ISL_1916387, EPI_ISL_1916388, EPI_ISL_1916389, EPI_ISL_1916390, EPI_ISL_1916391, EPI_ISL_1916392, EPI_ISL_1916393, EPI_ISL_1916394, EPI_ISL_1916395 | | | |
| see above | CSIR-National Environmental Engineering Research Institute | CSIR-Centre for Cellular and Molecular Biology - INSACOG | Lamuk Zaveri, Sofia Banu, Onkar Kulkarni, Ara Sreenivas, Shreekant Verma, Amareshwar Vodapalli , Viswagithe S L, B Himasri, Sharath Chandra Thota, Karthik Bharadwaj Tallapaka, Krishna Khairnar, Rakesh K Mishra,Divya Tej Sowpati |
| EPI_ISL_1916396, EPI_ISL_1916397, EPI_ISL_1916398, EPI_ISL_1916399, EPI_ISL_1916400, EPI_ISL_1916401, EPI_ISL_1916402, EPI_ISL_1916403, EPI_ISL_1916404, EPI_ISL_1916405, EPI_ISL_1916406, EPI_ISL_1916407, EPI_ISL_1916408, EPI_ISL_1916409, EPI_ISL_1916410, EPI_ISL_1916411, EPI_ISL_1916412, EPI_ISL_1916413, EPI_ISL_1916414, EPI_ISL_1916415, EPI_ISL_1916416, EPI_ISL_1916417, EPI_ISL_1916418, EPI_ISL_1916419, EPI_ISL_1916420, EPI_ISL_1916421, EPI_ISL_1916422, EPI_ISL_1916423, EPI_ISL_1916424 | | | |
| see above | CSIR-National Environmental Engineering Research Institute | CSIR-Centre for Cellular and Molecular Biology - INSACOG | Onkar Kulkarni, Lamuk Zaveri, Ara Sreenivas, Sofia Banu, Shreekant Verma, Amareshwar Vodapalli , Viswagithe S L, B Himasri, Sharath Chandra Thota, Karthik Bharadwaj Tallapaka, Krishna Khairnar, Rakesh K Mishra,Divya Tej Sowpati |

| | | | |
|---|---|---|---|
| EPI_ISL_1916425, EPI_ISL_1916426, EPI_ISL_1916427, EPI_ISL_1916428, EPI_ISL_1916429, EPI_ISL_1916430, EPI_ISL_1916431, EPI_ISL_1916432, EPI_ISL_1916433, EPI_ISL_1916434, EPI_ISL_1916435, EPI_ISL_1916436, EPI_ISL_1916437, EPI_ISL_1916438, EPI_ISL_1916439, EPI_ISL_1916440, EPI_ISL_1916441, EPI_ISL_1916442, EPI_ISL_1916443, EPI_ISL_1916444, EPI_ISL_1916445, EPI_ISL_1916446, EPI_ISL_1916447, EPI_ISL_1916448, EPI_ISL_1916449, EPI_ISL_1916450, EPI_ISL_1916451, EPI_ISL_1916452, EPI_ISL_1916453, EPI_ISL_1916454, EPI_ISL_1916455, EPI_ISL_1916456, EPI_ISL_1916457, EPI_ISL_1916458, EPI_ISL_1916459, EPI_ISL_1916460, EPI_ISL_1916461, EPI_ISL_1916462, EPI_ISL_1916463, EPI_ISL_1916464, EPI_ISL_1916465, EPI_ISL_1916466, EPI_ISL_1916467, EPI_ISL_1916468, EPI_ISL_1916469, EPI_ISL_1916470, EPI_ISL_1916471, EPI_ISL_1916472, EPI_ISL_1916473, EPI_ISL_1916474, EPI_ISL_1916475, EPI_ISL_1916476, EPI_ISL_1916477, EPI_ISL_1916478, EPI_ISL_1916479, EPI_ISL_1916480, EPI_ISL_1916481, EPI_ISL_1916482, EPI_ISL_1916483, EPI_ISL_1916484, EPI_ISL_1916485, EPI_ISL_1916486, EPI_ISL_1916487, EPI_ISL_1916488, EPI_ISL_1916489, EPI_ISL_1916490, EPI_ISL_1916491, EPI_ISL_1916492, EPI_ISL_1916493, EPI_ISL_1916494, EPI_ISL_1916495, EPI_ISL_1916496, EPI_ISL_1916497, EPI_ISL_1916498 | | | |
| see above | CSIR-National Environmental Engineering Research Institute | CSIR-Centre for Cellular and Molecular Biology - INSACOG | Ara Sreenivas, Onkar Kulkarni, Lamuk Zaveri, Sofia Banu, Shreekant Verma, Amareshwar Vodapalli , Viswagithe S L, B Himasri, Sharath Chandra Thota, Karthik Bharadwaj Tallapaka, Krishna Khairnar, Rakesh K Mishra,Divya Tej Sowpati |
| EPI_ISL_1928385, EPI_ISL_1928387, EPI_ISL_1928388, EPI_ISL_1928389, EPI_ISL_1928391, EPI_ISL_1928392, EPI_ISL_1928394, EPI_ISL_1928395, EPI_ISL_1928396, EPI_ISL_1928398, EPI_ISL_1928399, EPI_ISL_1928401, EPI_ISL_1928402, EPI_ISL_1928404, EPI_ISL_1928405, EPI_ISL_1928407, EPI_ISL_1928408, EPI_ISL_1928410, EPI_ISL_1928411, EPI_ISL_1928413, EPI_ISL_1928414, EPI_ISL_1928416, EPI_ISL_1928417, EPI_ISL_1928419, EPI_ISL_1928420, EPI_ISL_1928422, EPI_ISL_1928423, EPI_ISL_1928425, EPI_ISL_1928426, EPI_ISL_1928427, EPI_ISL_1928429, EPI_ISL_1928430, EPI_ISL_1928432, EPI_ISL_1928433, EPI_ISL_1928435, EPI_ISL_1928436, EPI_ISL_1928438, EPI_ISL_1928439, EPI_ISL_1928441, EPI_ISL_1928442, EPI_ISL_1928444, EPI_ISL_1928445, EPI_ISL_1928447, EPI_ISL_1928448, EPI_ISL_1928450, EPI_ISL_1928451, EPI_ISL_1928453, EPI_ISL_1928454, EPI_ISL_1928456, EPI_ISL_1928457, EPI_ISL_1928459, EPI_ISL_1928460, EPI_ISL_1928462, EPI_ISL_1928463, EPI_ISL_1928464, EPI_ISL_1928465, EPI_ISL_1928466, EPI_ISL_1928467, EPI_ISL_1928468, EPI_ISL_1928469, EPI_ISL_1928471, EPI_ISL_1928472, EPI_ISL_1928474, EPI_ISL_1928475, EPI_ISL_1928477, EPI_ISL_1928478, EPI_ISL_1928480, EPI_ISL_1928481, EPI_ISL_1928483, EPI_ISL_1928484, EPI_ISL_1928485, EPI_ISL_1928487, EPI_ISL_1928488, EPI_ISL_1928490, EPI_ISL_1928491, EPI_ISL_1928493, EPI_ISL_1928494, EPI_ISL_1928496, EPI_ISL_1928497, EPI_ISL_1928499, EPI_ISL_1928500, EPI_ISL_1928502, EPI_ISL_1928503, EPI_ISL_1928505, EPI_ISL_1928518, EPI_ISL_1928520, EPI_ISL_1928521, EPI_ISL_1928522, EPI_ISL_1928524, EPI_ISL_1928525, EPI_ISL_1928527, EPI_ISL_1928539, EPI_ISL_1928540, EPI_ISL_1928541, EPI_ISL_1928579, EPI_ISL_1928581, EPI_ISL_1928583, EPI_ISL_1928584, EPI_ISL_1928586, EPI_ISL_1928593, EPI_ISL_1928595, EPI_ISL_1928596, EPI_ISL_1928598, EPI_ISL_1928608, EPI_ISL_1928610, EPI_ISL_1928611, EPI_ISL_1928613, EPI_ISL_1928614, EPI_ISL_1928615, EPI_ISL_1928617, EPI_ISL_1928618, EPI_ISL_1928620, EPI_ISL_1928622, EPI_ISL_1928623, EPI_ISL_1928624, EPI_ISL_1928626, EPI_ISL_1928633, EPI_ISL_1928635, EPI_ISL_1928636, EPI_ISL_1928642, EPI_ISL_1928644, EPI_ISL_1928645, EPI_ISL_1928647, EPI_ISL_1928648, EPI_ISL_1928650 | | | |
| see above | ICMR-National Institute of Virology - INSACOG | NIV Influenza | Dr. Varsha Potdar |
| EPI_ISL_1939875, EPI_ISL_1939876, EPI_ISL_1939877, EPI_ISL_1939878, EPI_ISL_1939880, EPI_ISL_1939881, EPI_ISL_1939882, EPI_ISL_1939883, EPI_ISL_1939886, EPI_ISL_1939887, EPI_ISL_1939888, EPI_ISL_1939889, EPI_ISL_1939890, EPI_ISL_1939891, EPI_ISL_1939892, EPI_ISL_1939893, EPI_ISL_1939894, EPI_ISL_1939895, EPI_ISL_1939896, EPI_ISL_1939897, EPI_ISL_1939898, EPI_ISL_1939899, EPI_ISL_1939900, EPI_ISL_1939901, EPI_ISL_1939902, EPI_ISL_1939903, EPI_ISL_1939904, EPI_ISL_1939905, EPI_ISL_1939906, EPI_ISL_1939907, EPI_ISL_1939908, EPI_ISL_1939909, EPI_ISL_1939910, EPI_ISL_1939911, EPI_ISL_1939912, EPI_ISL_1939913, EPI_ISL_1939914, EPI_ISL_1939915, EPI_ISL_1939916, EPI_ISL_1939917, EPI_ISL_1939918, EPI_ISL_1939920, EPI_ISL_1939921, EPI_ISL_1939922, EPI_ISL_1939923, EPI_ISL_1939924, EPI_ISL_1939925, EPI_ISL_1939926, EPI_ISL_1939927, EPI_ISL_1939928 | | | |
| see above | NCCS | inStem NCBS | Uma Ramakrishnan Dasaradhi Palakodeti Aswin SaiNarain |
| EPI_ISL_1970419, EPI_ISL_1970420, EPI_ISL_1970421, EPI_ISL_1970422, EPI_ISL_1970423, EPI_ISL_1970424, EPI_ISL_1970425, EPI_ISL_1970426, EPI_ISL_1970427, EPI_ISL_1970428, EPI_ISL_1970429, EPI_ISL_1970430, EPI_ISL_1970431, EPI_ISL_1970432, EPI_ISL_1970433, EPI_ISL_1970434, EPI_ISL_1970435, EPI_ISL_1970436, EPI_ISL_1970437, EPI_ISL_1970438, EPI_ISL_1970439, EPI_ISL_1970440, EPI_ISL_1970441, EPI_ISL_1970442, EPI_ISL_1970443, EPI_ISL_1970444, EPI_ISL_1970445, EPI_ISL_1970446, EPI_ISL_1970447, EPI_ISL_1970448, EPI_ISL_1970449, EPI_ISL_1970450, EPI_ISL_1970451, EPI_ISL_1970452, EPI_ISL_1970513, EPI_ISL_1970514, EPI_ISL_1970515, EPI_ISL_1970516, EPI_ISL_1970517, EPI_ISL_1970518, EPI_ISL_1970519, EPI_ISL_1970520, EPI_ISL_1970521, EPI_ISL_1970522, EPI_ISL_1970523, EPI_ISL_1970524, EPI_ISL_1970525, EPI_ISL_1970526, EPI_ISL_1970527, EPI_ISL_1970528, EPI_ISL_1970529, EPI_ISL_1970530, EPI_ISL_1970531, EPI_ISL_1970532, EPI_ISL_1970533, EPI_ISL_1970534, EPI_ISL_1970535, EPI_ISL_1970536, EPI_ISL_1970537, EPI_ISL_1970538, EPI_ISL_1970539, EPI_ISL_1970540, EPI_ISL_1970541, EPI_ISL_1970542, EPI_ISL_1970543, EPI_ISL_1970544, EPI_ISL_1970545, EPI_ISL_1970546 | | | |
| see above | ICMR-National Institute of Virology - INSACOG | NIV Influenza | Dr. Varsha Potdar |
| EPI_ISL_1970625, EPI_ISL_1970626 | CSIR-National Environmental Engineering Research Institute | CSIR-Centre for Cellular and Molecular Biology - INSACOG | Sofia Banu, Onkar Kulkarni, Lamuk Zaveri, Ara Sreenivas, Shreekant Verma, Amareshwar Vodapalli , Viswagithe S L, B Himasri, Sharath Chandra Thota, Karthik Bharadwaj Tallapaka, Krishna Khairnar, Rakesh K Mishra,Divya Tej Sowpati |
| EPI_ISL_1970627 | CSIR-National Environmental Engineering Research Institute | CSIR-Centre for Cellular and Molecular Biology - INSACOG | Lamuk Zaveri, Sofia Banu, Onkar Kulkarni, Ara Sreenivas, Shreekant Verma, Amareshwar Vodapalli , Viswagithe S L, B Himasri, Sharath Chandra Thota, Karthik Bharadwaj Tallapaka, Krishna Khairnar, Rakesh K Mishra,Divya Tej Sowpati |
| EPI_ISL_1970628, EPI_ISL_1970629 | CSIR-National Environmental Engineering Research Institute | CSIR-Centre for Cellular and Molecular Biology - INSACOG | Ara Sreenivas, Onkar Kulkarni, Lamuk Zaveri, Sofia Banu, Shreekant Verma, Amareshwar Vodapalli , Viswagithe S L, B Himasri, Sharath Chandra Thota, Karthik Bharadwaj Tallapaka, Krishna Khairnar, Rakesh K Mishra,Divya Tej Sowpati |
| EPI_ISL_1972346, EPI_ISL_1972348, EPI_ISL_1972350 | The National Centre for Cell Science | CSIR-Centre for Cellular and Molecular Biology-INSACOG | Dhiraj Paul, Mohak P Gujare, Shivang P. Bhanushali, Mitali Inamdar, Sonal Manik Chavan, Manoj Kumar Bhat, Ajay Pillai, INSACOG Consortium team, Yogesh Shouche,Payel Mukherjee,Lamuk Zaveri,Tulasi Nagabandi, Ara Sreenivas,Valli Nagalakshmi Undamatla,Shreekant Verma, Amareshwar Vodapalli ,Blessy B John,Viswagithe S L,B Himasri,Onkar Kulkarni,Sofia Banu,Archana Bharadwaj Siva,Sharath Chandra Thota,Karthik Bharadwaj Tallapaka,Rakesh K Mishra,Divya Tej Sowpati |
| EPI_ISL_2013028, EPI_ISL_2013029, EPI_ISL_2013030, EPI_ISL_2013031 | Indian Council of Medical Research-National Institute of Virology, Microbial Containment Complex | Indian Council of Medical Research-National Institute of Virology, Microbial Containment Complex | Pragya D Yadav |
| EPI_ISL_2023383 | The Foundation for Medical Research | The Foundation for Medical Research | Ayan Mandal, Kayzad Nilgiriwala, Ambreen Shaikh, Grishma Patel, Smriti Vaswani, Swapneil Parikh, Nerges Mistry |
| EPI_ISL_2032336, EPI_ISL_2032338, EPI_ISL_2036287, EPI_ISL_2036288, EPI_ISL_2036289, EPI_ISL_2036290, EPI_ISL_2036291, EPI_ISL_2036292, EPI_ISL_2036293 | Indian Council of Medical Research-National Institute of Virology, Microbial Containment Complex | Indian Council of Medical Research-National Institute of Virology, Microbial Containment Complex | Pragya D Yadav |
| EPI_ISL_2099770 | Indian Council of Medical Research-National Institute of Virology, Microbial Containment Complex | Indian Council of Medical Research-National Institute of Virology, Microbial Containment Complex | Pragya D Yadav |
| EPI_ISL_2107008, EPI_ISL_2107009, EPI_ISL_2107010, EPI_ISL_2107011, EPI_ISL_2107012, EPI_ISL_2107013, EPI_ISL_2107014, EPI_ISL_2107015, EPI_ISL_2107016, EPI_ISL_2107017, EPI_ISL_2107018, EPI_ISL_2107019, EPI_ISL_2107020, EPI_ISL_2107021, EPI_ISL_2107022, EPI_ISL_2107023, EPI_ISL_2107024, EPI_ISL_2107025, EPI_ISL_2107026, EPI_ISL_2107027, EPI_ISL_2107028, EPI_ISL_2107029, EPI_ISL_2107030, EPI_ISL_2107031, EPI_ISL_2107032, EPI_ISL_2107033, EPI_ISL_2107034, EPI_ISL_2107035, EPI_ISL_2107036, EPI_ISL_2107037, EPI_ISL_2107038, EPI_ISL_2107039, EPI_ISL_2107040, EPI_ISL_2107041, EPI_ISL_2107042, EPI_ISL_2107043, EPI_ISL_2107044, EPI_ISL_2107045, EPI_ISL_2107046, EPI_ISL_2107047, EPI_ISL_2107048, EPI_ISL_2107049, EPI_ISL_2107050, EPI_ISL_2107051, EPI_ISL_2107052, EPI_ISL_2107053, EPI_ISL_2107054, EPI_ISL_2107055, EPI_ISL_2107056, EPI_ISL_2107057, EPI_ISL_2107058, EPI_ISL_2107059, EPI_ISL_2107060, EPI_ISL_2107061, EPI_ISL_2107062, EPI_ISL_2107063, EPI_ISL_2107064, EPI_ISL_2107065, EPI_ISL_2107066, EPI_ISL_2107067, EPI_ISL_2107068, EPI_ISL_2107069, EPI_ISL_2107070, EPI_ISL_2107071, EPI_ISL_2107072, EPI_ISL_2107073, EPI_ISL_2107074, EPI_ISL_2107075, EPI_ISL_2107076, EPI_ISL_2107077, EPI_ISL_2107078, EPI_ISL_2107079, EPI_ISL_2107080, EPI_ISL_2107081, EPI_ISL_2107082, EPI_ISL_2107083, EPI_ISL_2107084, EPI_ISL_2107085, EPI_ISL_2107086, EPI_ISL_2107087, EPI_ISL_2107088, EPI_ISL_2107089, EPI_ISL_2107090, EPI_ISL_2107091, EPI_ISL_2107092 | | | |
| see above | National Centre For Cell Science - INSACOG | National Centre For Cell Science | Dhiraj Paul, Shivang P. Bhanushali, Mohak P Gujare, Mitali Inamdar, Manoj Kumar Bhat, Ajay Pillai, INSACOG Consortium team, Yogesh Shouche |
| EPI_ISL_2131509, EPI_ISL_2131510, EPI_ISL_2131511, EPI_ISL_2131512, EPI_ISL_2131513, EPI_ISL_2131514, EPI_ISL_2131515, EPI_ISL_2131516, EPI_ISL_2131517, EPI_ISL_2131518, EPI_ISL_2131519, EPI_ISL_2131520, EPI_ISL_2131521, EPI_ISL_2131522, EPI_ISL_2131523, EPI_ISL_2131524, EPI_ISL_2131525, EPI_ISL_2131526, EPI_ISL_2131527, EPI_ISL_2131528, EPI_ISL_2131529, EPI_ISL_2131530, EPI_ISL_2131531, EPI_ISL_2131532, EPI_ISL_2131533, EPI_ISL_2131534, EPI_ISL_2131547, EPI_ISL_2131548, EPI_ISL_2131549, EPI_ISL_2131550, EPI_ISL_2131551, EPI_ISL_2131552, EPI_ISL_2131553, EPI_ISL_2131554, EPI_ISL_2131555, EPI_ISL_2131556, EPI_ISL_2131557, EPI_ISL_2131558, EPI_ISL_2131559, EPI_ISL_2131560, EPI_ISL_2131561, EPI_ISL_2131562, EPI_ISL_2131563, EPI_ISL_2131564, EPI_ISL_2131565, EPI_ISL_2131566, EPI_ISL_2131567, EPI_ISL_2131568, EPI_ISL_2131569, EPI_ISL_2131570, EPI_ISL_2131571, EPI_ISL_2131572 | | | |
| see above | ICMR-National Institute of Virology - INSACOG | NIV Influenza | Dr. Varsha Potdar |
| EPI_ISL_2151097 | INSACOG-Sikkim | National Institute of Biomedical Genomics - INSACOG | Arindam Maitra, Tara D Sharma, Nidhan Kumar Biswas, Kaden Zangmu Bhutia, Sreedhar Chinnaswamy Tshering Ongmu Bhutia, Saumitra Das |
| EPI_ISL_2162386, EPI_ISL_2162387, EPI_ISL_2162388, EPI_ISL_2162389, EPI_ISL_2162390, EPI_ISL_2162391, EPI_ISL_2162392, EPI_ISL_2162393, EPI_ISL_2162394, EPI_ISL_2162395, EPI_ISL_2162396, EPI_ISL_2162397, EPI_ISL_2162398, EPI_ISL_2162399, EPI_ISL_2162400, EPI_ISL_2162401, EPI_ISL_2162402, EPI_ISL_2162403, EPI_ISL_2162404, EPI_ISL_2162405, EPI_ISL_2162406, EPI_ISL_2162407, EPI_ISL_2162408, EPI_ISL_2162409, EPI_ISL_2162410, EPI_ISL_2162411, EPI_ISL_2162412, EPI_ISL_2162413, EPI_ISL_2162414, EPI_ISL_2162415, EPI_ISL_2162416, EPI_ISL_2162417, EPI_ISL_2162418, EPI_ISL_2162419, EPI_ISL_2162420, EPI_ISL_2162421, EPI_ISL_2162422, EPI_ISL_2162423, EPI_ISL_2162424, EPI_ISL_2162425, EPI_ISL_2162426, EPI_ISL_2162427, EPI_ISL_2162428, EPI_ISL_2162429, EPI_ISL_2162430, EPI_ISL_2162431, EPI_ISL_2162432, EPI_ISL_2162433, EPI_ISL_2162434, EPI_ISL_2162435, EPI_ISL_2162436, EPI_ISL_2162437, EPI_ISL_2162438, EPI_ISL_2162439, EPI_ISL_2162440, EPI_ISL_2162441, EPI_ISL_2162442, EPI_ISL_2162443, EPI_ISL_2162444, EPI_ISL_2162445, EPI_ISL_2162446, EPI_ISL_2162447, EPI_ISL_2162448, EPI_ISL_2162449, EPI_ISL_2162450, EPI_ISL_2162451, EPI_ISL_2162452, EPI_ISL_2162453, EPI_ISL_2162454, EPI_ISL_2162455, EPI_ISL_2162456, EPI_ISL_2162457, EPI_ISL_2162458, EPI_ISL_2162459, EPI_ISL_2162460, EPI_ISL_2162461, EPI_ISL_2162462, EPI_ISL_2162463, EPI_ISL_2162464, EPI_ISL_2162465, EPI_ISL_2162466, EPI_ISL_2162467 | | | |
| see above | National Centre For Cell Science | National Centre For Cell Science - INSACOG | Dhiraj Paul, Mohak P Gujare, Mitali Inamdar, Shivang P. Bhanushali, Manoj Kumar Bhat, Ajay Pillai, INSACOG Consortium team, Yogesh Shouche |
| EPI_ISL_2189714, EPI_ISL_2189715, EPI_ISL_2189716, EPI_ISL_2189719, EPI_ISL_2189720, EPI_ISL_2189721, EPI_ISL_2189722, EPI_ISL_2189723, EPI_ISL_2189725, EPI_ISL_2189726, EPI_ISL_2189727, EPI_ISL_2189728, EPI_ISL_2189729, EPI_ISL_2189730, EPI_ISL_2189733, EPI_ISL_2189734, EPI_ISL_2189735, EPI_ISL_2189736, EPI_ISL_2189737, EPI_ISL_2189740, EPI_ISL_2189741, EPI_ISL_2189742, EPI_ISL_2189743, EPI_ISL_2189745, EPI_ISL_2189746, EPI_ISL_2189747, EPI_ISL_2189748, EPI_ISL_2189749, EPI_ISL_2189750, EPI_ISL_2189753, EPI_ISL_2189754, EPI_ISL_2189755, EPI_ISL_2189756, EPI_ISL_2189758, EPI_ISL_2189759, EPI_ISL_2189760, EPI_ISL_2189761, EPI_ISL_2189762, EPI_ISL_2189764, EPI_ISL_2189765, EPI_ISL_2189766, EPI_ISL_2189767, EPI_ISL_2189769, EPI_ISL_2189770, EPI_ISL_2189771, EPI_ISL_2189772, EPI_ISL_2189773, EPI_ISL_2189775, EPI_ISL_2189776, EPI_ISL_2189777, EPI_ISL_2189778, EPI_ISL_2189779, EPI_ISL_2189781, EPI_ISL_2189783, EPI_ISL_2189784, EPI_ISL_2189785, EPI_ISL_2189786, EPI_ISL_2189787, EPI_ISL_2189788, EPI_ISL_2189789, EPI_ISL_2189790, EPI_ISL_2189791, EPI_ISL_2189792, EPI_ISL_2189793, EPI_ISL_2189794, EPI_ISL_2189795, EPI_ISL_2189796, EPI_ISL_2189797, EPI_ISL_2189798, EPI_ISL_2189799, EPI_ISL_2189800, EPI_ISL_2189801, EPI_ISL_2189803, EPI_ISL_2189804, EPI_ISL_2189805, EPI_ISL_2189806, EPI_ISL_2189807, EPI_ISL_2189808, EPI_ISL_2189809, EPI_ISL_2189810, EPI_ISL_2189811, EPI_ISL_2189812, EPI_ISL_2189813, EPI_ISL_2189815, EPI_ISL_2189816, EPI_ISL_2189817, EPI_ISL_2189818, EPI_ISL_2189819, EPI_ISL_2189820, EPI_ISL_2189821, EPI_ISL_2189822, EPI_ISL_2189823, EPI_ISL_2189824, EPI_ISL_2189825, EPI_ISL_2189826, EPI_ISL_2189827, EPI_ISL_2189828, EPI_ISL_2189829, EPI_ISL_2189830, EPI_ISL_2189831, EPI_ISL_2189832, EPI_ISL_2189833, EPI_ISL_2189834, EPI_ISL_2189835, EPI_ISL_2189836, EPI_ISL_2189837, EPI_ISL_2189838, EPI_ISL_2189839, EPI_ISL_2189840, EPI_ISL_2189841, EPI_ISL_2189842, EPI_ISL_2189843, EPI_ISL_2189844, EPI_ISL_2189845, EPI_ISL_2189846, EPI_ISL_2189848, EPI_ISL_2189849, EPI_ISL_2189850, EPI_ISL_2189851, EPI_ISL_2189852, EPI_ISL_2189853, EPI_ISL_2189854, EPI_ISL_2189855, EPI_ISL_2189856, EPI_ISL_2189857, EPI_ISL_2189858, EPI_ISL_2189859, EPI_ISL_2189860, EPI_ISL_2189861, EPI_ISL_2189862, EPI_ISL_2189863 | | | |
| see above | ICMR-National Institute of Virology | NCDC Delhi, Biotechnology Division | Meena Datta,V.Vipat,S.Jadhav,Priyanka Singh,U.Saha ,M.Das,Uma Sharma,G.Divekar,T.Sanjeevi,Manoj K Singh,S Jadhav,Radhakrishnan V. S,A Walimbe ,Robin Marwal,V Malik,Mahesh S Dhar,K.Iyengar,Kalaiarasan Ponnusamy,K P.Shinde,Hemlata Lall,K.Korabu,Z. Sayyed,Hema Gogia, S.Shelkande,R.Verma,Preeti Madan,S.Bhorekar,B.Apoorva,Sandhya Kabra,A.Titkare,H.Kengale,Sujeet K Singh,V.Autude,A.Awhale,M L Choudhary,Partha Rakshit |

| | | | |
|---|---|---|---|
| EPI_ISL_2225612 | National Centre For Cell Science | National Centre For Cell Science - INSACOG | Dhiraj Paul, Shivang P. Bhanushali, Mitali Inamdar, Mohak P Gujare, Manoj Kumar Bhat, Ajay Pillai, INSACOG Consortium team, Yogesh Shouche |
| EPI_ISL_2225613, EPI_ISL_2225614, EPI_ISL_2225615, EPI_ISL_2225616, EPI_ISL_2225617, EPI_ISL_2225618, EPI_ISL_2225619, EPI_ISL_2225620, EPI_ISL_2225621, EPI_ISL_2225622, EPI_ISL_2225623, EPI_ISL_2225624, EPI_ISL_2225625, EPI_ISL_2225626, EPI_ISL_2225627, EPI_ISL_2225628, EPI_ISL_2225629, EPI_ISL_2225630, EPI_ISL_2225631, EPI_ISL_2225632, EPI_ISL_2225633, EPI_ISL_2225634, EPI_ISL_2225635, EPI_ISL_2225636, EPI_ISL_2225637, EPI_ISL_2225638, EPI_ISL_2225639, EPI_ISL_2225640, EPI_ISL_2225641, EPI_ISL_2225642, EPI_ISL_2225643, EPI_ISL_2225644, EPI_ISL_2225645, EPI_ISL_2225646, EPI_ISL_2225647, EPI_ISL_2225648, EPI_ISL_2225649, EPI_ISL_2225650, EPI_ISL_2225651, EPI_ISL_2225652, EPI_ISL_2225653, EPI_ISL_2225654, EPI_ISL_2225655, EPI_ISL_2225656, EPI_ISL_2225657, EPI_ISL_2225658, EPI_ISL_2225659, EPI_ISL_2225660, EPI_ISL_2225661, EPI_ISL_2225662, EPI_ISL_2225663, EPI_ISL_2225664, EPI_ISL_2225665, EPI_ISL_2225666, EPI_ISL_2225667, EPI_ISL_2225668, EPI_ISL_2225669, EPI_ISL_2225670, EPI_ISL_2225671, EPI_ISL_2225672, EPI_ISL_2225673, EPI_ISL_2225674, EPI_ISL_2225675, EPI_ISL_2225676, EPI_ISL_2225677, EPI_ISL_2225678, EPI_ISL_2225679, EPI_ISL_2225680, EPI_ISL_2225681, EPI_ISL_2225682, EPI_ISL_2225683, EPI_ISL_2225684, EPI_ISL_2225685, EPI_ISL_2225686, EPI_ISL_2225687, EPI_ISL_2225688, EPI_ISL_2225689, EPI_ISL_2225690, EPI_ISL_2225691, EPI_ISL_2225692, EPI_ISL_2225693, EPI_ISL_2225694, EPI_ISL_2225695, EPI_ISL_2225696, EPI_ISL_2225697, EPI_ISL_2233145, EPI_ISL_2233146 | | | |
| see above | National Centre For Cell Science | National Centre For Cell Science - INSACOG | Dhiraj Paul, Shivang P. Bhanushali, Mitali Inamdar, Mohak P Gujare, Manoj Kumar Bhat, Ajay Pillai, INSACOG Consortium team, Yogesh Shouche |
| EPI_ISL_2233147 | National Centre For Cell Science | National Centre For Cell Science - INSACOG | Dhiraj Paul, Mohak P Gujare, Shivang P. Bhanushali, Mitali Inamdar, Manoj Kumar Bhat, Ajay Pillai, INSACOG Consortium team, Yogesh Shouche |
| EPI_ISL_2272490 | Indian Council of Medical Research-National Institute of Virology, Microbial Containment Complex | Indian Council of Medical Research-National Institute of Virology, Microbial Containment Complex | Pragya D Yadav |
| EPI_ISL_2304047, EPI_ISL_2304048, EPI_ISL_2304049, EPI_ISL_2304050, EPI_ISL_2304051, EPI_ISL_2304052, EPI_ISL_2304053, EPI_ISL_2304054 | National Centre For Cell Science | National Centre For Cell Science - INSACOG | Dhiraj Paul, Mohak P Gujare, Shivang P. Bhanushali, Mitali Inamdar, Manoj Kumar Bhat, Ajay Pillai, INSACOG Consortium team, Yogesh Shouche |
| EPI_ISL_2304055, EPI_ISL_2304056 | Armed Forces Medical College Pune | National Centre For Cell Science - INSACOG | Dhiraj Paul, Mohak P Gujare, Shivang P. Bhanushali, Mitali Inamdar, Manoj Kumar Bhat, Ajay Pillai, INSACOG Consortium team, Yogesh Shouche |
| EPI_ISL_2304057, EPI_ISL_2304058, EPI_ISL_2304059, EPI_ISL_2304060, EPI_ISL_2304061, EPI_ISL_2304062, EPI_ISL_2304063, EPI_ISL_2304064, EPI_ISL_2304065, EPI_ISL_2304066, EPI_ISL_2304067, EPI_ISL_2304068, EPI_ISL_2304069, EPI_ISL_2304070, EPI_ISL_2304071, EPI_ISL_2304072, EPI_ISL_2304073, EPI_ISL_2304074, EPI_ISL_2304075, EPI_ISL_2304076, EPI_ISL_2304077, EPI_ISL_2304078, EPI_ISL_2304079, EPI_ISL_2304080, EPI_ISL_2304081, EPI_ISL_2304082, EPI_ISL_2304083, EPI_ISL_2304084, EPI_ISL_2304085 | | | |
| see above | National Centre For Cell Science | National Centre For Cell Science - INSACOG | Dhiraj Paul, Mohak P Gujare, Shivang P. Bhanushali, Mitali Inamdar, Manoj Kumar Bhat, Ajay Pillai, INSACOG Consortium team, Yogesh Shouche |
| EPI_ISL_2304086, EPI_ISL_2304087, EPI_ISL_2304088, EPI_ISL_2304089 | Armed Forces Medical College Pune | National Centre For Cell Science - INSACOG | Dhiraj Paul, Mohak P Gujare, Shivang P. Bhanushali, Mitali Inamdar, Manoj Kumar Bhat, Ajay Pillai, INSACOG Consortium team, Yogesh Shouche |
| EPI_ISL_2304090, EPI_ISL_2304091 | National Centre For Cell Science | National Centre For Cell Science - INSACOG | Dhiraj Paul, Mohak P Gujare, Shivang P. Bhanushali, Mitali Inamdar, Manoj Kumar Bhat, Ajay Pillai, INSACOG Consortium team, Yogesh Shouche |
| EPI_ISL_2304092, EPI_ISL_2304093, EPI_ISL_2304094, EPI_ISL_2304095, EPI_ISL_2304096, EPI_ISL_2304097, EPI_ISL_2304098 | Armed Forces Medical College Pune | National Centre For Cell Science - INSACOG | Dhiraj Paul, Mohak P Gujare, Shivang P. Bhanushali, Mitali Inamdar, Manoj Kumar Bhat, Ajay Pillai, INSACOG Consortium team, Yogesh Shouche |
| EPI_ISL_2304099 | National Centre For Cell Science | National Centre For Cell Science - INSACOG | Dhiraj Paul, Mohak P Gujare, Shivang P. Bhanushali, Mitali Inamdar, Manoj Kumar Bhat, Ajay Pillai, INSACOG Consortium team, Yogesh Shouche |
| EPI_ISL_2304100, EPI_ISL_2304101, EPI_ISL_2304102 | Armed Forces Medical College Pune | National Centre For Cell Science - INSACOG | Dhiraj Paul, Mohak P Gujare, Shivang P. Bhanushali, Mitali Inamdar, Manoj Kumar Bhat, Ajay Pillai, INSACOG Consortium team, Yogesh Shouche |
| EPI_ISL_2304103 | National Centre For Cell Science | National Centre For Cell Science - INSACOG | Dhiraj Paul, Mohak P Gujare, Shivang P. Bhanushali, Mitali Inamdar, Manoj Kumar Bhat, Ajay Pillai, INSACOG Consortium team, Yogesh Shouche |
| EPI_ISL_2304104, EPI_ISL_2304105, EPI_ISL_2304106, EPI_ISL_2304107, EPI_ISL_2304108, EPI_ISL_2304109, EPI_ISL_2304110, EPI_ISL_2304111, EPI_ISL_2304112, EPI_ISL_2304113, EPI_ISL_2304114, EPI_ISL_2304115, EPI_ISL_2304116, EPI_ISL_2304117, EPI_ISL_2304118, EPI_ISL_2304119, EPI_ISL_2304120, EPI_ISL_2304121, EPI_ISL_2304122 | | | |
| see above | Armed Forces Medical College Pune | National Centre For Cell Science - INSACOG | Dhiraj Paul, Mohak P Gujare, Shivang P. Bhanushali, Mitali Inamdar, Manoj Kumar Bhat, Ajay Pillai, INSACOG Consortium team, Yogesh Shouche |
| EPI_ISL_2304123 | National Centre For Cell Science | National Centre For Cell Science - INSACOG | Dhiraj Paul, Mohak P Gujare, Shivang P. Bhanushali, Mitali Inamdar, Manoj Kumar Bhat, Ajay Pillai, INSACOG Consortium team, Yogesh Shouche |
| EPI_ISL_2304124, EPI_ISL_2304125, EPI_ISL_2304126, EPI_ISL_2304127, EPI_ISL_2304128, EPI_ISL_2304129 | Armed Forces Medical College Pune | National Centre For Cell Science - INSACOG | Dhiraj Paul, Mohak P Gujare, Shivang P. Bhanushali, Mitali Inamdar, Manoj Kumar Bhat, Ajay Pillai, INSACOG Consortium team, Yogesh Shouche |
| EPI_ISL_2304130 | National Centre For Cell Science | National Centre For Cell Science - INSACOG | Dhiraj Paul, Mohak P Gujare, Shivang P. Bhanushali, Mitali Inamdar, Manoj Kumar Bhat, Ajay Pillai, INSACOG Consortium team, Yogesh Shouche |
| EPI_ISL_2304131 | Armed Forces Medical College Pune | National Centre For Cell Science - INSACOG | Dhiraj Paul, Mohak P Gujare, Shivang P. Bhanushali, Mitali Inamdar, Manoj Kumar Bhat, Ajay Pillai, INSACOG Consortium team, Yogesh Shouche |
| EPI_ISL_2332704, EPI_ISL_2332705, EPI_ISL_2332706, EPI_ISL_2332707, EPI_ISL_2332708, EPI_ISL_2332709, EPI_ISL_2332710, EPI_ISL_2332711, EPI_ISL_2332712, EPI_ISL_2332713, EPI_ISL_2332714, EPI_ISL_2332715, EPI_ISL_2332716, EPI_ISL_2332717, EPI_ISL_2332718, EPI_ISL_2332719, EPI_ISL_2332720, EPI_ISL_2332721, EPI_ISL_2332722, EPI_ISL_2332723, EPI_ISL_2332724, EPI_ISL_2332725, EPI_ISL_2332726, EPI_ISL_2332727, EPI_ISL_2332728, EPI_ISL_2332729, EPI_ISL_2332730, EPI_ISL_2332731, EPI_ISL_2332732, EPI_ISL_2332733, EPI_ISL_2332734, EPI_ISL_2332735, EPI_ISL_2332736, EPI_ISL_2332737, EPI_ISL_2332738, EPI_ISL_2332739, EPI_ISL_2332740, EPI_ISL_2332741, EPI_ISL_2332742, EPI_ISL_2332743, EPI_ISL_2332744, EPI_ISL_2332745, EPI_ISL_2332746, EPI_ISL_2332747, EPI_ISL_2332748, EPI_ISL_2332749, EPI_ISL_2332750, EPI_ISL_2332751, EPI_ISL_2332752, EPI_ISL_2332753, EPI_ISL_2332754, EPI_ISL_2332755, EPI_ISL_2332756, EPI_ISL_2332757, EPI_ISL_2332758, EPI_ISL_2332759, EPI_ISL_2332760, EPI_ISL_2332761, EPI_ISL_2332762, EPI_ISL_2332763, EPI_ISL_2332764, EPI_ISL_2332765, EPI_ISL_2332766, EPI_ISL_2332767, EPI_ISL_2332768, EPI_ISL_2332769, EPI_ISL_2332770, EPI_ISL_2332771, EPI_ISL_2332772, EPI_ISL_2332773, EPI_ISL_2332774, EPI_ISL_2332775, EPI_ISL_2332776, EPI_ISL_2332777, EPI_ISL_2332778, EPI_ISL_2332779, EPI_ISL_2332780, EPI_ISL_2332781, EPI_ISL_2332782, EPI_ISL_2332783, EPI_ISL_2332784, EPI_ISL_2332785, EPI_ISL_2332786, EPI_ISL_2332787, EPI_ISL_2332788 | | | |
| see above | National Centre For Cell Science | National Centre For Cell Science - INSACOG | Dhiraj Paul, Shivang P. Bhanushali, Mitali Inamdar, Mohak P Gujare, Manoj Kumar Bhat, Ajay Pillai, INSACOG Consortium team, Yogesh Shouche |
| EPI_ISL_2341667, EPI_ISL_2341668, EPI_ISL_2341672, EPI_ISL_2341673, EPI_ISL_2341677, EPI_ISL_2341678, EPI_ISL_2341686, EPI_ISL_2341688, EPI_ISL_2341702, EPI_ISL_2341710, EPI_ISL_2341713, EPI_ISL_2341718, EPI_ISL_2341720, EPI_ISL_2341725, EPI_ISL_2341730, EPI_ISL_2341733, EPI_ISL_2341734, EPI_ISL_2341735, EPI_ISL_2341736, EPI_ISL_2341744, EPI_ISL_2341748, EPI_ISL_2341749, EPI_ISL_2341750, EPI_ISL_2341751, EPI_ISL_2341752, EPI_ISL_2341753, EPI_ISL_2341756, EPI_ISL_2341761, EPI_ISL_2341765, EPI_ISL_2341766, EPI_ISL_2341768, EPI_ISL_2341769, EPI_ISL_2341771, EPI_ISL_2341776, EPI_ISL_2341777, EPI_ISL_2341791, EPI_ISL_2341798, EPI_ISL_2341804, EPI_ISL_2341813, EPI_ISL_2341817, EPI_ISL_2341820, EPI_ISL_2341830, EPI_ISL_2341839, EPI_ISL_2341840, EPI_ISL_2341845, EPI_ISL_2341943, EPI_ISL_2341944, EPI_ISL_2341945, EPI_ISL_2341946, EPI_ISL_2341947, EPI_ISL_2341948, EPI_ISL_2341949, EPI_ISL_2341950, EPI_ISL_2341951, EPI_ISL_2341952, EPI_ISL_2341953, EPI_ISL_2341954, EPI_ISL_2341955, EPI_ISL_2341956, EPI_ISL_2341957, EPI_ISL_2341958, EPI_ISL_2341959, EPI_ISL_2341960, EPI_ISL_2341961, EPI_ISL_2341962, EPI_ISL_2341963, EPI_ISL_2341964, EPI_ISL_2341965, EPI_ISL_2341966, EPI_ISL_2341967, EPI_ISL_2341968, EPI_ISL_2341969, EPI_ISL_2341970, EPI_ISL_2341971, EPI_ISL_2341972, EPI_ISL_2341973, EPI_ISL_2341974, EPI_ISL_2341975, EPI_ISL_2341976, EPI_ISL_2341977, EPI_ISL_2341978, EPI_ISL_2341979, EPI_ISL_2341980, EPI_ISL_2341981, EPI_ISL_2341982, EPI_ISL_2341983, EPI_ISL_2341984, EPI_ISL_2341985, EPI_ISL_2341986, EPI_ISL_2341987, EPI_ISL_2341988, EPI_ISL_2341989, EPI_ISL_2341990, EPI_ISL_2341991, EPI_ISL_2341992, EPI_ISL_2342602, EPI_ISL_2342607, EPI_ISL_2342609, EPI_ISL_2342610, EPI_ISL_2342614, EPI_ISL_2342615, EPI_ISL_2342620, EPI_ISL_2342622, EPI_ISL_2342623, EPI_ISL_2342630, EPI_ISL_2342632, EPI_ISL_2342634, EPI_ISL_2342638, EPI_ISL_2342641, EPI_ISL_2342642, EPI_ISL_2342643, EPI_ISL_2342644, EPI_ISL_2342649, EPI_ISL_2342650, EPI_ISL_2342654, EPI_ISL_2342655, EPI_ISL_2342656, EPI_ISL_2342657, EPI_ISL_2342660, EPI_ISL_2342665, EPI_ISL_2342667, EPI_ISL_2342673, EPI_ISL_2342674, EPI_ISL_2342675, EPI_ISL_2342680, EPI_ISL_2342681, EPI_ISL_2342683, EPI_ISL_2342684, EPI_ISL_2342686, EPI_ISL_2342687, EPI_ISL_2342689, EPI_ISL_2342693, EPI_ISL_2342696, EPI_ISL_2342713, EPI_ISL_2342717, EPI_ISL_2342718, EPI_ISL_2342721, EPI_ISL_2342722, EPI_ISL_2342725, EPI_ISL_2342740, EPI_ISL_2342742, EPI_ISL_2342743, EPI_ISL_2342745, EPI_ISL_2342755, EPI_ISL_2342764, EPI_ISL_2342766, EPI_ISL_2342769, EPI_ISL_2342771, EPI_ISL_2342775, EPI_ISL_2342776, EPI_ISL_2342777, EPI_ISL_2342779, EPI_ISL_2342780, EPI_ISL_2342781, EPI_ISL_2342782, EPI_ISL_2342783, EPI_ISL_2342788, EPI_ISL_2342792, EPI_ISL_2342793, EPI_ISL_2342796, EPI_ISL_2342798, EPI_ISL_2342799, EPI_ISL_2342801, EPI_ISL_2342803, EPI_ISL_2342804, EPI_ISL_2342805, EPI_ISL_2342806, EPI_ISL_2342807, EPI_ISL_2342808, EPI_ISL_2342811, EPI_ISL_2342813, EPI_ISL_2342815, EPI_ISL_2342816, EPI_ISL_2342817, EPI_ISL_2342820, EPI_ISL_2342821, EPI_ISL_2342822, EPI_ISL_2342823, EPI_ISL_2342824, EPI_ISL_2342825, EPI_ISL_2342827, EPI_ISL_2342829, EPI_ISL_2342830, EPI_ISL_2342831, EPI_ISL_2342832, EPI_ISL_2342839, EPI_ISL_2342840, EPI_ISL_2342841, EPI_ISL_2342842, EPI_ISL_2342844, EPI_ISL_2342845, EPI_ISL_2342849, EPI_ISL_2342851, EPI_ISL_2342852, EPI_ISL_2342856, EPI_ISL_2342859, EPI_ISL_2342861, EPI_ISL_2342882, EPI_ISL_2342883, EPI_ISL_2342884, EPI_ISL_2342885, EPI_ISL_2342886, EPI_ISL_2342887, EPI_ISL_2342888, EPI_ISL_2342889, EPI_ISL_2342890, EPI_ISL_2342891, EPI_ISL_2342892, EPI_ISL_2342893, EPI_ISL_2342894, EPI_ISL_2342895, EPI_ISL_2342896, EPI_ISL_2342897, EPI_ISL_2342898, EPI_ISL_2342899, EPI_ISL_2342900, EPI_ISL_2342901, EPI_ISL_2342902, EPI_ISL_2342903, EPI_ISL_2342904, EPI_ISL_2342905, EPI_ISL_2342906, EPI_ISL_2342907, EPI_ISL_2342908, EPI_ISL_2342909, EPI_ISL_2342910, EPI_ISL_2342911, EPI_ISL_2342912, EPI_ISL_2342913, EPI_ISL_2342914, EPI_ISL_2342915, EPI_ISL_2342916, EPI_ISL_2342917, EPI_ISL_2342918, EPI_ISL_2342919, EPI_ISL_2342920, EPI_ISL_2342921, EPI_ISL_2342922, EPI_ISL_2342923, EPI_ISL_2342924, EPI_ISL_2342925, EPI_ISL_2342926, EPI_ISL_2342927, EPI_ISL_2342928, EPI_ISL_2342929, EPI_ISL_2342930, EPI_ISL_2342931, EPI_ISL_2342932, EPI_ISL_2342933, EPI_ISL_2342934, EPI_ISL_2342935, EPI_ISL_2342936, EPI_ISL_2342937, EPI_ISL_2342938, EPI_ISL_2342939, EPI_ISL_2342940, EPI_ISL_2342941, EPI_ISL_2342942, EPI_ISL_2342943, EPI_ISL_2342944, EPI_ISL_2342945, EPI_ISL_2342946, EPI_ISL_2342947, EPI_ISL_2342948, EPI_ISL_2342949, EPI_ISL_2342950, EPI_ISL_2342951, EPI_ISL_2342952, EPI_ISL_2342953, EPI_ISL_2342954, EPI_ISL_2342955, EPI_ISL_2342956, EPI_ISL_2342957, EPI_ISL_2342958, EPI_ISL_2342959, EPI_ISL_2342960, EPI_ISL_2342961, EPI_ISL_2342962, EPI_ISL_2342963, EPI_ISL_2342964, EPI_ISL_2342965, EPI_ISL_2342966, EPI_ISL_2342967, EPI_ISL_2342968, EPI_ISL_2342969, EPI_ISL_2342970 | | | |
| see above | NCCS, Pune | Institute of Life Sciences - INSACOG | Sunil K. Raghav, Safal Walia, Arup Ghosh, Atimukta Jha, Amol M. Kanampalliwar, Omprakash Shiriwas, Sana Fatma, Shifu Aggarwal, Rupesh Dash, Rajeeb Swain, Punit Prasad, INSACOG Consortium, Ajay Parida |
| EPI_ISL_2365423, EPI_ISL_2365424 | National Centre For Cell Science | National Centre For Cell Science-INSACOG | Dhiraj Paul, Shivang P. Bhanushali, Mohak P Gujare, Mitali Inamdar, Manoj Kumar Bhat, Ajay Pillai, INSACOG Consortium team, Yogesh Shouche |
| EPI_ISL_2382915, EPI_ISL_2382916, EPI_ISL_2382917, EPI_ISL_2382918, EPI_ISL_2382919, EPI_ISL_2382920, EPI_ISL_2382921, EPI_ISL_2382922, EPI_ISL_2382923, EPI_ISL_2382924, EPI_ISL_2382925, EPI_ISL_2382926, EPI_ISL_2382927, EPI_ISL_2382928, EPI_ISL_2382929, EPI_ISL_2382930, EPI_ISL_2382931, EPI_ISL_2382932, EPI_ISL_2382933, EPI_ISL_2382934, EPI_ISL_2382935, EPI_ISL_2382936, EPI_ISL_2382937, EPI_ISL_2382938, EPI_ISL_2382939, EPI_ISL_2382940, EPI_ISL_2382941, EPI_ISL_2382970, EPI_ISL_2382971, EPI_ISL_2382972, EPI_ISL_2382973, EPI_ISL_2382974, EPI_ISL_2382975, EPI_ISL_2382976, EPI_ISL_2382977, EPI_ISL_2382978, EPI_ISL_2382979, EPI_ISL_2382980, EPI_ISL_2382981, EPI_ISL_2382982, EPI_ISL_2382983, EPI_ISL_2382984, EPI_ISL_2382985, EPI_ISL_2382986, EPI_ISL_2382987, EPI_ISL_2382988, EPI_ISL_2382989, EPI_ISL_2382990, EPI_ISL_2382991, EPI_ISL_2382992, EPI_ISL_2382993, EPI_ISL_2382994, EPI_ISL_2382995, EPI_ISL_2382996, EPI_ISL_2382997, EPI_ISL_2382998, EPI_ISL_2382999, EPI_ISL_2383000, EPI_ISL_2383001, EPI_ISL_2383002, EPI_ISL_2383003, EPI_ISL_2383004, EPI_ISL_2383005, EPI_ISL_2383006, EPI_ISL_2383007 | | | |
| see above | National Centre For Cell Science | National Centre For Cell Science-INSACOG | Dhiraj Paul, Shivang P. Bhanushali, Mohak P Gujare, Mitali Inamdar, Manoj Kumar Bhat, Ajay Pillai, INSACOG Consortium team, Yogesh Shouche |
| EPI_ISL_2399418, EPI_ISL_2399419, EPI_ISL_2399420, EPI_ISL_2399422 | Indian Council of Medical Research-National Institute of Virology, Microbial Containment Complex | Indian Council of Medical Research-National Institute of Virology, Microbial Containment Complex | Pragya D Yadav |
| EPI_ISL_2441372, EPI_ISL_2441373, EPI_ISL_2441374, EPI_ISL_2441375, EPI_ISL_2441376, EPI_ISL_2441377, EPI_ISL_2441378, EPI_ISL_2441379, EPI_ISL_2441380, EPI_ISL_2441381, EPI_ISL_2441382, EPI_ISL_2441383, EPI_ISL_2441384, EPI_ISL_2441385, EPI_ISL_2441386, EPI_ISL_2441387, EPI_ISL_2441388, | | | |

| | | | |
|---|---|---|---|
| EPI_ISL_2441389, EPI_ISL_2441390, EPI_ISL_2441391, EPI_ISL_2441392, EPI_ISL_2441393, EPI_ISL_2441394, EPI_ISL_2441395, EPI_ISL_2441396, EPI_ISL_2441397, EPI_ISL_2441398, EPI_ISL_2441399, EPI_ISL_2441400, EPI_ISL_2441401, EPI_ISL_2441402, EPI_ISL_2441403, EPI_ISL_2441404, EPI_ISL_2441405, EPI_ISL_2441406, EPI_ISL_2441407, EPI_ISL_2441408, EPI_ISL_2441409, EPI_ISL_2441410, EPI_ISL_2441411, EPI_ISL_2441412, EPI_ISL_2441413, EPI_ISL_2441414, EPI_ISL_2441415, EPI_ISL_2441416, EPI_ISL_2441417, EPI_ISL_2441418, EPI_ISL_2441419, EPI_ISL_2441420, EPI_ISL_2441421, EPI_ISL_2441422, EPI_ISL_2441423, EPI_ISL_2441424, EPI_ISL_2441425, EPI_ISL_2441426, EPI_ISL_2441427, EPI_ISL_2441428, EPI_ISL_2441429, EPI_ISL_2441430, EPI_ISL_2441431, EPI_ISL_2441432, EPI_ISL_2441433, EPI_ISL_2441434, EPI_ISL_2441435, EPI_ISL_2441436, EPI_ISL_2441437, EPI_ISL_2441438, EPI_ISL_2441439, EPI_ISL_2441440, EPI_ISL_2441441, EPI_ISL_2441442, EPI_ISL_2441443, EPI_ISL_2441444, EPI_ISL_2441445, EPI_ISL_2441446, EPI_ISL_2441447, EPI_ISL_2441448, EPI_ISL_2441449, EPI_ISL_2441450, EPI_ISL_2441451, EPI_ISL_2441452, EPI_ISL_2441453, EPI_ISL_2441454, EPI_ISL_2441455, EPI_ISL_2441456, EPI_ISL_2441457, EPI_ISL_2441458, EPI_ISL_2441459, EPI_ISL_2441460, EPI_ISL_2441461, EPI_ISL_2441462, EPI_ISL_2441463, EPI_ISL_2441464, EPI_ISL_2441465, EPI_ISL_2441466, EPI_ISL_2441467, EPI_ISL_2441468, EPI_ISL_2441469, EPI_ISL_2441470, EPI_ISL_2441471 | | | |
| see above | CSIR-National Environmental Engineering Research Institute | CSIR-Centre for Cellular and Molecular Biology-INSACOG | Payel Mukherjee, Lamuk Zaveri, Onkar Kulkarni,Tulasi Nagabandi, Sofia Banu, Ara Sreenivas,Shreekant Verma, Amareshwar Vodapalli ,B Himasri,Valli Nagalakshmi Undamatla, Sumedha Avadhanula, Vidhyadhari Methuku, Priya Nurkurthy, Archana Bharadwaj Siva, Karthik Bharadwaj Tallapaka,Krishna Khairnar,Rakesh K Mishra,Divya Tej Sowpati |
| EPI_ISL_2443364, EPI_ISL_2443365, EPI_ISL_2443366, EPI_ISL_2443367, EPI_ISL_2443368, EPI_ISL_2443369, EPI_ISL_2443370, EPI_ISL_2443371, EPI_ISL_2443372, EPI_ISL_2443373, EPI_ISL_2443374, EPI_ISL_2443376, EPI_ISL_2443377, EPI_ISL_2443378, EPI_ISL_2443379, EPI_ISL_2443380 | | | |
| see above | Indian Council of Medical Research-National Institute of Virology, Microbial Containment Complex | Indian Council of Medical Research-National Institute of Virology, Microbial Containment Complex | Pragya D. Yadav |
| EPI_ISL_2459659 | National Centre for Disease Control (NCDC) Biotechnology Division, Delhi | NCDC Delhi, Biotechnology Division INSACOG | Kalaiarasan Ponnusamy, Meena Datta, Priyanka Singh, Uma Sharma, Manoj K Singh, Radhakrishnan V. S, Robin Marwal, Mahesh S Dhar, Hemlata Lall, Hema Gogia, Preeti Madan, Sandhya Kabra, Sujeet K Singh, Partha Rakshit |
| EPI_ISL_2459692, EPI_ISL_2459699, EPI_ISL_2459704, EPI_ISL_2459709, EPI_ISL_2459715, EPI_ISL_2459722, EPI_ISL_2459725, EPI_ISL_2459726, EPI_ISL_2459731, EPI_ISL_2459738, EPI_ISL_2459739, EPI_ISL_2459740, EPI_ISL_2459741, EPI_ISL_2459743, EPI_ISL_2459745, EPI_ISL_2459747, EPI_ISL_2459749, EPI_ISL_2459751, EPI_ISL_2459752, EPI_ISL_2459753, EPI_ISL_2459754, EPI_ISL_2459757, EPI_ISL_2459758, EPI_ISL_2459759, EPI_ISL_2459760, EPI_ISL_2459763, EPI_ISL_2459765, EPI_ISL_2459767, EPI_ISL_2459769, EPI_ISL_2459771, EPI_ISL_2459772, EPI_ISL_2459773, EPI_ISL_2459775, EPI_ISL_2459776, EPI_ISL_2459777, EPI_ISL_2459778, EPI_ISL_2459780, EPI_ISL_2459781 | | | |
| see above | National Centre for Disease Control (NCDC) Biotechnology Division, Delhi | NCDC Delhi, Biotechnology Division INSACOG | Mahesh S Dhar, Kalaiarasan Ponnusamy, Meena Datta, Priyanka Singh, Uma Sharma, Manoj K Singh, Radhakrishnan V. S, Robin Marwal, Hemlata Lall, Hema Gogia, Preeti Madan, Sandhya Kabra, Sujeet K Singh, Partha Rakshit |
| EPI_ISL_2459782 | National Centre for Disease Control (NCDC) Biotechnology Division, Delhi | NCDC Delhi, Biotechnology Division INSACOG | Kalaiarasan Ponnusamy, Meena Datta, Priyanka Singh, Uma Sharma, Manoj K Singh, Radhakrishnan V. S, Robin Marwal, Mahesh S Dhar, Hemlata Lall, Hema Gogia, Preeti Madan, Sandhya Kabra, Sujeet K Singh, Partha Rakshit |
| EPI_ISL_2459783, EPI_ISL_2459786, EPI_ISL_2459790, EPI_ISL_2459793, EPI_ISL_2459794 | National Centre for Disease Control (NCDC) Biotechnology Division, Delhi | NCDC Delhi, Biotechnology Division INSACOG | Mahesh S Dhar, Kalaiarasan Ponnusamy, Meena Datta, Priyanka Singh, Uma Sharma, Manoj K Singh, Radhakrishnan V. S, Robin Marwal, Hemlata Lall, Hema Gogia, Preeti Madan, Sandhya Kabra, Sujeet K Singh, Partha Rakshit |
| EPI_ISL_2459795 | National Centre for Disease Control (NCDC) Biotechnology Division, Delhi | NCDC Delhi, Biotechnology Division INSACOG | Kalaiarasan Ponnusamy, Meena Datta, Priyanka Singh, Uma Sharma, Manoj K Singh, Radhakrishnan V. S, Robin Marwal, Mahesh S Dhar, Hemlata Lall, Hema Gogia, Preeti Madan, Sandhya Kabra, Sujeet K Singh, Partha Rakshit |
| EPI_ISL_2459796, EPI_ISL_2459798, EPI_ISL_2459801, EPI_ISL_2459802, EPI_ISL_2459803, EPI_ISL_2459805, EPI_ISL_2459806, EPI_ISL_2459807, EPI_ISL_2459808, EPI_ISL_2459809, EPI_ISL_2459810, EPI_ISL_2459811, EPI_ISL_2459814, EPI_ISL_2459815, EPI_ISL_2459816, EPI_ISL_2459817, EPI_ISL_2459818, EPI_ISL_2459821, EPI_ISL_2459824, EPI_ISL_2459825, EPI_ISL_2459827, EPI_ISL_2459828, EPI_ISL_2459829, EPI_ISL_2459831, EPI_ISL_2459832, EPI_ISL_2459836, EPI_ISL_2459840, EPI_ISL_2459845, EPI_ISL_2459846, EPI_ISL_2459847, EPI_ISL_2459849, EPI_ISL_2459851, EPI_ISL_2459854, EPI_ISL_2459868, EPI_ISL_2459869, EPI_ISL_2459875 | | | |
| see above | National Centre for Disease Control (NCDC) Biotechnology Division, Delhi | NCDC Delhi, Biotechnology Division INSACOG | Mahesh S Dhar, Kalaiarasan Ponnusamy, Meena Datta, Priyanka Singh, Uma Sharma, Manoj K Singh, Radhakrishnan V. S, Robin Marwal, Hemlata Lall, Hema Gogia, Preeti Madan, Sandhya Kabra, Sujeet K Singh, Partha Rakshit |
| EPI_ISL_2459973 | National Centre for Disease Control (NCDC) Biotechnology Division, Delhi | NCDC Delhi, Biotechnology Division INSACOG | Priyanka Singh, Uma Sharma, Manoj K Singh, Radhakrishnan V. S, Robin Marwal, Mahesh S Dhar, Kalaiarasan Ponnusamy, Meena Datta, Hemlata Lall, Hema Gogia, Preeti Madan, Sandhya Kabra, Sujeet K Singh, Partha Rakshit |
| EPI_ISL_2459974, EPI_ISL_2459977, EPI_ISL_2459983, EPI_ISL_2459987, EPI_ISL_2459992, EPI_ISL_2459997 | National Centre for Disease Control (NCDC) Biotechnology Division, Delhi | NCDC Delhi, Biotechnology Division INSACOG | Uma Sharma, Manoj K Singh, Radhakrishnan V. S, Robin Marwal, Mahesh S Dhar, Kalaiarasan Ponnusamy, Meena Datta, Priyanka Singh, Hemlata Lall, Hema Gogia, Preeti Madan, Sandhya Kabra, Sujeet K Singh, Partha Rakshit |
| EPI_ISL_2460003 | National Centre for Disease Control (NCDC) Biotechnology Division, Delhi | NCDC Delhi, Biotechnology Division INSACOG | Priyanka Singh, Uma Sharma, Manoj K Singh, Radhakrishnan V. S, Robin Marwal, Mahesh S Dhar, Kalaiarasan Ponnusamy, Meena Datta, Hemlata Lall, Hema Gogia, Preeti Madan, Sandhya Kabra, Sujeet K Singh, Partha Rakshit |
| EPI_ISL_2460007, EPI_ISL_2460009, EPI_ISL_2460010, EPI_ISL_2460016, EPI_ISL_2460018 | National Centre for Disease Control (NCDC) Biotechnology Division, Delhi | NCDC Delhi, Biotechnology Division INSACOG | Uma Sharma, Manoj K Singh, Radhakrishnan V. S, Robin Marwal, Mahesh S Dhar, Kalaiarasan Ponnusamy, Meena Datta, Priyanka Singh, Hemlata Lall, Hema Gogia, Preeti Madan, Sandhya Kabra, Sujeet K Singh, Partha Rakshit |
| EPI_ISL_2460021, EPI_ISL_2460030, EPI_ISL_2460031, EPI_ISL_2460039 | National Centre for Disease Control (NCDC) Biotechnology Division, Delhi | NCDC Delhi, Biotechnology Division INSACOG | Priyanka Singh, Uma Sharma, Manoj K Singh, Radhakrishnan V. S, Robin Marwal, Mahesh S Dhar, Kalaiarasan Ponnusamy, Meena Datta, Hemlata Lall, Hema Gogia, Preeti Madan, Sandhya Kabra, Sujeet K Singh, Partha Rakshit |
| EPI_ISL_2460040, EPI_ISL_2460041, EPI_ISL_2460042, EPI_ISL_2460043, EPI_ISL_2460044, EPI_ISL_2460045, EPI_ISL_2460046, EPI_ISL_2460047, EPI_ISL_2460048, EPI_ISL_2460049, EPI_ISL_2460050, EPI_ISL_2460052, EPI_ISL_2460072, EPI_ISL_2460073, EPI_ISL_2460089, EPI_ISL_2460096, EPI_ISL_2460116, EPI_ISL_2460139, EPI_ISL_2460151 | | | |
| see above | National Centre for Disease Control (NCDC) Biotechnology Division, Delhi | NCDC Delhi, Biotechnology Division INSACOG | Uma Sharma, Manoj K Singh, Radhakrishnan V. S, Robin Marwal, Mahesh S Dhar, Kalaiarasan Ponnusamy, Meena Datta, Priyanka Singh, Hemlata Lall, Hema Gogia, Preeti Madan, Sandhya Kabra, Sujeet K Singh, Partha Rakshit |
| EPI_ISL_2460156 | National Centre for Disease Control (NCDC) Biotechnology Division, Delhi | NCDC Delhi, Biotechnology Division INSACOG | Priyanka Singh, Uma Sharma, Manoj K Singh, Radhakrishnan V. S, Robin Marwal, Mahesh S Dhar, Kalaiarasan Ponnusamy, Meena Datta, Hemlata Lall, Hema Gogia, Preeti Madan, Sandhya Kabra, Sujeet K Singh, Partha Rakshit |
| EPI_ISL_2460168, EPI_ISL_2460172 | National Centre for Disease Control (NCDC) Biotechnology Division, Delhi | NCDC Delhi, Biotechnology Division INSACOG | Uma Sharma, Manoj K Singh, Radhakrishnan V. S, Robin Marwal, Mahesh S Dhar, Kalaiarasan Ponnusamy, Meena Datta, Priyanka Singh, Hemlata Lall, Hema Gogia, Preeti Madan, Sandhya Kabra, Sujeet K Singh, Partha Rakshit |
| EPI_ISL_2460183, EPI_ISL_2460184, EPI_ISL_2460185, EPI_ISL_2460186, EPI_ISL_2460187 | National Centre for Disease Control (NCDC) Biotechnology Division, Delhi | NCDC Delhi, Biotechnology Division INSACOG | Priyanka Singh, Uma Sharma, Manoj K Singh, Radhakrishnan V. S, Robin Marwal, Mahesh S Dhar, Kalaiarasan Ponnusamy, Meena Datta, Hemlata Lall, Hema Gogia, Preeti Madan, Sandhya Kabra, Sujeet K Singh, Partha Rakshit |
| EPI_ISL_2460188, EPI_ISL_2460189, EPI_ISL_2460190, EPI_ISL_2460191, EPI_ISL_2460192, EPI_ISL_2460193, EPI_ISL_2460194, EPI_ISL_2460195, EPI_ISL_2460196 | National Centre for Disease Control (NCDC) Biotechnology Division, Delhi | NCDC Delhi, Biotechnology Division INSACOG | Uma Sharma, Manoj K Singh, Radhakrishnan V. S, Robin Marwal, Mahesh S Dhar, Kalaiarasan Ponnusamy, Meena Datta, Priyanka Singh, Hemlata Lall, Hema Gogia, Preeti Madan, Sandhya Kabra, Sujeet K Singh, Partha Rakshit |
| EPI_ISL_2460229 | National Centre for Disease Control (NCDC) Biotechnology Division, Delhi | NCDC Delhi, Biotechnology Division INSACOG | Priyanka Singh, Uma Sharma, Manoj K Singh, Radhakrishnan V. S, Robin Marwal, Mahesh S Dhar, Kalaiarasan Ponnusamy, Meena Datta, Hemlata Lall, Hema Gogia, Preeti Madan, Sandhya Kabra, Sujeet K Singh, Partha Rakshit |
| EPI_ISL_2460230, EPI_ISL_2460231, EPI_ISL_2460232, EPI_ISL_2460233 | National Centre for Disease Control (NCDC) Biotechnology Division, Delhi | NCDC Delhi, Biotechnology Division INSACOG | Uma Sharma, Manoj K Singh, Radhakrishnan V. S, Robin Marwal, Mahesh S Dhar, Kalaiarasan Ponnusamy, Meena Datta, Priyanka Singh, Hemlata Lall, Hema Gogia, Preeti Madan, Sandhya Kabra, Sujeet K Singh, Partha Rakshit |
| EPI_ISL_2460424, EPI_ISL_2460434, EPI_ISL_2460436, EPI_ISL_2460448, EPI_ISL_2460449 | National Centre for Disease Control (NCDC) Biotechnology Division, Delhi | NCDC Delhi, Biotechnology Division INSACOG | Radhakrishnan V. S, Robin Marwal, Mahesh S Dhar, Kalaiarasan Ponnusamy, Meena Datta, Priyanka Singh, Uma Sharma, Hemlata Lall, Manoj K Singh, Hema Gogia, Preeti Madan, Sandhya Kabra, Sujeet K Singh, Partha Rakshit |
| EPI_ISL_2460474 | National Centre for Disease Control (NCDC) Biotechnology Division, Delhi | NCDC Delhi, Biotechnology Division INSACOG | Robin Marwal, Mahesh S Dhar, Kalaiarasan Ponnusamy, Meena Datta, Priyanka Singh, Uma Sharma, Hemlata Lall, Manoj K Singh, Radhakrishnan V. S, Hema Gogia, Preeti Madan, Sandhya Kabra, Sujeet K Singh, Partha Rakshit |
| EPI_ISL_2483043, EPI_ISL_2483044, EPI_ISL_2483045, EPI_ISL_2483046, EPI_ISL_2483047, EPI_ISL_2483048, EPI_ISL_2483049, EPI_ISL_2483050, EPI_ISL_2483051, EPI_ISL_2483052, EPI_ISL_2483053, EPI_ISL_2483054, EPI_ISL_2483055, EPI_ISL_2483056, EPI_ISL_2483057, EPI_ISL_2497899, EPI_ISL_2497900, EPI_ISL_2497901, EPI_ISL_2497902, EPI_ISL_2497903, EPI_ISL_2497904, EPI_ISL_2497905 | | | |
| see above | Indian Council of Medical Research-National Institute of Virology, Microbial Containment Complex | Indian Council of Medical Research-National Institute of Virology, Microbial Containment Complex | Pragya D Yadav |
| EPI_ISL_2503370 | INSACOG-WB | National Institute of Biomedical Genomics - INSACOG | Arindam Maitra, Bhaswati Bandyopadhyay, Nidhan Kumar Biswas, Tamal Ghosh, Sreedhar Chinnaswamy, Ajay Chakraborti, Saumitra Das |
| EPI_ISL_2504231, EPI_ISL_2504232, | National Centre for Disease Control (NCDC) Biotechnology | NCDC Delhi, Biotechnology Division INSACOG | Mahesh S Dhar, Kalaiarasan Ponnusamy, Meena Datta, Priyanka Singh, Uma Sharma, Manoj K Singh, Radhakrishnan V. S, Robin Marwal, Hemlata Lall, |

| | | | |
|---|---|---|---|
| EPI_ISL_2504233, EPI_ISL_2504234, EPI_ISL_2504235, EPI_ISL_2504236, EPI_ISL_2504237 | Division, Delhi | | Hema Gogia, Preeti Madan, Sandhya Kabra, Sujeet K Singh, Partha Rakshit |
| EPI_ISL_2504245 | National Centre for Disease Control (NCDC) Biotechnology Division, Delhi | NCDC Delhi, Biotechnology Division INSACOG | Kalaiarasan Ponnusamy, Meena Datta, Priyanka Singh, Uma Sharma, Manoj K Singh, Radhakrishnan V. S, Robin Marwal, Mahesh S Dhar, Hemlata Lall, Hema Gogia, Preeti Madan, Sandhya Kabra, Sujeet K Singh, Partha Rakshit |
| EPI_ISL_2504324 | National Centre for Disease Control (NCDC) Biotechnology Division, Delhi | NCDC Delhi, Biotechnology Division INSACOG | Priyanka Singh, Uma Sharma, Manoj K Singh, Radhakrishnan V. S, Robin Marwal, Mahesh S Dhar, Kalaiarasan Ponnusamy, Meena Datta, Hemlata Lall, Hema Gogia, Preeti Madan, Sandhya Kabra, Sujeet K Singh, Partha Rakshit |
| EPI_ISL_2504325, EPI_ISL_2504326, EPI_ISL_2504327, EPI_ISL_2504328, EPI_ISL_2504329, EPI_ISL_2504330, EPI_ISL_2504331, EPI_ISL_2504332, EPI_ISL_2504333, EPI_ISL_2504334, EPI_ISL_2504335, EPI_ISL_2504336, EPI_ISL_2504337, EPI_ISL_2504338, EPI_ISL_2504339 | | | |
| see above | National Centre for Disease Control (NCDC) Biotechnology Division, Delhi | NCDC Delhi, Biotechnology Division INSACOG | Uma Sharma, Manoj K Singh, Radhakrishnan V. S, Robin Marwal, Mahesh S Dhar, Kalaiarasan Ponnusamy, Meena Datta, Priyanka Singh, Hemlata Lall, Hema Gogia, Preeti Madan, Sandhya Kabra, Sujeet K Singh, Partha Rakshit |
| EPI_ISL_2504833, EPI_ISL_2504834, EPI_ISL_2504835, EPI_ISL_2504836, EPI_ISL_2504837, EPI_ISL_2504838, EPI_ISL_2504839, EPI_ISL_2504840, EPI_ISL_2504841, EPI_ISL_2504842, EPI_ISL_2504843, EPI_ISL_2504844, EPI_ISL_2504845, EPI_ISL_2504846, EPI_ISL_2504847, EPI_ISL_2504848, EPI_ISL_2504849, EPI_ISL_2504850, EPI_ISL_2504851, EPI_ISL_2504852, EPI_ISL_2504853, EPI_ISL_2504854, EPI_ISL_2504855, EPI_ISL_2504856, EPI_ISL_2504857, EPI_ISL_2504858, EPI_ISL_2504859, EPI_ISL_2504860, EPI_ISL_2504861, EPI_ISL_2504862 | | | |
| see above | National Centre for Disease Control (NCDC) Biotechnology Division, Delhi | NCDC Delhi, Biotechnology Division INSACOG | Radhakrishnan V. S, Robin Marwal, Mahesh S Dhar, Kalaiarasan Ponnusamy, Meena Datta, Priyanka Singh, Uma Sharma, Hemlata Lall, Manoj K Singh, Hema Gogia, Preeti Madan, Sandhya Kabra, Sujeet K Singh, Partha Rakshit |
| EPI_ISL_2521748, EPI_ISL_2521749 | Indian Council of Medical Research-National Institute of Virology, Microbial Containment Complex | Indian Council of Medical Research-National Institute of Virology, Microbial Containment Complex | Pragya D Yadav |
| EPI_ISL_413522 | Indian Council of Medical Research - National Institute of Virology | National Influenza Center, Indian Council of Medical Research - National Institute of Virology | Potdar V, Yadav PD, Choudhary ML, Shete-Aich A |
| EPI_ISL_413523 | Indian Council of Medical Research-National Institute of Virology | National Influenza Center, Indian Council of Medical Research-National Institute of Virology | Potdar V, Yadav PD, Choudhary ML, Shete-Aich A |
| EPI_ISL_435077, EPI_ISL_435085, EPI_ISL_435086 | National Centre for Disease control (NCDC), CSIR-Institute of Genomics and Integrative Biology (CSIR-IGIB) | NCDC/CSIR-IGIB | Pramod Kumar, Rajesh Pandey, Pooja Sharma, Mahesh Dhar, Vivekanand A, Bharathram Uppili, Himanshu Vashisht, Saruchi Wadhwa, Nishu Tyagi, Uma Sharma, Priyanka Singh, Hemlata Lall, Meena Datta, Poonam Gupta, Nidhi Saini, Aarti Tewari, Bibhash Nandi, Dhirendra Kumar, Satyabrata Bag, Varun Jaiswal, Hema Gogia, Preeti Madan, Simrita Singh, Prateek Singh, Debasis Dash, Mitali Mukerji, Manju Bala, Sandhya Kabra, Sujeet Singh, Mohammed Faruq, Anurag Agrawal, Partha Rakshit |
| EPI_ISL_436438, EPI_ISL_436442, EPI_ISL_436444, EPI_ISL_436446 | National Centre for Disease control (NCDC) | NCDC/CSIR-IGIB | Pramod Kumar#, Rajesh Pandey#, Pooja Sharma, Mahesh S Dhar, Vivekanand A, Bharathram Uppili, Himanshu Vashisht, Saruchi Wadhwa, Nishu Tyagi, Uma Sharma, Priyanka Singh, Hemlata Lall, Meena Datta, Poonam Gupta, Nidhi Saini, Aarti Tewari, Bibhash Nandi, Dhirendra Kumar, Satyabrata Bag, Varun Jaiswal, Hema Gogia, Preeti Madan, Simrita Singh, Prateek Singh, Debasis Dash, Mitali Mukerji, Manju Bala, Sandhya Kabra, Sujeet Singh, Mohammed Faruq, Anurag Agrawal*, Partha Rakshit* |
| EPI_ISL_450321 | NIV Pune | CSIR-Centre for Cellular and Molecular Biology | Dr V A Potdar, Dr ML Choudhary,Dr Priya Abraham,V. Vipat, S. Jadhav, U. Saha, H. Kengle, A. Awhale, A. Jagtap, A. Gondhalikar, V Malik, N Srivastava, S. Digraskar, P. Malsane, S. Hundekar, K. Patel, Yogesh Balakartik, M. Kakade, S. Jadhav, R. Gunjikar, V. Awtade, S. Bhorekar, P Shinde, S. Salve, B. Minhas S. Bharadwaj, H Kaushal Y. Gurav, S. Tomar,Payel Mukherjee, Sofia Banu, Priya Singh, Dhiviya Vedagiri, Divya Gupta, Vishal Sah, Santosh Kumar Kuncha, Krishnan Harinivas Harshan, Archana Bharadwaj Siva, Karthik Bharadwaj Tallapaka, Shagufta Khan, Lamuk Zaveri, Namami Gaur, Sakshi Shambhavi, Tulasi Nagabandi, Purushotham Vodnala,G. Aditya Kumar, Koushick Sivakumar, Pooja Ramesh Gupta, Rajan Kumar Jha, Shraddha Vijay Lahoti, Deepak Kumar, Devi Prasad Vijayashankara, Disha Nanda, Divya Das, Jotin Gogoi, Manish |
| EPI_ISL_450322 | NIV Pune | CSIR-Centre for Cellular and Molecular Biology | Dr V A Potdar, Dr ML Choudhary,Dr Priya Abraham,V. Vipat, S. Jadhav, U. Saha, H. Kengle, A. Awhale, A. Jagtap, A. Gondhalikar, V Malik, N Srivastava, S. Digraskar, P. Malsane, S. Hundekar, K. Patel, Yogesh Balakartik, M. Kakade, S. Jadhav, R. Gunjikar, V. Awtade, S. Bhorekar, P Shinde, S. Salve, B. Minhas S. Bharadwaj, H Kaushal Y. Gurav, S. Tomar,Sofia Banu, Payel Mukherjee, Priya Singh, Dhiviya Vedagiri, Divya Gupta, Vishal Sah, Santosh Kumar Kuncha, Krishnan Harinivas Harshan, Archana Bharadwaj Siva, Karthik Bharadwaj Tallapaka, Shagufta Khan, Lamuk Zaveri, Namami Gaur, Sakshi Shambhavi, Tulasi Nagabandi, Purushotham Vodnala, Disha Nanda, Divya Das, Jotin Gogoi, Manish Bhattacharjee, Ravi Prasad Mukku, Renu Sudhakar, Somesh Gorde, Gangumala Srinivas Reddy, Sujoy Deb, Swati Bayyana, Zeba Rizvi, Rakesh K Mishra |
| EPI_ISL_450323 | NIV Pune | CSIR-Centre for Cellular and Molecular Biology | Dr V A Potdar, Dr ML Choudhary,Dr Priya Abraham,V. Vipat, S. Jadhav, U. Saha, H. Kengle, A. Awhale, A. Jagtap, A. Gondhalikar, V Malik, N Srivastava, S. Digraskar, P. Malsane, S. Hundekar, K. Patel, Yogesh Balakartik, M. Kakade, S. Jadhav, R. Gunjikar, V. Awtade, S. Bhorekar, P Shinde, S. Salve, B. Minhas S. Bharadwaj, H Kaushal Y. Gurav, S. Tomar,Payel Mukherjee, Sofia Banu, Priya Singh, Dhiviya Vedagiri, Divya Gupta, Vishal Sah, Santosh Kumar Kuncha, Krishnan Harinivas Harshan, Archana Bharadwaj Siva, Karthik Bharadwaj Tallapaka, Shagufta Khan, Lamuk Zaveri, Namami Gaur, Sakshi Shambhavi, Tulasi Nagabandi, Purushotham Vodnala,G. Aditya Kumar, Koushick Sivakumar, Pooja Ramesh Gupta, Rajan Kumar Jha, Shraddha Vijay Lahoti, Deepak Kumar, Devi Prasad Vijayashankara, Disha Nanda, Divya Das, Jotin Gogoi, Manish |
| EPI_ISL_450324 | NIV Pune | CSIR-Centre for Cellular and Molecular Biology | Dr V A Potdar, Dr ML Choudhary,Dr Priya Abraham,V. Vipat, S. Jadhav, U. Saha, H. Kengle, A. Awhale, A. Jagtap, A. Gondhalikar, V Malik, N Srivastava, S. Digraskar, P. Malsane, S. Hundekar, K. Patel, Yogesh Balakartik, M. Kakade, S. Jadhav, R. Gunjikar, V. Awtade, S. Bhorekar, P Shinde, S. Salve, B. Minhas S. Bharadwaj, H Kaushal Y. Gurav, S. Tomar,Sofia Banu, Payel Mukherjee, Priya Singh, Dhiviya Vedagiri, Divya Gupta, Vishal Sah, Santosh Kumar Kuncha, Krishnan Harinivas Harshan, Archana Bharadwaj Siva, Karthik Bharadwaj Tallapaka, Shagufta Khan, Lamuk Zaveri, Namami Gaur, Sakshi Shambhavi, Tulasi Nagabandi, Purushotham Vodnala, Disha Nanda, Divya Das, Jotin Gogoi, Manish Bhattacharjee, Ravi Prasad Mukku, Renu Sudhakar, Somesh Gorde, Gangumala Srinivas Reddy, Sujoy Deb, Swati Bayyana, Zeba Rizvi, Rakesh K Mishra |
| EPI_ISL_450325 | NIV Pune | CSIR-Centre for Cellular and Molecular Biology | Dr V A Potdar, Dr ML Choudhary,Dr Priya Abraham,V. Vipat, S. Jadhav, U. Saha, H. Kengle, A. Awhale, A. Jagtap, A. Gondhalikar, V Malik, N Srivastava, S. Digraskar, P. Malsane, S. Hundekar, K. Patel, Yogesh Balakartik, M. Kakade, S. Jadhav, R. Gunjikar, V. Awtade, S. Bhorekar, P Shinde, S. Salve, B. Minhas S. Bharadwaj, H Kaushal Y. Gurav, S. Tomar,Payel Mukherjee, Sofia Banu, Priya Singh, Dhiviya Vedagiri, Divya Gupta, Vishal Sah, Santosh Kumar Kuncha, Krishnan Harinivas Harshan, Archana Bharadwaj Siva, Karthik Bharadwaj Tallapaka, Shagufta Khan, Lamuk Zaveri, Namami Gaur, Sakshi Shambhavi, Tulasi Nagabandi, Purushotham Vodnala,G. Aditya Kumar, Koushick Sivakumar, Pooja Ramesh Gupta, Rajan Kumar Jha, Shraddha Vijay Lahoti, Deepak Kumar, Devi Prasad Vijayashankara, Disha Nanda, Divya Das, Jotin Gogoi, Manish |
| EPI_ISL_452192, EPI_ISL_452193, EPI_ISL_452194, EPI_ISL_452195, EPI_ISL_452196, EPI_ISL_452197, EPI_ISL_452198, EPI_ISL_452199, EPI_ISL_452200, EPI_ISL_452201, EPI_ISL_452202, EPI_ISL_452203, EPI_ISL_452204, EPI_ISL_452205, EPI_ISL_452206, EPI_ISL_452207, EPI_ISL_452208, EPI_ISL_452209, EPI_ISL_452210, EPI_ISL_452211, EPI_ISL_452212, EPI_ISL_452213, EPI_ISL_452214, EPI_ISL_452215, EPI_ISL_452216, EPI_ISL_452217, EPI_ISL_454521, EPI_ISL_454522, EPI_ISL_454523, EPI_ISL_454524, EPI_ISL_454525, EPI_ISL_454526, EPI_ISL_454527, EPI_ISL_454528, EPI_ISL_454529, EPI_ISL_454530, EPI_ISL_454531, EPI_ISL_454532, EPI_ISL_454533, EPI_ISL_454534, EPI_ISL_454535, EPI_ISL_454536, EPI_ISL_454537, EPI_ISL_454538, EPI_ISL_454539, EPI_ISL_454540, EPI_ISL_454541, EPI_ISL_454542, EPI_ISL_454543, EPI_ISL_454544, EPI_ISL_454545, EPI_ISL_454546, EPI_ISL_454547, EPI_ISL_454548, EPI_ISL_454549, EPI_ISL_454550, EPI_ISL_454551, EPI_ISL_454552, EPI_ISL_454553, EPI_ISL_454554, EPI_ISL_454555, EPI_ISL_454556, EPI_ISL_454557, EPI_ISL_454558, EPI_ISL_454559, EPI_ISL_454560, EPI_ISL_454561, EPI_ISL_454562, EPI_ISL_454563, EPI_ISL_454564, EPI_ISL_454565, EPI_ISL_454566, EPI_ISL_454567, EPI_ISL_454568, EPI_ISL_454569, EPI_ISL_454570, EPI_ISL_479493, EPI_ISL_479494, EPI_ISL_479495, EPI_ISL_479496, EPI_ISL_479497, EPI_ISL_479498, EPI_ISL_479499, EPI_ISL_479500, EPI_ISL_479501, EPI_ISL_479502, EPI_ISL_479503, EPI_ISL_479504, EPI_ISL_479505, EPI_ISL_479506, EPI_ISL_479507, EPI_ISL_479508, EPI_ISL_479509, EPI_ISL_479510, EPI_ISL_479511, EPI_ISL_479512, EPI_ISL_479513, EPI_ISL_479514, EPI_ISL_479515, EPI_ISL_479516, EPI_ISL_479517, EPI_ISL_479518, EPI_ISL_479519, EPI_ISL_479520, EPI_ISL_479521, EPI_ISL_479522, EPI_ISL_479523, EPI_ISL_479524, EPI_ISL_479525, EPI_ISL_479526, EPI_ISL_479527, EPI_ISL_479528, EPI_ISL_479529, EPI_ISL_479530, EPI_ISL_479531, EPI_ISL_479532, EPI_ISL_479533, EPI_ISL_479534, EPI_ISL_479535, EPI_ISL_479536, EPI_ISL_479537, EPI_ISL_479538, EPI_ISL_479539, EPI_ISL_479540, EPI_ISL_479541, EPI_ISL_479542, EPI_ISL_479543, EPI_ISL_479544, EPI_ISL_479545, EPI_ISL_479546, EPI_ISL_479547, EPI_ISL_479548, EPI_ISL_479549, EPI_ISL_479550, EPI_ISL_479551, EPI_ISL_479552, EPI_ISL_479553, EPI_ISL_479554, EPI_ISL_479555, EPI_ISL_479556, EPI_ISL_479557, EPI_ISL_479558, EPI_ISL_479559, EPI_ISL_479560, EPI_ISL_479561, EPI_ISL_479562, EPI_ISL_479563, EPI_ISL_479564, EPI_ISL_479565, EPI_ISL_479566, EPI_ISL_479567, EPI_ISL_479568, EPI_ISL_479569, EPI_ISL_479570, EPI_ISL_479571, EPI_ISL_479572, EPI_ISL_479573, EPI_ISL_479657, EPI_ISL_479658, EPI_ISL_479659, EPI_ISL_479660, EPI_ISL_479661, EPI_ISL_479776 | | | |
| see above | NIV Influenza | NIV Influenza | Potdar V |
| EPI_ISL_496518, EPI_ISL_496519, EPI_ISL_496520 | Armed Forces Medical College | National Centre For Cell Science | Dhiraj Paul, Kunal Jani, Radha Chauhan, Janesh Kumar, Vasudevan Seshadri, Girdhari Lal, Rajesh Karyakarte, Suvarna Joshi, Murlidhar Tambe, Sourav Sen, Santosh Karade, Kavita Bala Anand, Shelinder Pal Singh Shergill, Rajiv Mohan Gupta, Manoj Kumar Bhat, Arvind Sahu, Maharashtra COVID-19 Study Group, DBT's PAN-INDIA 1000 SARS-CoV2 RNA genome sequencing consortium, Yogesh S Shouche |
| EPI_ISL_496521, EPI_ISL_496522, EPI_ISL_496523 | B.J. Govt. Medical College | National Centre For Cell Science | Dhiraj Paul, Kunal Jani, Radha Chauhan, Janesh Kumar, Vasudevan Seshadri, Girdhari Lal, Rajesh Karyakarte, Suvarna Joshi, Murlidhar Tambe, Sourav Sen, Santosh Karade, Kavita Bala Anand, Shelinder Pal Singh Shergill, Rajiv Mohan Gupta, Manoj Kumar Bhat, Arvind Sahu, Maharashtra COVID-19 Study Group, DBT's PAN-INDIA 1000 SARS-CoV2 RNA genome sequencing consortium, Yogesh S Shouche |
| EPI_ISL_496524, EPI_ISL_496525, EPI_ISL_496526 | National Centre For Cell Science | National Centre For Cell Science | Dhiraj Paul, Kunal Jani, Radha Chauhan, Janesh Kumar, Vasudevan Seshadri, Girdhari Lal, Rajesh Karyakarte, Suvarna Joshi, Murlidhar Tambe, Sourav Sen, Santosh Karade, Kavita Bala Anand, Shelinder Pal Singh Shergill, Rajiv Mohan Gupta, Manoj Kumar Bhat, Arvind Sahu, Maharashtra COVID-19 |

| | | | |
|---|---|---|---|
| | | | Study Group, DBT's PAN-INDIA 1000 SARS-CoV2 RNA genome sequencing consortium, Yogesh S Shouche |
| EPI_ISL_496527, EPI_ISL_496528 | B.J. Govt. Medical College | National Centre For Cell Science | Dhiraj Paul, Kunal Jani, Radha Chauhan, Janesh Kumar, Vasudevan Seshadri, Girdhari Lal, Rajesh Karyakarte, Suvarna Joshi, Murlidhar Tambe, Sourav Sen, Santosh Karade, Kavita Bala Anand, Shelinder Pal Singh Shergill, Rajiv Mohan Gupta, Manoj Kumar Bhat, Arvind Sahu, Maharashtra COVID-19 Study Group, DBT's PAN-INDIA 1000 SARS-CoV2 RNA genome sequencing consortium, Yogesh S Shouche |
| EPI_ISL_496529 | Armed Forces Medical College | National Centre For Cell Science | Dhiraj Paul, Kunal Jani, Radha Chauhan, Janesh Kumar, Vasudevan Seshadri, Girdhari Lal, Rajesh Karyakarte, Suvarna Joshi, Murlidhar Tambe, Sourav Sen, Santosh Karade, Kavita Bala Anand, Shelinder Pal Singh Shergill, Rajiv Mohan Gupta, Manoj Kumar Bhat, Arvind Sahu, Maharashtra COVID-19 Study Group, DBT's PAN-INDIA 1000 SARS-CoV2 RNA genome sequencing consortium, Yogesh S Shouche |
| EPI_ISL_496530 | B.J. Govt. Medical College | National Centre For Cell Science | Dhiraj Paul, Kunal Jani, Radha Chauhan, Janesh Kumar, Vasudevan Seshadri, Girdhari Lal, Rajesh Karyakarte, Suvarna Joshi, Murlidhar Tambe, Sourav Sen, Santosh Karade, Kavita Bala Anand, Shelinder Pal Singh Shergill, Rajiv Mohan Gupta, Manoj Kumar Bhat, Arvind Sahu, Maharashtra COVID-19 Study Group, DBT's PAN-INDIA 1000 SARS-CoV2 RNA genome sequencing consortium, Yogesh S Shouche |
| EPI_ISL_496531, EPI_ISL_496532 | National Centre For Cell Science | National Centre For Cell Science | Dhiraj Paul, Kunal Jani, Radha Chauhan, Janesh Kumar, Vasudevan Seshadri, Girdhari Lal, Rajesh Karyakarte, Suvarna Joshi, Murlidhar Tambe, Sourav Sen, Santosh Karade, Kavita Bala Anand, Shelinder Pal Singh Shergill, Rajiv Mohan Gupta, Manoj Kumar Bhat, Arvind Sahu, Maharashtra COVID-19 Study Group, DBT's PAN-INDIA 1000 SARS-CoV2 RNA genome sequencing consortium, Yogesh S Shouche |
| EPI_ISL_496533 | Armed Forces Medical College | National Centre For Cell Science | Dhiraj Paul, Kunal Jani, Radha Chauhan, Janesh Kumar, Vasudevan Seshadri, Girdhari Lal, Rajesh Karyakarte, Suvarna Joshi, Murlidhar Tambe, Sourav Sen, Santosh Karade, Kavita Bala Anand, Shelinder Pal Singh Shergill, Rajiv Mohan Gupta, Manoj Kumar Bhat, Arvind Sahu, Maharashtra COVID-19 Study Group, DBT's PAN-INDIA 1000 SARS-CoV2 RNA genome sequencing consortium, Yogesh S Shouche |
| EPI_ISL_496534 | B.J. Govt. Medical College | National Centre For Cell Science | Dhiraj Paul, Kunal Jani, Radha Chauhan, Janesh Kumar, Vasudevan Seshadri, Girdhari Lal, Rajesh Karyakarte, Suvarna Joshi, Murlidhar Tambe, Sourav Sen, Santosh Karade, Kavita Bala Anand, Shelinder Pal Singh Shergill, Rajiv Mohan Gupta, Manoj Kumar Bhat, Arvind Sahu, Maharashtra COVID-19 Study Group, DBT's PAN-INDIA 1000 SARS-CoV2 RNA genome sequencing consortium, Yogesh S Shouche |
| EPI_ISL_496535, EPI_ISL_496536 | National Centre For Cell Science | National Centre For Cell Science | Dhiraj Paul, Kunal Jani, Radha Chauhan, Janesh Kumar, Vasudevan Seshadri, Girdhari Lal, Rajesh Karyakarte, Suvarna Joshi, Murlidhar Tambe, Sourav Sen, Santosh Karade, Kavita Bala Anand, Shelinder Pal Singh Shergill, Rajiv Mohan Gupta, Manoj Kumar Bhat, Arvind Sahu, Maharashtra COVID-19 Study Group, DBT's PAN-INDIA 1000 SARS-CoV2 RNA genome sequencing consortium, Yogesh S Shouche |
| EPI_ISL_496537, EPI_ISL_496538, EPI_ISL_496539, EPI_ISL_496540, EPI_ISL_496541, EPI_ISL_496542, EPI_ISL_496543, EPI_ISL_496544, EPI_ISL_496545 | Armed Forces Medical College | National Centre For Cell Science | Dhiraj Paul, Kunal Jani, Radha Chauhan, Janesh Kumar, Vasudevan Seshadri, Girdhari Lal, Rajesh Karyakarte, Suvarna Joshi, Murlidhar Tambe, Sourav Sen, Santosh Karade, Kavita Bala Anand, Shelinder Pal Singh Shergill, Rajiv Mohan Gupta, Manoj Kumar Bhat, Arvind Sahu, Maharashtra COVID-19 Study Group, DBT's PAN-INDIA 1000 SARS-CoV2 RNA genome sequencing consortium, Yogesh S Shouche |
| EPI_ISL_496546, EPI_ISL_496547, EPI_ISL_496548, EPI_ISL_496549, EPI_ISL_496550, EPI_ISL_496551, EPI_ISL_496552, EPI_ISL_496553, EPI_ISL_496554 | B.J. Govt. Medical College | National Centre For Cell Science | Dhiraj Paul, Kunal Jani, Radha Chauhan, Janesh Kumar, Vasudevan Seshadri, Girdhari Lal, Rajesh Karyakarte, Suvarna Joshi, Murlidhar Tambe, Sourav Sen, Santosh Karade, Kavita Bala Anand, Shelinder Pal Singh Shergill, Rajiv Mohan Gupta, Manoj Kumar Bhat, Arvind Sahu, Maharashtra COVID-19 Study Group, DBT's PAN-INDIA 1000 SARS-CoV2 RNA genome sequencing consortium, Yogesh S Shouche |
| EPI_ISL_496555, EPI_ISL_496556, EPI_ISL_496557, EPI_ISL_496558, EPI_ISL_496559, EPI_ISL_496560, EPI_ISL_496561, EPI_ISL_496562, EPI_ISL_496563, EPI_ISL_496564, EPI_ISL_496565, EPI_ISL_496566, EPI_ISL_496567, EPI_ISL_496568, EPI_ISL_496569, EPI_ISL_496570, EPI_ISL_496571, EPI_ISL_496572, EPI_ISL_496573, EPI_ISL_496574, EPI_ISL_496575, EPI_ISL_496576, EPI_ISL_496577, EPI_ISL_496578, EPI_ISL_496579, EPI_ISL_496580, EPI_ISL_496581, EPI_ISL_496582, EPI_ISL_496583, EPI_ISL_496584, EPI_ISL_496585, EPI_ISL_496586, EPI_ISL_496587 | | | |
| see above | National Centre For Cell Science | National Centre For Cell Science | Dhiraj Paul, Kunal Jani, Radha Chauhan, Janesh Kumar, Vasudevan Seshadri, Girdhari Lal, Rajesh Karyakarte, Suvarna Joshi, Murlidhar Tambe, Sourav Sen, Santosh Karade, Kavita Bala Anand, Shelinder Pal Singh Shergill, Rajiv Mohan Gupta, Manoj Kumar Bhat, Arvind Sahu, Maharashtra COVID-19 Study Group, DBT's PAN-INDIA 1000 SARS-CoV2 RNA genome sequencing consortium, Yogesh S Shouche |
| EPI_ISL_496602 | Armed Forces Medical College | National Centre For Cell Science | Dhiraj Paul, Kunal Jani, Radha Chauhan, Janesh Kumar, Vasudevan Seshadri, Girdhari Lal, Rajesh Karyakarte, Suvarna Joshi, Murlidhar Tambe, Sourav Sen, Santosh Karade, Kavita Bala Anand, Shelinder Pal Singh Shergill, Rajiv Mohan Gupta, Manoj Kumar Bhat, Arvind Sahu, Maharashtra COVID-19 Study Group, DBT's PAN-INDIA 1000 SARS-CoV2 RNA genome sequencing consortium, Yogesh S Shouche |
| EPI_ISL_497873, EPI_ISL_497874, EPI_ISL_497875, EPI_ISL_497876, EPI_ISL_497877, EPI_ISL_497878, EPI_ISL_497879 | National Centre For Cell Science | National Centre For Cell Science | Dhiraj Paul, Kunal Jani, Radha Chauhan, Janesh Kumar, Vasudevan Seshadri, Girdhari Lal, Rajesh Karyakarte, Suvarna Joshi, Murlidhar Tambe, Sourav Sen, Santosh Karade, Kavita Bala Anand, Shelinder Pal Singh Shergill, Rajiv Mohan Gupta, Manoj Kumar Bhat, Arvind Sahu, Maharashtra COVID-19 Study Group, DBT's PAN-INDIA 1000 SARS-CoV2 RNA genome sequencing consortium, Yogesh S Shouche |
| EPI_ISL_497880, EPI_ISL_497881, EPI_ISL_497882, EPI_ISL_497883, EPI_ISL_497884, EPI_ISL_497885, EPI_ISL_497886, EPI_ISL_497887 | Armed Forces Medical College | National Centre For Cell Science | Dhiraj Paul, Kunal Jani, Radha Chauhan, Janesh Kumar, Vasudevan Seshadri, Girdhari Lal, Rajesh Karyakarte, Suvarna Joshi, Murlidhar Tambe, Sourav Sen, Santosh Karade, Kavita Bala Anand, Shelinder Pal Singh Shergill, Rajiv Mohan Gupta, Manoj Kumar Bhat, Arvind Sahu, Maharashtra COVID-19 Study Group, DBT's PAN-INDIA 1000 SARS-CoV2 RNA genome sequencing consortium, Yogesh S Shouche |
| EPI_ISL_497888, EPI_ISL_497889, EPI_ISL_497890, EPI_ISL_497891 | B.J. Govt. Medical College | National Centre For Cell Science | Dhiraj Paul, Kunal Jani, Radha Chauhan, Janesh Kumar, Vasudevan Seshadri, Girdhari Lal, Rajesh Karyakarte, Suvarna Joshi, Murlidhar Tambe, Sourav Sen, Santosh Karade, Kavita Bala Anand, Shelinder Pal Singh Shergill, Rajiv Mohan Gupta, Manoj Kumar Bhat, Arvind Sahu, Maharashtra COVID-19 Study Group, DBT's PAN-INDIA 1000 SARS-CoV2 RNA genome sequencing consortium, Yogesh S Shouche |
| EPI_ISL_508207, EPI_ISL_508208, EPI_ISL_508209, EPI_ISL_508210, EPI_ISL_508211, EPI_ISL_508212, EPI_ISL_508213, EPI_ISL_508214, EPI_ISL_508215, EPI_ISL_508216, EPI_ISL_508217, EPI_ISL_508218, EPI_ISL_508219, EPI_ISL_508220, EPI_ISL_508221, EPI_ISL_508222, EPI_ISL_508223, EPI_ISL_508224, EPI_ISL_508225, EPI_ISL_508226, EPI_ISL_508227, EPI_ISL_508228, EPI_ISL_508229, EPI_ISL_508230, EPI_ISL_508231, EPI_ISL_508232, EPI_ISL_508233, EPI_ISL_508234, EPI_ISL_508235, EPI_ISL_508236, EPI_ISL_508237, EPI_ISL_508238, EPI_ISL_508239, EPI_ISL_508240, EPI_ISL_508241, EPI_ISL_508242, EPI_ISL_508243, EPI_ISL_508244, EPI_ISL_508245, EPI_ISL_508246, EPI_ISL_508247, EPI_ISL_508248, EPI_ISL_508249, EPI_ISL_508250, EPI_ISL_508251, EPI_ISL_508252, EPI_ISL_508253, EPI_ISL_508254, EPI_ISL_508255, EPI_ISL_508256, EPI_ISL_508257, EPI_ISL_508258, EPI_ISL_508259, EPI_ISL_508260, EPI_ISL_508261, EPI_ISL_508262, EPI_ISL_508263, EPI_ISL_508264, EPI_ISL_508265, EPI_ISL_508266, EPI_ISL_508267, EPI_ISL_508268, EPI_ISL_508269, EPI_ISL_508270, EPI_ISL_508271, EPI_ISL_508272, EPI_ISL_508273, EPI_ISL_508274, EPI_ISL_508275, EPI_ISL_508276, EPI_ISL_508277, EPI_ISL_508278, EPI_ISL_508279, EPI_ISL_508280, EPI_ISL_508281, EPI_ISL_508282, EPI_ISL_508283, EPI_ISL_508284, EPI_ISL_508285, EPI_ISL_508286 | | | |
| see above | Government Medical College | National Institute of Biomedical Genomics | Arindam Maitra, Jyoti Iravane, Dhaval Khatri, Maitrik Dave, Saumitra Das |
| EPI_ISL_508423, EPI_ISL_508424, EPI_ISL_508425, EPI_ISL_508426, EPI_ISL_508427, EPI_ISL_508429, EPI_ISL_508430, EPI_ISL_508431, EPI_ISL_508432, EPI_ISL_508433, EPI_ISL_508435, EPI_ISL_508436, EPI_ISL_508437, EPI_ISL_508438, EPI_ISL_508439, EPI_ISL_508440 | | | |
| see above | Mahatma Gandhi Institute of Medical Sciences | National Institute of Biomedical Genomics | Arindam Maitra, Vijayshri Deotale, Rahul Narang, Deepashri Maraskolhe, Saumitra Das |
| EPI_ISL_511923, EPI_ISL_511924, EPI_ISL_511925, EPI_ISL_511926, EPI_ISL_511927, EPI_ISL_511929 | Mahatma Gandhi Institute of Medical Sciences | National Institute of Biomedical Genomics - DBT's PAN-INDIA 1000 SARS--CoV-2 RNA Genome Sequencing Consortium | Arindam Maitra, Vijayshri Deotale, Rahul Narang, Deepashri Maraskolhe, Saumitra Das |
| EPI_ISL_511930, EPI_ISL_511931, EPI_ISL_511932, EPI_ISL_511933, EPI_ISL_511934, EPI_ISL_511935, EPI_ISL_511936, EPI_ISL_511937, EPI_ISL_511938, EPI_ISL_511939, EPI_ISL_511940, EPI_ISL_511941 | | | |
| see above | Government Medical College | National Institute of Biomedical Genomics - DBT's PAN-INDIA 1000 SARS--CoV-2 RNA Genome Sequencing Consortium | Arindam Maitra, Jyoti Iravane, Dhaval Khatri, Maitrik Dave, Saumitra Das |
| EPI_ISL_522437 | Mahatma Gandhi Institute of Medical Sciences | National Institute of Biomedical Genomics - DBT's PAN-INDIA 1000 SARS--CoV-2 RNA Genome Sequencing Consortium | Arindam Maitra, Vijayshri Deotale, Rahul Narang, Deepashri Maraskolhe, Saumitra Das |
| EPI_ISL_528419, EPI_ISL_528420, EPI_ISL_528421, EPI_ISL_528422, EPI_ISL_528423, EPI_ISL_528424 | TNMC & BYL NAIR CH. HOSPITAL | Institute of Genomics and Integrative Biology - Council of Scientific and Industrial Research | Rajesh Pandey, Jayanthi Shastri, Akshay Kanakan, Vivekanand A, Janani Srinivasa Vasudevan, Ranjeet Maurya, Sachee Agrawal, Nirjhar Chatterjee, Swapneil Parikh, Manish Pathak, Subrat Thanapati, Jasmina Savak, Suresh Poojari, Mahesh Sangar, Amol Borse, Shweta Kawankar, Vasil Nachan, Mayuresh Vishwanathan, Shruthi Sachidanandan, Shrutika Pophale, Utkarsha Yelve |
| EPI_ISL_528425, EPI_ISL_528426 | P. D. Hinduja Hospital and Medical Research Centre | Institute of Genomics and Integrative Biology - Council of Scientific and Industrial Research | Rajesh Pandey, Jayanthi Shastri, Akshay Kanakan, Vivekanand A, Janani Srinivasa Vasudevan, Ranjeet Maurya, Sachee Agrawal, Nirjhar Chatterjee, Swapneil Parikh, Manish Pathak, Subrat Thanapati, Jasmina Savak, Suresh Poojari, Mahesh Sangar, Amol Borse, Shweta Kawankar, Vasil Nachan, Mayuresh Vishwanathan, Shruthi Sachidanandan, Shrutika Pophale, Utkarsha Yelve |
| EPI_ISL_541681, EPI_ISL_541682, EPI_ISL_541683, EPI_ISL_541684, EPI_ISL_541685, EPI_ISL_541686, EPI_ISL_541687, EPI_ISL_541688, EPI_ISL_541689, EPI_ISL_541690, EPI_ISL_541691, EPI_ISL_541692, EPI_ISL_541693, EPI_ISL_541694, EPI_ISL_541695, EPI_ISL_541696, EPI_ISL_541697, EPI_ISL_541698, | | | |

| | | | |
|---|---|---|---|
| EPI_ISL_541699, EPI_ISL_541700, EPI_ISL_541701, EPI_ISL_541702, EPI_ISL_541703, EPI_ISL_541704, EPI_ISL_541705, EPI_ISL_541706, EPI_ISL_541707, EPI_ISL_541708, EPI_ISL_541709, EPI_ISL_541710, EPI_ISL_541711, EPI_ISL_541712, EPI_ISL_541713, EPI_ISL_541714, EPI_ISL_541715, EPI_ISL_541716, EPI_ISL_541717, EPI_ISL_541718, EPI_ISL_541719, EPI_ISL_541720, EPI_ISL_541721, EPI_ISL_541722, EPI_ISL_541723, EPI_ISL_541724, EPI_ISL_541725, EPI_ISL_541726, EPI_ISL_541727, EPI_ISL_541728, EPI_ISL_541729, EPI_ISL_541730, EPI_ISL_541731, EPI_ISL_541732, EPI_ISL_541733, EPI_ISL_541734, EPI_ISL_541735, EPI_ISL_541736, EPI_ISL_541737, EPI_ISL_541738, EPI_ISL_541739, EPI_ISL_541740, EPI_ISL_541741, EPI_ISL_541742, EPI_ISL_541743, EPI_ISL_541744, EPI_ISL_541745, EPI_ISL_541746, EPI_ISL_541747, EPI_ISL_541748, EPI_ISL_541749, EPI_ISL_541750, EPI_ISL_541751 | | | |
| see above | National Institute of Virology, NIV Influenza | National Institute of Virology, NIV Influenza | Potdar V |
| EPI_ISL_577639, EPI_ISL_577640, EPI_ISL_577641, EPI_ISL_577642, EPI_ISL_577643, EPI_ISL_577644, EPI_ISL_577645, EPI_ISL_577646, EPI_ISL_577647, EPI_ISL_577648, EPI_ISL_577649, EPI_ISL_577650, EPI_ISL_577651, EPI_ISL_577652, EPI_ISL_577653, EPI_ISL_577654, EPI_ISL_577655, EPI_ISL_577656, EPI_ISL_577657, EPI_ISL_577658, EPI_ISL_577659, EPI_ISL_577660, EPI_ISL_577661, EPI_ISL_577662, EPI_ISL_577663, EPI_ISL_577664, EPI_ISL_577665, EPI_ISL_577666, EPI_ISL_577667, EPI_ISL_577668, EPI_ISL_577669, EPI_ISL_577670, EPI_ISL_577671, EPI_ISL_577672, EPI_ISL_577673, EPI_ISL_577674, EPI_ISL_577675, EPI_ISL_577676, EPI_ISL_577677, EPI_ISL_577678, EPI_ISL_577679, EPI_ISL_577680, EPI_ISL_577681, EPI_ISL_577682, EPI_ISL_577683, EPI_ISL_577684, EPI_ISL_577685, EPI_ISL_577686, EPI_ISL_577687, EPI_ISL_577688, EPI_ISL_577689, EPI_ISL_577690, EPI_ISL_577691, EPI_ISL_577692, EPI_ISL_577693, EPI_ISL_577694, EPI_ISL_577695, EPI_ISL_577696, EPI_ISL_577697, EPI_ISL_577698, EPI_ISL_577699, EPI_ISL_577700, EPI_ISL_577701, EPI_ISL_577702, EPI_ISL_577703, EPI_ISL_577704, EPI_ISL_577705, EPI_ISL_577706, EPI_ISL_577707, EPI_ISL_577708, EPI_ISL_577709, EPI_ISL_577710, EPI_ISL_577711, EPI_ISL_577712, EPI_ISL_577713, EPI_ISL_577716, EPI_ISL_577717, EPI_ISL_577718, EPI_ISL_577719, EPI_ISL_577720, EPI_ISL_577721, EPI_ISL_577722, EPI_ISL_577723, EPI_ISL_577724, EPI_ISL_577725, EPI_ISL_577726, EPI_ISL_577727, EPI_ISL_577728, EPI_ISL_577729, EPI_ISL_577730, EPI_ISL_577731, EPI_ISL_577732, EPI_ISL_577733, EPI_ISL_582241 | | | |
| see above | NIV Influenza | NIV Influenza | Potdar V |
| EPI_ISL_676509, EPI_ISL_676541 | TNMC & Nair ch. Hospital | CSIR-Institute of Genomics and Integrative Biology | Rajesh Pandey, Jayanthi Shastri, Akshay Kanakan, Janani Srinivasa Vasudevan, Ranjeet Maurya, Sachee Agrawal, Nirjhar Chatterjee, Swapneil Parikh, Manish Pathak, Subrat Thanapati, Jasmina Savak, Suresh Poojari, Mahesh Sangar, Amol Borse, Shweta Kawankar, Vasil Nachan, Mayuresh Vishwanathan, Shruthi Sachidanandan, Shrutika Pophale, Utkarsha Yelve |
| EPI_ISL_699659, EPI_ISL_699660, EPI_ISL_699661, EPI_ISL_699662, EPI_ISL_699663, EPI_ISL_699664, EPI_ISL_699665, EPI_ISL_699666, EPI_ISL_699667, EPI_ISL_699668, EPI_ISL_699669, EPI_ISL_699670, EPI_ISL_699671, EPI_ISL_699672, EPI_ISL_699673, EPI_ISL_699674, EPI_ISL_699675, EPI_ISL_699676, EPI_ISL_699677, EPI_ISL_699678, EPI_ISL_699679, EPI_ISL_699680, EPI_ISL_699681, EPI_ISL_699682, EPI_ISL_699683, EPI_ISL_699684, EPI_ISL_699685, EPI_ISL_699686, EPI_ISL_699687, EPI_ISL_699688, EPI_ISL_699689, EPI_ISL_699690, EPI_ISL_699691, EPI_ISL_699692, EPI_ISL_699693, EPI_ISL_699694, EPI_ISL_699695, EPI_ISL_699696, EPI_ISL_699697, EPI_ISL_699698, EPI_ISL_699699, EPI_ISL_699700, EPI_ISL_699701, EPI_ISL_699702, EPI_ISL_699703, EPI_ISL_699704, EPI_ISL_699705, EPI_ISL_699706, EPI_ISL_699707, EPI_ISL_699708, EPI_ISL_699709, EPI_ISL_699710, EPI_ISL_699711, EPI_ISL_699712, EPI_ISL_699713, EPI_ISL_699714, EPI_ISL_699715, EPI_ISL_699716, EPI_ISL_699717, EPI_ISL_699718, EPI_ISL_699719, EPI_ISL_699720, EPI_ISL_699721, EPI_ISL_699722, EPI_ISL_699723, EPI_ISL_699724, EPI_ISL_699725, EPI_ISL_699726, EPI_ISL_699727, EPI_ISL_699728, EPI_ISL_699729, EPI_ISL_699730, EPI_ISL_699731, EPI_ISL_699733, EPI_ISL_699734, EPI_ISL_699735, EPI_ISL_699736, EPI_ISL_699737, EPI_ISL_699738, EPI_ISL_699739, EPI_ISL_699740, EPI_ISL_699741, EPI_ISL_699742, EPI_ISL_699743, EPI_ISL_699744, EPI_ISL_699745, EPI_ISL_699746, EPI_ISL_699747, EPI_ISL_699748, EPI_ISL_699749, EPI_ISL_699750, EPI_ISL_699751, EPI_ISL_699752, EPI_ISL_699753, EPI_ISL_699754, EPI_ISL_699755, EPI_ISL_699756, EPI_ISL_699757, EPI_ISL_699758, EPI_ISL_699759, EPI_ISL_699760, EPI_ISL_699761, EPI_ISL_699762, EPI_ISL_699763, EPI_ISL_699764, EPI_ISL_699765, EPI_ISL_699766, EPI_ISL_699767, EPI_ISL_699768, EPI_ISL_699769, EPI_ISL_699770, EPI_ISL_699771, EPI_ISL_699772, EPI_ISL_699773, EPI_ISL_699774, EPI_ISL_699775, EPI_ISL_699776, EPI_ISL_699777, EPI_ISL_699778, EPI_ISL_699779, EPI_ISL_699780, EPI_ISL_699781, EPI_ISL_699782, EPI_ISL_699783, EPI_ISL_699784, EPI_ISL_699785, EPI_ISL_699786, EPI_ISL_699787, EPI_ISL_699788, EPI_ISL_699789, EPI_ISL_699790, EPI_ISL_699791, EPI_ISL_699792, EPI_ISL_699793, EPI_ISL_699794, EPI_ISL_699795, EPI_ISL_699796, EPI_ISL_699797, EPI_ISL_699798, EPI_ISL_699799, EPI_ISL_699800, EPI_ISL_699801, EPI_ISL_699802, EPI_ISL_699803, EPI_ISL_699804, EPI_ISL_699805, EPI_ISL_699806, EPI_ISL_699807, EPI_ISL_699808, EPI_ISL_699809, EPI_ISL_699810, EPI_ISL_699811, EPI_ISL_699812, EPI_ISL_699813, EPI_ISL_699814, EPI_ISL_699815, EPI_ISL_699816, EPI_ISL_699817, EPI_ISL_699818, EPI_ISL_699819, EPI_ISL_699820, EPI_ISL_699821, EPI_ISL_699822, EPI_ISL_699823, EPI_ISL_699824, EPI_ISL_699825, EPI_ISL_699826, EPI_ISL_699827, EPI_ISL_699828, EPI_ISL_699829, EPI_ISL_699830, EPI_ISL_699831, EPI_ISL_699832, EPI_ISL_699833, EPI_ISL_699834, EPI_ISL_699835, EPI_ISL_699836, EPI_ISL_699837, EPI_ISL_699838, EPI_ISL_699839, EPI_ISL_699840, EPI_ISL_699841, EPI_ISL_699842, EPI_ISL_699843, EPI_ISL_699844, EPI_ISL_699845, EPI_ISL_699846, EPI_ISL_699847, EPI_ISL_699848, EPI_ISL_699849, EPI_ISL_699850, EPI_ISL_699851, EPI_ISL_699852, EPI_ISL_699853, EPI_ISL_699854, EPI_ISL_699855, EPI_ISL_699856, EPI_ISL_699857, EPI_ISL_699858, EPI_ISL_699859, EPI_ISL_699860, EPI_ISL_699861, EPI_ISL_699862, EPI_ISL_699863, EPI_ISL_699864, EPI_ISL_699865, EPI_ISL_699866, EPI_ISL_699867, EPI_ISL_699868, EPI_ISL_699869, EPI_ISL_699870, EPI_ISL_699871, EPI_ISL_699872, EPI_ISL_699873, EPI_ISL_699874, EPI_ISL_699875, EPI_ISL_699876, EPI_ISL_699877, EPI_ISL_699878, EPI_ISL_699879, EPI_ISL_699880, EPI_ISL_699881, EPI_ISL_699882, EPI_ISL_699883, EPI_ISL_699884, EPI_ISL_699885, EPI_ISL_699886, EPI_ISL_699887, EPI_ISL_699888, EPI_ISL_699889, EPI_ISL_699890, EPI_ISL_699891, EPI_ISL_699892, EPI_ISL_699893, EPI_ISL_699894, EPI_ISL_699895, EPI_ISL_699896, EPI_ISL_699897, EPI_ISL_699898, EPI_ISL_699899, EPI_ISL_699900, EPI_ISL_699901, EPI_ISL_699902, EPI_ISL_699903, EPI_ISL_699904, EPI_ISL_699905, EPI_ISL_699906, EPI_ISL_699907, EPI_ISL_699908, EPI_ISL_699909, EPI_ISL_699910, EPI_ISL_699911, EPI_ISL_699912, EPI_ISL_699913, EPI_ISL_699914, EPI_ISL_699915, EPI_ISL_699916, EPI_ISL_699917, EPI_ISL_699918, EPI_ISL_699919, EPI_ISL_699920, EPI_ISL_699921, EPI_ISL_699922, EPI_ISL_699923, EPI_ISL_699924, EPI_ISL_699925, EPI_ISL_699926, EPI_ISL_699927, EPI_ISL_699928, EPI_ISL_699929, EPI_ISL_699930, EPI_ISL_699931, EPI_ISL_699932, EPI_ISL_699933, EPI_ISL_699934, EPI_ISL_699935, EPI_ISL_699936, EPI_ISL_699937, EPI_ISL_699938, EPI_ISL_699939, EPI_ISL_699940, EPI_ISL_699941, EPI_ISL_699942, EPI_ISL_699943, EPI_ISL_699944, EPI_ISL_699945, EPI_ISL_699946, EPI_ISL_699947, EPI_ISL_699948, EPI_ISL_699949, EPI_ISL_699950, EPI_ISL_699951, EPI_ISL_699952, EPI_ISL_699953, EPI_ISL_699954, EPI_ISL_699955, EPI_ISL_699956, EPI_ISL_699957, EPI_ISL_699958, EPI_ISL_699959, EPI_ISL_699960, EPI_ISL_699961, EPI_ISL_699962, EPI_ISL_699963, EPI_ISL_699964, EPI_ISL_699965, EPI_ISL_699966, EPI_ISL_699967, EPI_ISL_699968, EPI_ISL_699969, EPI_ISL_699970, EPI_ISL_699971, EPI_ISL_699972, EPI_ISL_699973, EPI_ISL_699974, EPI_ISL_699975, EPI_ISL_699976, EPI_ISL_699977, EPI_ISL_699978, EPI_ISL_699979, EPI_ISL_699980, EPI_ISL_699981, EPI_ISL_699982, EPI_ISL_699983, EPI_ISL_699984, EPI_ISL_699985, EPI_ISL_699986, EPI_ISL_699987, EPI_ISL_699988, EPI_ISL_699989, EPI_ISL_699990, EPI_ISL_699991, EPI_ISL_699992, EPI_ISL_699993, EPI_ISL_699994, EPI_ISL_699995, EPI_ISL_699996, EPI_ISL_699997, EPI_ISL_699998, EPI_ISL_699999, EPI_ISL_700000, EPI_ISL_700002, EPI_ISL_700003, EPI_ISL_700004, EPI_ISL_700005, EPI_ISL_700006, EPI_ISL_700007, EPI_ISL_700008, EPI_ISL_700009, EPI_ISL_700010, EPI_ISL_700011, EPI_ISL_700012, EPI_ISL_700013, EPI_ISL_700014, EPI_ISL_700015, EPI_ISL_700016, EPI_ISL_700017, EPI_ISL_700018, EPI_ISL_700019, EPI_ISL_700020, EPI_ISL_700021, EPI_ISL_700022, EPI_ISL_700023, EPI_ISL_700024, EPI_ISL_700025, EPI_ISL_700026, EPI_ISL_700027, EPI_ISL_700028, EPI_ISL_700029, EPI_ISL_700030, EPI_ISL_700031, EPI_ISL_700032, EPI_ISL_700033, EPI_ISL_700034, EPI_ISL_700035, EPI_ISL_700036, EPI_ISL_700037, EPI_ISL_700038, EPI_ISL_700039, EPI_ISL_700040, EPI_ISL_700041, EPI_ISL_700042, EPI_ISL_700043, EPI_ISL_700044, EPI_ISL_700045, EPI_ISL_700046, EPI_ISL_700047, EPI_ISL_700048, EPI_ISL_700049, EPI_ISL_700050, EPI_ISL_700051, EPI_ISL_700052, EPI_ISL_700053, EPI_ISL_700054, EPI_ISL_700055, EPI_ISL_700056, EPI_ISL_700057, EPI_ISL_700058, EPI_ISL_700059, EPI_ISL_700060, EPI_ISL_700061, EPI_ISL_700062, EPI_ISL_700063, EPI_ISL_700064, EPI_ISL_700065, EPI_ISL_700066, EPI_ISL_700067, EPI_ISL_700068, EPI_ISL_700069, EPI_ISL_700071, EPI_ISL_700072, EPI_ISL_700073, EPI_ISL_700074, EPI_ISL_700075, EPI_ISL_700076, EPI_ISL_700077, EPI_ISL_700078, EPI_ISL_700079, EPI_ISL_700080, EPI_ISL_700081, EPI_ISL_700082, EPI_ISL_700083, EPI_ISL_700084, EPI_ISL_700085, EPI_ISL_700086, EPI_ISL_700087, EPI_ISL_700088, EPI_ISL_700089, EPI_ISL_700090, EPI_ISL_700091, EPI_ISL_700092, EPI_ISL_700093, EPI_ISL_700094, EPI_ISL_700095, EPI_ISL_700096, EPI_ISL_700097, EPI_ISL_700098, EPI_ISL_700099, EPI_ISL_700100, EPI_ISL_700101, EPI_ISL_700102, EPI_ISL_700103, EPI_ISL_700104, EPI_ISL_700105, EPI_ISL_700106, EPI_ISL_700107, EPI_ISL_700108, EPI_ISL_700109, EPI_ISL_700110, EPI_ISL_700111, EPI_ISL_700112, EPI_ISL_700113, EPI_ISL_700114, EPI_ISL_700115, EPI_ISL_700116, EPI_ISL_700117, EPI_ISL_700118, EPI_ISL_700119, EPI_ISL_700120, EPI_ISL_700121, EPI_ISL_700122, EPI_ISL_700123, EPI_ISL_700124, EPI_ISL_700125, EPI_ISL_700126, EPI_ISL_700127, EPI_ISL_700128, EPI_ISL_700129, EPI_ISL_700130, EPI_ISL_700131, EPI_ISL_700132, EPI_ISL_700133, EPI_ISL_700134, EPI_ISL_700135, EPI_ISL_700136, EPI_ISL_700137, EPI_ISL_700138, EPI_ISL_700139, EPI_ISL_700140, EPI_ISL_700141, EPI_ISL_700142, EPI_ISL_700143, EPI_ISL_700144, EPI_ISL_700145, EPI_ISL_700146, EPI_ISL_700147, EPI_ISL_700148, EPI_ISL_700149, EPI_ISL_700150, EPI_ISL_700151, EPI_ISL_700152, EPI_ISL_700153, EPI_ISL_700154, EPI_ISL_700155, EPI_ISL_700156, EPI_ISL_700157, EPI_ISL_700158, EPI_ISL_700159, EPI_ISL_700160, EPI_ISL_700161, EPI_ISL_700162, EPI_ISL_700163, EPI_ISL_700164, EPI_ISL_700165, EPI_ISL_700166, EPI_ISL_700167, EPI_ISL_700168, EPI_ISL_700169, EPI_ISL_700170, EPI_ISL_700171, EPI_ISL_700172, EPI_ISL_700173, EPI_ISL_700174, EPI_ISL_700175, EPI_ISL_700176, EPI_ISL_700177, EPI_ISL_700178, EPI_ISL_700179, EPI_ISL_700180, EPI_ISL_700181, EPI_ISL_700182, EPI_ISL_700183, EPI_ISL_700184, EPI_ISL_700185, EPI_ISL_700186, EPI_ISL_700187, EPI_ISL_700188, EPI_ISL_700189, EPI_ISL_700190, EPI_ISL_700191, EPI_ISL_700192, EPI_ISL_700193, EPI_ISL_700194, EPI_ISL_700195, EPI_ISL_700196, EPI_ISL_700197, EPI_ISL_700198, EPI_ISL_700199, EPI_ISL_700200, EPI_ISL_700201, EPI_ISL_700202, EPI_ISL_700203, EPI_ISL_700204, EPI_ISL_700205, EPI_ISL_700206, EPI_ISL_700207, EPI_ISL_700208, EPI_ISL_700209, EPI_ISL_700210, EPI_ISL_700211, EPI_ISL_700212, EPI_ISL_700213, EPI_ISL_700214, EPI_ISL_700215, EPI_ISL_700216, EPI_ISL_700217, EPI_ISL_700218, EPI_ISL_700219, EPI_ISL_700220, EPI_ISL_700221, EPI_ISL_700222, EPI_ISL_700223, EPI_ISL_700224, EPI_ISL_700225, EPI_ISL_700226, EPI_ISL_700227, EPI_ISL_700228, EPI_ISL_700229, EPI_ISL_700230, EPI_ISL_700231, EPI_ISL_700232, EPI_ISL_700233, EPI_ISL_700234, EPI_ISL_700235, EPI_ISL_700236, EPI_ISL_700237, EPI_ISL_700238, EPI_ISL_700239, EPI_ISL_700240, EPI_ISL_700241, EPI_ISL_700242, EPI_ISL_700243, EPI_ISL_700244, EPI_ISL_700245, EPI_ISL_700246, EPI_ISL_700247, EPI_ISL_700248, EPI_ISL_700249, EPI_ISL_700250, EPI_ISL_700251, EPI_ISL_700252, EPI_ISL_700253, EPI_ISL_700254, EPI_ISL_700255, EPI_ISL_700256, EPI_ISL_700257, EPI_ISL_700258, EPI_ISL_700259, EPI_ISL_700260, EPI_ISL_700261, EPI_ISL_700262, EPI_ISL_700263, EPI_ISL_700264, EPI_ISL_700265, EPI_ISL_700266, EPI_ISL_700267, EPI_ISL_700268, EPI_ISL_700269, EPI_ISL_700270, EPI_ISL_700271, EPI_ISL_700272, EPI_ISL_700273, EPI_ISL_700274, EPI_ISL_700275, EPI_ISL_700276, EPI_ISL_700277, EPI_ISL_700278, EPI_ISL_700279, EPI_ISL_700280, EPI_ISL_700281, EPI_ISL_700282, EPI_ISL_700283, EPI_ISL_700284, EPI_ISL_700285, EPI_ISL_700286, EPI_ISL_700287, EPI_ISL_700288, EPI_ISL_700289, EPI_ISL_700290, EPI_ISL_700291, EPI_ISL_700292, EPI_ISL_700293, EPI_ISL_700294, EPI_ISL_700295, EPI_ISL_700296, EPI_ISL_700297, EPI_ISL_700298, EPI_ISL_700299, EPI_ISL_700300, EPI_ISL_700301, EPI_ISL_700302, EPI_ISL_700303, EPI_ISL_700304, EPI_ISL_700305, EPI_ISL_700307, EPI_ISL_700308, EPI_ISL_700309, EPI_ISL_700310, EPI_ISL_700312, EPI_ISL_700314, EPI_ISL_700315, EPI_ISL_700316, EPI_ISL_700317, EPI_ISL_700318, EPI_ISL_700319, EPI_ISL_700320, EPI_ISL_700321, EPI_ISL_700322, EPI_ISL_700323, EPI_ISL_700324, EPI_ISL_700325 | | | |
| see above | Hematopathology Laboratory, ACTREC, TMC | Hematopathology Laboratory, ACTREC, TMC | Hematopathology Laboratory, ACTREC |
| EPI_ISL_721629, EPI_ISL_721630, EPI_ISL_721631, EPI_ISL_721632, EPI_ISL_721633, EPI_ISL_721634, EPI_ISL_721635, EPI_ISL_721636, EPI_ISL_721637, EPI_ISL_721638, EPI_ISL_721639, EPI_ISL_721640, EPI_ISL_721641, EPI_ISL_721642, EPI_ISL_721643, EPI_ISL_721644, EPI_ISL_721645 | | | |
| see above | Armed Forces Medical College | National Centre For Cell Science | Dhiraj Paul, Kunal Jani, Radha Chauhan, Janesh Kumar, Vasudevan Seshadri, Girdhari Lal, Rajesh Karyakarte, Suvarna Joshi, Murlidhar Tambe, Sourav Sen, Santosh Karade, Kavita Bala Anand, Shelinder Pal Singh Shergill, Rajiv Mohan Gupta, Manoj Kumar Bhat, Arvind Sahu, Yogesh S Shouche |
| EPI_ISL_721646, EPI_ISL_721647, EPI_ISL_721648, EPI_ISL_721649, EPI_ISL_721650, EPI_ISL_721651, EPI_ISL_721653, EPI_ISL_721654, EPI_ISL_721655, EPI_ISL_721656, EPI_ISL_721657, EPI_ISL_721658, EPI_ISL_721659, EPI_ISL_721660, EPI_ISL_721661, EPI_ISL_721662, EPI_ISL_721663, EPI_ISL_721664, EPI_ISL_721665, EPI_ISL_721666, EPI_ISL_721667, EPI_ISL_722177, EPI_ISL_722178, EPI_ISL_722179, EPI_ISL_722180, EPI_ISL_722181, EPI_ISL_722182, EPI_ISL_722183, EPI_ISL_722184, EPI_ISL_722185, EPI_ISL_722186 | | | |
| see above | National Centre For Cell Science | National Centre For Cell Science | Dhiraj Paul, Kunal Jani, Radha Chauhan, Janesh Kumar, Vasudevan Seshadri, Girdhari Lal, Rajesh Karyakarte, Suvarna Joshi, Murlidhar Tambe, Sourav Sen, Santosh Karade, Kavita Bala Anand, Shelinder Pal Singh Shergill, Rajiv Mohan Gupta, Manoj Kumar Bhat, Arvind Sahu, Yogesh S Shouche |
| EPI_ISL_722187, EPI_ISL_722188, EPI_ISL_722189, EPI_ISL_722190, EPI_ISL_722191, EPI_ISL_722192, EPI_ISL_722193, EPI_ISL_722194, EPI_ISL_722195, EPI_ISL_722196, EPI_ISL_722197, EPI_ISL_722198, EPI_ISL_722199, EPI_ISL_722200 | | | |
| see above | Armed Forces Medical College | National Centre For Cell Science | Dhiraj Paul, Kunal Jani, Radha Chauhan, Janesh Kumar, Vasudevan Seshadri, Girdhari Lal, Rajesh Karyakarte, Suvarna Joshi, Murlidhar Tambe, Sourav Sen, Santosh Karade, Kavita Bala Anand, Shelinder Pal Singh Shergill, Rajiv Mohan Gupta, Manoj Kumar Bhat, Arvind Sahu, Yogesh S Shouche |
| EPI_ISL_723045, EPI_ISL_723046, EPI_ISL_723047, EPI_ISL_723048, EPI_ISL_723049, EPI_ISL_723050, EPI_ISL_723051, EPI_ISL_723052, EPI_ISL_723053, EPI_ISL_723054, EPI_ISL_723055, EPI_ISL_723056, EPI_ISL_723057, EPI_ISL_723058, EPI_ISL_723059, EPI_ISL_723060, EPI_ISL_723061, EPI_ISL_723062, EPI_ISL_723063, EPI_ISL_723064, EPI_ISL_723065, EPI_ISL_723066, EPI_ISL_723067, EPI_ISL_723068, EPI_ISL_723069, EPI_ISL_723070, EPI_ISL_723071, EPI_ISL_723072, EPI_ISL_723073, EPI_ISL_723074, EPI_ISL_723075, EPI_ISL_723076 | | | |
| see above | Hematopathology Laboratory, ACTREC, TMC | Hematopathology Laboratory, ACTREC, TMC | Hematopathology Laboratory, ACTREC |
| EPI_ISL_728254, EPI_ISL_728255, EPI_ISL_728256, EPI_ISL_728257, EPI_ISL_728258, EPI_ISL_728259, EPI_ISL_728260, EPI_ISL_728261, EPI_ISL_728262, EPI_ISL_728263, EPI_ISL_728264, EPI_ISL_728265, EPI_ISL_728266, EPI_ISL_728267, EPI_ISL_728268, EPI_ISL_728269, EPI_ISL_728270, EPI_ISL_728271, EPI_ISL_728325, EPI_ISL_728326, EPI_ISL_728327, EPI_ISL_728328, EPI_ISL_728329, EPI_ISL_728330 | | | |
| see above | B.J. Govt. Medical College | National Centre For Cell Science | Dhiraj Paul, Kunal Jani, Radha Chauhan, Janesh Kumar, Vasudevan Seshadri, Girdhari Lal, Rajesh Karyakarte, Suvarna Joshi, Murlidhar Tambe, Sourav Sen, Santosh Karade, Kavita Bala Anand, Shelinder Pal Singh Shergill, Rajiv Mohan Gupta, Manoj Kumar Bhat, Arvind Sahu, Yogesh S Shouche |
| EPI_ISL_779703, EPI_ISL_779705, EPI_ISL_779706, EPI_ISL_779708, EPI_ISL_779710 | The Foundation for Medical Research | The Foundation for Medical Research | Ayan Mandal, Kayzad Nilgiriwala, Kalpana Sriraman, Ambreen Shaikh, Grishma Patel, Tejal Mestry, Smriti Vaswani, Swapneil Parikh, Shreevatsa Udupa, Nirjhar Chatterjee, Jayanthi Shastri, Nerges Mistry |
| EPI_ISL_801538 | TNMC & BYL NAIR CH. HOSPITAL | CSIR-IGIB | Rajesh Pandey, Jayanthi Shastri, Akshay Kanakan, Janani Srinivasa Vasudevan, Ranjeet Maurya, Sachee Agrawal, Nirjhar Chatterjee, Swapneil Parikh, Manish Pathak, Subrat Thanapati, Jasmina Savak, Suresh Poojari, Mahesh Sangar, Amol Borse, Shweta Kawankar, Vasil Nachan, Mayuresh Vishwanathan, Shruthi Sachidanandan, Shrutika Pophale, Utkarsha Yelve |
| EPI_ISL_969397, EPI_ISL_969509, EPI_ISL_969740 | Indian Council of Medical Research-National Institute of Virology, Pune | Indian Council of Medical Research-National Institute of Virology, Maximum Containment Laboratory | Pragya D. Yadav, Anita Shete-Aich, Dimpal A. Nyayanit, Abhinendra Kumar, Savita Patil, Triparna D. Majumdar |
